# TGIRT-seq of Inflammatory Breast Cancer Tumor and Blood Samples Reveals Widespread Enhanced Transcription Impacting RNA Splicing and Intronic RNAs in Plasma

**DOI:** 10.1101/2023.05.26.23290469

**Authors:** Dennis Wylie, Xiaoping Wang, Jun Yao, Hengyi Xu, Elizabeth A. Ferrick-Kiddie, Toshiaki Iwase, Savitri Krishnamurthy, Naoto T. Ueno, Alan M. Lambowitz

## Abstract

Inflammatory breast cancer (IBC) is the most aggressive and lethal breast cancer subtype but lacks unequivocal genomic differences or robust biomarkers that differentiate it from non-IBC. Here, Thermostable Group II intron Reverse Transcriptase RNA-sequencing (TGIRT-seq) revealed myriad differences in tumor samples, Peripheral Blood Mononuclear Cells (PBMCs), and plasma that distinguished IBC from non-IBC patients and healthy donors across all tested receptor-based subtypes. These included numerous differentially expressed protein-coding gene and non-coding RNAs in all three sample types, a granulocytic immune response in IBC PBMCs, and over- expression of antisense RNAs, suggesting wide-spread enhanced transcription in both IBC tumors and PBMCs. By using TGIRT-seq to quantitate Intron-exon Depth Ratios (IDRs) and mapping reads to both genome and transcriptome reference sequences, we developed methods for parallel analysis of transcriptional and post-transcriptional gene regulation. This analysis identified numerous differentially and non-differentially expressed protein-coding genes in IBC tumors and PBMCs with high IDRs, the latter reflecting rate-limiting RNA splicing that negatively impacts mRNA production. Mirroring gene expression differences in tumors and PBMCs, over-represented protein-coding gene RNAs in IBC patient plasma were largely intronic RNAs, while those in non-IBC patients and healthy donor plasma were largely mRNA fragments. Potential IBC biomarkers in plasma included T-cell receptor pre-mRNAs and intronic, LINE-1, and antisense RNAs. Our findings provide new insights into IBC and set the stage for monitoring disease progression and response to treatment by liquid biopsy. The methods developed for parallel transcriptional and post-transcriptional gene regulation analysis have potentially broad RNA-seq and clinical applications.

## INTRODUCTION

High-throughput RNA sequencing (RNA-seq) is widely used to identify RNA biomarkers for diseases and environmental stimuli (*1*). Commonly, an initial comprehensive RNA-seq of one or more RNA biotypes is performed to identify candidate biomarkers, which are then validated by targeted assays in larger numbers of samples. Most desired are biomarkers that can be detected by minimally invasive blood-based assays and reflect molecular aberrations and immune responses throughout the body. For this purpose, labile blood-based RNA biomarkers are potentially advantageous by enabling diagnosis and real-time monitoring of disease progression and response to treatment. Thus far, most cellular and blood-based RNA biomarker studies have focused on polyadenylated mRNAs or mature miRNAs. However, these approaches miss numerous long non-coding RNAs (lncRNAs) and small non-coding RNAs (sncRNAs), as well as intron and RNAs that can be detected in blood but have not been widely explored for biomarker potentials (*2, 3*).

Inflammatory breast cancer (IBC), the most lethal and aggressive breast cancer subtype (*4*), comprises only 2-4% of breast cancer cases in the United States but causes 8-10% of breast cancer deaths (*5*). Due to its high recurrence rate and propensity to metastasize, IBC patients have a shorter median overall survival time than those with other types of breast cancer (denoted non-IBC) (*6–8*). Triple-negative IBC patients (TN-IBC), negative for estrogen receptor (ER), progesterone receptor (PR), and human epidermal growth factor receptor-2 (HER2), have the worst prognosis among receptor-based molecular subtypes (*9*). Although a tri-modality treatment (*10*), consisting of anthracycline-based systemic chemotherapy, modified radical mastectomy, and postoperative radiation therapy, has substantially improved patient survival outcomes, the median survival of patients with stage III IBC is only 4.75 years (*11*). Whole-genome sequencing found that IBC and non-IBC tumors have similar mutation frequencies for most cancer-related genes and pathways, with no strongly distinguishing features (*12*). FDA-approved targeted therapies specific to IBC are unavailable, and IBC diagnosis is currently based solely on clinical features (*13*). A particular challenge for IBC is that invasive tumors are often distributed as small foci without forming a discrete tumor mass. Thus, it is difficult to obtain high-quality core biopsies with high tumor cellularity, and the rarity of this disease limits patient sample size for optimal analyses. These challenges reinforce the critical importance of discovering blood-based biomarkers for diagnosing and developing treatments for IBC patients.

RNA-seq methods used for biomarker identification typically employ engineered retroviral reverse transcriptases (RTs) to convert RNAs into more readily sequenced cDNAs (*1, 14*). However, retroviral RTs have inherently low fidelity and processivity, reflecting the loss of key regions used by ancestral RTs to bind template RNAs and constrain substrate binding pockets (*15*). Recent studies showed that group II intron and other non-long-terminal repeat (non-LTR)-retro-element RTs, evolutionary ancestors of retroviral RTs, can outperform retroviral RTs for RNA-seq and genome engineering applications (*16–21*). Previously, we identified Thermostable Group II Intron Reverse Transcriptases (TGIRTs) from bacterial thermophiles that have higher fidelity, processivity, and strand displacement activity than retroviral RTs, as well as a proficient template-switching activity, which enables efficient, seamless RNA-seq adapter addition from small amounts of RNA (*16*). These properties make it possible for TGIRTs to give full-length, end-to-end sequence reads of highly modified tRNAs and other structured sncRNAs and enabled the development of TGIRT-seq, a fully validated RNA-seq method for comprehensive profiling of coding and non-coding RNAs, including those in plasma and extracellular vesicles (*2, 3, 22–24*).

Here, we profiled RNAs from Formalin-Fixed Paraffin-Embedded (FFPE) tumor slices, PBMCs, and plasma from cohorts of IBC and non-IBC patients and healthy donors by using new methods for TGIRT-seq library preparation and parallel analysis of transcriptional and post-transcriptional gene regulation. In contrast to whole-genome DNA sequencing (*12*), we found pronounced differences in all 3 sample types that distinguished IBC from non-IBC patients with all tested hormone receptor and HER2 subtypes, obtained new insights into the dysregulation of gene expression in IBC, found a direct correlation between over-expressed RNAs in tumors and PBMCs with those in plasma, and identified potential IBC biomarkers in plasma.

## RESULTS

### RNA samples and TGIRT-seq library preparation

The samples collected at MD Anderson for this study included FFPE tumor slices, PBMCs, and plasma from 10 IBC and 4 non-IBC patients, plus PBMCs and plasma from 2 additional non-IBC patients and 13 healthy female donors. To supplement these samples, we purchased RNAs isolated from matched frozen healthy breast tissues from 4 other non-IBC patients (OriGene). Patient information, including hormone receptor (HR) and Human Epidermal Growth Factor 2 (HER2) status, is summarized in Table S1.

For TGIRT-seq library preparation, cellular RNA samples were rRNA-depleted and separated into long and short fractions using a commercial kit with a 200-nt size cutoff (Methods). The long RNAs were then chemically fragmented to enable optimal quantitation of mRNAs and lncRNAs (*22, 25*) and recombined with untreated small RNAs for TGIRT-seq library preparation. Plasma RNAs (1-2 ng of RNA from 200 ml of plasma) were used for TGIRT-seq library preparation without rRNA depletion or size selection. Libraries were sequenced on an Illumina NextSeq to obtain 12.3 to 81.6 million reads per sample. Mapping statistics are summarized in the Supplemental (Suppl.) File.

Principal component analysis (PCA) showed that sample type was the major source of RNA sequence differences (Fig. S1). PC1 separated non-IBC FFPE and frozen breast tissue samples, possibly reflecting different modes of tissue preservation. Notably, PCAs based on single categories of cellular RNA biotypes (protein-coding gene, lncRNAs, sncRNAs, and RepBase-annotated RNAs (denoted Repeat-Element RNAs, https://www.girinst.org/repbase/)) largely separated IBC patients from healthy donor and non-IBC patient samples in all cases except for sncRNAs in FFPE tumor samples, reflecting findings below that only a small number of sncRNAs were differentially expressed between these samples. Collectively, the PCA indicated that different categories of RNA biotypes, including Repeat-Element RNAs, not commonly thought of as a rich source of RNA biomarkers, included sufficient differentially expressed (DE) RNAs to distinguish disease states albeit with lower resolution than all RNA biotypes.

### RNA biotypes detected by TGIRT-seq in different samples

Stacked bar graphs depicting the percentages of TGIRT-seq reads corresponding to different RNA biotypes for each of the 80 samples are shown in Fig. 1 with differences noted below supported by paired Wilcoxon sign ranked tests in the Suppl. File. Reads mapping to protein-coding genes comprised the highest percentages of total reads in both the breast tissue and PBMC samples (31- 80%), but lower percentages in plasma samples (3-41%), whereas reads mapping to cellular and mitochondrial (MT) tRNAs, sncRNAs, and Repeat-Element RNAs comprised higher and more variable percentages in plasma (Fig. 1A, top panel). 7SL, snoRNAs, and snRNAs comprised the highest percentages of sncRNA reads in the cellular RNA samples, with snoRNAs comprising higher percentages of the sncRNA reads in PBMCs compared to the other sample types (Fig. 1B). Y RNAs comprised the highest percentage of sncRNA reads in most of the plasma samples (Fig. 1B). Based on the percentage of total reads, cellular and MT tRNAs appeared to be upregulated relative to other RNA biotypes in IBC compared to non-IBC FFPE tumors and even more so in IBC PBMCs, the latter likely reflecting a high degree of PBMC activation in IBC. By contrast, tRNAs appeared to be downregulated in non-IBC PBMCs compared to both healthy and IBC PBMCs, possibly reflecting suppression of immune responses in non-IBC patients (*26*), supported by additional findings below.

**Fig. 1.**
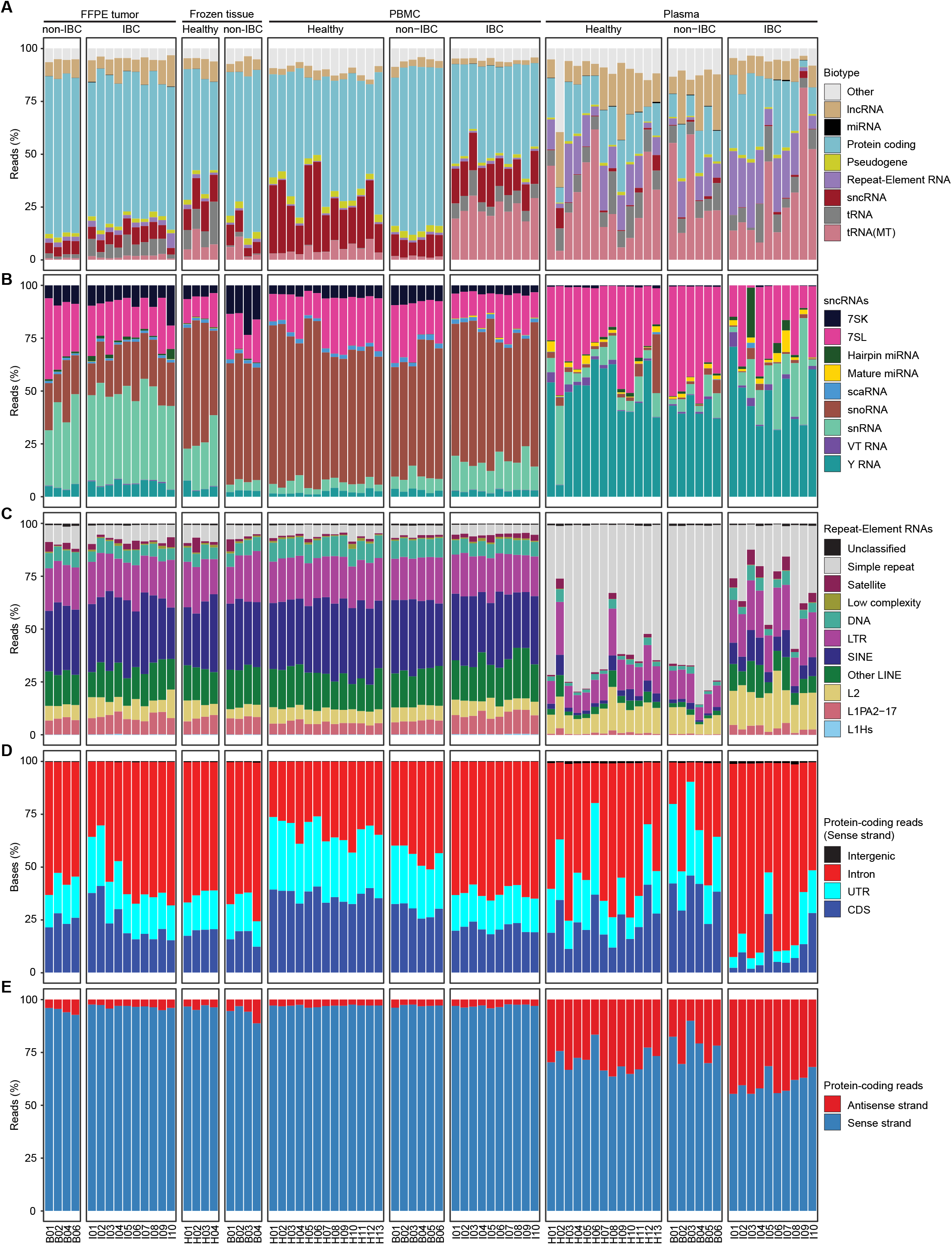
Distribution of reads mapping to different RNA biotypes in different sample sets. Stacked bar graphs showing the percentage of reads in each sample assigned to different RNA biotypes (Ensembl GRCh38 Release 93). (**A**) Major RNA biotype categories excluding rRNA; (**B**) sncRNA categories; (**C**) Repeat-Element RNA categories; (**D**) Different regions of protein-coding genes, including Introns, Coding Sequences (CDS), and Untranslated Exon Regions (UTR); (**E**) Protein-coding gene Antisense and Sense strands. Sample types (FFPE tumors, frozen Neighboring Healthy Breast Tissue, frozen non-IBC tumors, PBMCs, or Plasma), with disease status (Healthy, non-IBC, or IBC) are indicated above. Sample names corresponding to healthy donor or patient ID numbers are indicated below (H, Healthy; B, non-IBC; I, IBC). Paired Wilcoxon sign rank test results for differences in biotype read distributions between sample types can be found in the Suppl file.

The protein-coding gene reads for tumors and PBMCs included relatively high percentages mapping to intron RNAs (30-75% in tumors, 25-65% in PBMC; Fig. 1D), in agreement with previous findings for TGIRT-seq and TruSeq of chemically fragmented Universal Human Reference RNA (*22*). The protein-coding gene reads for breast tissue and PBMC samples mapped predominantly (>88%) to the annotated sense strand, as expected for cellular RNAs, while those for plasma samples included substantially higher proportions (11-45%) mapping to the antisense strand (Fig. 1E). Notably, the proportions of reads mapping to protein-gene intron and antisense RNAs tended to be higher in most of the IBC plasma samples (51-92% and 32-45%, respectively) than in most of the healthy and non-IBC plasma samples (9-74% and 10-36%; Fig. 1D and E).

Repeat-Element RNAs annotated in RepBase constituted only low proportions of total reads in the cellular tumor and PBMC samples (1.6-7.0% in FFPE tumors, 0.4-1.5% in PBMCs), but higher proportions of total reads in most of the plasma samples (up to 31%), indicating preferential export and/or higher stability of these RNAs in plasma (Fig. 1A). Notably, retroelement RNAs, including Long Interspersed Nuclear Elements (LINEs), abundant but normally repressed retrotransposons in the human genome; Short Interspersed Nuclear Elements (SINEs), non-coding RNAs mobilized by LINE RTs; and endogenous retrovirus and other LTR-containing retroelement RNAs comprised higher proportions of the Repeat-Element RNAs in most of the IBC plasma samples than in most of the healthy or non-IBC plasma samples (Fig. 1C).

The distribution of reads among RNA biotypes was more variable for the plasma than cellular RNA samples irrespective of disease state, with tRNAs being the most variably abundant RNA biotype (Fig. 1A). This variability may reflect different degrees of cell breakage, despite precautions in the method used for plasma preparation, and/or variable degradation of more labile RNA species in clinical plasma samples, which were prepared at different times and used for multiple purposes. Although these caveats should be kept in mind, they did not prevent us from detecting reproducible IBC-specific RNAs in plasma, as described below.

### Differential gene expression and intron-depth ratio (IDR) analyses

We performed gene-wise differential expression (DE) analysis of TGIRT-seq reads mapped separately to both the Ensembl GRCh38 genome (Exons + Introns) and transcriptome (Exons only) reference sequences using the DESeq2 R package (https://github.com/thelovelab/DESeq2) with size factors calculated based on all protein-coding gene reads to minimize the impact of variations in sncRNA composition. The protein-coding gene RNAs detected by TGIRT-seq in cellular RNA samples had different ratios of reads that mapped to introns or exons. To quantitate these differences, we developed a bioinformatic parameter termed Intron-exon Depth Ratio (IDR) based on genome annotations and comparing reads mapped to genome or transcriptome reference sequences to determine the intron inclusion frequency (θ*_g_*) for each gene in each cellular RNA sample (see equation in Methods).

Fig. 2A shows a plot of the fraction of intronic RNA detected by TGIRT-seq compared to that annotated in the genomic reference sequence for each protein-coding gene in a combined dataset for the 10 IBC FFPE tumor samples (dots color-coded by gene length). The plot shows an overall upward trend, reflecting that genes comprised predominantly of intron sequences contribute more intronically derived reads but with wide variance in IDR values due to differences in the relative rates of transcription, RNA splicing, and intron RNA or mRNA turnover. A gene lying on the black diagonal has an IDR of 1, indicating a high level of intronic relative to exonic RNA, in many cases extending across the length of the gene suggesting accumulation of unspliced pre-mRNAs (see below). Genes having a more typical IDR of 0.19 lie on the red curve. Upset plots showed that FFPE tumor samples from non-IBC and IBC patients and PBMC samples from healthy donor, non-IBC, and IBC patients each had large distinct subsets of genes with moderate 0.5 to ≤1.0 or high IDRs (≥1.0; black bars and dots; Fig. 2B, upper and lower plot, respectively). Protein-coding and lncRNA genes with IDRs ≥0.5 are indicated with an asterisk in descriptions below.

**Fig. 2.**
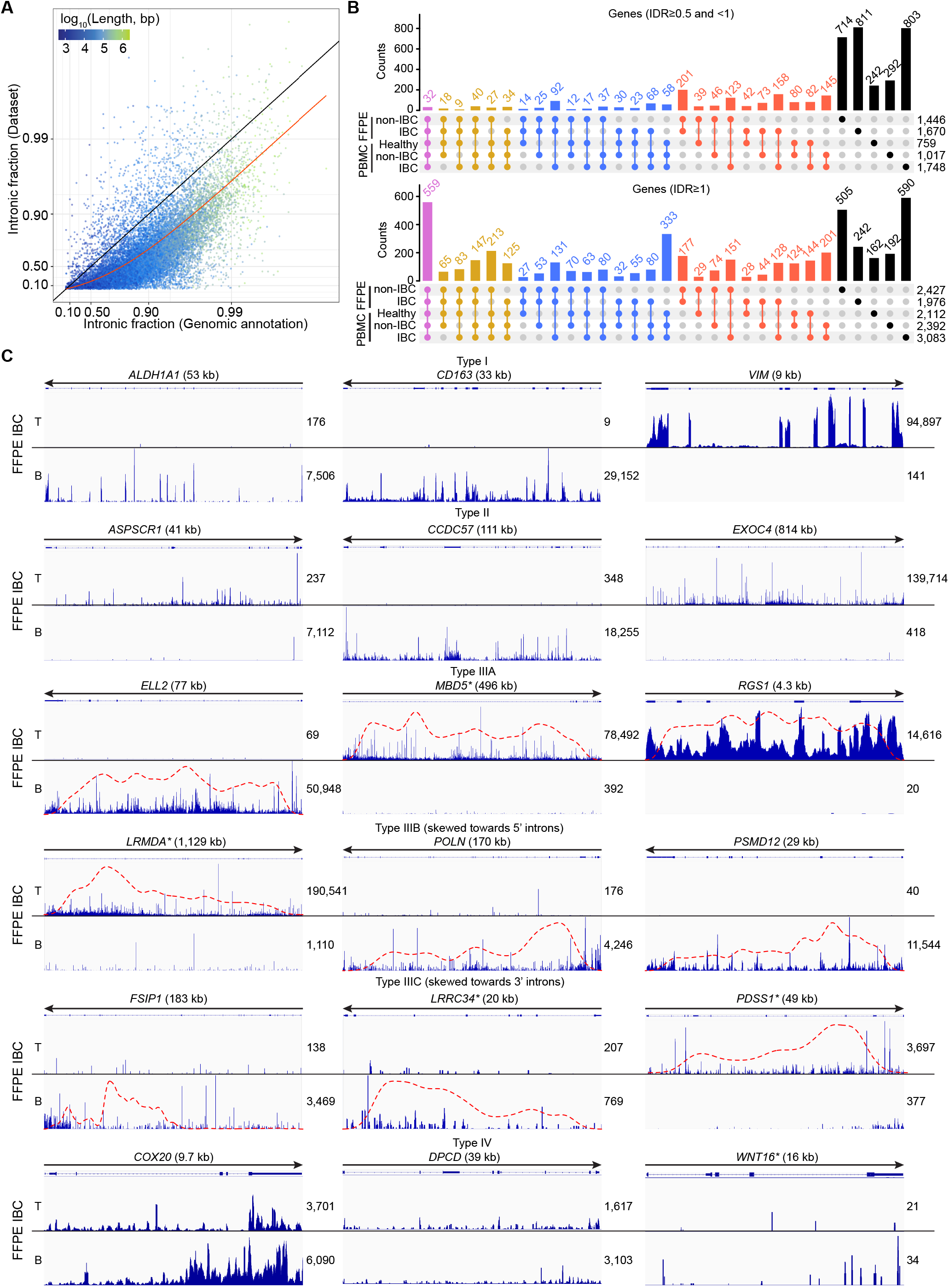
Categories of protein-coding genes transcripts identified by TGIRT-seq. (**A**) Intron- exon Depth Ratio (IDR) analysis. Each point indicates the fraction of reads for a protein-coding gene in IBC FFPE tumor samples derived from introns (*y*-axis, -log_10_(1-*y*) scaling) compared to the fraction in their genomic annotation (*x*-axis, -log_10_(1-*x*) scaling). Points are colored according to the log_10_-transformed length in base pairs (bp) of the genomic interval encoding the gene (exons and introns included): longer genomic intervals tend to be associated with relatively high genomic intron fractions, as is evident from color gradient from left to right. The black line indicates equality *y*=*x*, corresponding to the set of points representing genes with IDR=1; points above or below this line represent genes with IDRs greater or less than 1, respectively. The red trend line plots the solution to the IDR equation (see Methods) when applied to data for all genes taken together, yielding a typical IDR value of ∼0.19. (**B**) Upset plot showing the number of genes with IDRs between 0.5 and 1 (top panel) and IDR≥1 (bottom panel) in different combined datasets indicated to the left of the plots. (**C**) Integrated Genome Viewer (IGV) plots for representative protein-coding genes from combined IBC FFPE tumor datasets Arrows for gene maps above the IGV plots indicate the 5’ to 3’ orientation of the gene, with the number of reads that mapped to the top (T) or bottom (B) strands indicated to the right of the plot for each strand. Type I to IV genes are defined in Results and were classified into different categories as described in Methods. Density plots for skewness of intron body coverage are shown as red dashed lines. Protein-coding genes with IDR>0.5 are marked with an asterisk.

Examination of IGV alignments (Fig. 2C) showed that protein-coding gene RNAs could be divided into the following different categories based on the depth and distribution of intron RNA reads: Type I, mean coverage depth of exons significantly higher than that of introns (*t*-test *p*≤0.05), indicating a preponderance of spliced mature mRNA; Type II, no significant difference in mean coverage depth between exons and introns, likely reflecting rate-limiting RNA splicing, resulting in accumulation of unspliced or partially spliced pre-mRNAs if the reads spanned multiple introns; Type IIIA, mean coverage depth across introns significantly higher than that of exons (*t*-test *p*≤0.05) due to some combination of rate-limiting RNA splicing, accumulation of excised intron RNAs, or a higher rate of mRNA turnover; Type IIIB, similar to Type IIIA but with reads coverage skewed towards 5’ introns (skewness ≥0.5), possibly corresponding to nascent transcripts or reflecting gene regulation via transcriptional pausing or arrest; Type IIIC, similar to Type IIIA but with reads coverage skewed towards 3’ introns (skewness ≤ -0.5), possibly reflecting post-transcriptional regulation by discard pathways that abort RNA splicing (*27, 28*); and Type IV, genes with a high proportion of antisense reads (≥30%), suggesting activation of cryptic promoters within the same or neighboring genes, a potential indicator of widespread enhanced transcription. Mapping TGIRT-seq reads to both genome and transcriptome reference sequences and analyzing IDRs revealed wide-spread post-transcriptional regulation in IBC tumors and PBMCs that would be missed by only quantitating mRNAs.

All genes mentioned in the text are listed in the Suppl. File with their log_2_-fold-changes (LFCs), *p*-values, and false discovery rate (FDR) adjusted (adj.) *p*-values for reads mapped separately to genome and transcriptome reference sequences. Descriptions and cancer relevance of all highly DE genes in IBC (adj. *p*≤0.001, |LFC|≥2) are summarized in Table S2 with literature references. Genes were singled out for inclusion in heat maps and discussion below according to several criteria: (i) their individual DE statistics; (ii) their membership in potentially significant biological pathways or gene sets; (iii) previously reported findings of cancer relevance.

### Differential gene expression in IBC tumors

We focus first on RNAs detected in tumors. Given the potential issues with differences in tissue preservation methods found by PCA (Fig. S1), our primary comparison was IBC vs non-IBC patient FFPE tumor samples, a widely used comparison in the literature (*29–34*). For this comparison, TGIRT-seq reads mapped to the human genome reference sequence (Exons + Introns) identified 43 significantly DE genes (28 protein-coding + 15 non-coding RNAs) with adj. *p*≤0.001 and |LFC|≥2, increasing to 637 (222 protein-coding + 415 non-coding RNAs) with adj. *p*≤0.05 and |LFC|≥1; Suppl. File). A heat map of significantly DE protein-coding and non-coding RNAs in IBC versus non-IBC patient FFPE tumor samples at adj. *p*≤0.001 and |LFC|≥2 and detected in half or more of the IBC samples is shown in Fig. 3A with volcano plot comparisons and IGV alignments for different categories of RNAs shown in Fig. S2 and Fig. S3, respectively.

**Fig. 3.**
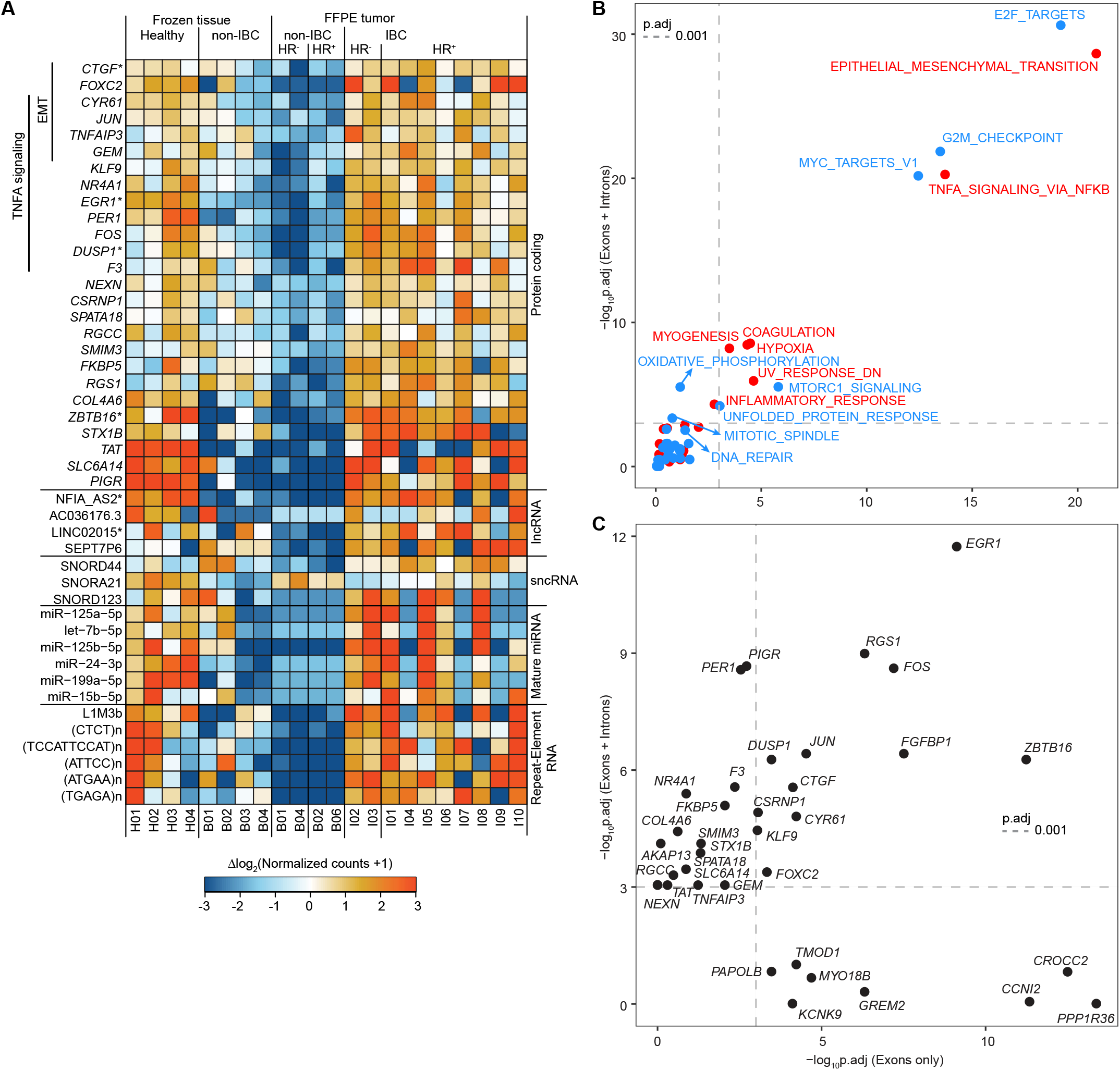
Heat map of differentially expressed genes in healthy breast tissue or tumor samples and scatter plots comparing significantly enriched gene sets and differentially expressed genes in IBC versus non-IBC FFPE tumor samples. **(A)** Heat map showing significantly differentially expressed genes (adj. *p*≤0.001, LFC≥2) detected in half or more of the IBC FFPE tumor samples. The color of the tile for gene *g* in sample *s* encodes the difference in the (log_2_-transformed DESeq2- normalized) expression level for sample *s* from the overall mean expression level for gene *g* for all samples of same type (Frozen tissue or FFPE). The color scale is shown below the heat map. Lanes are labeled at the bottom according to healthy donor and patient ID numbers (Table S1). An additional protein-coding gene RNA (*FGFBP1*) and 3 additional lncRNAs (AC044784.1, LINC02562, and AC006924.1*) met the significance and fold change criteria for over-expression in IBC FFPE tumors but were not detected in half of the IBC samples and thus not included in the heat map. Protein-coding and lncRNA genes with IDR>0.5 are marked with an asterisk. **(B**) Scatter plots comparing (log_10_-transformed and sign-inverted) FDR-adjusted *p*-values from GO-MWU enrichment of members of BROAD Hallmark gene sets in IBC versus non-IBC FFPE tumors using gene counts for reads mapped to transcriptome (Exons only; *x*-axis) or genome reference sequences (Exons + Introns; *y*-axis). Gene sets for which the average gene MWU test statistic indicated increased expression in IBC are colored red, while those with significantly decreased expression in IBC are colored blue. Vertical and horizontal dashed lines indicate adj. *p*=0.001. (**C**) Scatter plots comparing (log_10_-transformed and sign-inverted) FDR-adjusted *p*-values for DE of the most significantly over-expressed genes in IBC tumors based on Exon only mapping of reads (*x*-axis) to Exon + Intron mapping of reads (*y*-axis). Only genes for which adj. *p*≤0.001 and LFC≥1 in the analysis based on one or both of these mappings (Exons only or Exons + Introns) are shown.

The significantly DE protein-coding genes in IBC tumors that met the criteria for inclusion in Fig. 3A included 11 members of the Hallmark TNFα-signaling-via-NFκB (denoted TNFα-via- NFκB) gene set: *JUN*, *GEM*, *KLF9*, *NR4A1*, *EGR1*\*, *PER1*, *FOS*, *DUSP1*\*, *F3*, *CYR61*\* (aka. *CCN1*), and *TNFAIP3\** (Fig. 3A). *JUN*, *CYR61*\*, *GEM*, and *TNFAIP3*\* are also members of the Epithelial-Mesenchymal Transition (EMT) gene set, and *F3* is also a member of the Inflammatory Response gene set. Two other members of the EMT gene set (*CTGF*\* (aka. *CCN2*) and *FOXC2*) also met the criteria for inclusion in Fig. 3A. Three of the above genes — *EGR1*\*, *JUN*, and *DUSP1*\*— were the 3 most discriminatory genes between IBC and non-IBC samples in a study using an RT-qPCR panel to compare expression of 538 genes in 36 IBC and 22 non-IBC frozen tumor samples (*29*). *EGR1* is a transcriptional regulator that is essential for T and B cell development and mediates an EGFR pathway-regulated immunosuppressive tumor microenvironment in IBC (*35*), while *DUSP1* promotes tumorigenesis in a number of cancers and regulates immune escape in cancer cells (*36*). *JUN* and *FOS* encode components of Activator Protein 1 (AP-1), a modulator of inflammatory responses in cancers and immune disorders (*37*), which also plays a role in hypertranscription in aggressive cancers (*38*).

Other significantly differentially expressed protein-coding genes that met the criteria for inclusion in Fig. 3A and are Hallmark gene set members were *RGS1\** (Inflammatory Response); *ZBTB16\** and *SLC6A14* (KRAS Signaling Down); *PIGR* (KRAS Signaling Up), a prognostic biomarker for hepatocellular carcinoma and correlated with poor prognosis in patients with osteosarcoma and pancreatic cancer; *FKBP5* (Estrogen and Androgen Response), over-expressed in a variety of cancers; *NEXN* (Apical Junction); and *TAT* (Xenobiotic Metabolism). Non-Hallmark gene set members that met the criteria for inclusion in Fig. 3A were *CSRNP1*, *COL4A6*, *SMIM3*, *STX1B* (a marker for breast tumors with neuroendocrine features), *SPATA18*, and *RGCC* (regulator of cell cycle, which plays a key role in mediating cancer-related inflammation in a wide variety of cancers).

To assess the degree of post-transcriptional regulation in IBC tumors, we compared DESeq2 expression statistics for Hallmark gene sets and genes (adj. *p*≤0.001 and LFC≥1) with TGIRT-seq reads mapped to the genomic reference sequence (Exons + Introns, *y*-axis) to those mapped to the transcriptome reference sequence (Exons only, *x*-axis; Fig. 3B and C). The Hallmark gene sets that were significantly enriched in differentially over-expressed (red) or under-expressed (blue) genes in IBC tumors were similar regardless of the reference sequence used for mapping, with TNFα-via- NFκB and EMT being the two gene sets most significantly enriched in over-expressed gene in IBC FFPE tumors. Eleven of the 26 protein-coding genes included in Fig. 3A were significantly over- expressed at adj. *p*≤0.001 and LFC≥1 for reads mapped to either the genome or transcriptome reference sequence, while the remaining 15 genes plus an additional gene (*AKAP13*, Hallmark Mitotic Spindle gene) were significantly over-expressed at adj. *p*≤0.001 and LFC≥1 only for reads mapped to the genome reference sequence (Fig. 3C, upper left section). Eight additional genes were significantly over-expressed at adj. *p*≤0.001 and LFC≥1 only for reads mapped to the transcriptome reference sequence (Fig. 3C, lower right section). These included *TMOD1*, driven by NFκB to enhance breast cancer growth; *MYO18B*, a candidate tumor suppressor gene in lung cancer; *CCNI2*, involved in cell-cycle regulation and promoting progression of colorectal cancer; *KCNK9*, whose hypomethylation is associated with apoptosis resistance of breast cancer; *GREM2*, a suppressor of breast cancer progression; and *CROCC2*, a tumor microenvironment-related prognostic biomarker for ovarian serous cancer (Table S2). Only subsets of these potentially interesting and clinically relevant differentially expressed genes would have been identified by mapping reads solely to either a genome or transcriptome reference sequence. Among protein-coding genes previously reported as having elevated RNA levels in IBC tumors, we found little or no difference in RNA expression levels for *COX-2* (aka *PTGS2*), *IL6\**, *IL8* (aka *CXCL8*), or the various *VEGF* genes for reads mapped to either the genome or transcriptome reference sequence (expression statistics in the Suppl. File).

To identify genes whose post-transcriptional regulation limits differential expression between IBC and non-IBC tumors, we plotted ΔIDRs (the ratio of IDRs between two compared genes) vs. DE test statistic values for reads mapped to the transcriptome reference sequence (Fig. S4). The plots showed that mRNA (Exons only) expression levels in IBC versus non-IBC tumors trended downward for higher ΔIDR genes, as expected for rate-limiting or aborted RNA splicing. After filtering genes with low overall expression (RPM<1), we identified 1,542 genes with high ΔIDRs (>2) that were not significantly differentially over-expressed (LFC<1) in IBC versus non-IBC tumors for reads mapped to the transcriptome reference sequence (Suppl. file). These genes included 1,105 Type I, 184 Type II, 94 Type IIIA, 19 Type IIIB, 9 Type IIIC, and 131 Type IV (IGV examples shown in Fig. S4D). High ΔIDR non-DE genes in these categories included 26 members of the Hallmark Inflammatory Response gene set, 36 members of the TNFα-via-NFκB, 28 members of the EMT gene set, 14 members of the Coagulation gene set, and 29 members of the Hypoxia gene set (Fig. S4, Table S3). The high ΔIDR non-DE genes in the Inflammatory Response gene set included those encoding pro-inflammatory cytokines (*CCL20*, *CXCL8*, *CXCL10*, *IL1B*) and a cytokine receptor (*IL18R1\**), possibly reflecting post-transcriptional regulatory mechanisms that modulate over-expression of deleterious inflammatory response genes (Table S3).

Other categories of significantly over-expressed RNAs in IBC vs non-IBC FFPE tumor samples that met the criteria for potential IBC biomarkers in Fig. 3A included 7 lncRNAs, 5 corresponding to an annotated mature lncRNA (AC006924.1, AC036176.3, AC044784.1, LINC02562, and SEPT7P6) and two having high read coverage across introns (LINC02015*, which predicts survival in glioblastoma multiform patients, and antisense RNA NFIA-AS2*, which correlates with poor prognosis in glioma; Fig. 3 and Fig. S3B); SNORD44, which has an antitumor effect when over-expressed in colorectal cancer cells; 5 simple repeat or satellite sequences annotated in RepBase (ATGAA)n, (ATTCC)n, (CTCT)n, (TGAGA)n, and (TCCATTCCAT)n; and LINE-1 element L1M3b, which has been reported to modulate chromatin assembly (*39, 40*) (Fig. 3A and Fig. S2). Two other snoRNAs, SNORA21 and SNORD123, which were among the key discriminative sncRNAs used in predicting eight different cancer types (*41*), were just below the stringent fold-change or FDR cutoffs for inclusion in Fig. 4A (SNORD123: adj. *p*=0.003, LFC = +6.8; SNORA21: adj. *p*=4.3e-6, LFC = -1.8). Despite the elevated expression levels of tRNAs in IBC vs non-IBC FFPE tumors (Fig. 1A), we found no significant differences in the relative abundance of each individual full-length tRNA isodecoder species, 5’- or 3’ halves, or 3’ tRFs between these samples at adj. *p*≤0.001, |LFC|≥2 (Fig. S2).

**Fig. 4.**
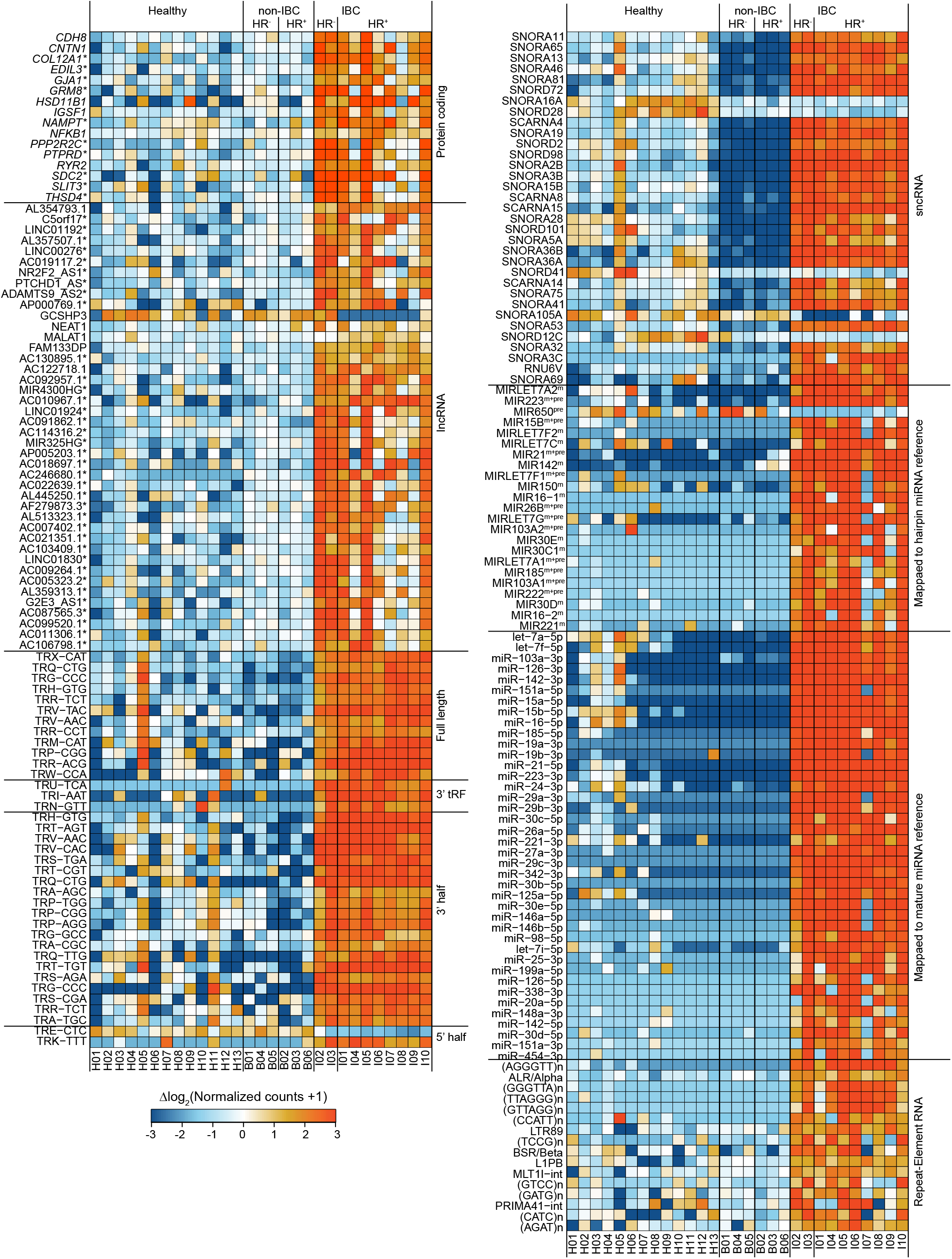
Heat map of expression levels of significantly differentially expressed genes in PBMCs. Tiles are colored by expression difference relative to the genewise mean taken across all PBMC samples, as in Fig. 3 above. The significantly over-expressed genes shown in the heat map (adj. *p*≤0.001, LFC≥2) are limited to those detected in half or more of the IBC samples. Lanes are labeled at the bottom according to healthy donor and patient ID numbers (Table S1). miRNAs mapping to the hairpin miRNA reference were categorized as: m, mature miRNAs, if reads were ≤2 nucleotides shorter or longer than the annotated mature miRNA sequence; pre, pre-miRNAs if reads were derived from pre-miRNA only, including relatively long RNAs that were derived from pre- miRNA but shorter than the annotated hairpin sequence; or m+pre, if reads were derived from both mature and pre-miRNAs. Examples of reads for different categories of miRNAs are shown in IGV alignments in Fig. S7E. Protein-coding and lncRNA genes with IDR>0.5 are marked with an asterisk.

Finally, we identified one mature miRNA (mir-125b-5p) as significantly upregulated in IBC versus non-IBC FFPE tumor samples at *p*≤0.001 and 5 others (let-7b-5p, mir-125a-5p, mir15b-5p, and mir-199a-5p, and mir-24-3p) upregulated but with a higher adj. *p*≤0.01 (Fig. 3A and Fig. S2). Although several of these miRNAs have cancer relevance (Table S2), none had been identified previously as upregulated in IBC tumors. Among the 12 miRNAs identified previously as upregulated in IBC (*42–45*), we found 10 to be nominally elevated (LFCs 0.05 to 3.32) either as mature miRNAs or as fragments derived from pre-miRNA hairpins but at adj. *p*>0.05, while miR- 520a-5p appeared to downregulated (adj. *p*=0.75, LFC= -0.95), and we did not detect miR-720 in our datasets (Suppl. file).

Comparisons between frozen healthy breast tissue and non-IBC tumor samples for the same set of IBC-focused genes in Fig. 3A showed mixed results, but with a trend toward lower expression levels of these genes in non-IBC tumor samples. These findings leave open the possibility that over-expression of some of genes in IBC vs non-IBC FFPE tumor samples may to some degree reflect under-expression in non-IBC tumors. Exceptions for which this was not the case included *RGS1\**, which was among the 10 most significantly elevated genes in the IBC vs non-IBC FFPE tumor comparison (LFC>3, adj. *p*<10^-6^ using either Exon + Intron or Exon only alignments) and *TNFAIP3*\*, *GEM*, *F3*, and *SMIM3*, which had similar expression levels between the paired frozen healthy breast tissue and non-IBC tumors samples (Fig. 3A).

### Differential gene expression in IBC PBMCs

TGIRT-seq with reads mapped to the human genome reference sequence identified 374 DE genes (149 protein-coding and 225 other) in IBC patient PBMCs versus both healthy donor and non-IBC patient PBMCs at adj. *p*≤0.001 and |LFC|≥2 (heat maps Fig. 4 and Fig. S5; Volcano plots Figs. S6 and S7) increasing to 807 genes at *p*≤0.001 and LFC≥1 (383 protein-coding and 424 other; Suppl. File). Scatter plots comparing Hallmark gene sets identified 5 that were that were significantly enriched (GO-MWU adj. *p*≤0.001) in up-regulated genes in IBC versus both non-IBC patient and healthy donor PBMCs with reads mapped to either the genome or transcriptome reference sequences: TNFα-via-NFκB, UV Response Down, Inflammatory Response, Spermatogenesis, and KRAS signaling down (red names and dots in Fig. 5A and B). Protein-coding genes that were significantly over-expressed at adj. *p*≤0.001, LFC≥1 in IBC patients compared to both healthy donor and non-IBC-patient PBMCs included 378 only for reads mapped to the genome reference sequence, only 5 for reads mapped to both the genome and transcriptome reference sequence (*COX7C, GOLGA4*, *NABP1*, *RGPD*1 and *RGPD2\**), and 48 only for reads mapped to the transcriptome reference sequence (Fig. 5C and D). These finding indicate pervasive rate-limiting RNA splicing in IBC patient PBMCs, possibly due to wide-spread enhanced transcription that results in saturation of the RNA splicing apparatus in addition to more specific post-transcriptional regulatory mechanisms.

**Fig. 5.**
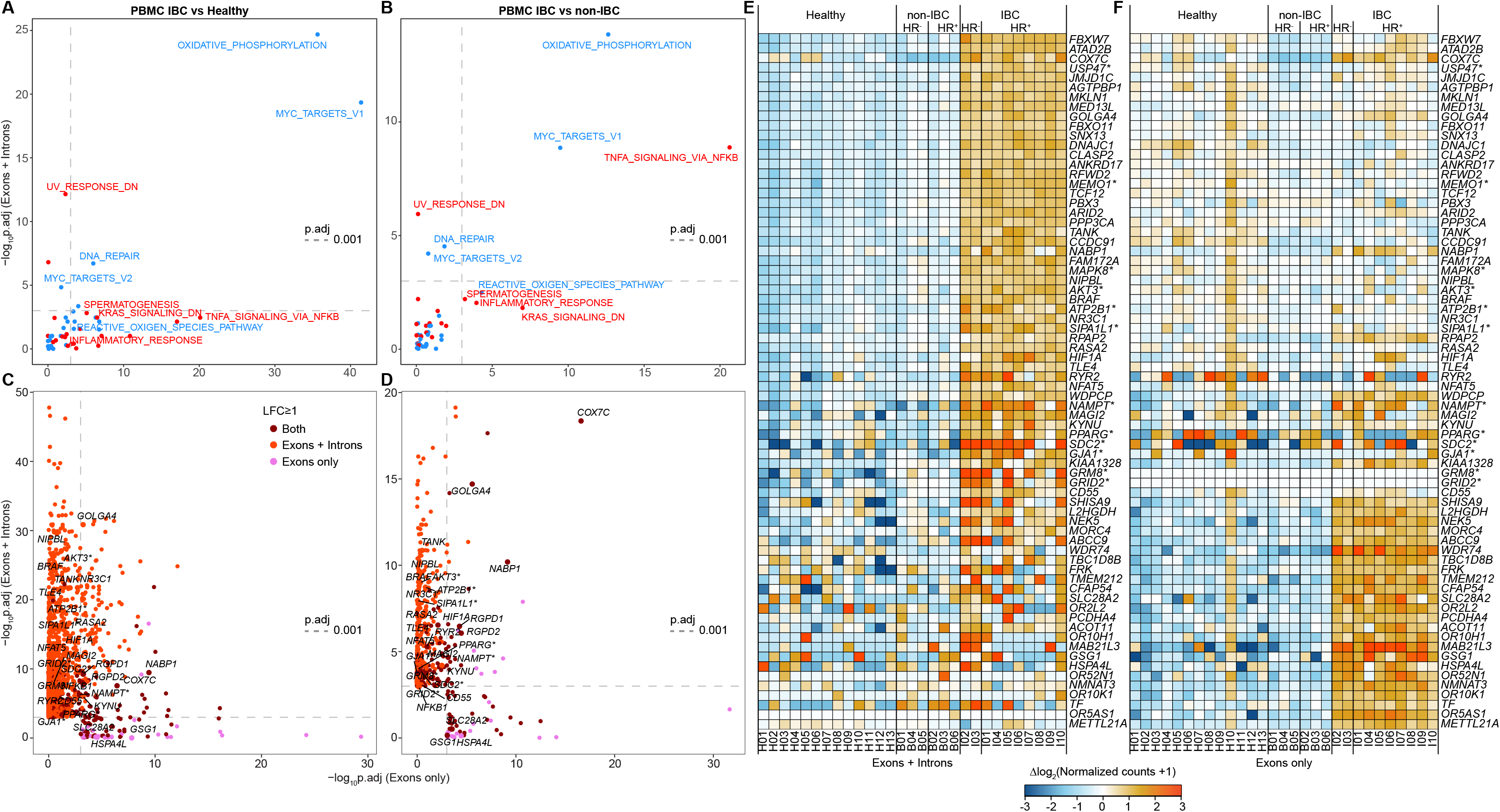
Comparison of PBMC differential expression for reads mapping to genome (Exons + Introns) versus transcriptome (Exons only) reference sequences. **(A)** Scatter plot comparing (log_10_-transformed and sign-inverted) FDR-adjusted *p*-values from GO-MWU IBC patient versus healthy donor analysis of BROAD Hallmark gene sets using gene counts generated by Exon only mapping reads (*x*-axis) or Exon + Intron mapping reads (*y*-axis). Gene sets for which the average gene MWU test statistic indicated increased expression in IBC are colored red, while those with decreased expression in IBC are colored blue. Vertical and horizontal dashed lines indicate adj. *p*=0.001. **(B)** Scatter plot formatted as in (A), but for IBC versus non-IBC patient PBMC comparison. **(C)** Scatter plot comparing (log_10_-transformed and sign-inverted) FDR-adjusted *p*- values for DE of most significantly IBC patient versus healthy donor PBMCs based on Exon only mapping reads (*x*-axis) or Exon + Intron mapping reads (*y*-axis). Genes name in plots are (i) those found in one of the 5 Hallmark gene sets enriched in up-regulated genes named in red in Fig. 5A and B and with LFC≥1 for which the adj. *p*-value <0.001 in the analysis based on at least one of the two mapping strategies (Exons only or Exons + Introns), and (ii) those with LFC≥1 for which the adj. *p*-value was <0.001 based on both of the two mapping strategies. **(D)** Scatter plot formatted as in (C) for an IBC versus non-IBC patient PBMC comparison. **(E)** Heat map showing expression profiles using Exon + Intron alignments of over-expressed protein-coding genes in IBC at adj. *p*≤0.001, LFC≥1 with reads mapped to the genome and/or the transcriptome reference sequences, which are either (1) members of one of the 5 Hallmark gene sets enriched in up-regulated genes named in red in Fig. 5A and B, or (2) among the top 50 over-expressed (LFC≥1) genes by *p*-value for each of the IBC vs. non-IBC and IBC vs. healthy comparisons in one or both mappings (Exons + Introns or Exons only). Tiles are color-coded as shown below the heat maps by expression difference relative to genewise mean taken across all PBMC samples, as in Fig. 3. Lanes are labeled at the bottom according to the healthy donor and patient ID numbers (Table S1). (**F**) Heat map formatted as in (E), but using Exon only instead of Exon + Intron mapping. Of the total 75 genes compared in Fig. 5E and F, only 3 (*COX7C*, *GOLGA4*, *NABP1*) were among the top 50 DE genes by *p*-value identified by both genome and transcriptome mapping. Protein-coding and lncRNA genes with IDR>0.5 are marked with an asterisk.

Similar comparisons for non-IBC patient versus healthy donor PBMCs for reads mapped to the genome or transcriptome reference sequences identified only 3 genes that were significantly over-expressed (adj. *p*≤0.001, LFC≥1) in non-IBC patient versus healthy donor PBMCs (*PRKCA*, *TSHZ2*, and *RPL28*), and 320 genes, a surprising large number, that were significantly under-expressed in non-IBC patient versus healthy donor PBMCs (adj. *p*≤0.001, LFC≤ -1; Fig. S8A). In contrast to IBC patient PBMCs (Fig. 5A), Hallmark gene sets that were significantly enriched in under-expressed genes in non-IBC patient versus healthy donor PBMCs (GO-MWU adj. *p*≤0.001) included Interferon Alpha Response, Interferon Gamma Response, and MTORC1 Signaling (Fig. S8B), suggesting an actively repressed PBMC response in non-IBC patient PBMCs.

Focusing on an IBC-specific PBMC response, members of the 5 Hallmark gene sets that were significantly enriched in up-regulated genes in IBC patient versus both non-IBC patient and healthy donor PBMCs at adj. *p*≤0.001 and LFC≥1 in one or both mappings (Exons + Introns or Exons only) included: TNFα-via-NFκB, *NAMPT\** (also over-expressed in PBMCs from patients with adult T-cell leukemia), *NFKB1\** (adj. *p*=0.0011, included in heat map Fig. 4A), *KYNU, NFAT5, TANK*, and *ATP2B1\** (also a member of the Hallmark Inflammatory Response and other gene sets and potentially significant for a granulocytic PBMC response in IBC; see below). In addition to *NAMPT\** and *ATP2B1\**, those in the Inflammatory Response gene set were *CD55, HIF1A*, and *SLC28A2*; those for the KRAS Signaling Down gene set were *RYR2* and *GRID2\**; those in the Spermatogenesis gene set were *GRM8\**, *GSG1, HSPA4L*, *TLE4,* and *BRAF* (a well-known cancer marker expressed at highest levels in B cells but also at higher levels in eosinophils and neutrophils); and those for UV Response Down gene were *SDC2*,* which encodes a transmembrane (Type I) heparan sulfate proteoglycan, a class of molecules involved at multiple steps in the progression of different cancers (*46*); *GJA1\**, also a member of the EMT and Estrogen Response Early gene sets, associated with recurrence, metastasis, and reduced survival in cancer; and *AKT3\**, *SIPA1L1*, NR3C1, PPARG*, NIPBL, RASA2, MAGI2,* and *ATP2B1\**. Scatter plots comparing the expression levels of genes belonging to the Hallmark TNFα-via-NFκB and Inflammatory Response gene sets between IBC tumors and PBMCs showed that *JUN* and *FOS,* which likely contribute to enhanced transcription, were similarly highly expressed in both samples sets for reads mapped to the genome or transcriptome reference sequences, but with other highly expressed members of these gene sets in IBC tumors outnumbering those in IBC PBMCs (Fig. S9).

Analysis of protein-coding gene expression data for PBMCs by CIBERSORTx (https://cibersortx.stanford.edu/), which estimates the relative abundance of different PBMC cell types from RNA-seq data, indicated IBC patient PBMCs had elevated levels of activated dendritic cells, which function in antigen presentation, and lower levels of resting mast cells, a class of granulocytes involved in immunoregulation, compared to both healthy donor and non-IBC patient PBMCs. CIBERSORTx profiling also indicated that eosinophils and neutrophils, granulocytes expected to be depleted by the centrifugation step during PBMC preparation (*47*) (see Methods), were enriched in multiple IBC patient samples compared to both non-IBC patient and healthy donor PBMC samples (box plots for cell types in Fig. S10, heat maps for a selection of CIBERSORTx genes in Fig. S11). The enrichment of these cell types in PBMC preparations from IBC patients likely reflects an increase in low-density, activated eosinophils and neutrophils, which contribute to inflammation and whose abundance can be modulated by cancer treatments (*48–50*).

Genes with high ΔIDRs (>2) included 2,316 that were not significantly over-expressed for reads mapped to the transcriptome reference sequence in IBC versus non-IBC patient and healthy donor PBMCs (LFC<1 in both comparisons), including 1,760 Type I, 188 Type II, 130 Type IIIA, 30 Type IIIB, 2 Type IIIC, and 206 Type IV (Fig. S12 and Suppl. File). These genes included 32 members of the Hallmark Inflammatory Response gene set, 26 members of the TNFα-via-NFκB gene set, 30 members of the EMT gene set, 14 members of the Coagulation gene set, and 21 members of the Interferon Gamma Response gene set, including *IL7\**, *IRF4*, and *STAT3* (Fig. S12 and Table S4). The heat maps of Fig. 5E and F identified 2 genes (*GRM8\** and *GRID2*\*) that were significantly over-expressed with reads mapped to Introns + Exons, but had zero reads that mapped to Exons, both found in IGV alignments to be long genes (815-1,507 kb) with a low depth of intron reads extending across the length of the gene (Fig. S7A).

Notably, heat maps for the top 50 DE genes (Fig. 5E and F) and CIBERSORTx marker panel genes (Fig. S11) with adj. *p*≤0.001, LFC≥1 showed that protein-coding gene reads mapped to the genome reference sequence, which quantifies both introns and exons, more clearly distinguished IBC patients from both healthy donor and non-IBC patient PBMCs than did those identified by mapping reads to the transcriptome reference sequence, which quantifies only exons. An example is healthy donor H10, whose protein-coding gene expression pattern was more similar to that of IBC patient than other healthy donors or non-IBC patient PBMCs for reads mapped to the transcriptome reference sequence (Fig. 5F), but more closely resembled other healthy donors for reads mapped to the genome reference sequence (Fig. 5E). These findings suggest an activated PBMC response in healthy donor H10 PBMCs that did not lead to rate-limiting RNA splicing or accumulation of excised intron RNAs, possibly reflecting a less saturated splicing apparatus in non-IBC than IBC patient PBMCs.

Other categories of significantly DE RNAs in IBC patient PBMCs versus both non-IBC patient and healthy donor PBMCs at adj. *p*≤0.001 and |LFC|≥2 included 56 lncRNAs; 52 over- expressed tRNA isodecoder families, tRNA halves, and 3’-tRFs fragments, potentially relevant to cell regulatory processes; 36 sncRNA including 28 snoRNAs (23 over-expressed and 5 under-expressed), potentially relevant to post-transcriptional regulation; 65 miRNAs (40 identified by mapping reads to the miRBase mature miRNA reference set and 25 identified by mapping reads to the miRBase hairpin reference set); and 16 Repeat-Element RNAs, including 12 simple repeats or satellite sequences, 3 LTR retroelement (LTR89, MLT1-int, and PRIMA41-int), and LINE-1 element L1PB RNAs (heat map Fig. 4, Volcano plots Fig. S6; IGV alignments for representative DE protein-coding gene, lncRNAs, sncRNAs, mature miRNAs and hairpin miRNAs in Fig. S7).

Notable differentially expressed RNAs in IBC patient PBMCs in Fig. 4A included: (i) lncRNAs AL354793.1, a biomarker for triple-negative breast cancer; LINC01192, associated with low survival of triple negative breast cancer; antisense RNA NR2F2-AS1, which promotes cancer cell proliferation in a number of different cancers; and pseudogene *GCSHP3*, which is under-expressed in gastric cancer tumors and in HR^-^ but not HR^+^ IBC patients; (ii) over-expressed SNORA11 and SNORA65, reported signatures of ameloblastoma; SNORA13, related to chemoresistance of osteosarcomas; and SNORD72, over-expressed in hepatocellular carcinoma tissue; and (iii) over-expressed miR-21, whose expression levels correlated with the histological grade of IBC (*51*) and miR-223 and miR-142, both identified as hematopoietic miRNAs capable of increasing the rate at which progenitor cells differentiate into T cells (*52*), with miR-223 identified as a potential diagnostic marker and therapeutic target for inflammatory disorders (*53*).

We found only a limited overall correlation of LFCs for significantly DE genes encoding different RNA biotypes between IBC PBMCs and tumors, indicating that significant gene expression changes in IBC tumors do not simply reflect the presence of tumor-infiltrating PBMCs (Fig. S13).

### Identification of potential IBC biomarkers in plasma

TGIRT-seq of the clinical plasma samples in this study gave datasets with reads mapping to as many as 22,000 different genes encompassing all RNA biotypes detected in tumors and PBMCs (Suppl. file). Potential RNA biomarkers identified in IBC plasma samples by DESeq2 (adj. *p*≤0.001 and LFC≥2) included 30 protein-coding gene RNAs, 6 lncRNAs, 1 sncRNA (RNY5), and 39 Repeat-Element RNAs, and those identified by peak calling (read coverage ≥10 reads at the peak maximum, *q*-value≤0.01), included 3 protein-coding gene RNAs, 1 lncRNA (a 165-nt RNA mapped to an intergenic region), and 64 Repeat-Element RNAs (heat maps Fig. 6A and B, Volcano plots Fig. S14, IGV alignments Fig. 6C-H and Fig. S15).

**Fig. 6.**
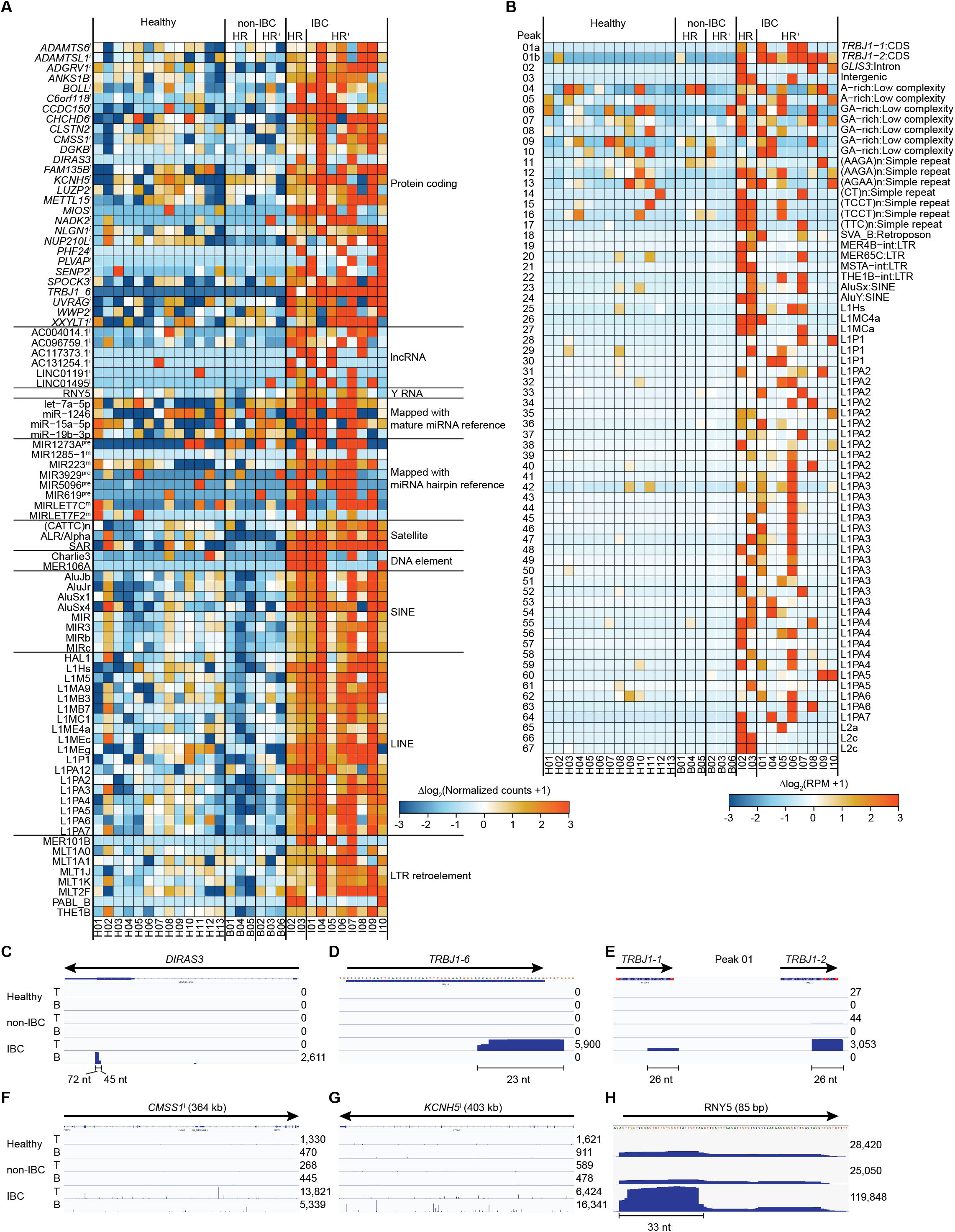
Heat map of the relative abundance and IGV plots for select genes in the plasma sample set. (**A**) Genes and features of interest, including those identified by DESeq2 analysis as significantly over-represented (adj. *p*≤0.001, LFC≥2) in the IBC plasma datasets. Tiles are colored by expression differences relative to the corresponding genewise means taken across all plasma samples. Color scale at bottom right of panel. miRNAs mapping to the hairpin miRNA reference were categorized as: m, mature miRNAs, if reads were 1 or 2 nucleotides shorter or longer than the annotated mature miRNA sequence; pre, pre-miRNAs if reads were derived from pre-miRNA only; or m+pre, if reads were derived from both mature and pre-miRNAs. i: protein-coding gene contains reads that are mostly derived from intronic regions. (**B**) Heat map for called peaks in the plasma sample set generated using MACS2. Tiles are colored by log_2_-transformed difference in RPM- normalized counts from the genewise mean across all plasma samples. Color scale at bottom right. Lanes are labeled at the bottom according to healthy donor and patient ID numbers (Table S1). (**C-H**) IGV plots for the indicated genes and peaks. Arrows for gene maps above the IGV plots indicate the 5’ to 3’ orientation of the gene, with the number of reads that mapped to the top (T) or bottom (B) strands indicated to the right of the plot for each strand.

Protein-coding gene RNAs that were significantly over-represented in IBC patient plasma included 4 corresponding to short, discrete, largely exonic RNA fragments, possibly stabilized in plasma by bound proteins (Fig. 6A, C-E). For one gene, *DIRAS3*, a tumor suppressor gene whose normally silenced maternal allele can be re-activated to induce autophagy in breast cancer cells (*54*), the over-represented species were 72- and 45-nt RNA fragments derived from the 3’ end of the coding sequence and 3’ UTR, respectively (Fig. 6A and C). The other 3 over-represented exon-containing RNAs were 23- or 26-nt RNA fragments of 3 different T-cell receptor beta one joining regions (*TRBJ1*-*1*, -*2*, and -*6*; Fig. 6A, B, D, and E). The *TRBJ1-6* RNAs were detected in all 10 of the IBC patient samples (highly abundant in 9, moderately abundant in 1, and not detected in any of the healthy or non-IBC patient samples; Fig. 6A). *TRBJ1-1* RNAs were abundant in 4 of the IBC patient samples and not detected in any of the healthy or non-IBC patient samples, and *TRBJ1-2* RNAs were found in all 10 of the IBC patient samples (highly abundant in 7, moderately abundant in 3) and detectable at lower abundance in one each of the healthy and non-IBC samples (Fig. 6B).

The remaining 26 protein-coding gene RNAs that were significantly over-represented in IBC patient plasma corresponded to 50- to 80-nt intron RNA fragments (denoted with superscript "i" in Fig. 6A; IGV examples in Fig. 6F, G and Fig. S15A). Intron RNA fragments were also the over-represented RNA species for all 6 lncRNAs that were significantly over-represented in IBC plasma (Fig. 6A, IGV plots in Fig. 15B). Some of the over-represented intron RNAs were highly specific for IBC plasma (e.g., *PHF24* and *PLVAP,* AC117373.1), while others were found at lower levels in healthy donor and/or non-IBC patient plasma (Fig. 6A).

For RNY5, the over-represented RNAs in IBC plasma were 5’ fragments of 28-33 nt (Fig. 6H), similar to those that were found to be enriched in cancer cell extracellular vesicles and cause rapid death of human primary cells but not cancer cells, possibly a mechanism by which cancer cells establish a favorable environment for proliferation (*55*).

A number of mature miRNAs (let-7a-5p, let-7c-5p, let-7f2-5p, miR-15a-5p, miR-1246-5p, miR-19b-3p and miR-223-3p) and partially or aberrantly processed miRNA transcripts (mir-619, mir-1273a, mir-1285-1, mir-3929, and mir-5096) detected in 5 or more IBC samples were over- represented (LFC≥2) in IBC compared to healthy and non-IBC samples, but fell short of statistical significance, likely due to low read counts and variable quality of the plasma samples (Fig. 6A). No tRNAs, tRNA halves or 3’ tRFs were found to be significantly over-represented in IBC plasma at adj. *p*≤0.001 and |LFC|≥2 compared to non-IBC and healthy plasma (Fig. S14A-C).

The 39 Repeat-Element RNAs identified as significantly over-represented in IBC patient plasma by DESeq2 included 18 LINEs, 8 SINEs, 8 LTR-containing retroelements, 2 DNA transposons, and 3 satellite sequences (Fig. 6A), while the 64 over-represented Repeat-Element RNA fragments identified by peak calling in IBC patient plasma ranged from 151 to 478 nt and included 43 mapping to LINE elements, 5 to LTR-containing retroelements, 2 to SINEs, and the remainder RepBase-annotated low complexity sequences, simple repeats and satellite sequences (Fig. 6B). Notably, all called peaks derived from LINE-1 elements corresponded to antisense RNAs (Suppl. file).

Reciprocally, we identified numerous genes whose transcripts were significantly over- represented and present in at least half of the healthy and non-IBC patient plasma samples, but sparsely represented in IBC patient plasma samples (heat maps Fig. 7A and B, Volcano plots Fig. S14A-C, IGV examples in Fig. 7D and Fig. S15). These included 125 protein-coding gene RNAs; lncRNA *SNHG6*, suggested to promote cancer development by miRNA sponging; 8 sncRNAs (7 snoRNA and snRNA U4ATAC); the 5’ half of AlaCGC and 3’-tRFs of AsnGTT and ArgTCG; 5 mature miRNAs (mir-16-1-5p, mir-30d-5p, mir-199a-5p, mir-101-3p, and mir-142-3p); 4 simple repeats (CCTCTTn, TCCTCCn, TTCTCTCn, and CTTCTCTTn); and 2 LTR retroelement RNAs (HERV1-LTRc and LTR12D) (Fig. 7A and B). Three other protein-coding gene RNAs (*ICAM3*, *RHOC,* and *TRA2B*) and one lncRNA (FAM212B-AS1) were found to be over-represented in non-IBC patient plasma compared to both healthy and IBC patient plasma (Fig. 7C; IGVs Fig. S15A and B).

**Fig. 7.**
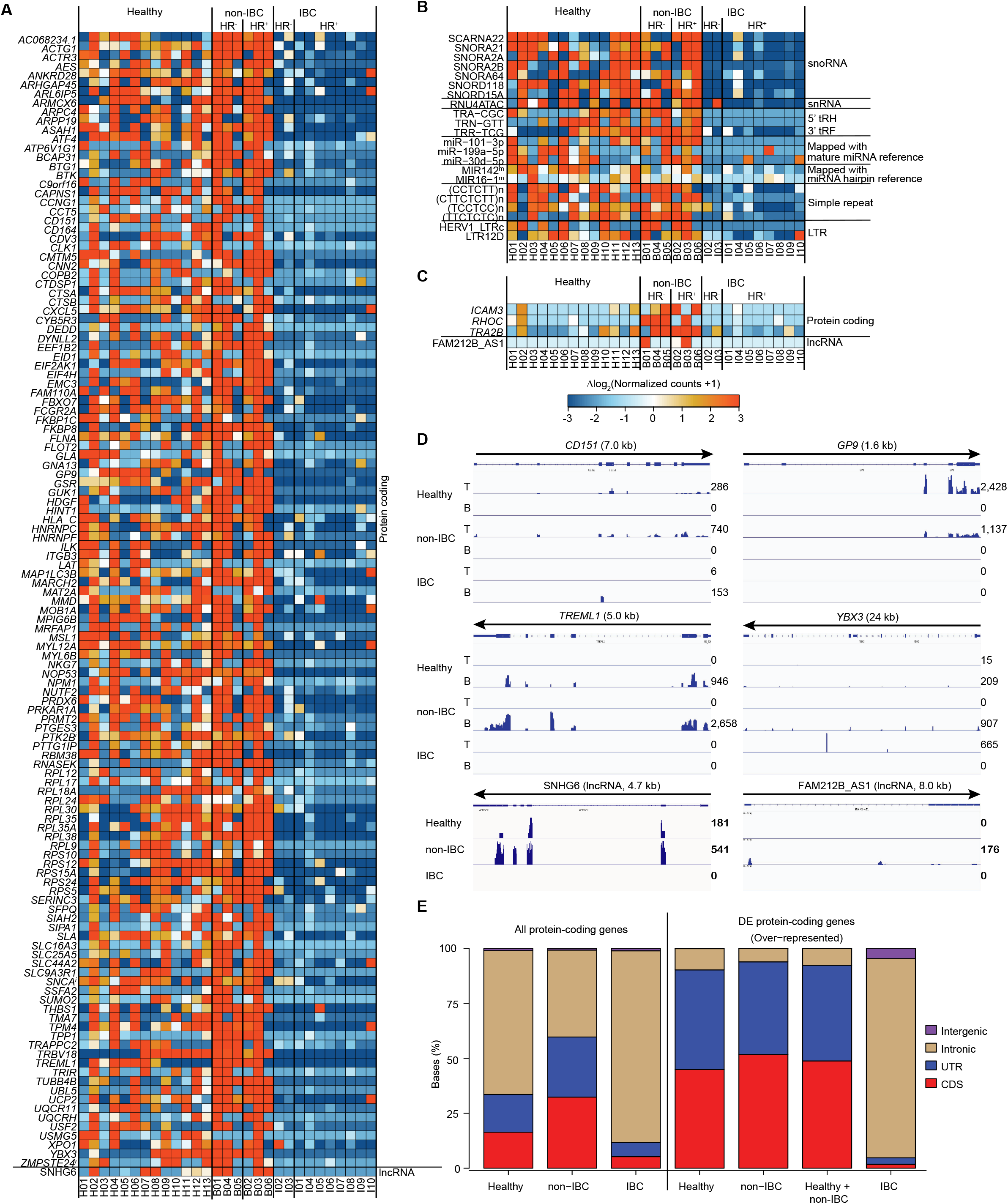
Heat map of the relative abundance for genes and features that were significantly over-represented in healthy and/or non-IBC plasma samples relative to the IBC plasma samples. (**A-C**) Heat maps for the indicated sample and RNA comparisons. Tiles are colored by expression differences relative to the corresponding genewise means taken across all plasma samples. Lanes are labeled at the bottom according to the healthy donor and patient ID numbers (Table S1). miRNAs mapping to the hairpin miRNA reference were categorized as: m, mature miRNAs, if reads were 1 or 2 nucleotides shorter or longer than the annotated mature miRNA sequence. (**D**) IGV plots of over-represented genes in healthy or non-IBC plasma compared to IBC plasma. T and B, top and bottom DNA strands, respectively. The number of reads for RNAs in each IGV plot is indicated to the right. (**E**) Stacked bar graphs showing the percentage of bases in reads mapped to coding sequences (CDS), untranslated regions (UTRs), introns, or intergenic regions for all protein- coding genes detected in the indicated plasma samples (left 3 bars) or for significantly over- represented protein-coding genes (adj. *p*≤0.001, LFC≥2) detected in ≥ 50% of the indicated plasma samples (right 4 bars).

In contrast to IBC patient plasma, where the over-represented protein-coding gene RNAs were largely intronic, the over-represented RNAs for protein-coding genes in healthy and non-IBC plasma mapped largely to exons, indicating that they likely correspond to fragments of mRNAs (IGV examples in Fig. 7D and Fig S15A). This stark difference between IBC and non-IBC plasma samples is illustrated by the finding that although introns comprised substantial proportions of the bases in reads mapping to protein-coding genes in plasma samples regardless of disease state (healthy, 65%; non-IBC, 40%; and IBC, 87%), they comprised >91% of the bases in reads from over-represented protein-coding genes in IBC plasma compared to only 10 and 6% for over- represented protein-coding genes in healthy and non-IBC plasma samples, respectively (Fig. 7E).

### Cellular origin of *TRBJ1* RNAs in IBC patient plasma

The cellular origin of the 3 *TRBJ1* RNA fragments identified above as potential plasma RNA biomarkers for IBC was revealed by IGV alignments (Fig. 8A), which showed peaks corresponding to the same *TRBJ1-2* and -*6* RNAs and to a lesser extent *TRBJ1-1* and *-5* RNAs rising above a background of reads spanning these regions in IBC tumor and PBMCs. Despite the presence of RNAs mapping across the *TRBJ1* locus in the cellular samples, the reads detected in plasma were short with relatively discrete lengths, suggesting they corresponded to RNA fragments protected from plasma RNase by structural features or bound proteins (*3*).

**Fig. 8.**
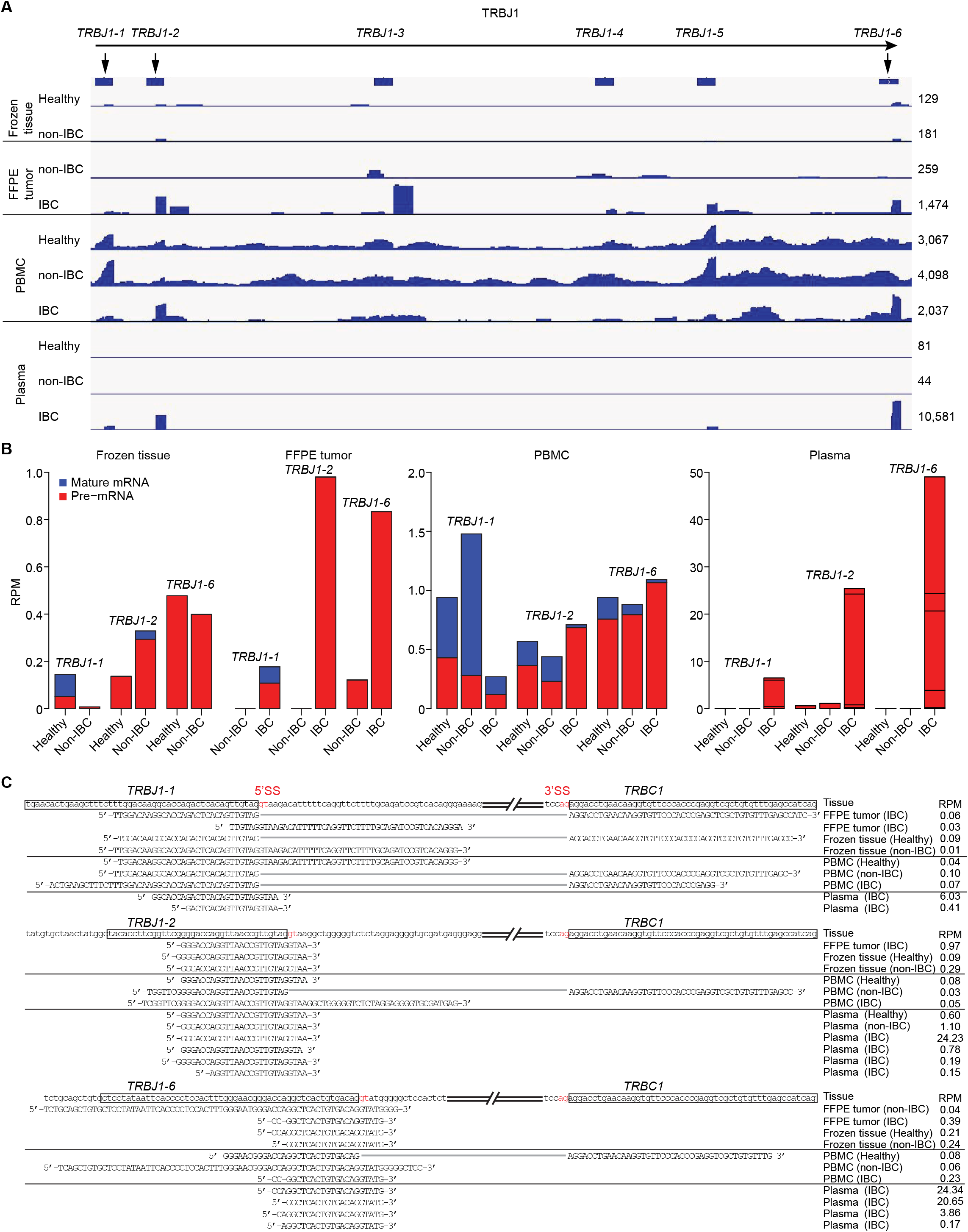
Cellular origin of TRBJ1 RNAs that were over-represented in IBC plasma samples. (**A**) IGV plots for RNAs mapping to genomic locus encoding *TRBJ1-1* through *TRBJ1-6* in combined datasets for different samples. The number of reads for RNAs in each IGV plot is indicated to the right. (**B**) Stacked bar graph showing RPM for spliced mRNAs (blue) and unspliced pre-mRNAs (red) for *TRBJ1-1*, *TRBJ1-2*, and *TRBJ1-6* in different sample types. Sequences corresponding to the splice site for *TRBJ1-1*, *TRBJ1-2*, or *TRBJ1-6* to *TRBC1* were extracted and categorized by sample class, sample type, and RNA type (unspliced pre-mRNA or spliced mRNA). Horizontal thin black lines in the bar graphs for plasma delineate unspliced pre-mRNA reads with different sequences. (**C**) Sequence alignments for the reads encompassing the splice-site (SS) region of *TRBJ1-1*, *TRBJ1-2*, or *TRBJ1-6* to *TRBC1* (5’SS, 3’SS, and intron 5’ GT and 3’ AG in red). The displayed reads are representative abundant RNA fragments from each category for samples in the same disease category and tissue type. Sequences of all unique RNA fragments detected in plasma datasets with >5 reads are shown. Reads containing spliced exons are connected by gray lines.

Closer scrutiny revealed that all of the exonic *TRBJ1* reads from plasma samples, regardless of disease state, extended a short distance into a downstream intron, while those in the cellular samples were a mixture of exon-intron and spliced mRNA reads in which the different *TRBJ1* segments were spliced to the downstream *TRBC1* constant region (Fig. 8B and C). Among the cellular RNA samples, the preponderance of *TRBJ1-2* and *-6* reads that extended into the intron was highest in IBC FFPE tumors (Fig. 8B), where they may have originated either from unspliced pre-mRNA in tumor infiltrating T cells or from aberrant transcription of the *TRBJ1-n* locus in non-T-cells (Fig. 8B and C). Regardless of their origin, the *TRBJ1-2* and *-6* RNAs were prevalent in IBC patient plasma samples but sparse or undetectable in non-IBC patient or healthy donor plasma samples, making them strong candidates for plasma RNA biomarkers for IBC.

### Cellular origin of exon and intron RNA fragments in plasma

To investigate the cellular origin of intron RNAs in plasma, we plotted log_2_-normalized read counts for RNAs mapped to Exons + Introns, Exons only, or Introns only for each protein-coding gene in plasma versus those in FFPE tumors and PBMCs (Fig. 9A). Dots and trend lines for genes with different IDRs are color-coded as shown in the Figure with trend lines for genes ≤100 kb and >100 kb indicated by solid and dashed lines, respectively. In all cases, the levels of both Exon and Intron RNAs in plasma trended higher for genes with higher log_2_-transformed normalized expression levels in tumors or PBMCs up to ∼7.5, the range found for most genes, but with the plots for Exon + Intron RNAs (top row) more closely resembling those for Intron RNAs (bottom row) than those for Exons RNAs (middle row), reflecting preferential enrichment of intron RNAs in plasma regardless of sample type or disease state (*2, 3*). All of the plots also showed preferential enrichment in plasma of Intron RNAs from long genes with high IDRs (>1) (red dots and dashed lines). Notably, the trends for long genes with high IDRs were more pronounced for intron RNAs from IBC tumors and PBMCs than for non-IBC patient or healthy donor samples, potentially reflecting wide-spread enhanced transcription in IBC tumors and PBMCs that results in rate-limiting or aborted RNA splicing and leads to accumulation of intronic RNAs that are preferentially exported and end up in plasma.

**Fig. 9.**
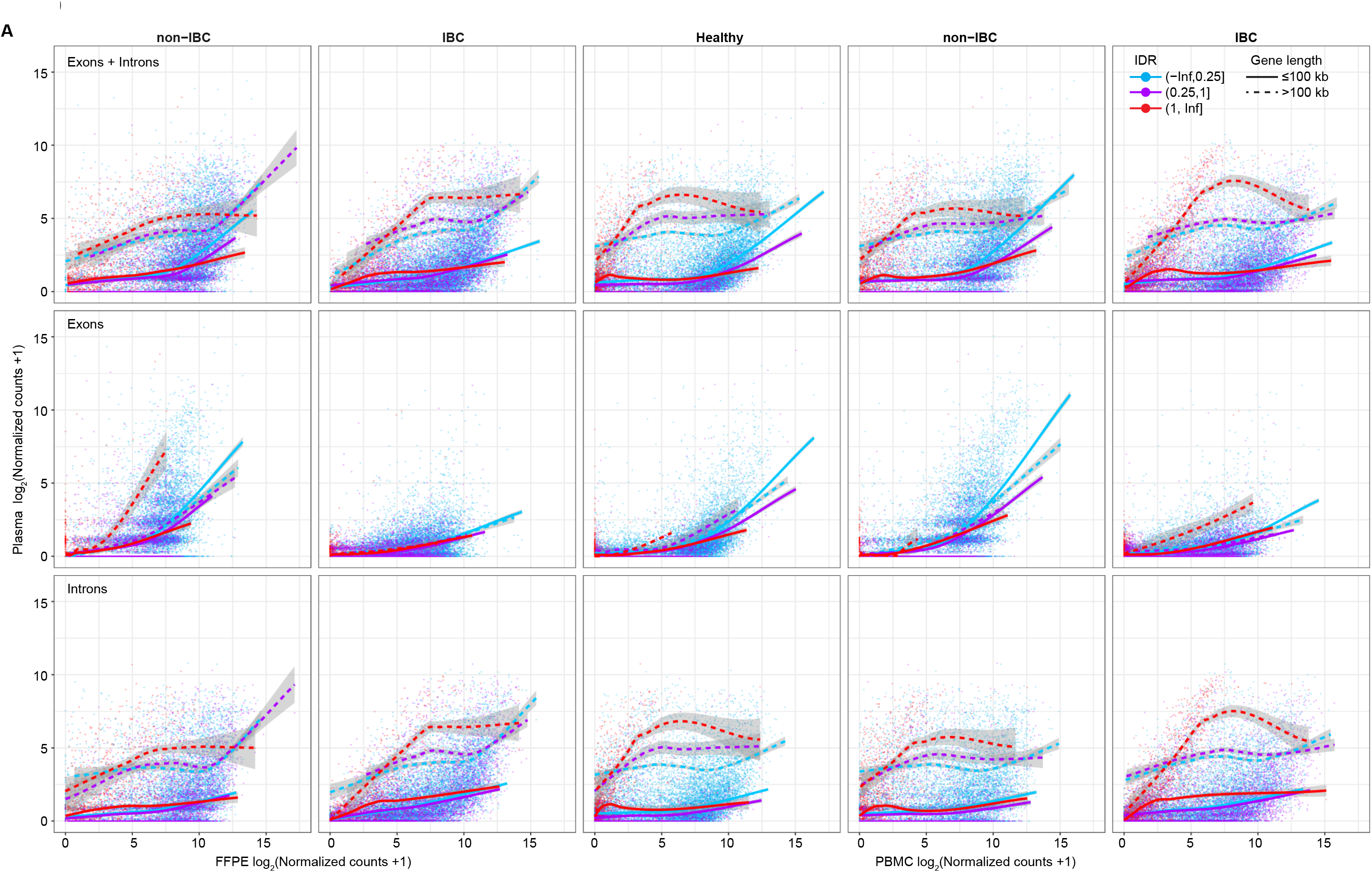
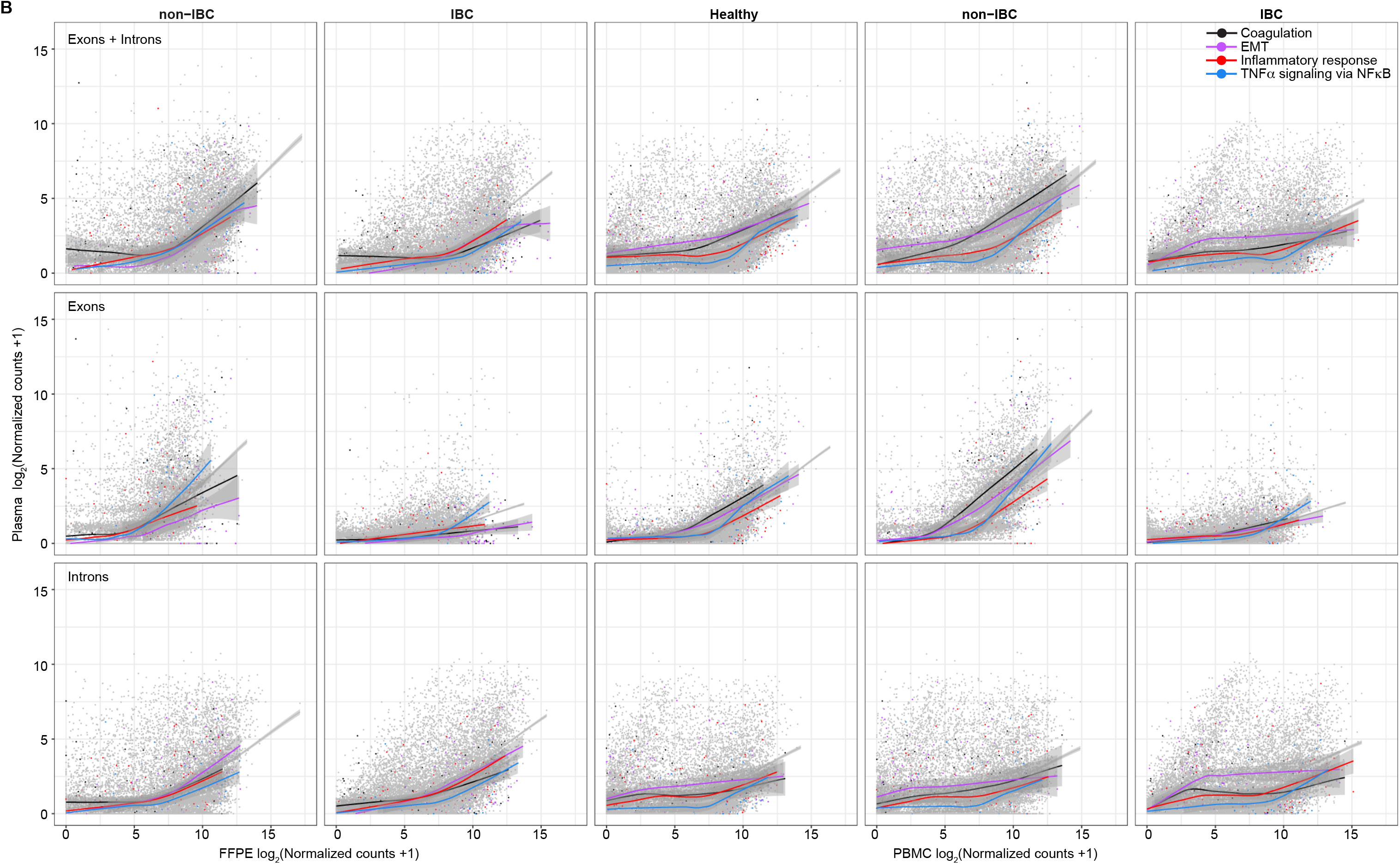
Cellular origin of intron RNAs that are over-represented in IBC plasma samples. **(A)** Average abundance based on DESeq2 normalized counts for each gene whose RNAs were detected in plasma samples plotted against its average normalized expression level for RNAs in non-IBC or IBC patient FFPE tumors samples (left two columns) or Healthy donor, non-IBC patient, or IBC patient PBMC samples (right three columns) for reads mapped to Exons + Introns (top row), Exons only (middle row), or Introns only (bottom row). Each point represents a single gene, colored according to its binned IDR values in FFPE tumor or PBMC samples, with color codes shown on the top right plot. Generalized additive model trendlines are fit for each group of IDR-binned genes, with 95% confidence intervals based on standard error estimates indicated by the ribbons around the trendlines. The trendlines show separation of gene sets with different IDRs and particularly for the region around 5 on the *x*-axis in each panel, with high-IDR genes (red points and trendlines) tending to show higher average plasma enrichment than lower-IDR genes with similar FFPE tumor or PBMC expression levels. Solid lines represent trends calculated from genes ≤100 kb and dashed lines for genes >100 kb. **(B)** The same comparisons as in panel A with points colored according to selected Hallmark gene sets with color codes shown on the top right plot. Generalized additive model trendlines are fit for each group of IDR-binned genes, with 95% confidence intervals based on standard error estimates indicated by the ribbons around the trendlines.

Fig. 9B shows the same plots with dots and color-coded trend lines for genes in 4 Hallmark gene sets (Coagulation, EMT, Inflammatory response and TNFα-via-NFκB). The plots for IBC patient plasma samples (columns 2 and 5) showed largely upward trends for Exon + Intron RNAs from all 4 pathways (top row) that again more closely resembled those for expression levels of Intron RNAs (bottom row) than Exon RNAs (middle row) in IBC tumors and PBMCs. By contrast, the plots for Exon + Intron RNAs in healthy donor and non-IBC patient plasma (columns 1, 3 and 4) more closely resembled those for Exon than Intron RNAs from these pathways in non-IBC patient tumors and PBMCs, likely reflecting the lower degree of post-transcriptional regulation in non-IBC patient tumors and PBMCs (see above). The plots for non-IBC patient plasma samples showed more pronounced upward trends for Exon RNAs from the TNFα-via-NFκB gene set in non-IBC patent tumors (column 1) and Exon RNAs from the Coagulation gene set in non-IBC patient PBMCs (column 4) than did those in IBC patient or healthy donor plasma (columns 2, 3, and 5; Fig. 9B).

For the 26 protein-coding genes identified by DESeq2 as having over-represented intron RNAs in IBC patient plasma (Fig. 6A), we identified a total of 2,481 intron fragments (where overlapping fragments were counted as one fragment). These included 25 intron RNA fragments from 12 genes that were over-represented in IBC patient plasma and found in at least half of those samples making them candidates for IBC biomarkers (Fig. S16). All of these intron RNAs were from relatively long (60 to 1,004 kb) genes with low expression levels and relatively high proportions of antisense-strand reads (11 to 89%) in IBC tumors and PBMCs (Fig. S16), likely reflecting activation of antisense promoters and/or preferential enrichment of antisense RNAs in plasma (Fig. 1E). Collectively, these findings show that RNAs in plasma samples mirror gene expression differences in tumors and PBMCs with exon RNAs from more highly expressed gene enriched in non-IBC-patient plasma and intron RNAs from high IDR genes enriched in IBC patient plasma, a difference likely reflecting widespread enhanced transcription and rate-limiting RNA splicing in IBC tumors and PBMCs.

### Over-expression of LINE and other retroelements in IBC

Finally, to investigate if the prevalence of differentially represented LINE and other Repeat-Element RNAs in IBC plasma (Fig. 6) might also reflect widespread over-expression in IBC patient tumors and PBMCs, we compared aggregated reads mapping to multiple loci of each Repeat-Element family in IBC patient versus non-IBC patient and healthy donor samples. The resulting scatter plots showed over-expression of the known active human LINE-1 element L1Hs (*56*) and normally silenced primate-specific LINE-1s (L1PA2-17), LINE-2s, SINEs, endogenous retroviruses and other LTR-containing retroelements, and to a lesser extent DNA transposons and other Repeat-Element RNAs (Fig. 10A and Fig. S17). The higher abundance of LINE and other Repeat-Element RNAs in IBC tumors and PBMCs was magnified in plasma, suggesting their preferential export or higher stability in plasma compared to other RNA biotypes (Fig. 10A and Fig. S17). By contrast, pairwise comparisons showed no similar enrichment of LINE and other Repeat-Element RNAs in frozen non-IBC tumor samples versus matched healthy breast tissue samples, nor between non-IBC patient versus healthy donor PBMC or plasma samples (Fig. 10A and Fig. S17).

**Fig. 10.**
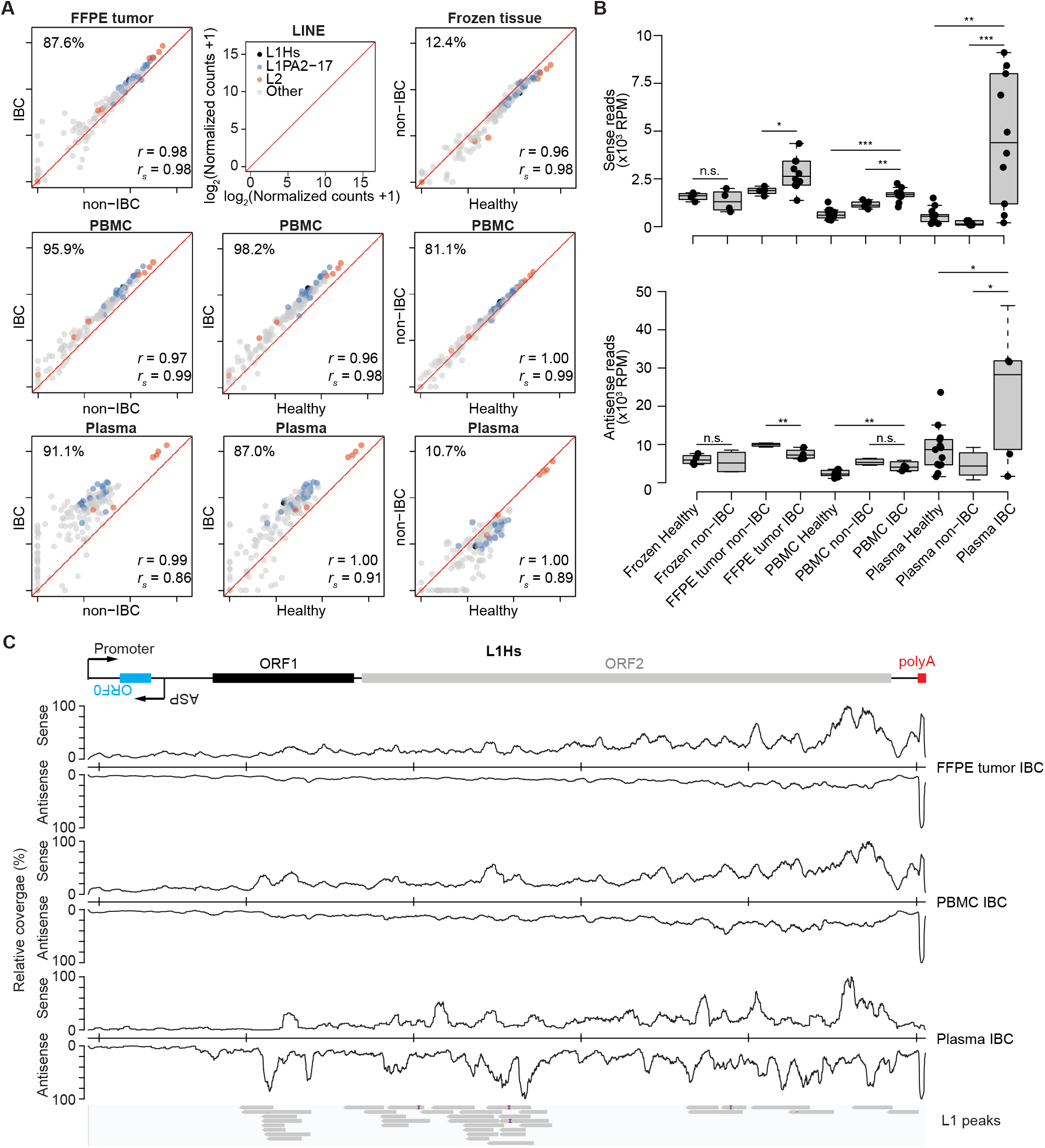
Activation of LINE-1 RNAs in IBC tumor and PBMCs and nature of LINE-1 RNA reads in plasma. (**A**) Scatter plots comparing the relative abundance of different families of LINE RNAs color coded as indicated in the top center panel detected in breast tissue, PBMC or plasma samples. DESeq2 normalized counts of LINE elements were averaged by tissue type and disease status and log_2_-transformed for pairwise comparisons. The number at the top left of each plot indicates the percentage of LINE RNAs above the diagonal, a measure of over-expression of LINE elements in each comparison. Pearson’s (*r*) and Spearman’s (*r_s_*) correlation coefficients are shown at the bottom right. (**B**) Box plots comparing the abundance (RPM) of reads remapped to the L1Hs consensus sequences of the sense and antisense strands in different tissue types. Wilcoxon tests were used to compare different groups of samples *: *p*≤0.05; **: *p*≤0.01; ***: *p*≤0.001, n.s. not significant. (**C**) Coverage plots of reads mapped to the L1Hs consensus sequence in combined dataset from different tissues. Coverage depths were normalized to the maximum coverage of the sense or antisense reads in each sample type. Abbreviation: ASP, LINE-1 antisense promoter.

To explore the origin of LINE-1 RNA reads, we focused on the active human LINE-1 element L1Hs, which is over-expressed in some aggressive cancers (*56*), by remapping all L1 reads to the L1Hs consensus sequence. Consistent with the activation of L1Hs in IBC tumors and PBMCS, we found that levels and ratios of L1Hs sense strand to antisense strand reads were significantly higher than in non-IBC patient tumors or PBMCs (Fig. 10B and C). L1Hs reads were even more highly enriched in IBC plasma samples, but with a higher ratio of antisense- to sense-strand reads (Fig. 10B) and with peaks of antisense-strand reads correlated with differentially represented LINE-1 RNA fragments identified by peak calling (Fig. 10C), likely reflecting read through transcription resulting from transcriptional activation of neighboring genes.

## Discussion

TGIRT-seq of FFPE tumor samples, PBMCs, and plasma RNAs revealed numerous striking differences in all 3 sample types that distinguished IBC from non-IBC patients across all tested HR and HER2 subtypes. By mapping TGIRT-seq reads to both genome (Exons + Introns) and transcriptome (Exons only) reference sequences and analyzing Intron-exon Depth Ratios (IDRs), we developed methods for parallel analysis of transcriptional and post-transcriptional gene regulation from TGIRT-seq datasets. This analysis showed wide variations in IDRs for different genes in all cell types, reflecting differences in the relative rates of transcription, RNA splicing, and intron or mRNA turnover. Many high ΔIDR genes in IBC versus non-IBC tumors remained DE for reads mapped to the transcriptome but with expression statistics trending downward for higher ΔIDR genes, reflecting rate-limiting RNA splicing (Fig. S4). We also identified subsets of high ΔIDR genes in IBC versus non-IBC tumors and IBC versus healthy donor and non-IBC PBMCs whose mRNAs were not differentially expressed, reflecting activated transcription combined with rate-limiting or aborted RNA splicing (Fig. S4 and S12). The differences in post-transcriptional regulation were particularly pronounced in IBC patient PBMC versus non-IBC patient and healthy donor PBMCs for which we found only 53 genes that were significantly elevated at adj. *p*≤0.001 and LFC≥1 for reads mapped to the transcriptome reference sequence compared to 383 genes for reads mapped to the genome reference sequence (Figs. 4, 5, and S5).

TGIRT-seq of IBC versus non-IBC tumor FFPE tumor samples RNAs showed over- expression of key members of the Hallmark TNFα-via-NFκB and EMT gene sets, including some found previously to be over-expressed in IBC patient (*30, 33, 57, 58*). These included JUN and FOS, which dimerize to form transcription factor AP-1 that functions in conjunction with other proteins to promote hypertranscription and inflammation-induced cancer progression in aggressive cancers (*37, 38, 59–61*). Activation of PBMCs in IBC patients was also correlated with over- expression of JUN, FOS, and other TNFα-via-NFκB and inflammatory response genes, albeit different subsets than in IBC tumors (Fig. S9).

CIBERSORTx analysis suggested that IBC patient PBMCs had elevated levels of granulocytic eosinophils and neutrophils compared to those from healthy donor and non-IBC patients (Fig. S10). Beyond their well-documented role in parasitic infection control, eosinophils play a pivotal role in anti-tumor responses (*62*). *ATP2B1\**, which encodes an ion transport ATPase that functions in granule secretion in granulocytes (*63*), stood out as being over-expressed in all IBC patient PBMC samples (adj. *p*<10^-8^ and LFC>1.5) compared to both healthy donor and non-IBC patient PBMC samples for reads mapped to the genomic reference sequence (Fig. 5 and Fig. S11), making it a promising biomarker for an immune response in IBC.

The larger numbers of significantly DE genes in both IBC tumors and PBMCs for reads mapped to a genome reference sequence compared to those for reads mapped to a transcriptome reference sequence suggests wide-spread enhanced transcription that results in rate-limiting RNA splicing and leads to the accumulation of unspliced pre-mRNAs for large subsets of genes in both sample types. Wide-spread enhanced transcription in IBC tumors and PBMCs was also indicated by over-expression of LINE-1 and other Repeat-Element RNAs (Fig. 10 and Fig. S17). These findings, together with the over-expression of AP1-components JUN and FOS, suggest that wide-spread enhanced transcription in IBC tumors and possibly also in PBMCs may be related to hypertranscription, a characteristic of aggressive cancers attributed to DNA hypomethylation and associated changes in chromatin structure that activate transcription of topologically linked genes, including LINE-1 and s (*64, 65*). The activation of LINE-1 elements in IBC tumors and PBMCs indicated by higher expression of L1Hs sense strand than antisense strand RNAs (Fig. 10) could also contribute to activation of distant genes (*66*) as well as to inflammatory responses and interferon production in IBC via RIG-I or MDA5 sensing of double-strand RNAs or cGAS/STING sensing of LINE-1 cDNA and/or RNA/cDNA duplexes (*67–69*).

IDR analysis also provided insights into post-transcriptional regulatory mechanisms that are potentially activated by wide-spread enhanced transcription in IBC patient tumors and PBMCs.

Thus, while many high ΔIDR genes in IBC versus non-IBC tumors an PBMCs remained DE for reads mapped to the transcriptome reference sequence, mRNA expression statistics for higher ΔIDR genes trended downward, reflecting rate-limiting or aborted RNA splicing (Figs. S4 and S12).

Additionally, we identified subsets of transcriptionally active high ΔIDR genes in IBC versus non-IBC tumors and PBMCs whose mRNAs quantified by reads mapping to transcriptome reference sequences were not differentially expressed due to post-transcriptional mechanisms (Fig. S4 and S12). These included subsets of transcriptionally activated inflammatory cytokines response and inflammation-driven cancer progression genes in IBC tumors, whose over-expression might be deleterious, reflecting regulatory mechanisms that would be missed solely by quantitating mRNAs. Wider implications of these findings include that RNA-seq of chemically fragmented RNAs with reads mapped solely to a genome reference can over-estimate levels of mature mRNA due to the inclusion of intron and antisense RNA reads unless they are selectively removed, while RNA-seq of polyadenylated mRNAs or chemically fragmented RNAs with reads mapped solely to a transcriptome reference sequence misses highly informative insights about post-transcriptional gene regulation.

Consistent with previous TGIRT-seq analysis of human plasma from healthy individuals (*2, 3*), we found that intron RNAs contributed higher proportions of protein-coding gene bases in plasma than in cellular RNA samples irrespective of disease state (Fig. 7E). However, while most of the abundant differentially represented protein-coding gene RNAs in plasma samples from IBC patients were fragments of unspliced pre-mRNAs or intron RNAs, most of differentially represented protein-coding gene RNAs in plasma samples from healthy and non-IBC patients were mRNA fragments with reads mapping to exons (Fig. 7E). Plots of read depth in cellular versus plasma samples showed that enrichment of differentially represented intron RNAs in IBC patient plasma was correlated with the expression levels of intron RNAs in IBC tumors and PBMCs, while enrichment of differentially represented Exon RNA fragments in non-IBC patient or healthy donor plasma was correlated with expression levels of mRNA exons in non-IBC tumors or PBMCs (Fig. 9). Since plasma contains highly active RNases, plasma RNAs are expected to turnover rapidly, providing higher time resolution than plasma DNAs.

Among the many DE RNAs in IBC, prime candidates for blood-based IBC biomarkers included 3 different sets of *TRBJ1* exon-intron RNA fragments, possibly related to aberrant enhanced transcription and/or induction of a specific immune response in IBC. All 3 *TRBJ1* RNAs could be traced to tumors and PBMCs in IBC patients, supporting their utility as blood-based RNA biomarkers with direct links to disease progression. The *TRBJ1-6* pre-mRNA fragments, which we detected in all 10 IBC plasma samples but none of the healthy or non-IBC plasma samples, were likewise not detected by TGIRT-seq of plasma prepared from a healthy individual at different times (*2*), from different lots of commercial plasma pooled from multiple healthy individuals (*3*), nor from COVID-19 patient PBMCs or plasma (15 and 20 patients, respectively; unpublished data), advancing them as a prime candidate for IBC-specific RNA biomarker validation in larger numbers of samples than the initial cohort analyzed here. Overall, the molecular characteristics found by TGIRT-seq for IBC provide a transformative landscape and road map for further understanding and diagnosing this disease and should enable liquid biopsies that can be used by clinicians to diagnose, monitor disease progression, predict therapeutic responses, and make more informed clinical decisions to improve patient outcomes. We anticipate this will also be the case for other diseases.

## Methods

### Patients and samples

Ten IBC patient FFPE tumor tissue and paired PBMC and plasma samples, 4 non-IBC patient FFPE tumor tissue and paired PBMC and plasma samples, 2 additional non-IBC patient PBMC and plasma samples, and 13 healthy donor PBMC and plasma samples were obtained from the Morgan Welch Inflammatory Breast Cancer Research Program and Clinic at MD Anderson Cancer Center (MDACC). Each patient and healthy donor gave written informed consent. This study was approved by the MDACC Institutional Review Board. IBC was diagnosed by strictly following the criteria of the American Joint Committee on Cancer (*13*). All IBC patient tissues were reviewed by a pathologist (Savitri K.) to ensure accurate diagnosis and tumor cellularity >60%.

For preparation of PBMC and plasma, fresh blood was collected in 10-mL K^+^/EDTA venous blood collection tubes and mixed with an equal volume of phosphate-buffered saline without calcium and magnesium (PBS −/−; Thermo Fisher), then gently layered over 15-mL Ficoll-Paque PLUS (GE Healthcare) in a 50-mL conical tube and centrifuged at 400 × *g* for 35 min at room temperature.

After centrifugation, plasma (top yellow layer) was transferred into a clean tube, aliquoted, and stored at −80°C. The PBMCs (thin white layer between the yellow plasma layer and clear Ficoll layer) were transferred to a 15-ml conical tube, mixed with 3 volumes of PBS −/−, and centrifuged at 450 × *g* for 10 min at room temperature. The pellets were washed twice with PBS−/−, centrifuged at 2,000 rpm for 10 min at room temperature, and used for RNA extraction as described below. Tumor RNA, PBMC RNA, and plasma samples were shipped to UT Austin for RNA analysis on dry ice.

Frozen matched tumor and normal breast tissue RNAs from 4 deidentified non-IBC patients were purchased from OriGene (CR525106 and CR560146, CR520829 and CR561033). Individual patient information, including HR and HER2 status for all MD Anderson patients and provided by the supplier for the frozen commercial samples, is summarized in Table S1.

### RNA preparation

Total cellular RNA was extracted from FFPE tumor slices and PBMCs at MD Anderson by using a miRVana miRNA isolation kit (ThermoFisher), shipped to UT Austin on dry ice, and stored frozen at -80°C until ready for use. For TGIRT-seq library preparation, the thawed tumor and PBMC RNAs (500 ng for most samples but as little as 200 ng for a few tumor samples) were incubated with Baseline-ZERO DNase (Lucigen; 2 units, 30 min at 37°C) to digest DNA followed by rRNA depletion using an Illumina RiboZero Gold (Human/Mouse/Rat) kit. The rRNA-depleted RNAs were then cleaned up with a Zymo RNA clean and concentrator kit by using the manufacturer’s two-fraction protocol to separate long and short RNAs. The long RNAs (>200 nt) were chemically fragmented to 70-90 nt length by using an NEBNext Magnesium RNA Fragmentation Module (New England Biolabs; 94°C for 5 min) and cleaned up with a Zymo RNA clean and concentrator kit using a modified 8× ethanol protocol (v/v ratio 1:2:8 for RNA sample: kit RNA Binding Buffer: 100% ethanol) to minimize loss of very small RNAs (*70, 71*). The chemically fragmented long RNAs were then combined with the non-chemically fragmented short RNAs (<200 nt), and the reconstituted total RNAs were treated with T4 polynucleotide kinase (Lucigen; 50 U for 30 min at 37°C) to remove 3’ phosphates and 2’,3’-cyclic phosphates, which impede TGIRT template switching, followed by a final clean-up with a Zymo RNA clean and concentrator kit using the modified 8× ethanol protocol above.

Plasma RNAs were isolated using a miRNeasy Serum/Plasma Advanced kit (Qiagen) with a modified lysis buffer step in which the plasma samples were incubated with proteinase K (20 mg/mL stock, 1% v/v) in the presence of SDS (0.5% stock, 1% v/v) and linear acrylamide carrier (5 mg/mL stock, 1-2 µl per sample) for 15 min at 50°C in order to improve the recovery of protein-bound RNAs. After neutralization, centrifugation, and column clean up as specified in the kit protocol, the isolated RNAs were treated with T4 polynucleotide kinase (Lucigen; 50 units, 30 min at 37°C) to remove 3’ phosphates and with DNase I (Zymo: 30 U, 30 min at 37°C) to degrade residual DNA, followed by a final clean up with a Zymo RNA clean and concentrator kit using the modified 8× ethanol protocol described above.

The Purchased frozen matched non-IBC tumor and healthy tissue RNAs (500 ng) from 4 non-IBC patients were treated with Baseline-ZERO DNase for 30 min at 37°C to remove residual DNA and then processed by using the Zymo kit two-fraction protocol to obtain total RNA samples with chemically fragmented long RNAs and intact short RNAs, as described above for the FFPE tumor and PBMC samples.

For all samples, RNA concentration and quality were checked by mRNA assay with a Pico Kit on an Agilent 2100 Bioanalyzer. The recovered RNAs (2-50 ng for cellular RNAs and 0.5-2 ng for plasma RNAs) were used for TGIRT-seq library preparation.

### Construction and sequencing of TGIRT-seq libraries

TGIRT-seq libraries were constructed as described (*70, 71*) by using TGIRT-template switching from a synthetic RNA template/DNA primer duplex to the 3’ end of the target RNA for 3’ RNA-seq adapter addition and a single-stranded DNA ligation to the 3’ end of the cDNA using Thermostable 5’ AppDNA/RNA Ligase (New England Biolabs) for 5’ RNA-seq adapter addition. The TGIRT-template switching reaction was done with 1 μM TGIRT-III RT (InGex, LLC) for 15 min at 60°C. The resulting cDNAs were amplified by PCR with primers that add capture sites and indices for Illumina sequence (denaturation 98°C for 5 sec, followed by 12 cycles of 98°C for 5 sec, 65°C for 10 sec, and 72°C for 10 sec) The PCR products were purified by using Agencourt AMPure XP beads (Beckman Coulter), and the libraries were sequenced on an Illumina NextSeq 500 to obtain 2×75-nt paired-end reads.

### Read mapping

Illumina TruSeq adapters and PCR primer sequences were trimmed from the reads with Cutadapt v2.8 (https://github.com/marcelm/cutadapt) (sequencing quality score cut-off at 20; *p*-value <0.01) and reads <15-nt after trimming were discarded. To minimize mismapping, we used a sequential mapping strategy. First, reads were mapped to the human mitochondrial genome (Ensembl GRCh38 Release 93) and the *Escherichia coli* genome (GeneBank: NC_000913) using HISAT2 v2.1.0 (https://github.com/DaehwanKimLab/hisat2) with customized settings (-k 10 --rfg 1,3 --rdg 1,3 -- mp 4,2 --no-mixed --no-discordant -- no-spliced-alignment) to identify reads derived from mitochondrial and *E. coli* RNAs (denoted Pass 1). Unmapped reads from Pass1 were then mapped to a collection of customized references sequences for human sncRNAs (miRNA, tRNA, Y RNA, Vault RNA, 7SL RNA, 7SK RNA) and rRNAs (2.2-kb 5S rRNA repeats from the 5S rRNA cluster on chromosome 1 (1q42, Gene- Bank: X12811) and 43-kb 45S rRNA containing 5.8S, 18S and 28S rRNAs from clusters on chromosomes 13,14,15, 21, and 22 (GeneBank: U13369), using HISAT2 with the following settings -k 20 --rdg 1,3 --rfg 1,3 --mp 2,1 --no-mixed --no-discordant --no-spliced-alignment --norc (denoted Pass 2). Unmapped reads from Pass 2 were then mapped to the human genome reference sequence (Ensembl GRCh38 Release 93) using HISAT2 with settings optimized for nonspliced mapping (-k 10 --rdg 1,3 --rfg 1,3 --mp 4,2 --no-mixed --no-discordant -- no-spliced- alignment) (denoted Pass 3) followed by splice aware mapping (-k 10 --rdg 1,3 --rfg 1,3 --mp 4,2 --no-mixed --no-discordant --dta) (denoted Pass 4). For reads that mapped to multiple genomic loci with the same mapping score in passes 3 to 4, the alignment with the shortest distance between the two paired ends (*i.e.*, the shortest read span) was selected. In the case of ties (*i.e.*, reads with the same mapping score and read span), reads mapping to a chromosome were selected over reads mapping to scaffold sequences, and in other cases, the read was assigned randomly to one of the tied choices. The filtered multiply mapped reads were then combined with the uniquely mapped reads from Passes 3-4 by using SAMtools v1.10 (https://github.com/samtools/samtools).

To generate counts for different genomic features, the mapped reads were intersected with Ensembl GRCh38 Release 93 gene annotations plus the RNY5 gene and its 10 pseudogenes, which were not annotated in this release. Annotations for s were from RepBase (https://www.girinst.org/repbase/), and annotations for hairpin miRNA and mature miRNA were from miRBase (https://www.mirbase.org/). Coverage of each feature was calculated by BEDTools v2.29.2 (https://bedtools.readthedocs.io/en/latest/content/bedtools-suite.html). To avoid miscounting reads with embedded sncRNAs that were not filtered out in Pass 2 (*e.g.*, snoRNAs), reads were first intersected with sncRNA annotations, and the remaining reads were then intersected with the annotations for protein-coding gene RNAs, lincRNAs, antisense RNAs, and other lncRNAs to get the read count for each annotated feature. Read coverages were calculated from merged BAM files and converted to BigWig format using UCSC command line utility tools (https://genome.ucsc.edu/index.html), and then visualized by using Integrative Genomics Viewer v2.6.2 (IGV) (https://software.broadinstitute.org/software/igv/).

### Normalization and differential expression analysis

Read counts from tumor tissue samples were normalized by using DESeq2 (https://github.com/thelovelab/DESeq2) with sample size factors estimated based on counts from nuclear protein-coding genes. Genewise differential expression analyses were also performed using DESeq2, while all other analyses were applied to log_2_-transformed DESeq2-normalized expression values (with log_2_ transformation applied after adding a pseudocount of 1 to avoid the singular behavior of log_2_0). Candidate genes were checked visually by IGV to see if differential expression reflected read coverage across the gene or was limited to specific regions and to filter out protein-coding, lncRNA, and miRNA genes whose differential expression was due to reads mapping to simple repeats or other low complexity sequences as defined by RepBase (https://www.girinst.org/repbase/). For DESeq2 analysis of Repeat-Element RNAs, reads mapping to the same were aggregated by the name of the element and normalized by protein-coding gene reads.

### Identification of gene sets enriched for differential expression

Elevated or depressed DESeq2 test statistics among genes composing either the BROAD Institute’s list of hallmark gene sets (*72*) or Gene Ontology (GO) biological process gene sets (*73*) was assessed by using GO-MWU (Gene Ontology Mann-Whitney U testing, https://github.com/z0on/GO_MWU), which analyzed each GO category G within specified size limits (minimum of 10 observed genes, maximum of 10% of all observed genes) and tests whether the scores (DESeq2 test statistics) associated with genes within G were significantly more likely to be found near the top or bottom of the global ranked list of genes than would randomly shuffled genes. GO categories with highly overlapping sets of associated genes were merged (retaining the name of the largest merged category) by complete linkage hierarchical clustering (based on gene set dissimilarities defined by the ratio of the number of overlapping genes to the number of genes in the smaller gene set) and then applying a dendrogram cut height of 0.25 (thereby merging together a cluster of gene sets only when the most dissimilar pair of them overlaps in ≥75% of genes contained in the smaller of the pair).

### Peak calling

For each sample, a BAM file with read pairs that mapped to the human-genome was processed as described above. Reads that overlapped sncRNAs (Ensembl annotation, www.ensembl.org), rRNA loci, Mt DNA (to eliminate mis-mapping of Mt RNAs to nuclear Mt DNA segments (NUMTs), and human genome blacklist regions (https://sites.google.com/site/anshulkundaje/projects/blacklists) were removed using BEDTools. BAM files from libraries within the same group (Healthy, non-IBC, and IBC) were combined and then split into separate BAM files containing only reads mapping to the forward or reverse strand. BAM files for each strand were then used as input for the MACS2 callpeak algorithm (https://pypi.org/project/MACS2/) using the combined BAM file for both strands from healthy or non-IBC plasma datasets (n=15 and 6, respectively) as the base-line control, via: macs2 callpeak --treatment {combined BAM from IBC dataset, forward or reverse strand} -- control {combined BAM from Healthy or non-IBC dataset, forward or reverse strand } --outdir {outdir} --name {comparison_strand} --nomodel --format BAMPE --qvalue 0.01 --keep-dup all -- gsize hs --min-length 150.

The called peaks from the forward and reverse strands were then combined and filtered to obtain candidate peaks with read coverage ≥10 reads at the peak maximum, a false discovery rate ≤ 0.01 (*q*-value assigned by MACS2), and a requirement that the peak be detected in at least 5 of the 10 IBC plasma samples to avoid batch effects. The annotation of peaks was done with BEDTools (https://bedtools.readthedocs.io/en/latest/content/bedtools-suite.html) intersect command against genomic features from Ensembl genes (https://useast.ensembl.org/index.html) and RepBase- annotated repeats (https://www.girinst.org/repbase/). An overlap score, defined as the number of bases overlapping the genomic feature divided by the peak width, was computed for each overlapping record between a peak and a genomic feature. A maximal overlap score of 1 indicates the peak can be explained by the full-length genomic feature. A best feature annotation for each peak was selected by the highest overlap score. In cases of identical overlap scores, a feature was selected with the priority: repeat regions (RepBase), protein-coding genes, lncRNAs.

### Intron-exon Depth Ratio (IDR) analysis

IDRs were considered both as a single quantity θ representing all genes and separately as an individual quantity θ*_g_* for each gene, through a simple model *y* = θ*x* / [(θ-1)*x*+1], where *y* is the estimated fraction of reads for a protein-coding gene in FFPE IBC tumor samples derived from introns and *x* is the fraction of the genomic annotation of the gene’s length corresponding to introns. This model arises from the assumption that the per-base rate of intronic region inclusion in sequencing reads is a fixed quantity θ times the per-base exonic region inclusion rate. The intronic read fractions *y_g_*for gene *g* are estimated as 1-(number of reads mapped to *g* using transcriptomic reference / number of reads mapped to *g* using genomic reference), capped at a maximum value of 1 and floored at a minimum value of 0. Mapping to the transcriptomic reference was done using HISAT2 (http://daehwankimlab.github.io/hisat2/) and Ensembl GRCh38 Release 93 reference, while genomic mapping was performed as described in read mapping section above. Genomic and transcriptomic mapping results included slightly different gene sets; we assigned IDR values only to genes included in both results sets.

All-genes θ (with no subscript) values were estimated by least-squares using data for all genes; in the FFPE IBC sample set, for example, this yields a value of ∼0.19, indicating that read depth for intronic regions tends to be about 0.19 times the read depth for the exonic regions on average across all genes. Individual genes *g* may have much higher or lower IDR values θ*_g_*, where θ*_g_* is the value of θ which solves *y_g_* = θ*_g_x_g_* / [(θ*_g_*-1)*x_g_*+1] for the values *y_g_* and *x_g_* associated specifically with gene *g* only. IDR values θ*_g_* for each gene were calculated separately for each combination of diagnostic group (healthy, non-IBC, and IBC) and sample type (Frozen tissue, FFPE, and PBMC).

We quantified differences in in IDR values for each gene *g* using a ΔIDR score defined as the ratio of IDR estimated within a target sample group (e.g., FFPE IBC samples) divided the IDR estimated within the reference sample group (e.g., FFPE non-IBC samples). These ΔIDR(*g*) scores were used in gene set ΔIDR enrichment analysis using the same GO-MWU technique (described in its own methods subsection) used for enrichment in DE test statistic scores.

### Categorizing transcripts by TGIRT-seq

Gene body (exons and UTRs only) and intron body (introns only) coverage of each protein-coding gene from combined datasets based on tissue type and disease status was calculated using RSeQC (https://github.com/MonashBioinformaticsPlatform/RSeQC) based on annotated protein-coding genes in Ensembl GRCh38 Release 93. Intron body coverage was analyzed by using a customized script to make hypothetical pseudogenes containing only introns for all protein-coding genes annotated in Ensembl GRCh38 Release 93. Significant differences in gene body and intron body coverage for each gene were determined by student *t*-test. Skewness, a measurement of the asymmetry of distribution in read coverage depth across the intron body, was calculated by R using moments package (https://cran.r-project.org/web/packages/moments/). To classify genes with different intron and exon expression patterns, those with ≥30% of all reads corresponding to the antisense strand were classified as Type IV and removed from further analysis. The remaining genes with mean coverage depth of exons significantly higher than that of introns (*t*-test *p*≤0.05) were classified as Type I; those with no significant difference in mean coverage depth between exons and introns were classified as Type II; those with mean coverage depth across introns significantly higher than that of exons (*t*-test *p*≤0.05) were classified as Type III. Type III genes then were further divided into 3 subtypes: Type IIIA, little or no skewness in intron body coverage (skewness between 0.5 and -0.5); Type IIIB, skewed towards 5’ introns (skewness ≥0.5); and Type IIIC, skewed towards 3’ introns (skewness ≤ -0.5).

## Data deposition

The TGIRT-seq datasets in this manuscript as listed in the Suppl. file have been deposited in the National Center for Biotechnology Information Sequence Read Archive (accession number: PRJNA954747).

## Acknowledgements

We thank the patients and healthy donors for contributing samples and the staff of the Morgan Welch Inflammatory Breast Cancer Research Program and Clinic for collecting those samples. We also thank Lauren Ehrlich (University of Texas at Austin), Tak Mak (University of Toronto), John Moran (University of Michigan), and Ryan Nottingham (University of Texas at Austin) for comments on the manuscript and Shanshan Wang (University of Texas at Austin) for contributing to TGIRT-seq library preparation. The Texas Advanced Computing Center (TACC) and the Center for Biomedical Research Support (CBRS) at the University of Texas at Austin provided high-performance computing resources. High-throughput sequencing was done by the Genomic Sequencing and Analysis Facility at the University of Texas at Austin.

---

**Fig. S1. Principal Component Analysis.** PCA applied to all genes in all sample sets (left panel) and PCA run separately on genes encoding different categories of RNA biotypes in each sample type (right panels) color coded as shown to the right of the plots. For all samples considered together (left panel), the first principal component (PC1) separated the plasma samples (right) from the tumor and PBMC samples, while the second principal component (PC2) split the PBMC samples (+, top left) from frozen breast tissue and FFPE tumor samples. Seven of the 10 IBC FFPE tumor samples (red open circles) clustered on one side of the breast tissue samples without overlapping the FFPE or frozen non-IBC breast tissue samples (blue open or closed circles, respectively), while the remaining 3 IBC tumor samples (FFPE-I04, FFPE-I07, and FFPE-I09) were outliers with lower gene detection rates than the other FFPE tumor samples (see heat map Fig. 3A). Frozen healthy breast tissue samples were separated from frozen non-IBC breast tissue samples, but overlapped the IBC FFPE tumor samples, limiting qualifying comparisons between frozen and FFPE breast tissue samples. PCAs based on single categories of cellular RNA biotypes (Protein coding, lncRNAs, sncRNAs, and Repeat-Element RNAs, right panels) largely separated IBC patients from healthy donor and non-IBC patient samples in all cases, except for sncRNAs in FFPE tumor samples, likely reflecting limited differential expression of sncRNA between IBC and non-IBC FFPE tumor samples (Fig. 3).

**Fig. S2. Volcano plots for DESeq2 comparisons of FFPE breast tissue samples.** The *y*-axis indicates the log_10_-transformed adj. *p*-value while the *x*-axis indicates the DESeq2-estimated LFC. Each point represents one gene with biotypes color coded for those genes with adj. *p*≤0.001 and |LFC|>2. Significantly over-represented and other notable genes that passed filtering steps and were detected in at least 50% of the samples of the group in which they were over-represented are labeled by their gene symbol. Protein-coding and lncRNA genes with IDR>0.5 are marked with an asterisk. nonsig: genes that are not significantly differentially expressed.

**Fig. S3. IGV plots of differentially expressed or notable genes in breast tissue samples.** (**A**) protein-coding genes, (**B**) lncRNAs, (**C**) snoRNAs, and (**D**) mature miRNAs. T and B, top strand and bottom DNA strands, respectively. The number of reads for RNAs in each IGV plot is indicated to the right. The primary comparison in Results is for FFPE IBC versus FFPE non-IBC patient tumors. Protein-coding and lncRNA genes with IDR>0.5 are marked with an asterisk.

**Fig. S4. High ΔIDR genes in IBC versus non-IBC FFPE tumors samples trend toward lower differential expression for reads mapped to the transcriptome reference. (A-C)** Plots of ΔIDR (*y*-axis) versus differential expression enrichment scores calculated by DESeq2 for reads mapped to the transcriptome reference sequence (*x*-axis). For each panel, different categories of genes are color coded as shown beneath the panels based on: (**A**) IDR in IBC tumor samples; (**B**) gene type based on the ratio of intron to exon read depth across the gene; (**C**) membership in Broad Institute Hallmark gene sets with separate trendlines shown for each gene set to highlight their varying patterns of differential expression and ΔIDRs. Select genes which were either: (i) differentially expressed; (ii) have particularly high or ΔIDR values; or (iii) represent the various intron expression patterns shown in panel (D) are named in the plot. **(D)** IGV plots of genes with high ΔIDRs (>2) that are not differentially expressed in IBC compared to non-IBC FFPE tumor samples for reads mapped to the transcriptome reference sequence. Examples are classified as Type I, II, IIIA, IIIB, and IIIC in IBC FFPE tumors based on definitions in Results illustrated in Fig. 2C. Gene length and IDR values for the gene in non-IBC and IBC datasets are shown in parentheses next to the gene name. The number of reads for RNAs in each IGV plot is indicated to the right. Red dashed lines are density plots of intron body coverage over the length of the gene. Protein-coding and lncRNA genes with IDR>0.5 are marked with an asterisk.

**Fig. S5. Heat map of the relative abundance for all 149 DE protein-coding genes (adj. p≤0.001, |LFC|≥2 in IBC PBMC samples relative to the healthy and non-IBC PBMC samples.** Tiles are colored by expression differences relative to the corresponding genewise means taken across all PBMC samples based on the color code top right. Lanes in the heat map are numbered according to the healthy donor or patient ID number (Table S1). Protein-coding genes with IDR>0.5 are marked with an asterisk.

**Fig. S6. Volcano plots for DESeq2 comparisons of PBMC samples.** (**A**) IBC patient vs Healthy donor PBMCs. (**B**) IBC vs non-IBC patient PBMCs. The *y*-axis indicates log_10_-transformed adj. *p*- values, while *x*-axis indicates LFC. Each point represents one gene with biotype color coded for those genes with adj. *p*≤0.001 and |LFC|>2. Significantly over-represented and other notable genes that passed filtering steps and were detected in at least 50% of the samples of the group in which they were over-represented are labeled by their gene symbol. Protein-coding genes with IDR>0.5 are marked with an asterisk; lncRNA genes with IDR>0.5 were not marked with an asterisk due to crowding of gene names (see Fig. 4 and Suppl. file for IDR status). nonsig: genes that are not significantly differentially expressed.

**Fig. S7. IGV plots of differentially expressed or notable genes in PBMC samples.** (**A**) protein-coding genes, (**B**) lncRNAs, (**C**) sncRNAs, (**D**) mature miRNAs, and (**E**) hairpin miRNAs. T and B, top strand and bottom DNA strands, respectively. The number of reads for RNAs in each IGV plot is indicated to the right. Protein-coding and lncRNA genes with IDR>0.5 are marked with an asterisk.

**Fig. S8. Non-IBC patient versus healthy donor PBMC scatter plots.** (**A**) Scatter plot comparing (log_10_-transformed and sign-inverted) FDR-adjusted *p*-values for DE genes in non-IBC patient versus healthy donor PBMCs based on Exon only mapping reads (*x*-axis) versus Exon + Intron mapping reads (*y*-axis). Only genes for with LFC≥1 are shown, and only those for which the adj. *p*- value was ≤0.001 based on at least one of the two mapping strategies (Exons only or Exons + Introns) are named in the plots. (**B**) Scatter plot comparing (log_10_-transformed and sign-inverted) FDR-adjusted *p*-values from GO-MWU non-IBC patient versus healthy donor analysis of BROAD Hallmark gene sets using gene counts generated by Exon only mapping reads (*x*-axis) or Exon + Intron mapping reads (*y*-axis). Gene sets for which the average gene MWU test statistic indicated increased expression in non-IBC are colored red, while those with decreased expression in non-IBC are colored blue. Vertical and horizontal dashed lines indicate adj. *p*=0.001.

**Fig. S9. Hallmark TNFα-via-NFκB and Inflammatory Response gene scatter plots for reads mapped to genome and transcriptome reference sequences.** Scatter plots of log_2_-transformed normalized counts of genes belonging to Hallmark TNFα-via-NFκB (blue) and Inflammatory Response (red) gene sets in IBC FFPE tumor samples (*y*-axis) versus IBC PBMC (*x*-axis) datasets. Genes belonging to both gene sets are colored in purple. Genes that have LFC≥2 are labeled along with *JUN* and *FOS*. Pearson correlation coefficients (*r*) are shown at the bottom right.

**Fig. S10. CIBERSORTx-estimated fractions of PBMC cell types for all samples grouped by diagnosis.** Points represent individual sample cell type fraction estimates (*y*-axis) separated by patient diagnosis (Healthy, non-IBC, or IBC). Midpoints of boxes indicate median values for each sample group, with bottom and top of boxes indicating 25^th^ and 75^th^ percentile estimates for within-group distributions of cell type fractions. **(A)** CIBERSORTx analyses conducted using Exon + Intron mapping gene counts, **(B)** CIBERSORTx analyses conducted using Exon only mapping gene counts. Mann-Whitney U-testing indicated significant differences between a number of groups: *p<*0.05 for eosinophil abundances in IBC PBMC samples compared to the combined healthy and non-IBC sample sets using either Exon only or Exon + Intron alignments; for neutrophils, *p*=0.0011 using Exon + Intron alignments and *p*=0.053 using Exon only alignments. Considering Exon only alignments, activated dendritic cells were found to be elevated in IBC (*p*=0.011; for Exon + Intron analysis, *p*=0.061) and significant evidence (*p*=0.010) of reduction in resting mast cells, another class of granulocytes, in IBC PBMCs relative to the combined non-IBC patient and healthy donor PBMC sample sets.

**Fig. S11. Heat map of the relative abundance of CIBERSORTx-related genes in PBMC samples.** Relevant CIBERSORTx marker panel genes (those most highly expressed in the CIBERSORTx training set data for activated dendritic cells, eosinophils, neutrophils, or resting mast cells, as well as *BRAF*) and one other gene (*ATP2B1\**) potentially relevant to the IBC PBMC response that were differentially expressed (adj. *p*≤0.001, |LFC|≥1) in IBC patient PBMCs compared to either non-IBC patient or healthy donors PBMCs in Exon + Intron alignments are shown. Tiles are colored by expression differences relative to the corresponding genewise means taken across all PBMC samples for (**A**) Exon + Intron alignments and (**B**) Exon only alignments. Lanes in heat maps are numbered according to the healthy donor or patient ID number (Table S1).

**Fig. S12. High ΔIDR genes sequenced in IBC versus non-IBC PBMC samples trend toward lower differential expression for reads mapped to the transcriptome reference. (A-C)** ΔIDR (*y*- axis) versus differential expression enrichment scores calculated by DESeq2 for reads mapped to the transcriptome reference sequence (*x*-axis) with genes (and, when applicable, their names) color coded as shown beneath the panels based on (**A**) IDR in IBC PBMC samples; (**B**) Gene type based on the ratio of intron to exon read depth across the gene as defined in Fig.2C; (**C**) Membership in Broad Institute Hallmark gene sets (with separate trendlines shown for each gene set to highlight their varying patterns of differential expression and ΔIDR). Select genes which were either: (i) differentially expressed; (ii) have particularly high or low ΔIDR values; or (iii) represent one of the different intronic expression patterns are named in panels (A-C).

**Fig. S13. Scatter plots comparing IBC vs non-IBC DE genes LFCs in FFPE tumors and PBMCs.** Each point represents an identified over-expressed gene in IBC vs non-IBC patient PBMCs with adj. *p*≤0.001, LFC>2 (left panel) or adj. *p*≤0.05, LFC>1 (right panel). The *y-*axis encodes DESeq2 LFC for the FFPE tumor IBC vs non-IBC comparison while the *x*-axis encodes the LFC for PBMC IBC vs non-IBC comparison. Colors indicate different gene biotype indicated to the right of the plots. Trendlines show the linear model fit summarizing relationship between two comparisons for all plotted genes, fit separately for each indicated gene biotype.

**Fig. S14. Volcano plots for DESeq2 two-group comparisons in plasma samples.** The *y*-axis indicates log_10_-transformed adj. *p*, while the *x*-axis indicates LFC. Each point represents one gene with biotypes color coded for those genes with adj. *p*≤0.001 and |LFC|>2. Significantly over- represented and other notable genes that passed filtering steps and were detected in at least 50% of the samples of the group in which they were over-represented are labeled by their gene symbol. i: protein-coding gene contains reads that are mostly derived from intronic regions. nonsig: genes that are not significantly differentially expressed.

**Fig. S15. IGV plots of differentially expressed or notable genes in plasma samples.** (**A**) protein-coding genes, (**B**) lncRNAs, (**C**) sncRNAs, and (**D**) miRNAs. T and B, top and bottom DNA strands, respectively. The number of reads is indicated to the right. i: protein-coding gene contains reads that are mostly derived from intronic regions.

**Fig. S16. IGV plots for protein-coding genes whose intron RNAs were significantly over-represented in IBC plasma and for which intron RNA fragments could be traced to tumors or PBMCs.** The IGV plots show coverage in combined datasets for IBC FFPE tumors, PBMC, and plasma with expanded regions at the bottom for intron RNA fragments found in 5 or more IBC plasma samples. The number of reads is indicated to the right. Types of gene based on IDR analysis in IBC FFPE tumor samples and PBMCs are indicated at the top right of each panel. Intron RNA fragments corresponding to RepBase-annotated simple repeats are indicated in red.

**Fig. S17. Scatter plots comparing the relative abundance of different Repeat-Element RNAs detected in breast tissues, PBMCs or Plasma.** (**A**) SINE elements, (**B**) LTR-containing retroelements, (**C**) DNA elements, and (**D**) other Repeat-Element RNAs. DESeq2-normalized counts of Repeat-Element RNAs were averaged by tissue type and disease status and log_2_- transformed for pairwise comparisons. The numbers at the top left of each indicate the percentage of Repeat-Element RNAs above the diagonal, a measure of over-expression of Repeat-Element RNAs in each comparison. Pearson’s (*r*) and Spearman’s (*r_s_*) correlation coefficients are shown at the bottom right. Subfamilies of the Repeat-Element RNAs are color coded as indicated in the top center box for each family of repeats.

