## Supplemental Table 1 for "TGIRT-seq of Inflammatory Breast Cancer Tumor and Blood Samples Reveals Widespread Enhanced Transcription Impacting RNA Splicing and Intronic RNAs in Plasma"

**Table S1. Donor information**

| Donor ID* (TGIRT-seq) | Tissue type (Dataset ID**) | | |  | Age | Race | ER^+/-^ | PR^+/-^ | HER2^+/-^ | HR^+/-^ | IBC |
| --- | --- | --- | --- | --- | --- | --- | --- | --- | --- | --- | --- |
|  | Frozen tissue | FFPE | PBMC | Plasma |  |  |  |  |  |  |  |
| H01 |  |  | PBMCH01 | PlasmaH01 |  |  |  |  |  |  |  |
| H02 |  |  | PBMCH02 | PlasmaH02 |  |  |  |  |  |  |  |
| H03 |  |  | PBMCH03 | PlasmaH03 |  |  |  |  |  |  |  |
| H04 |  |  | PBMCH04 | PlasmaH04 |  |  |  |  |  |  |  |
| H05 |  |  | PBMCH05 | PlasmaH05 |  |  |  |  |  |  |  |
| H06 |  |  | PBMCH06 | PlasmaH06 |  |  |  |  |  |  |  |
| H07 |  |  | PBMCH07 | PlasmaH07 |  |  |  |  |  |  |  |
| H08 |  |  | PBMCH08 | PlasmaH08 |  |  |  |  |  |  |  |
| H09 |  |  | PBMCH09 | PlasmaH09 |  |  |  |  |  |  |  |
| H10 |  |  | PBMCH10 | PlasmaH10 |  |  |  |  |  |  |  |
| H11 |  |  | PBMCH11 | PlasmaH11 |  |  |  |  |  |  |  |
| H12 |  |  | PBMCH12 | PlasmaH12 |  |  |  |  |  |  |  |
| H13 |  |  | PBMCH13 | PlasmaH13 |  |  |  |  |  |  |  |
| B01 |  | FFPEB01 | PBMCB01 | PlasmaB01 | 51-55 | White | Negative | Negative | Negative | Negative | N |
| B02 |  | FFPEB02 | PBMCB02 | PlasmaB02 | 61-65 | White | Positive | Positive | Negative | Positive | N |
| B03 |  |  | PBMCB03 | PlasmaB03 | 41-45 | White | Positive | Positive | Negative | Positive | N |
| B04 |  | FFPEB04 | PBMCB04 | PlasmaB04 | 66-70 | Black | Negative | Negative | Negative | Negative | N |
| B05 |  |  | PBMCB05 | PlasmaB05 | 46-50 | Black | Negative | Negative | Negative | Negative | N |
| B06 |  | FFPEB06 | PBMCB06 | PlasmaB06 | 46-50 | Black | Positive | Positive | Positive | Positive | N |
| I01 |  | FFPEI01 | PBMCI01 | PlasmaI01 | 71-75 | White | Positive | Negative | Negative | Positive | Y |
| I02 |  | FFPEI02 | PBMCI02 | PlasmaI02 | 66-70 | White | Negative | Negative | Negative | Negative | Y |
| I03 |  | FFPEI03 | PBMCI03 | PlasmaI03 | 41-45 | White | Negative | Negative | Negative | Negative | Y |
| I04 |  | FFPEI04 | PBMCI04 | PlasmaI04 | 56-60 | White | Positive | Positive | Negative | Positive | Y |
| I05 |  | FFPEI05 | PBMCI05 | PlasmaI05 | 21-25 | White | Positive | Negative | Negative | Positive | Y |
| I06 |  | FFPEI06 | PBMCI06 | PlasmaI06 | 51-55 | White | Positive | Positive | Negative | Positive | Y |
| I07 |  | FFPEI07 | PBMCI07 | PlasmaI07 | 41-45 | White | Positive | Positive | Negative | Positive | Y |
| I08 |  | FFPEI08 | PBMCI08 | PlasmaI08 | 51-55 | White | Positive | Positive | Negative | Positive | Y |
| I09 |  | FFPEI09 | PBMCI09 | PlasmaI09 | 46-50 | White | Positive | Negative | Negative | Positive | Y |
| I10 |  | FFPEI10 | PBMCI10 | PlasmaI10 | 56-60 | White | Positive^#^ | Negative | Negative | Positive | Y |
| H01 | FTH01 |  |  |  | 46-50 | White |  |  |  |  |  |
| H02 | FTH02 |  |  |  | 61-65 | Unknown |  |  |  |  |  |
| H03 | FTH03 |  |  |  | 56-60 | White |  |  |  |  |  |
| H04 | FTH04 |  |  |  | 71-75 | White |  |  |  |  |  |
| B01 | FTB01 |  |  |  | 46-50 | White | Negative | NA | Negative | NA | N |
| B02 | FTB02 |  |  |  | 61-65 | Unknown | Positive | Positive | Negative | Positive | N |
| B03 | FTB03 |  |  |  | 56-60 | White | Positive | Positive | Negative | Positive | N |
| B04 | FTB04 |  |  |  | 71-75 | White | Negative | NA | Negative | NA | N |

*: Donor ID: H, healthy; B, non-IBC; I, IBC

**: Dataset ID: PBMC, peripheral blood mononuclear cells; FFPE, formalin-fixed paraffin-embedded breast tissue; FT, frozen breast tissue; H, healthy; B, non-IBC; I, IBC

#: ER 1%, classified as low positive in a grading system which 1-10% is low positive and 10-100% is positive.

All other ER positive FFPE tumors were between 30 and 100
