## Supplemental Table 2 for "TGIRT-seq of Inflammatory Breast Cancer Tumor and Blood Samples Reveals Widespread Enhanced Transcription Impacting RNA Splicing and Intronic RNAs in Plasma"

**Table S2. Differentially expressed (DE) genes in IBC patient versus non-IBC patient and healthy donor samples with adj. p≤0.001 and |LFC|≥2.**

| **Gene Name** | **Encoded Protein or RNA** | **Cancer Relevance** | **References** |
| --- | --- | --- | --- |
| **FFPE samples** | | | |
| **Protein-coding genes** | | | |
| *AKAP13* | A-kinase Anchor Protein 13 | AKAP13 expression correlates with a non-favorable outcome after tamoxifen treatment and with ERαS305P positivity in breast cancer patients. | Bentin Toaldo C, Alexi X, Beelen K, Kok M, Hauptmann M, Jansen M, Berns E, Neefjes J, Linn S, Michalides R, Zwart W. Protein Kinase A-induced tamoxifen resistance is mediated by anchoring protein AKAP13. *BMC Cancer*. 2015 Aug 14;15:588. doi: 10.1186/s12885-015-1591-4. PMID: 26272591; PMCID: PMC4536754. |
| *ALDH1A1* | Aldehyde Dehydrogenases 1 Family Member A1 | ALDH1A1 overexpression is an oncogenic factor in most cancers. | H. Yue, Z. Hu, R. Hu, Z. Guo, Y. Zheng, Y. Wang, Y. Zhou, ALDH1A1 in Cancers: Bidirectional Function, Drug Resistance, and Regulatory Mechanism. *Front Oncol* **12**, 918778 (2022). |
| *ANKRD53* | Ankyrin Repeat Domain 53 | *ANKRD53* genes have differentially methylated regions in breast cancer patient samples but not a potential biomarker for breast cancer. | Noruzinia M, Tavakkoly-Bazzaz J, Ahmadvand M, Azimi F, Dehghanifard A, Khakpour G. Young Breast Cancer: Novel Gene Methylation in WBC. *Asian Pac J Cancer Prev.* 2021 Aug 1;22(8):2371-2375. doi: 10.31557/APJCP.2021.22.8.2371. PMID: 34452548; PMCID: PMC8629456. |
| *ARHGAP10* | Rho GTPase-Activating Protein 10 | ARHGAP10 is downregulated in ovarian cancer. It suppresses tumorigenicity of ovarian cancer cells. | Luo N, Guo J, Chen L, Yang W, Qu X, Cheng Z. ARHGAP10, downregulated in ovarian cancer, suppresses tumorigenicity of ovarian cancer cells. *Cell Death Dis.* 2016 Mar 24;7(3):e2157. doi: 10.1038/cddis.2015.401. PMID: 27010858; PMCID: PMC4823924. |
| *ASB4* | Ankyrin Repeat and SOCS Box Containing 4 | Loss of function of ASB4 hindered tumor cell migration and invasion in HCC. | W. H. Townley-Tilson, Y. Wu, J. E. Ferguson 3rd, C. Patterson, The ubiquitin ligase ASB4 promotes trophoblast differentiation through the degradation of ID2. PLoS One, e89451 (2014). |
| *CCDC33* | Coiled-Coil Domain Containing 33 | Not Reported. |  |
| *CCNI2* | Cyclin I Family Member 2 | CCNI2 is involved in cell cycle regulation and promoting progression of colorectal cancer. It also promotes the malignant phenotype of pancreatic cancer through PI3K pathway. | Lai DM, Bi JJ, Chen YH, Wu YD, Huang QW, Li HJ, Zhang S, Fu Z, Tong YX. CCNI2 plays a promoting role in the progression of colorectal cancer. *Cancer Med.* 2021 Mar;10(6):1913-1924. doi: 10.1002/cam4.3504. Epub 2021 Feb 23. PMID: 33620152; PMCID: PMC7957193.  Hu B, Zhang W, Zhang C, Li C, Zhang N, Pan K, Ge X, Wan T. CCNI2 promotes pancreatic cancer through PI3K/AKT signaling pathway. *Biomol Biomed.* 2023 Aug 1. doi: 10.17305/bb.2023.9337. Epub ahead of print. PMID: 37540586. |
| *CD24* | CD24 Antigen | CD24 is a regulator of cancer cell migration, invasion, and proliferation. | P. Altevogt, M. Sammar, L. Huser, G. Kristiansen, Novel insights into the function of CD24: A driving force in cancer. *Int J Cancer* **148**, 546-559 (2021). |
| *COL4A6* | Collagen Type IV Alpha 6 | Downregulation of collagen COL4A6 is associated with prostate cancer progression and metastasis. | J. B. Ma, J. Y. Bai, H. B. Zhang, L. Gu, D. He, P. Guo, Downregulation of Collagen COL4A6 Is Associated with Prostate Cancer Progression and Metastasis. *Genet Test Mol Biomarkers* **24**, 399-408 (2020). |
| *CROCC2* | Ciliary Rootlet Coiled-Coil 2 | CROCC2 is a tumor microenvironment-related prognostic biomarker for ovarian serous cancer (OSC). High expression of CROCC2 is associated with poor 3-year OSC mortality. | Wang L, Sun X, Jin C, Fan Y, Xue F. Identification of Tumor Microenvironment-Related Prognostic Biomarkers for Ovarian Serous Cancer 3-Year Mortality Using Targeted Maximum Likelihood Estimation: A TCGA Data Mining Study. *Front Genet.* 2021 Jun 3;12:625145. doi: 10.3389/fgene.2021.625145. PMID: 34149794; PMCID: PMC8211425. |
| *CSRNP1* | Cysteine and Serine Rich Nuclear Protein 1 | CSRNP1 may serve as a prognostic biomarker for predicting overall survival in patients with clear cell renal cell carcinoma (ccRCC). A higher expression of CSRNP1 is associated with a better prognosis of ccRCC. | H. Zhang, X. Qiu, G. Yang, The CSRNP Gene Family Serves as a Prognostic Biomarker in Clear Cell Renal Cell Carcinoma. *Front Oncol* **11**, 620126 (2021). |
| *CTGF* (*CCN2*) | Cellular Communication Network Factor 2 | CTGF promotes cancer initiation, progression, and metastasis by regulating cell proliferation, migration, invasion, drug resistance, and epithelial-mesenchymal transition (EMT). | Y. W. Shen, Y. D. Zhou, H. Z. Chen, X. Luan, W. D. Zhang, Targeting CTGF in Cancer: An Emerging Therapeutic Opportunity. *Trends Cancer* **7**, 511-524 (2021). |
| *CYR61 (CCN1)* | Cellular Communication Network Factor 1 | CYR61 is an extracellular matrix protein involved in the transduction of growth factor and hormone signaling that is frequently altered in expression in several types of cancers. | N. Terada, P. Kulkarni, R. H. Getzenberg, Cyr61 is a potential prognostic marker for prostate cancer. *Asian J Androl* **14**, 405-408 (2012). |
| *C6orf15* | Chromosome 6 Open Reading Frame 15 | C6orf15 is associated with significantly different overall survival rates of liver cancer patients. | S. Oh, H. J. Kwon, J. Jung, Estrogen exposure causes the progressive growth of SK-Hep1-derived tumor in ovariectomized mice. *Toxicol Res* **38**, 1-7 (2022). |
| *DCD* | Dermcidin | DCD is an antibacterial gene identified as a neural survival factor that is overexpressed in some invasive breast carcinomas that may promote breast tumorigenesis via modulation of ERBB signaling pathways. | J. Bancovik, D. F. Moreira, D. Carrasco, J. Yao, D. Porter, R. Moura, A. Camargo, C. C. Fontes-Oliveira, M. G. Malpartida, S. Carambula, E. Vannier, B. E. Strauss, A. Wakamatsu, V. A. Alves, A. F. Logullo, F. A. Soares, K. Polyak, J. E. Belizario, Dermcidin exerts its oncogenic effects in breast cancer via modulation of ERBB signaling. *BMC Cancer* **15**, 70 (2015). |
| *DNAH9* | Dynein Heavy Chain 9 | Gastric cancer patients with somatic mutation of DNAH gene have better prognosis than patients with wild type DNAH gene. | Zhu, C., Yang, Q., Xu, J. et al. Somatic mutation of DNAH genes implicated higher chemotherapy response rate in gastric adenocarcinoma patients. *J Transl Med* 17, 109 (2019). https://doi.org/10.1186/s12967-019-1867-6. |
| *DUSP1* | Dual Specificity Phosphatase 1 | DUSP1 promotes tumorigenesis in a number of cancers and regulates immune escape in cancer cells including prostate cancer, gastric cancer and lung cancer. | V. Moncho-Amor, I. Ibanez de Caceres, E. Bandres, B. Martinez-Poveda, J. L. Orgaz, I. Sanchez-Perez, S. Zazo, A. Rovira, J. Albanell, B. Jimenez, F. Rojo, C. Belda-Iniesta, J. Garcia-Foncillas, R. Perona, DUSP1/MKP1 promotes angiogenesis, invasion and metastasis in non-small-cell lung cancer. *Oncogene* **30**, 668-678 (2011). |
| *EGFR* | Epidermal Growth Factor Receptor | EGFR is overexpressed in a variety of cancers and is associated with poor prognosis, low survival rate, and resistance of cancer cells to treatment. | M. L. Uribe, I. Marrocco, Y. Yarden, EGFR in Cancer: Signaling Mechanisms, Drugs, and Acquired Resistance. *Cancers (Basel)* **13**, (2021). |
| *EGR1* | Early Growth Response Protein 1 | EGR1 is closely related to the initiation and progression of cancer and may participate in tumor cell proliferation, invasion, metastasis, and angiogenesis. EGR1 is essential for T and B cell development. It mediates an EGFR pathway-regulated immunosuppressive tumor microenvironment in IBC. | B. Wang, H. Guo, H. Yu, Y. Chen, H. Xu, G. Zhao, The Role of the Transcription Factor EGR1 in Cancer. *Front Oncol* **11**, 642547 (2021).  Bettini M, Xi H, Milbrandt J, Kersh GJ. Thymocyte development in early growth response gene 1-deficient mice. *J Immunol.* 2002;169(4):1713-20. doi: 10.4049/jimmunol.169.4.1713. PubMed PMID: 12165491.  McMahon SB, Monroe JG. The role of early growth response gene 1 (egr-1) in regulation of the immune response. J Leukoc Biol. 1996;60(2):159-66. doi: 10.1002/jlb.60.2.159. PubMed PMID: 8773576.  X. Wang, T. Semba, G. C. Manyam, J. Wang, S. Shao, F. Bertucci, P. Finetti, S. Krishnamurthy, L. T. H. Phi, T. Pearson, S. J. Van Laere, J. K. Burks, E. N. Cohen, J. M. Reuben, F. Yang, H. Min, N. Navin, V. N. Trinh, T. Iwase, H. Batra, Y. Shen, X. Zhang, D. Tripathy, N. T. Ueno, EGFR is a master switch between immunosuppressive and immunoactive tumor microenvironment in inflammatory breast cancer. *Sci Adv* **8**, eabn7983 (2022). |
| *F3* | Coagulation Factor III; Tissue Factor | TF expression is a marker for  malignant angiogenesis in human breast carcinoma. | J. Contrino, G. Hair, D. L. Kreutzer, F. R. Rickles, In situ detection of tissue factor in vascular endothelial cells: correlation with the malignant phenotype of human breast disease. *Nat Med* **2**, 209-215 (1996). |
| *FAH* | Fumarylacetoacetate Hydrolase | FAH induces metabolic reprogramming in melanoma. | Z. Gu, H. Zhang, Y. Li, S. Shen, X. Yin, W. Zhang, R. Cheng, Y. Zhang, X. Zhang, H. Chen, B. Huang, Y. Cao, CDC5L drives FAH expression to promote metabolic reprogramming in melanoma. *Oncotarget* **8**, 114328-114343 (2017). |
| *FAM46A* | Family with Sequence Similarity 46 Member A | FAM46A expression is elevated in glioblastoma and predicts poor prognosis of patients. | Wang Y, Cai R, Wang P, Huang C, Zhang C, Liu Z. FAM46A expression is elevated in glioblastoma and predicts poor prognosis of patients. *Clin Neurol Neurosurg*. 2021 Feb;201:106421. doi: 10.1016/j.clineuro.2020.106421. Epub 2020 Dec 8. PMID: 33370626. |
| *FGFBP1* | Fibroblast Growth Factor Binding Protein 1 | FGFBP1 promotes cell proliferation and migration in pancreatic cancer. | Z. Zhang, M. Liu, Q. Hu, W. Xu, W. Liu, Q. Sun, Z. Ye, G. Fan, X. Xu, X. Yu, S. Ji, Y. Qin, FGFBP1, a downstream target of the FBW7/c-Myc axis, promotes cell proliferation and migration in pancreatic cancer. *Am J Cancer Res* **9**, 2650-2664 (2019). |
| *FKBP5* | FKBP Prolyl Isomerase 5 | FKBP5 is overexpressed in brain cancers, prostate cancer, lymphoma, head and neck cancer, and melanoma. | L. Li, Z. Lou, L. Wang, The role of FKBP5 in cancer aetiology and chemoresistance. *Br J Cancer* **104**, 19-23 (2011). |
| *FOS* | Fos Proto-Oncogene | c-Fos regulates EMT and cancer stem cell reprogramming in Head and neck squamous cell carcinoma cells. FOS encodes components of Activator Protein 1 (AP-1), a modulator of inflammatory responses in cancers and immune disorders. | N. Muhammad, S. Bhattacharya, R. Steele, N. Phillips, R. B. Ray, Involvement of c-Fos in the Promotion of Cancer Stem-like Cell Properties in Head and Neck Squamous Cell Carcinoma. *Clin Cancer Res* **23**, 3120-3128 (2017).  Trop-Steinberg S, Azar Y. AP-1 Expression and its Clinical Relevance in Immune Disorders and Cancer. *Am J Med Sci.* 2017 May;353(5):474-483. doi: 10.1016/j.amjms.2017.01.019. Epub 2017 Feb 1. PMID: 28502334. |
| *GEM* | GTP-Binding Protein GEM | Expression of GEM is associated with metastasis and poor outcome in advanced bladder cancer. | J. R. Laurberg, J. B. Jensen, T. Schepeler, M. Borre, T. F. Orntoft, L. Dyrskjot, High expression of GEM and EDNRA is associated with metastasis and poor outcome in patients with advanced bladder cancer. *BMC Cancer* **14**, 638 (2014). |
| *GP2* | Glycoprotein 2 | GP2 coding variants correlate with the risk of pancreatic cancer. | Y. Lin, M. Nakatochi, N. Sasahira, M. Ueno, N. Egawa, Y. Adachi, S. Kikuchi, Glycoprotein 2 in health and disease: lifting the veil. *Genes Environ* **43**, 53 (2021). |
| *GREM2* | Gremlin 2, DAN Family BMP Antagonist | GREM2 overexpression in adipocytes can inhibit adipogenesis, reduce the expression and secretion of several adipokines, including IL-6, and ultimately inhibit breast cancer progression.  Gremlin 2 limits inflammation after myocardial infarction. | Jung J, Kim NH, Kwon M, Park J, Lim D, Kim Y, Gil W, Cheong YH, Park SA. The inhibitory effect of Gremlin-2 on adipogenesis suppresses breast cancer cell growth and metastasis. *Breast Cancer Res.* 2023 Oct 25;25(1):128. doi: 10.1186/s13058-023-01732-2. PMID: 37880751; PMCID: PMC10599028.  Sanders LN, Schoenhard JA, Saleh MA, Mukherjee A, Ryzhov S, McMaster WG Jr, Nolan K, Gumina RJ, Thompson TB, Magnuson MA, Harrison DG, Hatzopoulos AK. BMP Antagonist Gremlin 2 Limits Inflammation After Myocardial Infarction. *Circ Res.* 2016 Jul 22;119(3):434-49. doi: 10.1161/CIRCRESAHA.116.308700. Epub 2016 Jun 9. PMID: 27283840; PMCID: PMC4961528. |
| *HPSE2* | Heparanase 2 | HPSE2 promotes colorectal cancer proliferation and is associated with poor prognosis.  HPSE2 protein has a protective effect on LPS-induced inflammatory conditions and subsequent glomerulonephritis *in vivo*. | Zhang, H., Xu, C., Shi, C. *et al.* Hypermethylation of heparanase 2 promotes colorectal cancer proliferation and is associated with poor prognosis. *J Transl Med* **19**, 98 (2021). <https://doi.org/10.1186/s12967-021-02770-0>.  Buijsers B, Garsen M, de Graaf M, Bakker-van Bebber M, Guo C, Li X, van der Vlag J. Heparanase-2 protein and peptides have a protective effect on experimental glomerulonephritis and diabetic nephropathy. *Front Pharmacol.* 2023 Apr 27;14:1098184. doi: 10.3389/fphar.2023.1098184. PMID: 37180718; PMCID: PMC10172501. |
| *IRF9* | Interferon Regulatory Factor 9 | IRF9 is a marker of active intratumoral type I and II interferon (IFN) signaling and it reduces risk of distant relapse. Reduced IRF9 expression predicts poor outcome in TNBC despite chemotherapy. The loss of IRF9 predicts relapse in those patients who do not have a complete response to chemotherapy. | Brockwell NK, Rautela J, Owen KL, Gearing LJ, Deb S, Harvey K, Spurling A, Zanker D, Chan CL, Cumming HE, Deng N, Zakhour JM, Duivenvoorden HM, Robinson T, Harris M, White M, Fox J, Ooi C, Kumar B, Thomson J, Potasz N, Swarbrick A, Hertzog PJ, Molloy TJ, Toole SO, Ganju V, Parker BS. Tumor inherent interferon regulators as biomarkers of long-term chemotherapeutic response in TNBC. *NPJ Precis Oncol.* 2019 Aug 29;3:21. doi: 10.1038/s41698-019-0093-2. PMID: 31482136; PMCID: PMC6715634. |
| *KCNJ16* | Potassium Inwardly-Rectifying Channel, Subfamily J, Member 16 | KCNJ16 is highly expressed in the renal tubule and collecting duct system. It plays an important role in the development of clear cell renal cell carcinoma (ccRCC). | Zhang Y., Narayanan S. P., Mannan R., Raskind G., Wang X., Vats P., et al. (2021). Single-cell Analyses of Renal Cell Cancers Reveal Insights into Tumor Microenvironment, Cell of Origin, and Therapy Response. *Proc. Natl. Acad. Sci. U.S.A.* **118** (24). 10.1073/pnas.2103240118. |
| *KLF9* | Kruppel Like Factor 9 | KLF9 functions as a transcriptional repressor and thereby regulates multiple signaling pathways involved in oncogenesis and stem cell regulation. | M. Ying, J. Tilghman, Y. Wei, H. Guerrero-Cazares, A. Quinones-Hinojosa, H. Ji, J. Laterra, Kruppel-like factor-9 (KLF9) inhibits glioblastoma stemness through global transcription repression and integrin alpha6 inhibition. *J Biol Chem* **289**, 32742-32756 (2014). |
| *KCNK9* | Potassium Two Pore Domain Channel Subfamily K Member 9 | Hypomethylation of KCNK9 in breast cancer is associated with increases in both mitochondrial membrane potential and apoptosis resistance. | Skaar DA, Dietze EC, Alva-Ornelas JA, Ann D, Schones DE, Hyslop T, Sistrunk C, Zalles C, Ambrose A, Kennedy K, Idassi O, Miranda Carboni G, Gould MN, Jirtle RL, Seewaldt VL. Epigenetic Dysregulation of KCNK9 Imprinting and Triple-Negative Breast Cancer. *Cancers (Basel).* 2021 Nov 30;13(23):6031. doi: 10.3390/cancers13236031. PMID: 34885139; PMCID: PMC8656495. |
| *MT_ATP8* | Mitochondrially Encoded ATP Synthase Membrane Subunit 8 | *MT_ATP8* gene mutations have been observed in various cancer types, such as prostate cancer osteosarcoma and breast cancer. | Grzybowska-Szatkowska, L.; Slaska, B.; Rzymowska, J.; Brzozowska, A.; Florianczyk, B. Novel mitochondrial mutations in the ATP6 and ATP8 genes in patients with breast cancer. *Mol. Med. Rep.* 2014, 10, 1772–1778. |
| *MYO18B* | Myosin-XVIIIb | MYO18B gene is a strong candidate for a novel tumor suppressor gene whose inactivation is involved in lung cancer progression. | Nishioka M, Kohno T, Tani M, Yanaihara N, Tomizawa Y, Otsuka A, Sasaki S, Kobayashi K, Niki T, Maeshima A, Sekido Y, Minna JD, Sone S, Yokota J. MYO18B, a candidate tumor suppressor gene at chromosome 22q12.1, deleted, mutated, and methylated in human lung cancer. *Proc Natl Acad Sci U S A*. 2002 Sep 17;99(19):12269-74. doi: 10.1073/pnas.192445899. Epub 2002 Sep 3. PMID: 12209013; PMCID: PMC129434. |
| *NEXN* | Nexilin F-Actin Binding Protein | Nexilin regulates oligodendrocyte progenitor cell migration. | Q. Li, H. Zhao, P. Pan, X. Ru, S. Zuo, J. Qu, B. Liao, Y. Chen, H. Ruan, H. Feng, Nexilin Regulates Oligodendrocyte Progenitor Cell Migration and Remyelination and Is Negatively Regulated by Protease-Activated Receptor 1/Ras-Proximate-1 Signaling Following Subarachnoid Hemorrhage. *Front Neurol* **9**, 282 (2018). |
| *NR4A1* | Nuclear Receptor Subfamily 4 Group A Member 1 | NR4A1 regulates cell growth, apoptosis, metastasis, and metabolism in many tumors, including melanoma, colorectal cancer, breast cancer, and hepatocellular cancer.  NR4A1 has an anti-inflammatory role for NR4A1 in macrophages in both atherosclerosis and chronic inflammation models. | S. Deng, B. Chen, J. Huo, X. Liu, Therapeutic potential of NR4A1 in cancer: Focus on metabolism. *Front Oncol* **12**, 972984 (2022).  Hanna RN, Shaked I, Hubbeling HG, Punt JA, Wu R, Herrley E, et al. NR4A1 (Nur77) deletion polarizes macrophages toward an inflammatory phenotype and increases atherosclerosis. *Circ Res*. 2012;110(3):416–427. pmid:22194622  Koenis DS, Medzikovic L, van Loenen PB, van Weeghel M, Huveneers S, Vos M, et al. Nuclear Receptor Nur77 Limits the Macrophage Inflammatory Response through Transcriptional Reprogramming of Mitochondrial Metabolism. *Cell Rep.* 2018;24(8):2127–2140 e7. pmid:30134173 |
| *NRXN1* | Neurexin-1-Alpha | NRXN1 as a novel potential target of antibody-drug conjugates for small cell lung cancer. | Yotsumoto T, Maemura K, Watanabe K, Amano Y, Matsumoto Y, Zokumasu K, Ando T, Kawakami M, Kage H, Nakajima J, Yatomi Y, Nagase T, Takai D. NRXN1 as a novel potential target of antibody-drug conjugates for small cell lung cancer. *Oncotarget.* 2020 Sep 29;11(39):3590-3600. doi: 10.18632/oncotarget.27718. PMID: 33062195; PMCID: PMC7533074. |
| *PAK3* | p21-Activated Kinase 3 | PAK3 promotes the metastasis of hepatocellular carcinoma by regulating EMT process. | Gao Z, Zhong M, Ye Z, Wu Z, Xiong Y, Ma J, Chen H, Zhu Y, Yang Y, Zhao Y, Zhang Z. PAK3 promotes the metastasis of hepatocellular carcinoma by regulating EMT process. *J Cancer*. 2022 Jan 1;13(1):153-161. doi: 10.7150/jca.61918. PMID: 34976179; PMCID: PMC8692680. |
| *PAX7* | Paired Box Protein Pax-7 | Pax7 dysregulation in the muscle microenvironment promotes cancer cachexia. | He WA, Berardi E, Cardillo VM, Acharyya S, Aulino P, Thomas-Ahner J, Wang J, Bloomston M, Muscarella P, Nau P, Shah N, Butchbach ME, Ladner K, Adamo S, Rudnicki MA, Keller C, Coletti D, Montanaro F, Guttridge DC. NF-κB-mediated Pax7 dysregulation in the muscle microenvironment promotes cancer cachexia. *J Clin Invest*. 2013 Nov;123(11):4821-35. doi: 10.1172/JCI68523. PMID: 24084740; PMCID: PMC3809785. |
| *PER1* | Period Circadian Regulator 1 | Ectopic expression of Per1 and Per2 in human cancer cells is associated with cell cycle arrest and apoptosis. | S. Gery, H. P. Koeffler, The role of circadian regulation in cancer. *Cold Spring Harb Symp Quant Biol* **72**, 459-464 (2007). |
| *PIGR* | Polymeric Immunoglobulin Receptor | PIGR is a prognostic biomarker for hepatocellular carcinoma and correlated with poor prognosis in patients with osteosarcoma and pancreatic cancer. | Ai J, Tang Q, Wu Y, Xu Y, Feng T, Zhou R, Chen Y, Gao X, Zhu Q, Yue X, Pan Q, Xu S, Li J, Huang M, Daugherty-Holtrop J, He Y, Xu HE, Fan J, Ding J, Geng M. The role of polymeric immunoglobulin receptor in inflammation-induced tumor metastasis of human hepatocellular carcinoma. *J Natl Cancer Inst*. 2011 Nov 16;103(22):1696-712. doi: 10.1093/jnci/djr360. Epub 2011 Oct 24. PMID: 22025622; PMCID: PMC3216966.  N. Sphyris, S. A. Mani, pIgR: frenemy of inflammation, EMT, and HCC progression. *J Natl Cancer Inst* **103**, 1644-1645 (2011). |
| *PRICKLE2* | Prickle Planar Cell Polarity Protein 2 | PRICKLE2 is differentially expressed in breast cancer | Jaeger, H., and Delacretaz, J. (1953). Carcinoma en cuirasse of the breast and prickle cell epithelioma of the vulva. *Dermatologica* 107, 257–259. doi: 10.1159/000256802 |
| *PPP1R36* | Protein Phosphatase 1 Regulatory Subunit 36 | Germline-enriched PPP1R36 promotes autophagy. | Zhang, Q., Gao, M., Zhang, Y. *et al.* The germline-enriched Ppp1r36 promotes autophagy. *Sci Rep* **6**, 24609 (2016). https://doi.org/10.1038/srep24609 |
| *RARRES1* | Retinoic Acid Receptor Responder Protein 1 (TIG1) | TIG1 (RARRES1) promotes the development and progression of inflammatory breast cancer through activation of Axl kinase. | X. Wang, H. Saso, T. Iwamoto, W. Xia, Y. Gong, L. Pusztai, W. A. Woodward, J. M. Reuben, S. L. Warner, D. J. Bearss, G. N. Hortobagyi, M. C. Hung, N. T. Ueno, TIG1 promotes the development and progression of inflammatory breast cancer through activation of Axl kinase. *Cancer Res* **73**, 6516-6525 (2013). |
| *RGCC* | Regulator of Cell Cycle (also known as Response Gene to Complement-32, RGC-32) | RGC-32 is expressed by tumor cells and plays a dual role in cancer, functioning as either a tumor promoter by endorsing malignancy initiation, progression, invasion, metastasis, and angiogenesis, or as a tumor suppressor. RGCC also plays a key role in mediating cancer-related inflammation in a wide variety of cancer types. | S. I. Vlaicu, A. Tatomir, V. Rus, H. Rus, Role of C5b-9 and RGC-32 in Cancer. *Front Immunol* **10**, 1054 (2019).  Zhao P., Gao D., Wang Q., Song B., Shao Q., Sun J., Ji C., Li X., Li P., Qu X. Response gene to complement 32 (RGC-32) expression on M2-polarized and tumor-associated macrophages is M-CSF-dependent and enhanced by tumor-derived IL-4. *Cell Mol Immunol.* 2015; 12(6): 692–9. |
| *RGS1* | Regulator of G Protein Signaling 1 | RGS1 is a new marker and promoting factor for T-Cell exhaustion in multiple cancers. | Y. Bai, M. Hu, Z. Chen, J. Wei, H. Du, Single-Cell Transcriptome Analysis Reveals RGS1 as a New Marker and Promoting Factor for T-Cell Exhaustion in Multiple Cancers. *Front Immunol* **12**, 767070 (2021). |
| *RPGR* | X-linked Retinitis Pigmentosa GTPase Regulator | RPGR is a ciliary protein, which acts as a guanine exchange factor for RAB8A and plays a role in protein trafficking in photoreceptors. Mutations in RPGR have been reported in breast cancer. | Anand M, Khanna H. Ciliary transition zone (TZ) proteins RPGR and CEP290: role in photoreceptor cilia and degenerative diseases. *Expert Opin Ther Targets.* 2012 Jun;16(6):541-51. doi: 10.1517/14728222.2012.680956. Epub 2012 May 7. PMID: 22563985; PMCID: PMC3724338.  Pongor L, Kormos M, Hatzis C, Pusztai L, Szabó A, Győrffy B. A genome-wide approach to link genotype to clinical outcome by utilizing next generation sequencing and gene chip data of 6,697 breast cancer patients. *Genome Med.* 2015 Oct 16;7:104. doi: 10.1186/s13073-015-0228-1. PMID: 26474971; PMCID: PMC4609150. |
| *SH3PXD2B* | SH3 and PX Domains 2B | SH3PXD2B expression was up-regulated in Hepatocellular carcinoma (HCC) tissues may promote the invasion and metastasis of HCC and be a valuable therapeutic target and biomarker for treatment and prognosis of HCC. | Kui X, Wang Y, Zhang C, Li H, Li Q, Ke Y, Wang L. Prognostic value of SH3PXD2B (Tks4) in human hepatocellular carcinoma: a combined multi-omics and experimental study. *BMC Med Genomics.* 2021 Apr 28;14(1):115. doi: 10.1186/s12920-021-00963-6. PMID: 33906640; PMCID: PMC8080318. |
| *SLC6A14* | Solute Carrier Family 6 Member 14 | Aberrant SLC6A14 expression promotes proliferation and metastasis of colorectal cancer via enhancing the JAK2/STAT3 pathway. | H. Mao, J. Sheng, J. Jia, C. Wang, S. Zhang, H. Li, F. He, Aberrant SLC6A14 Expression Promotes Proliferation and Metastasis of Colorectal Cancer via Enhancing the JAK2/STAT3 Pathway. *Onco Targets Ther* **14**, 379-392 (2021). |
| *SMIM3* | Small Integral Membrane Protein 3 | SMIM3 is one of the 11 gene signature that is robust in segregating tumors by Extracapsular spread (ECS) status. In node negative patients, patients harboring this ECS signature had a significantly worse overall survival. | W. Wang, W. K. Lim, H. S. Leong, F. T. Chong, T. K. Lim, D. S. Tan, B. T. Teh, N. G. Iyer, An eleven gene molecular signature for extra-capsular spread in oral squamous cell carcinoma serves as a prognosticator of outcome in patients without nodal metastases. *Oral Oncol* **51**, 355-362 (2015). |
| *SPATA18* | Spermatogenesis Associated 18 | SPATA18 expression predicts favorable clinical outcome in colorectal cancer. | A. Sugimura-Nagata, A. Koshino, K. Nagao, A. Nagano, M. Komura, A. Ueki, M. Ebi, N. Ogasawara, T. Tsuzuki, K. Kasai, S. Takahashi, K. Kasugai, S. Inaguma, SPATA18 Expression Predicts Favorable Clinical Outcome in Colorectal Cancer. *Int J Mol Sci* **23**, (2022). |
| *STX1B* | Syntaxin 1B | Syntaxin-1 is a markers of breast tumors with neuroendocrine features. | S. Turkevi-Nagy, A. Bathori, J. Bocz, L. Krenacs, G. Cserni, B. Kovari, Syntaxin-1 and Insulinoma-Associated Protein 1 Expression in Breast Neoplasms with Neuroendocrine Features. *Pathol Oncol Res* **27**, 1610039 (2021). |
| *TMOD1* | Ttropomodulin 1 | NF-κB regulates TMOD1 expression. TMOD1 elevation is associated with enhanced breast tumor growth in a mouse tumor xenograft model and in a 3D type I collagen culture. | Ito-Kureha T, Koshikawa N, Yamamoto M, Semba K, Yamaguchi N, Yamamoto T, Seiki M, Inoue J. Tropomodulin 1 expression driven by NF-κB enhances breast cancer growth. *Cancer Res.* 2015 Jan 1;75(1):62-72. doi: 10.1158/0008-5472.CAN-13-3455. Epub 2014 Nov 14. PMID: 25398440. |
| *ZBTB16* | Zinc Finger and BTB Domain Containing 16 | ZBTB16 inhibits breast cancer proliferation and metastasis through upregulating ZBTB28 and antagonizing BCL6. | J. He, M. Wu, L. Xiong, Y. Gong, R. Yu, W. Peng, L. Li, L. Li, S. Tian, Y. Wang, Q. Tao, T. Xiang, BTB/POZ zinc finger protein ZBTB16 inhibits breast cancer proliferation and metastasis through upregulating ZBTB28 and antagonizing BCL6/ZBTB27. *Clin Epigenetics* **12**, 82 (2020). |
| **lncRNA** | | | |
| *AC006924.1* | N/A* | Not Reported. |  |
| *AC036176.3* | N/A* | Not Reported. |  |
| *AC044784.1* | N/A* | Not Reported. |  |
| *LINC02015* | Non-Protein Coding RNA 2015 | It is a novel prognostic marker to predict survival and TMZ-chemoradiation response in Glioblastoma multiform patients after surgery. | W. Z. Gao, L. M. Guo, T. Q. Xu, Y. H. Yin, F. Jia, Identification of a multidimensional transcriptome signature for survival prediction of postoperative glioblastoma multiforme patients. *J Transl Med* **16**, 368 (2018). |
| *LINC02562* | Long Intergenic Non-Protein Coding RNA 2562 | LINC02562 is differentially expressed in lung adenocarcinoma and lung squamous cell carcinoma compared to normal tissues. | https://ccsm.uth.edu/lncRNAfunc/gene_search_result.cgi?page=page&type=quick_search&quick_search=ENSG00000260265 |
| *NFIA_AS2* | NFIA Antisense RNA 2 | NFIA-AS2 promotes glioma progression and its high expression is closely correlated with poor prognosis. | Xin J, Zhao YH, Zhang XY, Tian LQ. LncRNA NFIA-AS2 promotes glioma progression through modulating the miR-655-3p/ZFX axis. *Hum Cell.* 2020 Oct;33(4):1273-1280. doi: 10.1007/s13577-020-00408-9. Epub 2020 Aug 10. PMID: 32779154. |
| *SEPT7P6* | Septin 7 Pseudogene 6 | Not Reported. |  |
| *SNHG6* | Small Nucleolar RNA Host Gene 6 | High expression level of SNHG6 is associated with tumor progression and poor prognosis. It promotes cancer development by miRNA sponging. | Khan K, Irfan M, Sattar AA, Faiz MB, Rahman AU, Athar H, Calina D, Sharifi-Rad J, Cho WC. LncRNA SNHG6 role in clinicopathological parameters in cancers. *Eur J Med Res.* 2023 Sep 21;28(1):363. doi: 10.1186/s40001-023-01358-2. PMID: 37735423; PMCID: PMC10515066.  Wang, Hs., Zhang, W., Zhu, Hl. et al. Long noncoding RNA SNHG6 mainly functions as a competing endogenous RNA in human tumors. *Cancer Cell Int* 20, 219 (2020). |
| **sncRNA** | | | |
| *SNORA21* | Small Nucleolar RNA, H/ACA Box 21 | SNORA21 suppresses tumorigenesis of gallbladder cancer and colorectal cancer (CRC). It might serve as an important prognostic biomarker in CRC and gastric cancer. | Y. Qin, Y. Zhou, A. Ge, L. Chang, H. Shi, Y. Fu, Q. Luo, Overexpression of SNORA21 suppresses tumorigenesis of gallbladder cancer in vitro and in vivo. *Biomed Pharmacother* **118**, 109266 (2019).  Yoshida K, Toden S, Weng W, Shigeyasu K, Miyoshi J, Turner J, Nagasaka T, Ma Y, Takayama T, Fujiwara T, Goel A. SNORA21 - An Oncogenic Small Nucleolar RNA, with a Prognostic Biomarker Potential in Human Colorectal Cancer. *EBioMedicine*. 2017 Aug;22:68-77. doi: 10.1016/j.ebiom.2017.07.009. Epub 2017 Jul 12. PMID: 28734806; PMCID: PMC5552212.  Liu CX, Qiao XJ, Xing ZW, Hou MX. The SNORA21 expression is upregulated and acts as a novel independent indicator in human gastric cancer prognosis. *Eur Rev Med Pharmacol Sci.* 2018 Sep;22(17):5519-5524. doi: 10.26355/eurrev_201809_15812. PMID: 30229823. |
| *SNORD123* | Small Nucleolar RNA, C/D Box 123 | SNORD123 is overexpressed in ameloblastoma, gastric, colorectal and lung cancer and differentially enriched in a basal-like subtype of triple-negative breast cancer. | H. J. Ferreira, H. Heyn, C. Moutinho, M. Esteller, CpG island hypermethylation-associated silencing of small nucleolar RNAs in human cancer. *RNA Biol* **9**, 881-890 (2012). |
| *SNORD44* | Small Nucleolar RNA, C/D Box 44 | The overexpression of SNORD44 from an adenovirus vector was found to have an antitumor effect in colorectal cancer cells. | Yuan S, Wu Y, Wang Y, Chen J, Chu L. An Oncolytic Adenovirus Expressing SNORD44 and GAS5 Exhibits Antitumor Effect in Colorectal Cancer Cells. *Hum Gene Ther.* 2017 Aug;28(8):690-700. doi: 10.1089/hum.2017.041. Epub 2017 May 19. PMID: 28530127. |
| **miRNA** | | | |
| *let−7b−5p* | MicroRNA Let-7b | microRNA let-7 functions as a tumor suppressor in cancer cells. | X. Han, Y. Chen, N. Yao, H. Liu, Z. Wang, MicroRNA let-7b suppresses human gastric cancer malignancy by targeting ING1. *Cancer Gene Ther* **22**, 122-129 (2015).  Chirshev E, Oberg KC, Ioffe YJ, Unternaehrer JJ. Let-7 as biomarker, prognostic indicator, and therapy for precision medicine in cancer. *Clin Transl Med.* 2019 Aug 28;8(1):24. doi: 10.1186/s40169-019-0240-y. PMID: 31468250; PMCID: PMC6715759. |
| *miR−15b−5p* | MicroRNA 15b-5p | miR-15b-5p promotes cell growth and metastasis of breast cancer. | B. Wu, G. Liu, Y. Jin, T. Yang, D. Zhang, L. Ding, F. Zhou, Y. Pan, Y. Wei, miR-15b-5p Promotes Growth and Metastasis in Breast Cancer by Targeting HPSE2. *Front Oncol* **10**, 108 (2020). |
| *miR−24−3p* | MicroRNA 24b-3p | High level of miR-24-3p is associated with shorter survival rate and metastasis in breast cancer. | A. Khodadadi-Jamayran, B. Akgol-Oksuz, Y. Afanasyeva, A. Heguy, M. Thompson, K. Ray, A. Giro-Perafita, I. Sanchez, X. Wu, D. Tripathy, A. Zeleniuch-Jacquotte, A. Tsirigos, F. J. Esteva, Prognostic role of elevated mir-24-3p in breast cancer and its association with the metastatic process. *Oncotarget* **9**, 12868-12878 (2018). |
| *miR−125a−5p* | MicroRNA 125a | Dysregulation of miR-125a promotes tumorigenesis. | J. K. Wang, Z. Wang, G. Li, MicroRNA-125 in immunity and cancer. *Cancer Lett* **454**, 134-145 (2019). |
| *miR−125b−5p* | MicroRNA 125b-5p | miR-125b is a tumor suppressor gene in esophageal squamous cell carcinoma. | L. L. Mei, W. J. Wang, Y. T. Qiu, X. F. Xie, J. Bai, Z. Z. Shi, miR-125b-5p functions as a tumor suppressor gene partially by regulating HMGA2 in esophageal squamous cell carcinoma. *PLoS One* **12**, e0185636 (2017).  Wang S, Huang J, Lyu H, Lee CK, Tan J, Wang J, Liu B. Functional cooperation of miR-125a, miR-125b, and miR-205 in entinostat-induced downregulation of erbB2/erbB3 and apoptosis in breast cancer cells. *Cell Death Dis.* 2013 Mar 21;4(3):e556. doi: 10.1038/cddis.2013.79. PMID: 23519125; PMCID: PMC3615747. |
| *miR−199a−5p* | MicroRNA 199a-5p | miR-199a inhibits the growth and metastasis of colorectal cancer cells. | L. Tian, M. Chen, Q. He, Q. Yan, C. Zhai, MicroRNA‑199a‑5p suppresses cell proliferation, migration and invasion by targeting ITGA3 in colorectal cancer. *Mol Med Rep* **22**, 2307-2317 (2020). |
| **LINE and Repeat Elements** | | | |
| *(ATGAA)n* | N/A* | Not Reported. |  |
| *(ATTCC)n* | N/A* | Not Reported. |  |
| *L1M3b* | N/A* | L1 element L1M3b is downregulated in hepatocellular carcinoma. It modulates chromatin assembly. | G. Karakulah, C. Yandim, Identification of differentially expressed genomic repeats in primary hepatocellular carcinoma and their potential links to biological processes and survival. *Turk J Biol* **45**, 599-612 (2021). |
| *(CTCT)n* | N/A* | Not Reported. |  |
| *(TCCATTCCAT)n* | N/A* | Not Reported. |  |
| *(TGAGA)n* | N/A* | Not Reported. |  |
| **PBMC Samples** | | | |
| **Protein-coding genes** | | | |
| AC022826.2 | N/A* | Not Reported. |  |
| *AGTPBP1* | ATP/GTP Binding Carboxypeptidase 1 | AGTPBP1 expression is correlated with cancer progression and immune infiltration in lung cancer. | Kwak HJ, Gil M, Chae HS, Seok J, Soundrarajan N, Saha SK, Kim A, Park KS, Park C, Cho SG. Expression of ATP/GTP Binding Protein 1 Has Prognostic Value for the Clinical Outcomes in Non-Small Cell Lung Carcinoma. *J Pers Med.* 2020 Dec 2;10(4):263. doi: 10.3390/jpm10040263. PMID: 33276627; PMCID: PMC7761608. |
| *ANKRD12* | Ankyrin Repeat Domain 12 | ANKRD12 mRNA were down regulated in CRC tumor tissues and low ANKRD12 expression was correlated with liver metastasis and poor survival of CRC patients. | Bai R, Li D, Shi Z, Fang X, Ge W, Zheng S. Clinical significance of Ankyrin repeat domain 12 expression in colorectal cancer. *J Exp Clin Cancer Res.* 2013 May 29;32(1):35. doi: 10.1186/1756-9966-32-35. PMID: 23718802; PMCID: PMC3689078. |
| *ANKRD17* | Ankyrin Repeat Domain 17 | In HeLa cells Ankrd17 contributed to pro-inflammatory responses induced by Shigella flexneri.  Ankrd17 is a positive regulator of inflammatory responses via the retinoic acid-inducible gene-I (RIG-I)-like receptor (RLR) signaling pathway. | Menning M, Kufer TA. A role for the Ankyrin repeat containing protein Ankrd17 in Nod1- and Nod2-mediated inflammatory responses. *FEBS Lett.* 2013 Jul 11;587(14):2137-42. doi: 10.1016/j.febslet.2013.05.037. Epub 2013 May 24. PMID: 23711367.  Li M, Qi W, Chang Q, Chen R, Zhen D, Liao M, Wen J, Deng Y. Influenza A virus protein PA-X suppresses host Ankrd17-mediated immune responses. *Microbiol Immunol.* 2021 Jan;65(1):48-59. doi: 10.1111/1348-0421.12863. Epub 2021 Jan 7. PMID: 33241870. |
| *ARID2* | AT-rich Interactive Domain-Containing Protein 2 | ARID2 has a tumor suppressor role in *TP53* mutated oral squamous cell carcinoma. *ARID2* deficiency promotes tumor progression and is associated with higher sensitivity to chemotherapy in lung cancer.  The loss of Arid2 impairs hematopoietic stem cells differentiation ability, and this effect may be mediated through upregulation of inflammatory pathways. | Shukla, P., Dange, P., Mohanty, B.S. *et al.* ARID2 suppression promotes tumor progression and upregulates cytokeratin 8, 18 and β-4 integrin expression in *TP53*-mutated tobacco-related oral cancer and has prognostic implications. *Cancer Gene Ther* 29, 1908–1917 (2022).  Moreno T, Monterde B, González-Silva L, Betancor-Fernández I, Revilla C, Agraz-Doblas A, Freire J, Isidro P, Quevedo L, Blanco R, Montes-Moreno S, Cereceda L, Astudillo A, Casar B, Crespo P, Morales Torres C, Scaffidi P, Gómez-Román J, Salido E, Varela I. ARID2 deficiency promotes tumor progression and is associated with higher sensitivity to chemotherapy in lung cancer. *Oncogene.* 2021 Apr;40(16):2923-2935. doi: 10.1038/s41388-021-01748-y. Epub 2021 Mar 19. PMID: 33742126; PMCID: PMC7610680.  Bluemn T, Schmitz J, Chen Y, Zheng Y, Zhang Y, Zheng S, Burns R, DeJong J, Christiansen L, Izaguirre-Carbonell J, Wang D, Zhu N. Arid2 regulates hematopoietic stem cell differentiation in normal hematopoiesis. *Exp Hematol.* 2021 Feb;94:37-46. doi: 10.1016/j.exphem.2020.12.004. Epub 2020 Dec 17. PMID: 33346030; PMCID: PMC10041880. |
| *ARID4B* | AT-rich Interactive Domain-Containing Protein 4B | ARID4B knockdown suppresses PI3K/AKT signaling and induces apoptosis in human glioma cells. | Luo SM, Tsai WC, Tsai CK, Chen Y, Hueng DY. *ARID4B* Knockdown Suppresses PI3K/AKT Signaling and Induces Apoptosis in Human Glioma Cells. *Onco Targets Ther.* 2021 Mar 10;14:1843-1855. doi: 10.2147/OTT.S286837. PMID: 33732001; PMCID: PMC7956898. |
| *ATAD2B* | ATPase Family AAA Domain Containing 2B | The abnormal expression of ATAD2 was found to be closely related to the occurrence and development of a variety of tumors. | Yalin Tong, Jinbei Li, Mengle Peng, Qilan Qian, Wen Shi, Zefeng Chen, Bin Liu. ATAD2 drives colorectal cancer progression by regulating TRIM25 expression via a positive feedback loop with E2F transcriptional factors,  *Biochemical and Biophysical Research Communications*,  Volume 594, 2022, Pages 146-152. |
| *ATP2B1* | Plasma Membrane Calcium-Transporting ATPase 1 | The protein encoded by this gene is an enzyme that removes bivalent calcium ions from eukaryotic cells against very large concentration gradients and play a critical role in intracellular calcium homeostasis.  ATP2B1 is upregulated in tumorigenic breast cancer cell lines. | Olson S, Wang MG, Carafoli E, Strehler EE, McBride OW. Localization of two genes encoding plasma membrane Ca2(+)-transporting ATPases to human chromosomes 1q25-32 and 12q21-23. *Genomics.* 1991 Apr;9(4):629-41. doi: 10.1016/0888-7543(91)90356-j. PMID: 1674727.  W. J. Lee, S. J. Roberts-Thomson, N. A. Holman, F. J. May, G. M. Lehrbach, G. R. Monteith, Expression of plasma membrane calcium pump isoform mRNAs in breast cancer cell lines. *Cell Signal* **14**, 1015-1022 (2002). |
| *BAZ1A* | Bromodomain Adjacent to Zinc Finger Domain 1A | BAZ1A acting as a novel regulator of cellular senescence in both normal and cancer cells, indicating a new target for potential cancer treatment. | Li X, Ding D, Yao J, Zhou B, Shen T, Qi Y, Ni T, Wei G. Chromatin remodeling factor BAZ1A regulates cellular senescence in both cancer and normal cells. *Life Sci.* 2019 Jul 15;229:225-232. doi: 10.1016/j.lfs.2019.05.023. Epub 2019 May 11. PMID: 31085244. |
| *BRAF* | Serine/threonine-Protein Kinase B-raf | BRAF plays a role in regulating the MAP kinase/ERKs signaling pathway, which affects cell division, differentiation, and secretion. BRAF mutations occur in human tumors such as melanoma, colorectal and thyroid cancers. | A. Gorden, I. Osman, W. Gai, D. He, W. Huang, A. Davidson, A. N. Houghton, K. Busam, D. Polsky, Analysis of BRAF and N-RAS mutations in metastatic melanoma tissues. *Cancer Res* **63**, 3955-3957 (2003). |
| *CCDC91* | Coiled-Coil Domain Containing 91 | The CCDC91 isoform acts as a [competitive endogenous RNA](https://www.sciencedirect.com/topics/biochemistry-genetics-and-molecular-biology/competitive-endogenous-rna) by sponging MIR890 to increase RUNX2 expression. | Nakajima M, Koido M, Guo L, Terao C, Ikegawa S. A novel CCDC91 isoform associated with ossification of the posterior longitudinal ligament of the spine works as a non-coding RNA to regulate osteogenic genes. *Am J Hum Genet.* 2023 Apr 6;110(4):638-647. doi: 10.1016/j.ajhg.2023.03.004. Epub 2023 Mar 28. PMID: 36990086; PMCID: PMC10119134. |
| *CDH8* | Cadherin-8 | CHD8 gene expression is significantly associated with genes involved in the Wnt/β‑catenin pathway and in the cell cycle in gastric cancer. | Sawada G, Ueo H, Matsumura T, Uchi R, Ishibashi M, Mima K, Kurashige J, Takahashi Y, Akiyoshi S, Sudo T, Sugimachi K, Doki Y, Mori M, Mimori K. CHD8 is an independent prognostic indicator that regulates Wnt/β-catenin signaling and the cell cycle in gastric cancer. *Oncol Rep.* 2013 Sep;30(3):1137-42. doi: 10.3892/or.2013.2597. Epub 2013 Jul 8. PMID: 23835524. |
| *CLASP2* | Cytoplasmic Linker Associated Protein 2 | CLASP2 is involved in the EMT and progression of bladder urothelial cancer.  The interaction among SOCS3, CLIP-170, and CLASP2 is critical for modulation of endothelial inflammation and lung injury. | Zhu, B., Qi, L., Liu, S. *et al.* CLASP2 is involved in the EMT and early progression after transurethral resection of the bladder tumor. *BMC Cancer* 17, 105 (2017). <https://doi.org/10.1186/s12885-017-3101-3>  Karki P, Ke Y, Zhang CO, Li Y, Tian Y, Son S, Yoshimura A, Kaibuchi K, Birukov KG, Birukova AA. SOCS3-microtubule interaction via CLIP-170 and CLASP2 is critical for modulation of endothelial inflammation and lung injury. *J Biol Chem.* 2021 Jan-Jun;296:100239. doi: 10.1074/jbc.RA120.014232. Epub 2021 Jan 9. PMID: 33372035; PMCID: PMC7949054. |
| *CNTN1* | Contactin 1 | CNTN1 is involved in cancer cell invasion, migration, metastasis, and chemoresistance by promoting epithelial-mesenchymal transition and mediating several signal transduction pathways. | Liang Y, Ma C, Li F, Nie G, Zhang H. The Role of Contactin 1 in Cancers: What We Know So Far. *Front Oncol.* 2020 Oct 29;10:574208. doi: 10.3389/fonc.2020.574208. PMID: 33194679; PMCID: PMC7658624. |
| *COL12A1* | Collagen Alpha-1(XII) Chain | COL12A1 is upregulated in breast cancer and promotes migration, invasion and metastasis. High expression of COL12A1 is associated with poor immunotherapy response. | Xu YH, Deng JL, Wang LP, Zhang HB, Tang L, Huang Y, Tang J, Wang SM, Wang G. Identification of Candidate Genes Associated with Breast Cancer Prognosis. *DNA Cell Biol.* 2020 Jul;39(7):1205-1227. doi: 10.1089/dna.2020.5482. Epub 2020 May 22. PMID: 32456464.  Yan Y, Liang Q, Liu Y, Zhou S, Xu Z. COL12A1 as a prognostic biomarker links immunotherapy response in breast cancer. *Endocr Relat Cancer.* 2023 Apr 3;30(5):e230012. doi: 10.1530/ERC-23-0012. PMID: 36877531. |
| *DNAJC1* | DnaJ Homolog Subfamily C Member 1 | DNAJC1 had a predictive value for the prognosis of HCC. Knockdown of DNAJC1 may inhibit HCC cell proliferation, migration and invasion and promote the HCC cell apoptosis through p53 and EMT signaling pathways. | Fan YC, Meng ZY, Zhang CS, Wei DW, Wei WS, Xie XD, Huang ML, Jiang LH. DNAJ heat shock protein family member C1 can regulate proliferation and migration in hepatocellular carcinoma. *PeerJ.* 2023 Jul 26;11:e15700. doi: 10.7717/peerj.15700. PMID: 37520264; PMCID: PMC10386825. |
| *EDIL3* | EGF Like Repeats and Discoidin Domains 3, also known as DEL1 | EDIL3 acts as a pro-angiogenic factor, mediator of the immune and anti-inflammatory response, and a regulator of endothelial cell adhesion and migration. | Choi EY, Chavakis E, Czabanka MA, Langer HF, Fraemohs L, Economopoulou M, Kundu RK, Orlandi A, Zheng YY, Prieto DA, Ballantyne CM, Constant SL, Aird WC, Papayannopoulou T, Gahmberg CG, Udey MC, Vajkoczy P, Quertermous T, Dimmeler S, Weber C, Chavakis T. Del-1, an endogenous leukocyte-endothelial adhesion inhibitor, limits inflammatory cell recruitment. *Science.* 2008 Nov 14;322(5904):1101-4. doi: 10.1126/science.1165218. |
| *ELF2* | E74-Like Factor 2 | ELF2 promoted the proliferation of osteosarcoma cells. | Zhang J, Hou W, Jia J, et al. MiR-409-3p regulates cell proliferation and tumor growth by targeting E74-like factor 2 in osteosarcoma. FEBS Open Bio. 2017;7(3):348–357. doi: 10.1002/2211-5463.12177 |
| *FAM172A* | Family with Sequence Similarity 172 Member A | FAM172A inhibits EMT in pancreatic cancer via ERK-MAPK signaling. | Chen Y, Liu P, Shen D, Liu H, Xu L, Wang J, Shen D, Sun H, Wu H. FAM172A inhibits EMT in pancreatic cancer via ERK-MAPK signaling. *Biol Open.* 2020 Feb 7;9(2):bio048462. doi: 10.1242/bio.048462. PMID: 31988090; PMCID: PMC7044457. |
| *FBXO11* | F-box Protein 11 | FBXO11 is a candidate tumor suppressor in the leukemic transformation of myelodysplastic syndrome. It governs macrophage cell death and inflammation in response to bacterial toxins. | Schieber M, Marinaccio C, Bolanos LC, Haffey WD, Greis KD, Starczynowski DT, Crispino JD. FBXO11 is a candidate tumor suppressor in the leukemic transformation of myelodysplastic syndrome. *Blood Cancer J.* 2020 Oct 6;10(10):98. doi: 10.1038/s41408-020-00362-7. PMID: 33024076; PMCID: PMC7538974.  Jeon Y, Chow SH, Stuart I, Weir A, Yeung AT, Hale C, Sridhar S, Dougan G, Vince JE, Naderer T. FBXO11 governs macrophage cell death and inflammation in response to bacterial toxins. *Life Sci Alliance.* 2023 Mar 28;6(6):e202201735. doi: 10.26508/lsa.202201735. PMID: 36977592; PMCID: PMC10053445. |
| *FBXW7* | F-box/WD Repeat-Containing Protein 7 | FBXW7α (FBXW7) attenuates inflammatory signaling. It is frequently inactivated in human lung, colon, and hematopoietic cancers. The loss of FBXW7 can serve as an independent prognostic marker and is significantly correlated with the resistance of tumor cells to chemotherapeutic agents and poorer disease outcomes. | Balamurugan K, Sharan S, Klarmann KD, Zhang Y, Coppola V, Summers GH, Roger T, Morrison DK, Keller JR, Sterneck E. FBXW7α attenuates inflammatory signalling by downregulating C/EBPδ and its target gene Tlr4. *Nat Commun.* 2013;4:1662. doi: 10.1038/ncomms2677. PMID: 23575666; PMCID: PMC3625980.  Fan, J., Bellon, M., Ju, M. *et al.* Clinical significance of FBXW7 loss of function in human cancers. *Mol Cancer* **21**, 87 (2022). https://doi.org/10.1186/s12943-022-01548-2 |
| *FNIP1* | Folliculin-Interacting Protein 1 | Folliculin-interacting proteins Fnip1 and Fnip2 play critical roles in kidney tumor suppression. | Hasumi H, Baba M, Hasumi Y, Lang M, Huang Y, Oh HF, Matsuo M, Merino MJ, Yao M, Ito Y, Furuya M, Iribe Y, Kodama T, Southon E, Tessarollo L, Nagashima K, Haines DC, Linehan WM, Schmidt LS. Folliculin-interacting proteins Fnip1 and Fnip2 play critical roles in kidney tumor suppression in cooperation with Flcn. *Proc Natl Acad Sci U S A.* 2015 Mar 31;112(13):E1624-31. doi: 10.1073/pnas.1419502112. Epub 2015 Mar 16. PMID: 25775561; PMCID: PMC4386336. |
| *GJA1* | Gap Junction Alpha-1 Protein (also known as connexin43) | GJA1 is implicated in many biological processes including inflammation. GJA1 expression has been associated with a wide variety of cancers, including breast cancer. It is associated with recurrence, metastasis, and reduced survival in cancer. GJA1 is also expressed in eosinophils and T cells to promote their maturation and activation. | Bonacquisti EE, Nguyen J. Connexin 43 (Cx43) in cancer: Implications for therapeutic approaches via gap junctions. *Cancer Lett.* 2019 Feb 1;442:439-444. doi: 10.1016/j.canlet.2018.10.043. Epub 2018 Nov 22. PMID: 30472182.  Chevallier, D., Carette, D., Segretain, D. *et al.* Connexin 43 a check-point component of cell proliferation implicated in a wide range of human testis diseases. Cell. *Mol. Life Sci.* **70**, 1207–1220 (2013).  Vliagoftis H, Ebeling C, Ilarraza R, Mahmudi-Azer S, Abel M, Adamko D, Befus AD, Moqbel R. Connexin 43 expression on peripheral blood eosinophils: role of gap junctions in transendothelial migration. *Biomed Res Int.* 2014;2014:803257. doi: 10.1155/2014/803257. Epub 2014 Jul 6. PMID: 25110696; PMCID: PMC4109672. |
| *GOLGB1* | Golgin B1 | Mutants of *GOLGB1* can promote HCC progression, providing new mechanistic insights on the HCC relapse. | Choi J., Kim M., Park Y., Im J., Kwon S., Kim H., Goo Woo H., Wang H. Mutations acquired by hepatocellular carcinoma recurrence give rise to an aggressive phenotype. *Oncotarget.* 2017; 8: 22903-22916. |
| *GRID2* | Glutamate Receptor, Ionotropic, Delta 2 | GRID2 was significantly downregulated in endometrial carcinoma. | Chen X, Zhang W, Zhu H, Lin F. Development and Validation of a 5-Gene Autophagy-Based Prognostic Index in Endometrial Carcinoma. *Med Sci Monit.* 2021 Feb 12;27:e928949. doi: 10.12659/MSM.928949. PMID: 33577492; PMCID: PMC7885295. |
| *GRM8* | Metabotropic Glutamate Receptor 8 | GRM8 signaling pathway could serve as potential targets of squamous cell lung cancer carrying GRM8 activating variants. | Zhang P, Kang B, Xie G, Li S, Gu Y, Shen Y, Zhao X, Ma Y, Li F, Si J, Wang J, Chen J, Yang H, Xu X, Yang Y. Genomic sequencing and editing revealed the GRM8 signaling pathway as potential therapeutic targets of squamous cell lung cancer. *Cancer Lett.* 2019 Feb 1;442:53-67. doi: 10.1016/j.canlet.2018.10.035. Epub 2018 Oct 27. PMID: 30391781. |
| *GTF2F2* | General Transcription Factor IIF Subunit 2 | Not Reported. |  |
| *HSD11B1* | 11β-Hydroxysteroid Dehydrogenase Type 1, also known as Cortisone Reductase | HSD11B1 is an enzyme that converts inert glucocorticoids into active forms in tissues. HSD11B1 inhibition in combination with PD-1 blockade augmented the production of interferon-γ by T cells. High levels of HSD11B1, predominantly expressed by tumor-associated macrophages, are associated with poor responses to immune checkpoint inhibitor therapy in patients with advanced melanomas. | Martins Nascentes Melo L, Herrera-Rios D, Hinze D, Löffek S, Oezel I, Turiello R, Klein J, Leonardelli S, Westedt IV, Al-Matary Y, Egea-Rodriguez S, Brenzel A, Bau M, Sucker A, Hadaschik E, Wirsdörfer F, Hanenberg H, Uhlenbrock N, Rauh D, Poźniak J, Rambow F, Marine JC, Effern M, Glodde N, Schadendorf D, Jablonska J, Hölzel M, Helfrich I. Glucocorticoid activation by HSD11B1 limits T cell-driven interferon signaling and response to PD-1 blockade in melanoma. *J Immunother Cancer.* 2023 Apr 7;11(4):e004150. doi: 10.1136/jitc-2021-004150. PMID: 37028818; PMCID: PMC10083881. |
| *JMJD1C* | Jumonji Domain Containing 1C | JMJD1C functions as an oncogenic factor for AML by promoting cell survival and self-renewal. It is also overexpressed in colon cancer tissues and increases colon cancer metastasis via the inactivation of the ATF2 pathway.  JMJD1C could enhance M1 macrophage polarization to inhibit the onset of glioma. Overexpression of JMJD1C induced a decrease in TGFβ 1–3 expression as well as an increase in interleukin (IL)‐1β and IL‐6 expression in LN‐229 cells. | Wang, L. et al. Novel somatic and germline mutations in intracranial germ cell tumours. *Nature* **511**, 241–245 (2014).  Chen, C. et al. Downregulation of histone demethylase JMJD1C inhibits colorectal cancer metastasis through targeting ATF2. *Am. J. Cancer Res.* **8**, 852–865 (2018).  Zhong C, Tao B, Yang F, Xia K, Yang X, Chen L, Peng T, Xia X, Li X, Peng L. Histone demethylase JMJD1C promotes the polarization of M1 macrophages to prevent glioma by upregulating miR-302a. *Clin Transl Med.* 2021 Sep;11(9):e424. doi: 10.1002/ctm2.424. PMID: 34586733; PMCID: PMC8473479. |
| *MAPK8* | Mitogen-Activated Protein Kinase 8 (also known as JNK1) | Dysregulation of the JNK signaling cascade is associated with inflammatory disorders and cancers.  Mice with myeloid cell-specific JNK deficiency exhibit reduced hepatic inflammation and suppression of both hepatitis and hepatocellular carcinoma. These data identify myeloid cells as a site of pro-inflammatory signaling by JNK that can promote liver pathology. | Hammouda MB, Ford AE, Liu Y, Zhang JY. The JNK Signaling Pathway in Inflammatory Skin Disorders and Cancer. *Cells.* 2020 Apr 2;9(4):857. doi: 10.3390/cells9040857. PMID: 32252279; PMCID: PMC7226813.  Han MS, Barrett T, Brehm MA, Davis RJ. Inflammation Mediated by JNK in Myeloid Cells Promotes the Development of Hepatitis and Hepatocellular Carcinoma. *Cell Rep.* 2016 Apr 5;15(1):19-26. doi: 10.1016/j.celrep.2016.03.008. Epub 2016 Mar 24. PMID: 27052181; PMCID: PMC4826851. |
| *MAP3K2* | Mitogen-Activated Protein Kinase Kinase Kinase 2 (also known as MEKK2) | The mRNA expression of MAP3K2 was positively associated with survival in kidney renal clear cell carcinoma and sarcoma. MAP3K2 expression was correlated with decreased survival in breast cancer and kidney renal papillary cell carcinoma.  MEKK2-MKK7 pathway JNK activation by antigen cross-linking is dependent on the, and cytokine production in mast cells is regulated in part by the signaling complex MEKK2-MEK5-ERK5. | Khoa Nguyen, Minh N. Tran, Andrew Rivera, Thomas Cheng, Gabrielle O. Windsor, Abraham B. Chabot, Jane E. Cavanaugh, Bridgette M. Collins-Burow, Sean B. Lee, David H. Drewry, Patrick T. Flaherty, Matthew E. Burow. MAP3K Family Review and Correlations with Patient Survival Outcomes in Various Cancer Types. *Front. Biosci. (Landmark Ed)* **2022**, 27(5), 167.  Chayama K, Papst PJ, Garrington TP, Pratt JC, Ishizuka T, Webb S, Ganiatsas S, Zon LI, Sun W, Johnson GL, Gelfand EW. Role of MEKK2-MEK5 in the regulation of TNF-alpha gene expression and MEKK2-MKK7 in the activation of c-Jun N-terminal kinase in mast cells. *Proc Natl Acad Sci U S A.* 2001 Apr 10;98(8):4599-604. doi: 10.1073/pnas.081021898. Epub 2001 Mar 27. PMID: 11274363; PMCID: PMC31880. |
| *MED13L* | Mediator Complex Subunit 13L | *MED13L* play key roles in gene transcription and in the inflammatory response through participation in the Wnt signaling pathway. It can also participate in the inflammatory response and positively regulate the secretion of pro-inflammatory factors by regulating and activating transcription factor 4 expression.  MED13L has oncogenic functions in NSCLC. NSCLC patients with a relatively high MED13L expression exhibited significantly shorter survival time (PFS and OS) compared to cases with low MED13L expression. | Iwasaki, Y., Suganami, T., Hachiya, R., Shirakawa, I., Kim-Saijo, M., Tanaka, M., et al. (2014). Activating transcription factor 4 links metabolic stress to interleukin-6 expression in macrophages. *Diabetes* 63, 152–161. doi:10.2337/db13-0757  Asadollahi, R., Zweier, M., Gogoll, L., Schiffmann, R., Sticht, H., Steindl, K., et al. (2017). Genotype-phenotype evaluation of MED13L defects in the light of a novel truncating and a recurrent missense mutation. *Eur. J. Med. Genet.* 60, 451–464. doi:10.1016/j.ejmg.2017.06.004  Zhang N, Song Y, Xu Y, Liu J, Shen Y, Zhou L, Yu J, Yang M. MED13L integrates Mediator-regulated epigenetic control into lung cancer radiosensitivity. *Theranostics.* 2020 Jul 23;10(20):9378-9394. doi: 10.7150/thno.48247. PMID: 32802198; PMCID: PMC7415817. |
| *MEMO1* | Mediator of Cell Motility 1 | Memo1 is a negative regulator of cell proliferation in HPV-positive cervical cancer cell lines. | Trejo-Cerro O, Massimi P, Broniarczyk J, Myers M, Banks L. Repression of Memo1, a Novel Target of Human Papillomavirus Type 16 E7, Increases Cell Proliferation in Cervical Cancer Cells. *J Virol.* 2022 Oct 26;96(20):e0122922. doi: 10.1128/jvi.01229-22. Epub 2022 Oct 5. PMID: 36197110; PMCID: PMC9599245. |
| *METTL21A* | Methyltransferase 21A, HSPA Lysine | Not Reported. |  |
| *MGAM* | Maltase-Glucoamylase | MGAM tends to express in luminal A breast cancers. But there is no significant association with patient survival. | Xu S, Feng Y, Zhao S. Proteins with Evolutionarily Hypervariable Domains are Associated with Immune Response and Better Survival of Basal-like Breast Cancer Patients. *Comput Struct Biotechnol J.* 2019 Mar 19;17:430-440. doi: 10.1016/j.csbj.2019.03.008. PMID: 30996822; PMCID: PMC6451114. |
| *MKLN1* | Muskelin | Expression of MKLN1 in pancreatic tumors was correlated with overall survival in patients with pancreatic adenocarcinoma. MKLN1 expression changes may be important for the initiation or progression of human pancreatic adenocarcinoma. | https://osf.io/preprints/osf/36c5p |
| *MNAT1* | CDK-activating Kinase Assembly Factor | MNAT1 promotes proliferation and the chemo-resistance of osteosarcoma cell to cisplatin. | Qiu, C., Su, W., Shen, N. *et al.* MNAT1 promotes proliferation and the chemo-resistance of osteosarcoma cell to cisplatin through regulating PI3K/Akt/mTOR pathway. *BMC Cancer* 20, 1187 (2020). |
| *NAMPT* | Nicotinamide Phosphoribosyl-Transferase | Overexpression of NAMPT has been observed in colorectal, ovarian, breast, gastric, and prostate. | R. E. Shackelford, K. Mayhall, N. M. Maxwell, E. Kandil, D. Coppola, Nicotinamide phosphoribosyltransferase in malignancy: a review. *Genes Cancer* **4**, 447-456 (2013). |
| *NEK7* | NIMA Related Kinase 7 | NEK7 is a key player in cell cycle progression required for cytokinesis. | L. O'Regan, A. M. Fry, The Nek6 and Nek7 protein kinases are required for robust mitotic spindle formation and cytokinesis. *Mol Cell Biol* **29**, 3975-3990 (2009). |
| *NFKB1* | Nuclear Factor NF-Kappa-B p105 Subunit | NFkB1 promotes inflammation-associated cancer. | J. A. DiDonato, F. Mercurio, M. Karin, NF-kappaB and the link between inflammation and cancer. *Immunol Rev* **246**, 379-400 (2012). |
| *OLR1* | Oxidized Low-Density Lipoprotein Receptor 1 | LOX-1 overexpression is a significant prognosis of tumor progression in advanced-stage prostate cancer. | F. Wan, X. Qin, G. Zhang, X. Lu, Y. Zhu, H. Zhang, B. Dai, G. Shi, D. Ye, Oxidized low-density lipoprotein is associated with advanced-stage prostate cancer. *Tumour Biol* **36**, 3573-3582 (2015). |
| *OXSR1* | Oxidative Stress Responsive Kinase 1 (also known as OSR1) | High expression of OXSR1 as a predictive biomarker for poor prognosis and lymph node metastasis in breast cancer.  Osr1 regulates hepatic inflammation and cell survival in the progression of non-alcoholic fatty liver disease.  Osr1 regulates macrophage-mediated liver inflammation in nonalcoholic fatty liver disease progression. | Li Y, Qin J, Wu J, Dai X, Xu J. High expression of OSR1 as a predictive biomarker for poor prognosis and lymph node metastasis in breast cancer. *Breast Cancer Res Treat.* 2020 Jul;182(1):35-46. doi: 10.1007/s10549-020-05671-w. Epub 2020 May 18. PMID: 32424721.  Zhou, Y., Liu, Z., Lynch, E.C. et al. Osr1 regulates hepatic inflammation and cell survival in the progression of non-alcoholic fatty liver disease. *Lab Invest* **101**, 477–489 (2021).  Liu L, Zhou Y, Liu Z, Li J, Hu L, He L, Gao G, Kidd B, Walsh A, Jiang R, Wu C, Zhang K, Xie L. Osr1 Regulates Macrophage-mediated Liver Inflammation in Nonalcoholic Fatty Liver Disease Progression. *Cell Mol Gastroenterol Hepatol.* 2023;15(5):1117-1133. doi: 10.1016/j.jcmgh.2022.12.010. Epub 2022 Dec 27. PMID: 36581078; PMCID: PMC10036739. |
| *PAPD4* | PAP Associated Domain Containing 4 (also known as TENT2) | TENT2 (PAPD4) contributes to guanylation and uridylation on mature miRNAs. PAPD4 is significantly upregulated in ccRCC tumors. | Yang A, Bofill-De Ros X, Stanton R, Shao TJ, Villanueva P, Gu S. TENT2, TUT4, and TUT7 selectively regulate miRNA sequence and abundance. *Nat Commun.* 2022 Sep 7;13(1):5260. doi: 10.1038/s41467-022-32969-8. PMID: 36071058; PMCID: PMC9452540.  Kajdasz A, Majer W, Kluzek K, Sobkowiak J, Milecki T, Derebecka N, Kwias Z, Bluyssen HAR, Wesoly J. Identification of RCC Subtype-Specific microRNAs-Meta-Analysis of High-Throughput RCC Tumor microRNA Expression Data. *Cancers (Basel).* 2021 Feb 1;13(3):548. doi: 10.3390/cancers13030548. PMID: 33535553; PMCID: PMC7867039. |
| *PICALM* | Phosphatidylinositol Binding Clathrin Assembly Protein | PICALM promotes colorectal cancer progression through ERK/MAPK signaling pathway. | Zhang X, Liu T, Huang J, He J. PICALM exerts a role in promoting CRC progression through ERK/MAPK signaling pathway. *Cancer Cell Int.* 2022 May 2;22(1):178. doi: 10.1186/s12935-022-02577-z. PMID: 35501863; PMCID: PMC9063212. |
| *PPP3CA* | Protein Phosphatase 3 Catalytic Subunit Alpha | PPP3CA is significantly associated with reduced overall survival in ovarian serous carcinoma. | Xin, B., Ji, KQ., Liu, YS. *et al.* Higher expression of calcineurin predicts poor prognosis in unique subtype of ovarian cancer. *J Ovarian Res* 12, 75 (2019). |
| *PTPRD* | Receptor-Type Tyrosine-Protein Phosphatase Delta | PTPRD as a tumor suppressor that is involved in the development of glioblastoma multiforme and multiple human cancers. | Veeriah S, Brennan C, Meng S, Singh B, Fagin JA, Solit DB, Paty PB, Rohle D, Vivanco I, Chmielecki J, Pao W, Ladanyi M, Gerald WL, Liau L, Cloughesy TC, Mischel PS, Sander C, Taylor B, Schultz N, Major J, Heguy A, Fang F, Mellinghoff IK, Chan TA. The tyrosine phosphatase PTPRD is a tumor suppressor that is frequently inactivated and mutated in glioblastoma and other human cancers. *Proc Natl Acad Sci U S A.* 2009 Jun 9;106(23):9435-40. doi: 10.1073/pnas.0900571106. Epub 2009 May 28. PMID: 19478061; PMCID: PMC2687998. |
| *RAPGEF2* | Rap Guanine Nucleotide Exchange Factor 2 | RAPGEF2 degradation is required for HGF-induced cell migration. | Magliozzi R, Low TY, Weijts BG, Cheng T, Spanjaard E, Mohammed S, van Veen A, Ovaa H, de Rooij J, Zwartkruis FJ, Bos JL, de Bruin A, Heck AJ, Guardavaccaro D. Control of epithelial cell migration and invasion by the IKKβ- and CK1α-mediated degradation of RAPGEF2. *Dev Cell.* 2013 Dec 9;27(5):574-85. doi: 10.1016/j.devcel.2013.10.023. Epub 2013 Nov 27. PMID: 24290981. |
| *RAP1A* | Ras-related Protein Rap-1A | High expression of Ras-related protein 1A promotes an aggressive phenotype in colorectal cancer via PTEN/FOXO3/CCND1 pathway.  Rap1 induces cytokine production in pro-inflammatory macrophages through NFκB signaling and is highly expressed in human atherosclerotic lesions. | Liu, L., Yan, X., Wu, D. *et al.* High expression of Ras-related protein 1A promotes an aggressive phenotype in colorectal cancer via PTEN/FOXO3/CCND1 pathway. *J Exp Clin Cancer Res* 37, 178 (2018).  Cai Y, Sukhova GK, Wong HK, Xu A, Tergaonkar V, Vanhoutte PM, Tang EH. Rap1 induces cytokine production in pro-inflammatory macrophages through NFκB signaling and is highly expressed in human atherosclerotic lesions. *Cell Cycle.* 2015;14(22):3580-92. doi: 10.1080/15384101.2015.1100771. PMID: 26505215; PMCID: PMC4825742. |
| *RFWD2* | E3 Ubiquitin-Protein Ligase RFWD2 (also known as COP1) | RFWD2 knockdown significantly suppresses cell proliferation and induces the apoptosis of human hepatocellular carcinoma cells.  E3 ubiquitin ligase COP1/RFWD2 was revealed to control pro-inflammatory gene expression in microglia by promoting degradation of the transcription factor c/EBPβ. | Lee YH, Andersen JB, Song HT, Judge AD, Seo D, Ishikawa T, et al. Definition of Ubiquitination Modulator COP1 as a Novel Therapeutic Target in Human Hepatocellular Carcinoma. *Cancer Res* (2010) 70:8264–9. doi:  10.1158/0008-5472.CAN-10-0749  Ndoja A, Reja R, Lee SH, Webster JD, Ngu H, Rose CM, et al. Ubiquitin ligase COP1 suppresses neuroinflammation by degrading c/EBPbeta in microglia. *Cell*. 2020;182:1156–69 e1112. |
| *RPAP2* | RNA Polymerase II-Associated Protein 2 | RPAP2 functions as a gatekeeper to inhibit d pre-initiation complex assembly and transcription initiation and suggests a transcription checkpoint. | Wang X, Qi Y, Wang Z, Wang L, Song A, Tao B, Li J, Zhao D, Zhang H, Jin Q, Jiang YZ, Chen FX, Xu Y, Chen X. RPAP2 regulates a transcription initiation checkpoint by inhibiting assembly of pre-initiation complex. *Cell Rep.* 2022 Apr 26;39(4):110732. doi: 10.1016/j.celrep.2022.110732. PMID: 35476980. |
| *RYR2* | Ryanodine Receptor 2 | The RYR2 protein functions as the major component of a calcium channel located in the sarcoplasmic reticulum that supplies ions to the cardiac muscle during systole. RYR2 mutation is associated with tumor mutation burden, prognosis, and antitumor immunity in patients with esophageal adenocarcinoma. | Liu Z, Liu L, Jiao D, Guo C, Wang L, Li Z, Sun Z, Zhao Y, Han X. Association of RYR2 Mutation With Tumor Mutation Burden, Prognosis, and Antitumor Immunity in Patients With Esophageal Adenocarcinoma. *Front Genet.* 2021 May 17;12:669694. doi: 10.3389/fgene.2021.669694. PMID: 34079583; PMCID: PMC8166246. |
| *SAMSN1* | SAM Domain-Containing Protein SAMSN-1 | Downregulation of *SAMSN1* transcription may affect the progression and recurrence of gastric cancer. | M. Kanda, D. Shimizu, S. Sueoka, S. Nomoto, H. Oya, H. Takami, K. Ezaka, R. Hashimoto, Y. Tanaka, D. Kobayashi, C. Tanaka, S. Yamada, T. Fujii, G. Nakayama, H. Sugimoto, M. Koike, M. Fujiwara, Y. Kodera, Prognostic relevance of SAMSN1 expression in gastric cancer. *Oncol Lett* **12**, 4708-4716 (2016). |
| *SBF2* | SET Binding Factor 2 (also known as myotubularin-related protein 13) | Not Reported. |  |
| *SDC2* | Syndecan 2 | SDC2 regulates cell proliferation, cell migration and cell-matrix interactions via its receptor for extracellular matrix proteins. | B. Jang, A. Kim, J. Hwang, H. K. Song, Y. Kim, E. S. Oh, Emerging Role of Syndecans in Extracellular Matrix Remodeling in Cancer. *J Histochem Cytochem* **68**, 863-870 (2020). |
| *SERPINB2* | Plasminogen Activator Inhibitor Type 2 | Elevated SerpinB2 expression has been linked with prolonged survival, decreased metastasis or decreased tumour growth in a number of cancer types including breast cancer and PDAC. | D. R. Croucher, D. N. Saunders, S. Lobov, M. Ranson, Revisiting the biological roles of PAI2 (SERPINB2) in cancer. *Nat Rev Cancer* **8**, 535-545 (2008). |
| *SLIT3* | Slit Homolog 3 Protein | Slit3 acts as a tumor suppressor in hepatocellular carcinoma (HCC) by repressing the tumor growth and thus tumor progression. Low Slit3 level indicates a poor response of HCC cells to chemotherapy. | Ng L, Chow AKM, Man JHW, Yau TCC, Wan TMH, Iyer DN, Kwan VHT, Poon RTP, Pang RWC, Law WL. Suppression of Slit3 induces tumor proliferation and chemoresistance in hepatocellular carcinoma through activation of GSK3β/β-catenin pathway. *BMC Cancer*. 2018 Jun 1;18(1):621. doi: 10.1186/s12885-018-4326-5. PMID: 29859044; PMCID: PMC5984734. |
| *SMURF2* | E3 Ubiquitin-Protein Ligase | The expression of Smurf2 was downregulated in HCC specimens and affected the survival of patients. Smurf2 inhibited the EMT of HCC by enhancing Smad2 ubiquitin-dependent proteasome degradation.  Smurf2 affects inflammation and collagen processing in cutaneous wounds by down-regulating TGF-β/Smad3 signaling. | Song D, Li S, Ning L, Zhang S, Cai Y. Smurf2 suppresses the metastasis of hepatocellular carcinoma via ubiquitin degradation of Smad2. *Open Med (Wars).* 2022 Feb 24;17(1):384-396. doi: 10.1515/med-2022-0437. PMID: 35509688; PMCID: PMC8874264.  Stuelten CH, Melis N, Subramanian B, Tang Y, Kimicata M, Fisher JP, Weigert R, Zhang YE. Smurf2 Regulates Inflammation and Collagen Processing in Cutaneous Wound Healing through Transforming Growth Factor-β/Smad3 Signaling. *Am J Pathol.* 2022 Dec;192(12):1699-1711. doi: 10.1016/j.ajpath.2022.08.002. Epub 2022 Sep 3. PMID: 36063900; PMCID: PMC9765313. |
| *SNX13* | Sorting Nexin-13 | SNX13 is a favorable prognostic marker in renal cancer. | https://www.proteinatlas.org/ENSG00000071189-SNX13/pathology |
| *STAG1* | Stromal Antigen 1 | STAG1 inactivation inhibits the proliferation of STAG2 mutated but not wild-type bladder cancer and Ewing sarcoma cell lines. | van der Lelij P, Lieb S, Jude J, Wutz G, Santos CP, Falkenberg K, Schlattl A, Ban J, Schwentner R, Hoffmann T, Kovar H, Real FX, Waldman T, Pearson MA, Kraut N, Peters JM, Zuber J, Petronczki M. Synthetic lethality between the cohesin subunits STAG1 and STAG2 in diverse cancer contexts. Elife. 2017 Jul 10;6:e26980. doi: 10.7554/eLife.26980. PMID: 28691904; PMCID: PMC5531830. |
| *STRN3* | Striatin 3 | STRN3-RARA is a new fusion gene for acute promyelocytic leukemia. | Zhang Q, Li H, Chen X, Gu F, Zhang L, Zhang L, Chen T, Chen Q, Meng W, Wu Y, Chang H, Liu T, Chen C, Ma H, Liu Y. Identifying STRN3-RARA as a new fusion gene for acute promyelocytic leukemia. *Blood*. 2023 Oct 26;142(17):1494-1499. doi: 10.1182/blood.2023020619. PMID: 37624915. |
| *TANK* | TRAF Family Member Associated NFKB Activator | Deregulated expression of TANK in glioblastomas triggers pro-tumorigenic ERK1/2 and AKT signaling pathways.  TANK is a protein with dual functions in activating NF-kB that is indispensable for immune responses and inflammatory processes, as well as for activating survival and proinflammatory genes within the tumor microenvironment.  TANK is also an adaptor protein that interacts with canonical IKKs (NEMO and IKKγ) and IKK-related kinases (TBK1 and IKKϵ) to modulate NF-κB and TLR-induced antiviral pathways and prevent autoimmunity. | Stellzig, J., Chariot, A., Shostak, K. *et al.* Deregulated expression of TANK in glioblastomas triggers pro-tumorigenic ERK1/2 and AKT signaling pathways. *Oncogenesis* 2, e79 (2013).  Cheng G, Baltimore D. TANK, a co-inducer with TRAF2 of TNF- and CD 40L-mediated NF-kappaB activation. *Genes Dev.* 1996 Apr 15;10(8):963-73. doi: 10.1101/gad.10.8.963. PMID: 8608943.  Chariot A, Leonardi A, Muller J, Bonif M, Brown K, Siebenlist U. Association of the adaptor TANK with the I kappa B kinase (IKK) regulator NEMO connects IKK complexes with IKK epsilon and TBK1 kinases. *J Biol Chem.* 2002 Oct 4;277(40):37029-36. doi: 10.1074/jbc.M205069200. Epub 2002 Jul 19. PMID: 12133833.  Guo B, Cheng G. Modulation of the interferon antiviral response by the TBK1/IKKi adaptor protein TANK. *J Biol Chem.* 2007 Apr 20;282(16):11817-26. doi: 10.1074/jbc.M700017200. Epub 2007 Feb 26. PMID: 17327220. |
| *TCF12* | Transcription Factor 12 | TCF12 promotes the tumorigenesis and metastasis of hepatocellular carcinoma via upregulation of CXCR4 expression. | Yang J, Zhang L, Jiang Z, Ge C, Zhao F, Jiang J, Tian H, Chen T, Xie H, Cui Y, Yao M, Li H, Li J. TCF12 promotes the tumorigenesis and metastasis of hepatocellular carcinoma via upregulation of CXCR4 expression. *Theranostics.* 2019 Aug 12;9(20):5810-5827. doi: 10.7150/thno.34973. PMID: 31534521; PMCID: PMC6735379. |
| *THSD4* | Thrombospondin Type 1 Domain Containing 4, also known as ADAMTSL6 | In a non-small cell lung cancer (NSCLC) cohort, THSD4 (ADAMTSL6) and Papilin (PAPLN) were associated with immune checkpoint inhibitor (ICI) sensitivity. | Zhang X, Yang W, Chen K, Zheng T, Guo Z, Peng Y, Yang Z. The potential prognostic values of the ADAMTS-like protein family: an integrative pan-cancer analysis. *Ann Transl Med.* 2021 Oct;9(20):1562. doi: 10.21037/atm-21-4946. PMID: 34790768; PMCID: PMC8576672. |
| *TMEM212* | Transmembrane Protein 212 | TMEM211 promotes tumor progression and metastasis in colon cancer. | Chang YF, Wang HH, Shu CW, Tsai WL, Lee CH, Chen CL, Liu PF. TMEM211 Promotes Tumor Progression and Metastasis in Colon Cancer. *Curr Issues Mol Biol.* 2023 May 24;45(6):4529-4543. doi: 10.3390/cimb45060287. PMID: 37367036; PMCID: PMC10297151. |
| *USP47* | Ubiquitin Specific Peptidase 47 | USP47 has a role in inflammation by regulating inflammasome activation and the release of proinflammatory cytokines, such as IL-1β and IL-18.  USP47 exhibits critical functions in cell growth and genome integrity that are closely related to cancer. | Palazon-Riquelme, P. et al. USP7 and USP47 deubiquitinases regulate NLRP3 inflammasome activation. EMBO Rep. **19**, e44766 (2018).  Shin, S.C., Park, J., Kim, K.H. *et al.* Structural and functional characterization of USP47 reveals a hot spot for inhibitor design. *Commun Biol* **6**, 970 (2023). |
| *VNN3* | Vascular Non-Inflammatory Molecule 3 | VNN3 is a potential novel predictive biomarker for clear cell renal cell carcinoma. | M. Ha, H. Jeong, J. S. Roh, B. Lee, D. Lee, M. E. Han, S. O. Oh, D. H. Sohn, Y. H. Kim, VNN3 is a potential novel biomarker for predicting prognosis in clear cell renal cell carcinoma. *Anim Cells Syst (Seoul)* **23**, 112-117 (2019). |
| *VPS54* | VPS54 Subunit of GARP Complex | VPS54 is a prognostic biomarker, high expression is unfavorable in liver cancer. | https://www.proteinatlas.org/ENSG00000143952-VPS54/pathology/liver+cancer |
| *WAC* | WW Domain Containing Adaptor With Coiled-Coil | WAC is a candidate tumor suppressor gene in colorectal cancer. | Christopher R. Clark, Caitlin Conboy, Makayla Maile, Callie Janik, Julia Hatler, Robert Cormier, David Largaespada, Timothy K. Starr. WAC: A candidate tumor suppressor gene in colorectal cancer. [abstract]. In: Proceedings of the 107th Annual Meeting of the American Association for Cancer Research; 2016 Apr 16-20; New Orleans, LA. Philadelphia (PA): AACR; Cancer Res 2016;76(14 Suppl):Abstract nr 3665. |
| *WDR74* | WD Repeat-Containing Protein 74 | WDR74 promotes proliferation and metastasis in colorectal cancer cells through regulating the Wnt/β-catenin signaling pathway.  WDR74 facilitates TGF-β/Smad pathway activation to promote M2 macrophage polarization. | Cai Z, Mei Y, Jiang X, Shi X. WDR74 promotes proliferation and metastasis in colorectal cancer cells through regulating the Wnt/β-catenin signaling pathway. *Open Life Sci.* 2021 Sep 6;16(1):920-929. doi: 10.1515/biol-2021-0096. PMID: 34553072; PMCID: PMC8422980.  Geng K, Ma X, Jiang Z, Gu J, Huang W, Wang W, Xu Y, Xu Y. WDR74 facilitates TGF-β/Smad pathway activation to promote M2 macrophage polarization and diabetic foot ulcer wound healing in mice. *Cell Biol Toxicol.* 2023 Aug;39(4):1577-1591. doi: 10.1007/s10565-022-09748-8. Epub 2022 Aug 19. PMID: 35982296. |
| *ZNF644* | Zing Finger Protein 644 | Among breast cancer patients who did receive radiotherapy, those with high ZNF644 expression had significantly longer survival than patients with low level expression. | Yan D, Shen M, Du Z, Cao J, Tian Y, Zeng P, Tang Z. Developing ZNF Gene Signatures Predicting Radiosensitivity of Patients with Breast Cancer. *J Oncol.* 2021 Aug 31;2021:9255494. doi: 10.1155/2021/9255494. PMID: 34504527; PMCID: PMC8423582. |
| **lncRNA** | | | |
| *AC019117.2* | N/A* | AC019117.2 infers a positive correlation with clinical stages of stage I to stage IV gastric cancer. | N. M. Beeraka, H. Gu, N. Xue, Y. Liu, H. Yu, J. Liu, K. Chen, V. N. Nikolenko, R. Fan, Testing lncRNAs signature as clinical stage-related prognostic markers in gastric cancer progression using TCGA database. *Exp Biol Med (Maywood)* **247**, 658-671 (2022). |
| *AC130895.1* | N/A* | Not Reported. |  |
| *AC122718.1* | N/A* | Not Reported. |  |
| *AC092957.1* | N/A* | Not Reported. |  |
| *AC010967.1* | N/A* | Not Reported. |  |
| *AC091862.1* | N/A* | Not Reported. |  |
| *AC114316.2* | N/A* | Not Reported. |  |
| *AC018697.1* | N/A* | Not Reported. |  |
| *AC246680.1* | N/A* | Not Reported. |  |
| *AC022639.1* | N/A* | Not Reported. |  |
| *AC007402.1* | N/A* | Not Reported. |  |
| *AC021351.1* | N/A* | Not Reported. |  |
| *AC103409.1* | N/A* | Not Reported. |  |
| *AC009264.1* | N/A* | Not Reported. |  |
| *AC005323.2* | N/A* | Not Reported. |  |
| *AC087565.3* | N/A* | Not Reported. |  |
| *AC099520.1* | N/A* | Not Reported. |  |
| *AC011306.1* | N/A* | Not Reported. |  |
| *AC106798.1* | N/A* | Not Reported. |  |
| *ADAMTS9_AS2* | ADAMTS9-AS2 Isoform 3 | ADAMTS9_AS2 inhibits tumor growth of breast cancer. | K. Ni, Z. Huang, Y. Zhu, D. Xue, Q. Jin, C. Zhang, C. Gu, The lncRNA ADAMTS9-AS2 Regulates RPL22 to Modulate TNBC Progression via Controlling the TGF-beta Signaling Pathway. *Front Oncol* **11**, 654472 (2021). |
| *AF279873.3* | N/A* | Not Reported. |  |
| *AL354793.1* | lncRNA | *AL354793.1* is associated with the prognosis of triple-negative breast cancer (TNBC) patients. | C. N. Fan, L. Ma, N. Liu, Comprehensive analysis of novel three-long noncoding RNA signatures as a diagnostic and prognostic biomarkers of human triple-negative breast cancer. J Cell Biochem **120**, 3185-3196 (2019). |
| *AL357507.1* | N/A* | High expression of AL357507.1 predicts a worse overall survival in clear cell renal cell carcinoma patients. | F. Yang, C. Liu, G. Zhao, L. Ge, Y. Song, Z. Chen, Z. Liu, K. Hong, L. Ma, Long non-coding RNA LINC01234 regulates proliferation, migration and invasion via HIF-2alpha pathways in clear cell renal cell carcinoma cells. *PeerJ* **8**, e10149 (2020). |
| *AL359313.1* | N/A* | Not Reported. |  |
| *AL445250.1* | N/A* | Not Reported. |  |
| *AL513323.1* | N/A* | Not Reported. |  |
| *AP005203.1* | N/A* | Not Reported. |  |
| *AP000769.1* | N/A* | Not Reported. |  |
| *C5orf17* | Chromosome 5 Open Reading Frame 17 | Not Reported. |  |
| *FAM133DP* | Family With Sequence Similarity 133 Member D, Pseudogene | Not Reported. |  |
| *GCSHP3* | Glycine Cleavage System Protein H Pseudogene 3 | GCSH-antisense regulation determines cancerous cell viability. | Adamus A, Müller P, Nissen B, Kasten A, Timm S, Bauwe H, Seitz G, Engel N. GCSH antisense regulation determines breast cancer cells' viability. *Sci Rep.* 2018 Oct 18;8(1):15399. doi: 10.1038/s41598-018-33677-4. PMID: 30337557; PMCID: PMC6193953.  Mo X, Wu Y, Chen L, Zhai M, Gao Z, Hu K, Guo J. Global expression profiling of metabolic pathway-related lncRNAs in human gastric cancer and the identification of RP11-555H23.1 as a new diagnostic biomarker. *J Clin Lab Anal.* 2019 Feb;33(2):e22692. doi: 10.1002/jcla.22692. Epub 2018 Oct 15. PMID: 30320481; PMCID: PMC6818562. |
| *G2E3_AS1* | G2E3 Antisense RNA 1 | Breast cancer patients with higher expression of G2E3-AS1 had poorer prognosis. | X. Li, Z. Ma, L. Mei, Cuproptosis-related gene SLC31A1 is a potential predictor for diagnosis, prognosis and therapeutic response of breast cancer. *Am J Cancer Res* **12**, 3561-3580 (2022). |
| *LINC00276* | Long Intergenic Non-Protein Coding RNA 276 | LINC00276 and other 5 lncRNAs are constructed for predicting colorectal cancer prognosis. | X. Zhao, J. Liu, S. Liu, F. Yang, E. Chen, Construction and Validation of an Immune-Related Prognostic Model Based on TP53 Status in Colorectal Cancer. *Cancers (Basel)* **11**, (2019). |
| *LINC01192* | Long Intergenic Non-Protein Coding RNA 1192 | High expression of LINC01192 is associated with low survival possibility in triple-negative breast cancer. | Li XX, Wang LJ, Hou J, Liu HY, Wang R, Wang C, Xie WH. Identification of Long Noncoding RNAs as Predictors of Survival in Triple-Negative Breast Cancer Based on Network Analysis. *Biomed Res Int.* 2020 Mar 3;2020:8970340. doi: 10.1155/2020/8970340. PMID: 32190687; PMCID: PMC7073484. |
| *LINC01830* | Long Intergenic Non-Protein Coding RNA 1830 | Not Reported. |  |
| *LINC01924* | Long Intergenic Non-Protein Coding RNA 1924 | Not Reported. |  |
| *MALAT1* | Metastasis Associated Lung Adenocarcinoma Transcript 1. Also known as NEAT2 (Noncoding Nuclear-Enriched Abundant Transcript 2) | MALAT1 is a prognostic marker for metastasis and patient survival in non–small cell lung carcinoma (NSCLC). | P. Ji, S. Diederichs, W. Wang, S. Boing, R. Metzger, P. M. Schneider, N. Tidow, B. Brandt, H. Buerger, E. Bulk, M. Thomas, W. E. Berdel, H. Serve, C. Muller-Tidow, MALAT-1, a novel noncoding RNA, and thymosin beta4 predict metastasis and survival in early-stage non-small cell lung cancer. *Oncogene* **22**, 8031-8041 (2003). |
| *MIR325HG* | MIR325 Host Gene | m^6^A-modified MIR325HG is correlated with worse patient survival in both low-grade glioma and glioblastoma. | K. Xu, Y. Cai, M. Zhang, H. Zou, Z. Chang, D. Li, J. Bai, J. Xu, Y. Li, Pan-cancer characterization of expression and clinical relevance of m(6)A-related tissue-elevated long non-coding RNAs. *Mol Cancer* **20**, 31 (2021). |
| *MIR4300HG* | N/A* | N/A |  |
| *NEAT1* | Nuclear Enriched Abundant Transcript 1 | TNBC tissues have high expression of NEAT1. NEAT1 confers oncogenic role by regulating apoptosis and cell cycle progression in TNBC cells. | V. Y. Shin, J. Chen, I. W. Cheuk, M. T. Siu, C. W. Ho, X. Wang, H. Jin, A. Kwong, Long non-coding RNA NEAT1 confers oncogenic role in triple-negative breast cancer through modulating chemoresistance and cancer stemness. *Cell Death Dis* **10**, 270 (2019). |
| *NR2F2_AS1* | NR2F2 Antisense RNA 1 | NR2F2-AS1 has oncogenic role. Its dysregulation promotes cancerous cell proliferation, dissemination, and migration. | Ghorbanzadeh S, Poor-Ghassem N, Afsa M, Nikbakht M, Malekzadeh K. Long non-coding RNA NR2F2-AS1: its expanding oncogenic roles in tumor progression. *Hum Cell*. 2022 Sep;35(5):1355-1363. doi: 10.1007/s13577-022-00733-1. Epub 2022 Jul 7. PMID: 35796938. |
| *PTCHD1_AS* | PTCHD1 Antisense RNA | Not Reported. |  |
| **PBMC Samples** | | | |
| **sncRNA** | | | |
| *RNU6V* | U6 Small Nuclear Variant Sequence With SNRPE Pseudogene Sequence | Not Reported. |  |
| *SCARNA8* | Small Cajal Body-Specific RNA 8 | Not Reported. |  |
| *SCARNA14* | Small Cajal Body-Specific RNA 14 | Not Reported. |  |
| *SCARNA15* | Small Cajal Body-Specific RNA 15 | Not Reported. |  |
| *SNORA2B* | Small Nucleolar RNA, H/ACA Box 2B | Low expression of SNORA2B is related to poor prognosis of ovarian cancer patients and may play roles as a suppressor gene in ovarian cancer. | W. Zhu, J. Niu, M. He, L. Zhang, X. Lv, F. Liu, L. Jiang, J. Zhang, Z. Yu, L. Zhao, J. Bi, Y. Yan, Q. Wei, H. Huo, Y. Fan, Y. Chen, J. Ding, M. Wei, SNORD89 promotes stemness phenotype of ovarian cancer cells by regulating Notch1-c-Myc pathway. *J Transl Med* **17**, 259 (2019). |
| *SNORA3B* | Small Nucleolar RNA, H/ACA Box 3B | Not Reported. |  |
| *SNORA3C* | Small Nucleolar RNA, H/ACA Box 3C | Not Reported. |  |
| *SCARNA4* | Small Cajal Body-Specific RNA 4 | SCARNA4-derived RNAs is a potential uncharacterized small RNA marker for cancer immunity and clinical outcome across tumors from diverse tissues of origin. | R. D. Chow, S. Chen, Sno-derived RNAs are prevalent molecular markers of cancer immunity. *Oncogene* **37**, 6442-6462 (2018). |
| *SNORA5A* | Small Nucleolar RNA, H/ACA Box 5A | Lung adenocarcinoma patients with high risk score of an expression signature containing SNOU109, SNORA5A, SNORA70, SNORD104 and U3 are significantly related to an unfavorable overall survival. | L. Zhang, M. Xin, P. Wang, Identification of a novel snoRNA expression signature associated with overall survival in patients with lung adenocarcinoma: A comprehensive analysis based on RNA sequencing dataset. *Math Biosci Eng* **18**, 7837-7860 (2021). |
| *SNORA11* | Small Nucleolar RNA, H/ACA Box 11 | SNORA11 is associated with luminal breast cancer. It is also a signature of ameloblastoma. | E. Karkkainen, S. Heikkinen, M. Tengstrom, V. M. Kosma, A. Mannermaa, J. M. Hartikainen, Expression profiles of small non-coding RNAs in breast cancer tumors characterize clinicopathological features and show prognostic and predictive potential. *Sci Rep* **12**, 22614 (2022).  Davanian H, Balasiddaiah A, Heymann R, Sundström M, Redenström P, Silfverberg M, Brodin D, Sällberg M, Lindskog S, Kruger Weiner C, Chen M. Ameloblastoma RNA profiling uncovers a distinct non-coding RNA signature. *Oncotarget*. 2017 Jan 17;8(3):4530-4542. doi: 10.18632/oncotarget.13889. PMID: 27965463; PMCID: PMC5354851. |
| *SNORA13* | Small Nucleolar RNA, H/ACA Box 13 | SNORA13 is involve in doxorubicin resistance in human doxorubicin resistant osteosarcoma cells. | Godel M, Morena D, Ananthanarayanan P, Buondonno I, Ferrero G, Hattinger CM, Di Nicolantonio F, Serra M, Taulli R, Cordero F, Riganti C, Kopecka J. Small Nucleolar RNAs Determine Resistance to Doxorubicin in Human Osteosarcoma. *Int J Mol Sci.* 2020 Jun 24;21(12):4500. doi: 10.3390/ijms21124500. PMID: 32599901; PMCID: PMC7349977. |
| *SNORA15B* | Small Nucleolar RNA, H/ACA Box 15B | Not Reported. |  |
| *SNORA16A* | Small Nucleolar RNA, H/ACA Box 16A | SNORA16A is upregulated in uterine leiomyoma. | T. D. Chuang, Y. Xie, W. Yan, O. Khorram, Next-generation sequencing reveals differentially expressed small noncoding RNAs in uterine leiomyoma. *Fertil Steril* **109**, 919-929 (2018). |
| *SNORA19* | Small Nucleolar RNA, H/ACA Box 19 | SNORA19 promotes cell survival by inhibiting apoptosis and cell death of ovarian cancer cells. | L. Faucher-Giguere, A. Roy, G. Deschamps-Francoeur, S. Couture, R. M. Nottingham, A. M. Lambowitz, M. S. Scott, S. Abou Elela, High-grade ovarian cancer associated H/ACA snoRNAs promote cancer cell proliferation and survival. *NAR Cancer* **4**, zcab050 (2022). |
| *SNORA28* | Small Nucleolar RNA, H/ACA Box 28 | SNORA28 is involve in doxorubicin resistance in human doxorubicin resistant osteosarcoma cells. | M. Godel, D. Morena, P. Ananthanarayanan, I. Buondonno, G. Ferrero, C. M. Hattinger, F. Di Nicolantonio, M. Serra, R. Taulli, F. Cordero, C. Riganti, J. Kopecka, Small Nucleolar RNAs Determine Resistance to Doxorubicin in Human Osteosarcoma. *Int J Mol Sci* **21**, (2020). |
| *SNORA32* | Small Nucleolar RNA, H/ACA Box 32 | Not Reported. |  |
| *SNORA36A* | Small Nucleolar RNA, H/ACA Box 36A | Not Reported. |  |
| *SNORA36B* | Small Nucleolar RNA, H/ACA Box 36B | Not Reported. |  |
| *SNORA41* | Small Nucleolar RNA, H/ACA Box 41 | SNORA41 expression is increased in colorectal cancer. | X. Yang, Y. Li, L. Li, J. Liu, M. Wu, M. Ye, SnoRNAs are involved in the progression of ulcerative colitis and colorectal cancer. *Dig Liver Dis* **49**, 545-551 (2017). |
| *SNORA46* | Small Nucleolar RNA, H/ACA Box 46 | SNORA46 is expressed at higher levels in low grade meningiomas than in higher grade tumors. | A. N. Viaene, B. Zhang, M. Martinez-Lage, C. Xiang, U. Tosi, J. P. Thawani, B. Gungor, Y. Zhu, L. Roccograndi, L. Zhang, R. L. Bailey, P. B. Storm, D. M. O'Rourke, A. C. Resnick, M. S. Grady, N. Dahmane, Transcriptome signatures associated with meningioma progression. *Acta Neuropathol Commun* **7**, 67 (2019). |
| *SNORA53* | Small Nucleolar RNA, H/ACA Box 53 | SNORA53 expression is reduced by RAS stimulation in human hepatocellular carcinoma. | M. McMahon, A. Contreras, M. Holm, T. Uechi, C. M. Forester, X. Pang, C. Jackson, M. E. Calvert, B. Chen, D. A. Quigley, J. M. Luk, R. K. Kelley, J. D. Gordan, R. M. Gill, S. C. Blanchard, D. Ruggero, A single H/ACA small nucleolar RNA mediates tumor suppression downstream of oncogenic RAS. *Elife* **8**, (2019) |
| *SNORA65* | Small Nucleolar RNA, H/ACA Box 65 | High level of SNORA65 is observed in primary lung cancer specimens with associated changes in rRNA pseudouridylation. It is also a signature of ameloblastoma. | C. Cui, Y. Liu, D. Gerloff, C. Rohde, C. Pauli, M. Kohn, D. Misiak, T. Oellerich, S. Schwartz, L. H. Schmidt, R. Wiewrodt, A. Marra, L. Hillejan, F. Bartel, C. Wickenhauser, S. Huttelmaier, S. Gollner, F. Zhou, B. Edemir, C. Muller-Tidow, NOP10 predicts lung cancer prognosis and its associated small nucleolar RNAs drive proliferation and migration. *Oncogene* **40**, 909-921 (2021).  Davanian H, Balasiddaiah A, Heymann R, Sundström M, Redenström P, Silfverberg M, Brodin D, Sällberg M, Lindskog S, Kruger Weiner C, Chen M. Ameloblastoma RNA profiling uncovers a distinct non-coding RNA signature. *Oncotarget*. 2017 Jan 17;8(3):4530-4542. doi: 10.18632/oncotarget.13889. PMID: 27965463; PMCID: PMC5354851. |
| *SNORA69* | Small Nucleolar RNA, H/ACA Box 69 | SNORA69 is significantly associated with PD-L1 expression, intratumoral CD8^+^ T cell abundance, and GZMA levels in pancreatic adenocarcinoma. | R. D. Chow, S. Chen, Sno-derived RNAs are prevalent molecular markers of cancer immunity. *Oncogene* **37**, 6442-6462 (2018). |
| *SNORA75* | Small Nucleolar RNA, H/ACA Box 75 | Overexpression of SNORA75 significantly promotes the proliferation, migration, and invasion of endometrial cells. | P. Wei, H. Wang, Y. Li, R. Guo, Nucleolar small molecule RNA SNORA75 promotes endometrial receptivity by regulating the function of miR-146a-3p and ZNF23. *Aging (Albany NY)* **13**, 14924-14939 (2021). |
| *SNORA81* | Small Nucleolar RNA, H/ACA Box 81 | SNORA81 knockdown inhibits 28S rRNA pseudouridylation and accumulation leading to reduced ovarian cancer cell proliferation and migration. | L. Faucher-Giguere, A. Roy, G. Deschamps-Francoeur, S. Couture, R. M. Nottingham, A. M. Lambowitz, M. S. Scott, S. Abou Elela, High-grade ovarian cancer associated H/ACA snoRNAs promote cancer cell proliferation and survival. *NAR Cancer* **4**, zcab050 (2022). |
| *SNORA105A* | Small Nucleolar RNA, H/ACA Box 105A | Not Reported. |  |
| *SNORD2* | Small Nucleolar RNA, C/D Box 2 | The expression of *SNORD2* is increased in cancerous endometrial tissues compared to paired normal and hyperplastic tissues. | H. J. Bao, X. Chen, X. Liu, W. Wu, Q. H. Li, J. Y. Xian, Y. Zhao, S. Chen, Box C/D snoRNA SNORD89 influences the occurrence and development of endometrial cancer through 2'-O-methylation modification of Bim. *Cell Death Discov* **8**, 309 (2022). |
| *SNORD12C* | Small Nucleolar RNA, C/D Box 12C | SNORD12C is upregulated in colon adenocarcinoma. | L. Huang, X. Z. Liang, Y. Deng, Y. B. Liang, X. Zhu, X. Y. Liang, D. Z. Luo, G. Chen, Y. Y. Fang, H. H. Lan, J. H. Zeng, Prognostic value of small nucleolar RNAs (snoRNAs) for colon adenocarcinoma based on RNA sequencing data. *Pathol Res Pract* **216**, 152937 (2020). |
| *SNORD28* | Small Nucleolar RNA, C/D Box 28 | SNORA28 is upregulated in both lung cancer. | L. Gao, J. Ma, K. Mannoor, M. A. Guarnera, A. Shetty, M. Zhan, L. Xing, S. A. Stass, F. Jiang, Genome-wide small nucleolar RNA expression analysis of lung cancer by next-generation deep sequencing. *Int J Cancer* **136**, E623-629 (2015). |
| *SNORD41* | Small Nucleolar RNA, C/D Box 41 | SNORD41 is upregulated in breast tumor tissue. | E. Karkkainen, S. Heikkinen, M. Tengstrom, V. M. Kosma, A. Mannermaa, J. M. Hartikainen, Expression profiles of small non-coding RNAs in breast cancer tumors characterize clinicopathological features and show prognostic and predictive potential. *Sci Rep* **12**, 22614 (2022). |
| *SNORD72* | Small Nucleolar RNA, C/D Box 72 | SNORA72 is overexpressed in hepatocellular carcinoma. | Mao LH, Chen SY, Li XQ, Xu F, Lei J, Wang QL, Luo LY, Cao HY, Ge X, Ran T, Li X, Zou M, Zhou ZH, Wu XL, He S. LncRNA-LALR1 upregulates small nucleolar RNA SNORD72 to promote growth and invasion of hepatocellular carcinoma. *Aging (Albany NY)*. 2020 Mar 11;12(5):4527-4546. doi: 10.18632/aging.102907. Epub 2020 Mar 11. PMID: 32160589; PMCID: PMC7093170. |
| *SNORD98* | Small Nucleolar RNA, C/D Box 98 | Not Reported. |  |
| *SNORD101* | Small Nucleolar RNA, C/D Box 101 | SNORD101 is highly expressed and upregulated in high-grade serous ovarian carcinoma. | L. Faucher-Giguere, A. Roy, G. Deschamps-Francoeur, S. Couture, R. M. Nottingham, A. M. Lambowitz, M. S. Scott, S. Abou Elela, High-grade ovarian cancer associated H/ACA snoRNAs promote cancer cell proliferation and survival. *NAR Cancer* **4**, zcab050 (2022). |
| **Mapped to hairpin miRNA reference** | | | |
| *MIR15B* | MicroRNA 15b | miR-15b is significantly upregulated in NSCLC tissues and cell lines. It promotes the proliferation and invasion of NSCLC cells. | H. Wang, Y. Zhan, J. Jin, C. Zhang, W. Li, MicroRNA-15b promotes proliferation and invasion of non‑small cell lung carcinoma cells by directly targeting TIMP2. *Oncol Rep* **37**, 3305-3312 (2017). |
| *MIR16−1* | MicroRNA 16-1 | miR-15a/miR-16-1 complex functions as tumor suppressors in chronic lymphocytic leukemia and solid tumors. | D. Bonci, V. Coppola, M. Musumeci, A. Addario, R. Giuffrida, L. Memeo, L. D'Urso, A. Pagliuca, M. Biffoni, C. Labbaye, M. Bartucci, G. Muto, C. Peschle, R. De Maria, The miR-15a-miR-16-1 cluster controls prostate cancer by targeting multiple oncogenic activities. *Nat Med* 14, 1271-1277 (2008). |
| *MIR16−2* | MicroRNA 16-2 | miR-16 inhibits the migration and invasion of ovarian cancer cells. | N. Li, L. Yang, Y. Sun, X. Wu, MicroRNA-16 inhibits migration and invasion via regulation of the Wnt/beta-catenin signaling pathway in ovarian cancer. *Oncol Lett* **17**, 2631-2638 (2019). |
| *MIR21* | MicroRNA 21 | miR-21 is highly expressed in many cancer types including breast cancer, compared to the corresponding normal tissue. miR-21 expression correlated with the histological grade of IBC. | L. Chen, Y. Li, Y. Fu, J. Peng, M. H. Mo, M. Stamatakos, C. B. Teal, R. F. Brem, A. Stojadinovic, M. Grinkemeyer, T. A. McCaffrey, Y. G. Man, S. W. Fu, Role of deregulated microRNAs in breast cancer progression using FFPE tissue. *PLoS One* **8**, e54213 (2013).  Lerebours F, Cizeron-Clairac G, Susini A, Vacher S, Mouret-Fourme E, Belichard C, Brain E, Alberini JL, Spyratos F, Lidereau R, Bieche I. miRNA expression profiling of inflammatory breast cancer identifies a 5-miRNA signature predictive of breast tumor aggressiveness. *Int J Cancer.* 2013 Oct 1;133(7):1614-23. doi: 10.1002/ijc.28171. Epub 2013 Apr 13. PMID: 23526361.  Wang S, Huang J, Lyu H, Lee CK, Tan J, Wang J, Liu B. Functional cooperation of miR-125a, miR-125b, and miR-205 in entinostat-induced downregulation of erbB2/erbB3 and apoptosis in breast cancer cells. *Cell Death Dis.* 2013 Mar 21;4(3):e556. doi: 10.1038/cddis.2013.79. PMID: 23519125; PMCID: PMC3615747. |
| *MIR26B* | MicroRNA 26B | miR-26B inhibits cell proliferation of TNBC, colorectal cancer, and cervical cancer. | Y. Li, Z. Sun, B. Liu, Y. Shan, L. Zhao, L. Jia, Tumor-suppressive miR-26a and miR-26b inhibit cell aggressiveness by regulating FUT4 in colorectal cancer. *Cell Death Dis* **8**, e2892 (2017). |
| *MIR30C1* | MicroRNA 30C1 | miR-30c is a prognostic biomarker in breast cancer. It regulates chemoresistance of breast cancer cells. | J. Bockhorn, R. Dalton, C. Nwachukwu, S. Huang, A. Prat, K. Yee, Y. F. Chang, D. Huo, Y. Wen, K. E. Swanson, T. Qiu, J. Lu, S. Y. Park, M. E. Dolan, C. M. Perou, O. I. Olopade, M. F. Clarke, G. L. Greene, H. Liu, MicroRNA-30c inhibits human breast tumour chemotherapy resistance by regulating TWF1 and IL-11. *Nat Commun* 4, 1393 (2013). |
| *MIR30D* | MicroRNA 30d | microRNA-30d mediates the invasion, migration, and EMT of breast cancer. | M. Han, Y. Wang, G. Guo, L. Li, D. Dou, X. Ge, P. Lv, F. Wang, Y. Gu, microRNA-30d mediated breast cancer invasion, migration, and EMT by targeting KLF11 and activating STAT3 pathway. *J Cell Biochem* **119**, 8138-8145 (2018). |
| *MIR30E* | MicroRNA 30E | miR-30e inhibits tumor growth and chemoresistance via targeting IRS1 in breast cancer. | M. M. Liu, Z. Li, X. D. Han, J. H. Shi, D. Y. Tu, W. Song, J. Zhang, X. L. Qiu, Y. Ren, L. L. Zhen, MiR-30e inhibits tumor growth and chemoresistance via targeting IRS1 in Breast Cancer. *Sci Rep* **7**, 15929 (2017). |
| *MIR103A1* | MicroRNA 103a-1 | MicroRNA-103a-3p promotes cell proliferation and invasion in NSCLC cells.  Promotes Cell Proliferation and Invasion in Non-Small-Cell Lung Cancer Cells  Promotes Cell Proliferation and Invasion in Non-Small-Cell Lung Cancer Cells | H. Li, M. Huhe, J. Lou, MicroRNA-103a-3p Promotes Cell Proliferation and Invasion in Non-Small-Cell Lung Cancer Cells through Akt Pathway by Targeting PTEN. *Biomed Res Int* **2021**, 7590976 (2021). |
| *MIR103A2* | MicroRNA 103a-2 | Exosome miR-103a-2-5p promotes the proliferation and migration of esophageal squamous cell carcinoma (ESCC) cells. | D. C. Gao, B. Hou, D. Zhou, Q. X. Liu, K. Zhang, X. Lu, J. Zhang, H. Zheng, J. G. Dai, Tumor-derived exosomal miR-103a-2-5p facilitates esophageal squamous cell carcinoma cell proliferation and migration. *Eur Rev Med Pharmacol Sci* 24, 6097-6110 (2020). |
| *MIR142* | MicroRNA 142 | miR-142 regulates the tumorigenicity of breast cancer stem cells. miR-142 is identified as hematopoietic miRNAs capable of increasing the rate at which progenitor cells differentiate into T cells. | T. Isobe, S. Hisamori, D. J. Hogan, M. Zabala, D. G. Hendrickson, P. Dalerba, S. Cai, F. Scheeren, A. H. Kuo, S. S. Sikandar, J. S. Lam, D. Qian, F. M. Dirbas, G. Somlo, K. Lao, P. O. Brown, M. F. Clarke, Y. Shimono, miR-142 regulates the tumorigenicity of human breast cancer stem cells through the canonical WNT signaling pathway. *Elife* **3**, (2014).  Chen CZ, Li L, Lodish HF, Bartel DP. MicroRNAs modulate hematopoietic lineage differentiation. *Science*. 2004 Jan 2;303(5654):83-6. doi: 10.1126/science.1091903. Epub 2003 Dec 4. PMID: 14657504. |
| *MIR150* | MicroRNA 150 | The miR-150 expression is associated with survival and response to adjuvant chemotherapy status of patients with colorectal cancer. | Y. Ma, P. Zhang, F. Wang, H. Zhang, J. Yang, J. Peng, W. Liu, H. Qin, miR-150 as a potential biomarker associated with prognosis and therapeutic outcome in colorectal cancer. *Gut* **61**, 1447-1453 (2012). |
| *MIR185* | MicroRNA 185 | miR-185 suppresses TNBC cell proliferation. | H. Tang, P. Liu, L. Yang, X. Xie, F. Ye, M. Wu, X. Liu, B. Chen, L. Zhang, X. Xie, miR-185 suppresses tumor proliferation by directly targeting E2F6 and DNMT1 and indirectly upregulating BRCA1 in triple-negative breast cancer. *Mol Cancer Ther* **13**, 3185-3197 (2014). |
| *MIR221* | MicroRNA 221 | High expression of miR-221/miR-222 promotes the malignant proliferation, immune escape, invasion, and metastasis of tumor cells. | S. Dufresne, A. Rebillard, P. Muti, C. M. Friedenreich, D. R. Brenner, A Review of Physical Activity and Circulating miRNA Expression: Implications in Cancer Risk and Progression. *Cancer Epidemiol Biomarkers Prev* **27**, 11-24 (2018). |
| *MIR222* | MicroRNA 222 | High expression of miR-221/miR-222 promotes the malignant proliferation, immune escape, invasion, and metastasis of tumor cells. | S. Dufresne, A. Rebillard, P. Muti, C. M. Friedenreich, D. R. Brenner, A Review of Physical Activity and Circulating miRNA Expression: Implications in Cancer Risk and Progression. *Cancer Epidemiol Biomarkers Prev* **27**, 11-24 (2018). |
| *MIR223* | MicroRNA 223 | Higher expression levels of miR-223 are linked to colorectal cancer.  miR-223 is identified as hematopoietic miRNAs capable of increasing the rate at which progenitor cells differentiate into T cells with miR-223 identified as a potential diagnostic marker and therapeutic target for inflammatory disorders. | J. Zhang, X. Luo, H. Li, X. Yue, L. Deng, Y. Cui, Y. Lu, MicroRNA-223 functions as an oncogene in human colorectal cancer cells. Oncol Rep **32**, 115-120 (2014).  Chen CZ, Li L, Lodish HF, Bartel DP. MicroRNAs modulate hematopoietic lineage differentiation. *Science*. 2004 Jan 2;303(5654):83-6. doi: 10.1126/science.1091903. Epub 2003 Dec 4. PMID: 14657504.  Aziz F. The emerging role of miR-223 as novel potential diagnostic and therapeutic target for inflammatory disorders. *Cell Immunol*. 2016 May;303:1-6. doi: 10.1016/j.cellimm.2016.04.003. Epub 2016 Apr 9. PMID: 27129807. |
| *MIR650* | MicroRNA 650 | MiR-650 promotes proliferation, migration, invasion, and EMT of hepatocellular carcinoma. | L. L. Han, X. R. Yin, S. Q. Zhang, miR-650 Promotes the Metastasis and Epithelial-Mesenchymal Transition of Hepatocellular Carcinoma by Directly Inhibiting LATS2 Expression. *Cell Physiol Biochem* **51**, 1179-1192 (2018). |
| *MIRLET7A1* | MicroRNA Let-7a-1 | MicroRNA let-7a suppresses breast cancer cell migration. | S. J. Kim, J. Y. Shin, K. D. Lee, Y. K. Bae, K. W. Sung, S. J. Nam, K. H. Chun, MicroRNA let-7a suppresses breast cancer cell migration and invasion through downregulation of C-C chemokine receptor type 7. *Breast Cancer Res* 14, R14 (2012). |
| *MIRLET7A2* | MicroRNA let-7a-2 | MIRLET-7A2 is a member of the MIRLET-7 family. MIRLET-7 acts as an oncogene in some cancer types such as hepatic cancer. | Z. Tang, G. S. Ow, J. P. Thiery, A. V. Ivshina, V. A. Kuznetsov, Meta-analysis of transcriptome reveals let-7b as an unfavorable prognostic biomarker and predicts molecular and clinical subclasses in high-grade serous ovarian carcinoma. *Int J Cancer* **134**, 306-318 (2014). |
| *MIRLET7C* | MicroRNA Let-7c | miR-let-7c suppresses the tumourigenesis of human mucosal melanoma and enhances the sensitivity to chemotherapy. | H. Tang, M. Ma, J. Dai, C. Cui, L. Si, X. Sheng, Z. Chi, L. Xu, S. Yu, T. Xu, J. Yan, H. Yu, L. Yang, Y. Kong, J. Guo, miR-let-7b and miR-let-7c suppress tumourigenesis of human mucosal melanoma and enhance the sensitivity to chemotherapy. *J Exp Clin Cancer Res* **38**, 212 (2019). |
| *MIRLET7F1* | MicroRNA Let-7f-1 | miRNA-let-7f-1 inhibits cell proliferation and triggers apoptosis in prostate cancer. | D. Li, L. Chen, W. Zhao, J. Hao, R. An, MicroRNA-let-7f-1 is induced by lycopene and inhibits cell proliferation and triggers apoptosis in prostate cancer. *Mol Med Rep* **13**, 2708-2714 (2016). |
| *MIRLET7F2* | MicroRNA Let-7f-2 | Not Reported. |  |
| *MIRLET7G* | MicroRNA Let-7g | miR-let-7g acts as a tumor suppressor and a predictive biomarker for chemoresistance in human epithelial ovarian cancer. | F. Biamonte, G. Santamaria, A. Sacco, F. M. Perrone, A. Di Cello, A. M. Battaglia, A. Salatino, A. Di Vito, I. Aversa, R. Venturella, F. Zullo, F. Costanzo, MicroRNA let-7g acts as tumor suppressor and predictive biomarker for chemoresistance in human epithelial ovarian cancer. *Sci Rep* **9**, 5668 (2019). |
| **Mapped to mature miRNA reference** | | | |
| *let−7i−5p* | MicroRNA Let-7i-5p | Let-7i-5p enhances cell proliferation, migration, and invasion of clear cell renal cell carcinoma. | Y. Liu, X. Hu, L. Hu, C. Xu, X. Liang, Let-7i-5p enhances cell proliferation, migration and invasion of ccRCC by targeting HABP4. *BMC Urol* **21**, 49 (2021). |
| *miR−15a−5p* | MicroRNA 15a-5p | miR-15a-5p inhibits proliferation and migration of colon cancer cells. | Z. Li, Z. Zhu, Y. Wang, Y. Wang, W. Li, Z. Wang, X. Zhou, Y. Bao, hsa‑miR‑15a‑5p inhibits colon cell carcinoma via targeting CCND1. *Mol Med Rep* **24**, (2021). |
| *miR−19a−3p* | MicroRNA 19a-3p | miR-19a-3p promotes tumor metastasis and chemoresistance through the PTEN/Akt pathway in hepatocellular carcinoma. | X. M. Jiang, X. N. Yu, T. T. Liu, H. R. Zhu, X. Shi, E. Bilegsaikhan, H. Y. Guo, G. Q. Song, S. Q. Weng, X. X. Huang, L. Dong, H. L. A. Janssen, X. Z. Shen, J. M. Zhu, microRNA-19a-3p promotes tumor metastasis and chemoresistance through the PTEN/Akt pathway in hepatocellular carcinoma. *Biomed Pharmacother* **105**, 1147-1154 (2018). |
| *miR−19b−3p* | MicroRNA 19b-3p | miR-19b-3p promotes proliferation of human colon cancer. | T. Jiang, L. Ye, Z. Han, Y. Liu, Y. Yang, Z. Peng, J. Fan, miR-19b-3p promotes colon cancer proliferation and oxaliplatin-based chemoresistance by targeting SMAD4: validation by bioinformatics and experimental analyses. *J Exp Clin Cancer Res* **36**, 131 (2017). |
| *miR−20a−5p* | MicroRNA 20a-5p | miR-20a-5p inhibits endometrial cancer progression. | Y. He, H. Ma, J. Wang, Y. Kang, Q. Xue, miR-20a-5p inhibits endometrial cancer progression by targeting janus kinase 1. *Oncol Lett* **21**, 427 (2021). |
| *miR−24−3p* | MicroRNA 24-3p | miR-24-3p promotes colon carcinogenesis. | Z. Gao, L. Zhou, S. Hua, H. Wu, L. Luo, L. Li, S. Wang, Y. Liu, Z. Zhou, X. Chen, miR-24-3p promotes colon cancer progression by targeting ING1. Signal Transduct Target Ther **5**, 171 (2020). |
| *miR−25−3p* | MicroRNA 25-3p | miR-25-3p promotes the proliferation of triple negative breast cancer. | H. Chen, H. Pan, Y. Qian, W. Zhou, X. Liu, MiR-25-3p promotes the proliferation of triple negative breast cancer by targeting BTG2. *Mol Cancer* **17**, 4 (2018). |
| *miR−26a−5p* | MicroRNA 26a-5p | miR-26a-5p inhibits the proliferation and invasion of gastric cancer cell. | Y. Li, P. Wang, L. L. Wu, J. Yan, X. Y. Pang, S. J. Liu, miR-26a-5p Inhibit Gastric Cancer Cell Proliferation and Invasion Through Mediated Wnt5a. *Onco Targets Ther* **13**, 2537-2550 (2020). |
| *miR−27a−3p* | MicroRNA 27a-3p | miR-27a-3p promotes gastric cancer cell proliferation *in vitro* and tumor growth *in vivo*. | L. Zhou, X. Liang, L. Zhang, L. Yang, N. Nagao, H. Wu, C. Liu, S. Lin, G. Cai, J. Liu, MiR-27a-3p functions as an oncogene in gastric cancer by targeting BTG2. *Oncotarget* **7**, 51943-51954 (2016). |
| *miR−29a−3p* | MicroRNA 29a-3p | miR-29a-3p inhibits the migration and proliferation of endometrial cancer cell. | A. Geng, L. Luo, F. Ren, L. Zhang, H. Zhou, X. Gao, miR-29a-3p inhibits endometrial cancer cell proliferation, migration and invasion by targeting VEGFA/CD C42/PAK1. *BMC Cancer* **21**, 843 (2021). |
| *miR−29b−3p* | MicroRNA 29b-3p | miR-29b-3p inhibits angiogenesis and migration and invasion of pancreatic cancer cell. | L. Wang, N. Mu, N. Qu, Methylation of the miR‑29b‑3p promoter contributes to angiogenesis, invasion, and migration in pancreatic cancer. *Oncol Rep* **45**, 65-72 (2021). |
| *miR−29c−3p* | MicroRNA 29c-3p | miR-29c-3p regulates the proliferation and migration of ovarian cancer. | S. Feng, S. Luo, C. Ji, J. Shi, miR-29c-3p regulates proliferation and migration in ovarian cancer by targeting KIF4A. *World J Surg Oncol* **18**, 315 (2020). |
| *miR−30b−5p* | MicroRNA 30b-5p | miR-30b-5p acts as a tumor suppressor. It inhibits the proliferation and metastasis of renal cell carcinoma. | W. Liu, H. Li, Y. Wang, X. Zhao, Y. Guo, J. Jin, R. Chi, MiR-30b-5p functions as a tumor suppressor in cell proliferation, metastasis and epithelial-to-mesenchymal transition by targeting G-protein subunit alpha-13 in renal cell carcinoma. *Gene* **626**, 275-281 (2017). |
| *miR−98−5p* | MicroRNA 98-5p | miRNA-98-5p inhibits the proliferation and metastasis of NSCLC cells. | F. Jiang, Q. Yu, Y. Chu, X. Zhu, W. Lu, Q. Liu, Q. Wang, MicroRNA-98-5p inhibits proliferation and metastasis in non-small cell lung cancer by targeting TGFBR1. *Int J Oncol* **54**, 128-138 (2019). |
| *miR−126−3p* | MicroRNA 126-3p | MicroRNA‑126‑3p inhibits the proliferation, migration, invasion, and angiogenesis of triple‑negative breast cancer cells. | Z. Hong, C. Hong, B. Ma, Q. Wang, X. Zhang, L. Li, C. Wang, D. Chen, MicroRNA‑126‑3p inhibits the proliferation, migration, invasion, and angiogenesis of triple‑negative breast cancer cells by targeting RGS3. *Oncol Rep* **42**, 1569-1579 (2019). |
| *miR−125a−5p* | MicroRNA 125a-5p | miR-125a-5p inhibits the proliferation and migration of breast cancer cells. | Z. Liang, Q. Pan, Z. Zhang, C. Huang, Z. Yan, Y. Zhang, J. Li, MicroRNA‑125a‑5p controls the proliferation, apoptosis, migration and PTEN/MEK1/2/ERK1/2 signaling pathway in MCF‑7 breast cancer cells. *Mol Med Rep* **20**, 4507-4514 (2019).  Wang S, Huang J, Lyu H, Lee CK, Tan J, Wang J, Liu B. Functional cooperation of miR-125a, miR-125b, and miR-205 in entinostat-induced downregulation of erbB2/erbB3 and apoptosis in breast cancer cells. *Cell Death Dis.* 2013 Mar 21;4(3):e556. doi: 10.1038/cddis.2013.79. PMID: 23519125; PMCID: PMC3615747. |
| *miR−126−5p* | MicroRNA 126-5p | miR-126-5p inhibits the migration of breast cancer cells. | Y. Miao, J. Lu, B. Fan, L. Sun, MicroRNA-126-5p Inhibits the Migration of Breast Cancer Cells by Directly Targeting CNOT7. *Technol Cancer Res Treat* **19**, 1533033820977545 (2020). |
| *miR−146a−5p* | MicroRNA 146a-5p | miR‑146a‑5p inhibits TNBC cell proliferation, migration, and invasion. | C. Si, Q. Yu, Y. Yao, Effect of miR-146a-5p on proliferation and metastasis of triple-negative breast cancer via regulation of SOX5. *Exp Ther Med* **15**, 4515-4521 (2018). |
| *miR−146b−5p* | MicroRNA 146b-5p | miR-146b-5p inhibits tumorigenesis and metastasis of gallbladder cancer. | B. Ouyang, N. Pan, H. Zhang, C. Xing, W. Ji, miR‑146b‑5p inhibits tumorigenesis and metastasis of gallbladder cancer by targeting Toll‑like receptor 4 via the nuclear factor‑kappaB pathway. *Oncol Rep* **45**, (2021). |
| *miR−148a−3p* | MicroRNA 148a-3p | miR‑148a‑3p inhibits the proliferation of cervical cancer cells. | Q. Chen, Y. Wang, H. Dang, X. Wu, MicroRNA-148a-3p inhibits the proliferation of cervical cancer cells by regulating the expression levels of DNMT1 and UTF1. *Oncol Lett* **22**, 617 (2021). |
| *miR−151a−3p* | MicroRNA 151a-3p | miR-151a induces partial EMT by regulating E-cadherin in NSCLC cells. | I. Daugaard, K. J. Sanders, A. Idica, K. Vittayarukskul, M. Hamdorf, J. D. Krog, R. Chow, D. Jury, L. L. Hansen, H. Hager, P. Lamy, C. L. Choi, D. Agalliu, D. G. Zisoulis, I. M. Pedersen, miR-151a induces partial EMT by regulating E-cadherin in NSCLC cells. *Oncogenesis* **6**, e366 (2017). |
| *miR−151a−5p* | MicroRNA 151a-5p | miR-151a-5p is highly expressed in lung cancer, and its inhibition reduces the proliferation, invasion and migration of lung cancer cells. | S. Guo, J. Zhang, Y. Y. Zhao, L. Y. Zhou, Y. Xie, X. Y. Wu, X. Bian, X. Y. Yu, The expressions of miR-151a-5p and miR-23b in lung cancer tissues and their effects on the biological functions of lung cancer A549 cells. *Eur Rev Med Pharmacol Sci* **24**, 6779-6785 (2020). |
| *miR−338−3p* | MicroRNA 338-3p | miR-338-3p suppresses ovarian cancer cells growth and metastasis. | R. Zhang, H. Shi, F. Ren, W. Feng, Y. Cao, G. Li, Z. Liu, P. Ji, M. Zhang, MicroRNA-338-3p suppresses ovarian cancer cells growth and metastasis: implication of Wnt/catenin beta and MEK/ERK signaling pathways. *J Exp Clin Cancer Res* **38**, 494 (2019). |
| *miR−342−3p* | MicroRNA 342-3p | miR-342-3p inhibits the growth of NSCLC. | Z. Chen, J. Ying, W. Shang, D. Ding, M. Guo, H. Wang, miR-342-3p Regulates the Proliferation and Apoptosis of NSCLC Cells by Targeting BCL-2. *Technol Cancer Res Treat* **20**, 15330338211041193 (2021). |
| *miR−454−3p* | MicroRNA 454-3p | miR-454-3p inhibits NSCLC cell proliferation and metastasis. | H. Liao, Y. Liang, L. Kang, Y. Xiao, T. Yu, R. Wan, miR‑454‑3p inhibits non‑small cell lung cancer cell proliferation and metastasis by targeting TGFB2. *Oncol Rep* **45**, (2021). |
| **Plasma Samples** | | | |
| ***Protein-coding genes*** | | | |
| *ADAMTS6* | ADAM Metallopeptidase With Thrombospondin Type 1 Motif 6 | ADAMTS6 inhibits tumor development by regulating the ERK pathway via binding of miR-221-3p in breast cancer. | Y. Xie, Q. Gou, K. Xie, Z. Wang, Y. Wang, H. Zheng, ADAMTS6 suppresses tumor progression via the ERK signaling pathway and serves as a prognostic marker in human breast cancer. *Oncotarget* **7**, 61273-61283 (2016). |
| *ADAMTSL1* | ADAMTS-Like Protein 1 | Germline variation in ADAMTSL1 is associated with prognosis following breast cancer treatment in young women. | L. Kadalayil, S. Khan, H. Nevanlinna, P. A. Fasching, F. J. Couch, J. L. Hopper, J. Liu, T. Maishman, L. Durcan, S. Gerty, C. Blomqvist, B. Rack, W. Janni, A. Collins, D. Eccles, W. Tapper, Germline variation in ADAMTSL1 is associated with prognosis following breast cancer treatment in young women. *Nat Commun* **8**, 1632 (2017). |
| *ADGRV1* | Adhesion G Protein-Coupled Receptor V1 | Not Reported. |  |
| *ANKS1B* | Ankyrin Repeat and Sterile Alpha Motif Domain-Containing Protein 1B | *ANKS1B* gene expression is elevated in patients with pre-B cell acute lymphocytic leukemia associated with t(1;19) translocation. | G. Casagrande, G. te Kronnie, G. Basso, The effects of siRNA-mediated inhibition of E2A-PBX1 on EB-1 and Wnt16b expression in the 697 pre-B leukemia cell line. *Haematologica* **91**, 765-771 (2006). |
| *BOLL* | Boule-like RNA- Binding Protein | Expression of BOLL in colorectal cancer cell significantly enhances cell proliferation, colony formation, and migration. | K. J. Kang, J. H. Pyo, K. J. Ryu, S. J. Kim, J. M. Ha, K. Choi, S. N. Hong, B. H. Min, D. K. Chang, H. J. Son, P. L. Rhee, J. J. Kim, Y. H. Kim, Oncogenic Role of BOLL in Colorectal Cancer. *Dig Dis Sci* **60**, 1663-1673 (2015). |
| *CCDC150* | Coiled-Coil Domain Containing 150 | Not Reported. |  |
| *CHCHD6* | Coiled-Coil-Helix-Coiled-Coil-Helix Domain Containing 6 | Alterations in CHCHD6 expression affect chemosensitivity of human cancer cells to genotoxic anticancer drugs. | J. An, J. Shi, Q. He, K. Lui, Y. Liu, Y. Huang, M. S. Sheikh, CHCM1/CHCHD6, novel mitochondrial protein linked to regulation of mitofilin and mitochondrial cristae morphology. *J Biol Chem* **287**, 7411-7426 (2012). |
| *CLSTN2* | Calsyntenin 2 | CLSTN2 expression is associated with the prognosis of colon adenocarcinoma patients. | M. Wu, W. Lou, M. Lou, P. Fu, X. F. Yu, Integrated Analysis of Distant Metastasis-Associated Genes and Potential Drugs in Colon Adenocarcinoma. *Front Oncol* **10**, 576615 (2020). |
| *CMSS1* | Cms1 Ribosomal Small Subunit Homolog | CMSS1 is closely related to the overall survival of patients with diffuse large B-cell lymphoma. | Y. Xie, X. Luo, H. He, T. Pan, Y. He, Identification of an individualized RNA binding protein-based prognostic signature for diffuse large B-cell lymphoma. *Cancer Med* **10**, 2703-2713 (2021). |
| *C6orf118* | Chromosome 6 Open Reading Frame 118 | Most thyroid cancers, pancreatic, urothelial, and renal cancers show moderate cytoplasmic positivity of C6orf118. | https://www.proteinatlas.org/ENSG00000112539-C6orf118/pathology |
| *DGKB* | Diacylglycerol Kinase Beta | DGKβ is a member of diacylglycerol kinase family. The function of DGKβ in cancer is not known. But DGKα is highly expressed in several refractory cancer cells including melanoma, hepatocellular carcinoma, and glioblastoma cells and it is a therapeutic target. | F. Sakane, F. Hoshino, M. Ebina, H. Sakai, D. Takahashi, The Roles of Diacylglycerol Kinase alpha in Cancer Cell Proliferation and Apoptosis. *Cancers (Basel)* **13**, (2021). |
| *DIRAS3* | DIRAS Family GTPase 3 | DIRAS3 is a maternally imprinted gene that is discovered in the context of ovarian cancer.  It is a tumor suppressor gene whose normally silenced maternal allele can be re-activated to induce autophagy in breast cancer cells. | Y. Yu, F. Xu, H. Peng, X. Fang, S. Zhao, Y. Li, B. Cuevas, W. L. Kuo, J. W. Gray, M. Siciliano, G. B. Mills, R. C. Bast, Jr., NOEY2 (ARHI), an imprinted putative tumor suppressor gene in ovarian and breast carcinomas. *Proc Natl Acad Sci U S A* **96**, 214-219 (1999).  Zou, CF., Jia, L., Jin, H. et al. Re-expression of ARHI (DIRAS3) induces autophagy in breast cancer cells and enhances the inhibitory effect of paclitaxel. *BMC Cancer* 11, 22 (2011). https://doi.org/10.1186/1471-2407-11-22 |
| *FAM135B* | Family with Sequence Similarity 135 Member B | FAM135B gene is associated with a carcinogenic risk of esophageal squamous cell carcinoma. | T. Wang, X. Lv, S. Jiang, S. Han, Y. Wang, Expression of ADAM29 and FAM135B in the pathological evolution from normal esophageal epithelium to esophageal cancer: Their differences and clinical significance. Oncol Lett **19**, 1727-1734 (2020). |
| *KCNH5* | Potassium Voltage-Gated Channel Subfamily H Member 5 | Lung, ovarian and breast cancers exhibit weak to moderate immunohistochemistry staining of KCNH5. | https://www.proteinatlas.org/ENSG00000140015-KCNH5/pathology |
| *LUZP2* | Leucine Zipper Protein 2 | Downregulation of LUZP2 is correlated with poor prognosis of low-grade Glioma. | Y. Li, G. Deng, Y. Qi, H. Zhang, H. Jiang, R. Geng, Z. Ye, B. Liu, Q. Chen, Downregulation of LUZP2 Is Correlated with Poor Prognosis of Low-Grade Glioma. *Biomed Res Int* **2020**, 9716720 (2020). |
| *METTL15* | Methyltransferase Like 15 | Circ-METTL15 contributes to the proliferation, metastasis, immune escape, and restrains apoptosis in lung cancer. | R. Zhang, L. Shang, J. Nan, K. Niu, J. Dai, X. Jin, X. Zhang, Circ-METTL15 contributes to the proliferation, metastasis, immune escape and restrains apoptosis in lung cancer by regulating miR-1299/PDL1 axis. *Autoimmunity* **55**, 8-20 (2022). |
| *MIOS* | Meiosis Regulator for Oocyte Development | Not Reported. |  |
| *NADK2* | NAD Kinase 2 | Knockdown of NADK2 affects pancreatic ductal adenocarcinomas growth to a lesser degree. | T. Schild, M. R. McReynolds, C. Shea, V. Low, B. E. Schaffer, J. M. Asara, E. Piskounova, N. Dephoure, J. D. Rabinowitz, A. P. Gomes, J. Blenis, NADK is activated by oncogenic signaling to sustain pancreatic ductal adenocarcinoma. *Cell Rep* **35**, 109238 (2021). |
| *NLGN1* | Neuroligin 1 | Upregulated NLGN1 predicts poor survival in colorectal cancer. | Q. Yu, X. Wang, Y. Yang, P. Chi, J. Huang, S. Qiu, X. Zheng, X. Chen, Upregulated NLGN1 predicts poor survival in colorectal cancer. BMC Cancer **21**, 884 (2021). |
| *NUP210L* | Nucleoporin 210 Like | *Nup210*, a gene encoding a nuclear pore complex (NPC) protein, is a potential metastasis susceptibility gene. NPC proteins are associated with several developmental disorders and cancers. | S. Sakuma, M. A. D'Angelo, The roles of the nuclear pore complex in cellular dysfunction, aging and disease. *Semin Cell Dev Biol* **68**, 72-84 (2017). |
| *PHF24* | PHD Finger Protein 24 | N/A |  |
| *PLVAP* | Plasmalemma Vesicle Associated Protein | PLVAP is associated with glioma-associated malignant processes. | K. Ma, X. Chen, X. Zhao, S. Chen, J. Yang, PLVAP is associated with glioma-associated malignant processes and immunosuppressive cell infiltration as a promising marker for prognosis. *Heliyon* **8**, e10298 (2022). |
| *SENP2* | Sentrin-Specific Protease 2 | SENP2 decreases bladder cell migration and invasion. | M. Tan, H. Gong, J. Wang, L. Tao, D. Xu, E. Bao, Z. Liu, J. Qiu, SENP2 regulates MMP13 expression in a bladder cancer cell line through SUMOylation of TBL1/TBLR1. *Sci Rep* **5**, 13996 (2015) |
| *SPOCK3* | SPARC/Osteonectin, Cwcv And Kazal Like Domains Proteoglycan 3 | SPOCK3 expresses in lung squamous cell carcinoma. | https://www.intogen.org/search?gene=SPOCK3&cancer=LUSC |
| *TRBJ1-6* | T Cell Receptor Beta Joining 1-6 | J region of the variable domain of T cell receptor (TR) beta chain that participates in the antigen recognition. | M. P. Lefranc, Immunoglobulin and T Cell Receptor Genes: IMGT((R)) and the Birth and Rise of Immunoinformatics. *Front Immunol* **5**, 22 (2014). |
| *UVRAG* | UV Radiation Resistance Associated Gene | UVRAG is frequently mutated in various cancer types, and mutations of UVRAG increase sensitivity to chemotherapy by impairing DNA-damage repair. | Y. K. Jo, N. Y. Park, J. H. Shin, D. S. Jo, J. E. Bae, E. S. Choi, S. Maeng, H. B. Jeon, S. A. Roh, J. W. Chang, J. C. Kim, D. H. Cho, Up-regulation of UVRAG by HDAC1 Inhibition Attenuates 5FU-induced Cell Death in HCT116 Colorectal Cancer Cells. *Anticancer Res* **38**, 271-277 (2018). |
| *WWP2* | WW Domain Containing E3 Ubiquitin Protein Ligase 2 | WWP2 plays a crucial part in the pathogenesis in different types of tumors. | R. Zhang, J. Zhang, W. Luo, Z. Luo, S. Shi, WWP2 Is One Promising Novel Oncogene. *Pathol Oncol Res* **25**, 443-446 (2019). |
| *XXYLT1* | Xyloside Xylosyltransferase 1 | *XXYLT1* methylation contributes to the occurrence of lung adenocarcinoma. | H. Zeng, Y. Wang, Y. Wang, Y. Zhang, XXYLT1 methylation contributes to the occurrence of lung adenocarcinoma: Methylation and lung adenocarcinoma. *Medicine (Baltimore)* **100**, e24150 (2021). |
| **lncRNA** | | | |
| *AC004014.1* | N/A* | Not Reported. |  |
| *AC096759.1* | N/A* | Not Reported. |  |
| *AC117373.1* | N/A* | Not Reported. |  |
| *AC131254.1* | N/A* | Not Reported. |  |
| *LINC01191* | Long Intergenic Non-Protein Coding RNA 1191 | A set of lncRNAs (LINC01191, RP4-639F20.1 and CTC-429P9.3) is associated with distant metastasis-free survival. | R. Liu, R. Hu, W. Zhang, H. H. Zhou, Long noncoding RNA signature in predicting metastasis following tamoxifen treatment for ER-positive breast cancer. *Pharmacogenomics*, (2018). |
| *LINC01495* | Long Intergenic Non-Protein Coding RNA 1495 | Not Reported. |  |
| **Y RNA** | | | |
| *RNY5* | RNA, Ro60-Associated Y5 | Higher RNY5 expression is associated with poor prognosis of prostate cancer patients.  The ectopic overexpression of RNY5 fragments induce cell death in human primary cells but not cancer cells, possibly a mechanism by which cancer cells might establish a favorable environment for proliferation. | Y. Tolkach, E. M. Niehoff, A. F. Stahl, C. Zhao, G. Kristiansen, S. C. Muller, J. Ellinger, YRNA expression in prostate cancer patients: diagnostic and prognostic implications. *World J Urol* **36**, 1073-1078 (2018).  Chakrabortty SK, Prakash A, Nechooshtan G, Hearn S, Gingeras TR. Extracellular vesicle-mediated transfer of processed and functional RNY5 RNA. *RNA*. 2015 Nov;21(11):1966-79. doi: 10.1261/rna.053629.115. Epub 2015 Sep 21. PMID: 26392588; PMCID: PMC4604435. |
| **Mapped with mature miRNA reference** | | | |
| *Let-7f2-5p* |  | Not Reported. |  |
| *miR-1246* | MicroRNA 1246 | Tumour-initiating cell-specific miR-1246 expression promotes NSCLC progression. | W. C. Zhang, T. M. Chin, H. Yang, M. E. Nga, D. P. Lunny, E. K. Lim, L. L. Sun, Y. H. Pang, Y. N. Leow, S. R. Malusay, P. X. Lim, J. Z. Lee, B. J. Tan, N. Shyh-Chang, E. H. Lim, W. T. Lim, D. S. Tan, E. H. Tan, B. C. Tai, R. A. Soo, W. L. Tam, B. Lim, Tumour-initiating cell-specific miR-1246 and miR-1290 expression converge to promote non-small cell lung cancer progression. *Nat Commun* **7**, 11702 (2016). |
| **miRNA** | | | |
| *MIR619* | MicroRNA 619 | miR-619-5p overexpression inhibits proliferative, migratory and invasive abilities of cisplatin-resistant oral squamous cell carcinoma cells. | A. Song, Y. Wu, W. Chu, X. Yang, Z. Zhu, E. Yan, W. Zhang, J. Zhou, X. Ding, J. Liu, H. Zhu, J. Ye, Y. Wu, Y. Zheng, X. Song, Involvement of miR-619-5p in resistance to cisplatin by regulating ATXN3 in oral squamous cell carcinoma. *Int J Biol Sci* **17**, 430-447 (2021). |
| *MIR1273* | MicroRNA 1273A | Downregulated expression of miR-1273a predicts poor prognosis for colon cancer and enhances tumor cell proliferation, migration, and invasion. | L. Sun, X. Zhou, Q. Jiang, Y. Zhuang, D. Li, Low miR-1273a expression predicts poor prognosis of colon cancer and facilitates tumor cell proliferation, migration, and invasion. *Braz J Med Biol Res* **54**, e10394 (2021). |
| *MIR1285* | MicroRNA 1285 | miR-1285 expression is significantly reduced in renal cell carcinoma (RCC) clinical specimens compared with adjacent non-cancerous tissues. | H. Hidaka, N. Seki, H. Yoshino, T. Yamasaki, Y. Yamada, N. Nohata, M. Fuse, M. Nakagawa, H. Enokida, Tumor suppressive microRNA-1285 regulates novel molecular targets: aberrant expression and functional significance in renal cell carcinoma. *Oncotarget* **3**, 44-57 (2012). |
| *MIR3929* | MicroRNA 3929 | miR-3929 inhibits proliferation and promotes apoptosis of cervical cancer cells. | Y. Wang, X. Li, S. Wang, Z. Song, Y. Bao, L. Zheng, G. Wang, Y. Sun, miR-3929 Inhibits Proliferation and Promotes Apoptosis by Downregulating Cripto-1 Expression in Cervical Cancer Cells. *Cytogenet Genome Res* **161**, 425-436 (2021). |
| *MIR5096* | MicroRNA 5096 | miR-5096 inhibits colony formation, transwell migration, and invasion of breast cancer cells. | P. Yadav, P. Sharma, S. Sundaram, G. Venkatraman, A. K. Bera, D. Karunagaran, SLC7A11/ xCT is a target of miR-5096 and its restoration partially rescues miR-5096-mediated ferroptosis and anti-tumor effects in human breast cancer cells. *Cancer Lett* **522**, 211-224 (2021). |
| **Satellite** | | | |
| *(CATTC)n* | N/A* | Not Reported. |  |
| *ALR/Alpha* | N/A* | Not Reported. |  |
| *SAR* | N/A* | Not Reported. |  |
| **DNA element** | | | |
| *Charlie3* | N/A* | Not Reported. |  |
| *MER106A* | N/A* | Not Reported. |  |
| **SINE** | | | |
| *AluJb* | N/A* | AluJb drives LIN28B expression and contributes to oncogenesis in lung cancer cell lines. | H. S. Jang, N. M. Shah, A. Y. Du, Z. Z. Dailey, E. C. Pehrsson, P. M. Godoy, D. Zhang, D. Li, X. Xing, S. Kim, D. O'Donnell, J. I. Gordon, T. Wang, Transposable elements drive widespread expression of oncogenes in human cancers. *Nat Genet* **51**, 611-617 (2019). |
| *AluJr* | N/A* | Not Reported. |  |
| *AluSx1* | N/A* | Not Reported. |  |
| *AluSx4* | N/A* | Not Reported. |  |
| *MIR* | N/A* | Not Reported. |  |
| *MIR3* | Retrotransposon Enhancer-Blocking Element | HBV-encoded miR-3 contributes to the development of HBV-related hepatocellular carcinoma and may be a therapeutic target for this cancer. | J. Tang, X. Xiao, Y. Jiang, Y. Tian, Z. Peng, M. Yang, Z. Xu, G. Gong, miR-3 Encoded by Hepatitis B Virus Downregulates PTEN Protein Expression and Promotes Cell Proliferation. *J Hepatocell Carcinoma* **7**, 257-269 (2020). |
| *MIRb* | N/A* | Not Reported. |  |
| *MIRc* | N/A* | Not Reported. |  |
| *AC092421.1* |  | Not Reported. |  |

N/A*: Not Available;
