## Supplemental Table 3 and 4 for "TGIRT-seq of Inflammatory Breast Cancer Tumor and Blood Samples Reveals Widespread Enhanced Transcription Impacting RNA Splicing and Intronic RNAs in Plasma"

**Table S3. Genes in Hallmark gene sets that have high ΔIDRs (>2) that were not significantly differentially over-expressed (LFC<1) in IBC versus non-IBC FFPE tumor samples for reads mapped to the transcriptome reference sequences.**

| Inflammatory Response | | | TNFα-via-NFκB | | | Epithelial Mesenchymal Transition | | | |
| --- | --- | --- | --- | --- | --- | --- | --- | --- | --- |
| ID | IDR | LFC | ID | IDR | LFC | | ID | IDR | LFC |
| ADGRE1* | 0.51 | -1.49 | BHLHE40* | 0.67 | 0.12 | | CDH2 | 0.17 | -1.71 |
| ADM | 0.40 | -0.08 | BTG1* | 0.81 | 0.51 | | COL12A1 | 0.13 | 0.35 |
| C3AR1 | 0.48 | -0.67 | BTG3 | 0.33 | 0.65 | | COL4A1 | 0.13 | -0.46 |
| CCL20^1^ | 0.03 | -0.08 | CCL20 | 0.03 | -0.08 | | COL5A2 | 0.09 | 0.98 |
| CD14 | 0.48 | 0.06 | CCND1 | 0.11 | 0.47 | | CXCL8 | 0.26 | -0.76 |
| CD69 | 0.40 | 0.97 | CCNL1* | 0.99 | -0.39 | | DPYSL3 | 0.14 | 0.57 |
| CHST2* | 3.13 | -0.06 | CD69 | 0.40 | 0.97 | | ECM1 | 0.13 | -0.54 |
| CXCL10^2^ | 0.41 | -0.84 | CXCL10 | 0.41 | -0.84 | | ELN | 0.10 | 0.06 |
| CXCL11 | 0.15 | -1.20 | CXCL11 | 0.15 | -1.20 | | FLNA | 0.15 | -0.16 |
| CXCL8^3^ | 0.26 | -0.76 | EIF1 | 0.33 | 0.08 | | FMOD | 0.03 | 0.19 |
| GCH1 | 0.38 | -1.48 | GADD45B | 0.30 | 0.57 | | FN1 | 0.07 | 0.69 |
| GPR183* | 0.58 | 0.76 | GCH1 | 0.38 | -1.48 | | FSTL1 | 0.10 | 0.70 |
| IL18R1*^4^ | 0.77 | -0.95 | GFPT2 | 0.27 | -0.06 | | GADD45B | 0.30 | 0.57 |
| IL1B^5^ | 0.27 | -0.91 | GPR183* | 0.58 | 0.76 | | GJA1 | 0.07 | 0.45 |
| ITGA5 | 0.24 | 0.51 | IL1B | 0.27 | -0.91 | | IGFBP3 | 0.09 | 0.53 |
| NAMPT* | 0.55 | 0.14 | KDM6B | 0.24 | 0.97 | | ITGA5 | 0.24 | 0.51 |
| NFKBIA | 0.31 | 0.96 | KLF2* | 0.55 | 0.93 | | ITGAV | 0.14 | -0.12 |
| NPFFR2 | 0.03 | -1.08 | KLF4 | 0.42 | 0.64 | | LAMA2 | 0.17 | 0.78 |
| OLR1 | 0.10 | 0.20 | KYNU | 0.05 | 0.23 | | LOX | 0.12 | 0.60 |
| PDPN | 0.19 | 0.63 | MARCKS | 0.29 | -0.02 | | MFAP5 | 0.14 | 0.20 |
| PTAFR | 0.14 | -0.73 | MCL1 | 0.08 | 0.47 | | MMP1 | 0.07 | -1.94 |
| PTGER2 | 0.31 | -0.12 | MSC | 0.15 | -0.88 | | MMP3 | 0.30 | 0.69 |
| ROS1* | 2.61 | -8.03 | NAMPT* | 0.55 | 0.14 | | TAGLN | 0.42 | -0.15 |
| SELENOS | 0.23 | -0.62 | NFKBIA | 0.31 | 0.96 | | THBS1 | 0.32 | 0.35 |
| SPHK1 | 0.06 | 0.48 | NR4A3 | 0.41 | -0.17 | | THY1 | 0.08 | 0.51 |
| TNFAIP6 | 0.11 | -0.32 | OLR1 | 0.10 | 0.20 | | TNC | 0.18 | 0.81 |
|  |  |  | PHLDA2* | 0.84 | -0.65 | | TNFRSF11B | 0.22 | -0.57 |
|  |  |  | PPP1R15A* | 0.59 | 0.92 | | TPM4 | 0.09 | 0.50 |
|  |  |  | PTGS2* | 0.58 | -0.05 | |  |  |  |
|  |  |  | REL | 0.08 | 0.26 | |  |  |  |
|  |  |  | SLC2A3* | 0.63 | -0.42 | |  |  |  |
|  |  |  | SNN | 0.14 | -1.00 | |  |  |  |
|  |  |  | SOCS3* | 0.55 | 0.54 | |  |  |  |
|  |  |  | SPHK1 | 0.06 | 0.48 | |  |  |  |
|  |  |  | TNC | 0.18 | 0.81 | |  |  |  |
|  |  |  | TNFAIP6 | 0.11 | -0.32 | |  |  |  |

**Table S3 (Continued).**

| Coagulation | | | Hypoxia | | |
| --- | --- | --- | --- | --- | --- |
| ID | IDR | LFC | ID | IDR | LFC |
| ANXA1 | 0.34 | 0.76 | ADM | 0.40 | -0.08 |
| C1R | 0.05 | 0.53 | AK4 | 0.10 | 0.43 |
| DUSP6 | 0.47 | 0.74 | BHLHE40* | 0.67 | 0.12 |
| FN1 | 0.07 | 0.69 | BTG1* | 0.81 | 0.51 |
| GDA* | 0.55 | -0.01 | CASP6 | 0.22 | -0.88 |
| MMP1 | 0.07 | -1.94 | CAV1 | 0.09 | -0.22 |
| MMP3 | 0.30 | 0.69 | CHST2* | 3.13 | -0.06 |
| MST1* | 0.88 | 0.34 | EDN2 | 0.45 | -1.53 |
| OLR1 | 0.10 | 0.20 | EFNA3 | 0.38 | -1.33 |
| PRSS23 | 0.06 | -0.34 | GAPDH | 0.04 | -0.39 |
| S100A1 | 0.12 | 0.42 | HAS1* | 0.56 | -1.43 |
| SERPING1 | 0.04 | 0.31 | IGFBP3 | 0.09 | 0.53 |
| THBS1 | 0.32 | 0.35 | LDHA | 0.05 | 0.09 |
| WDR1 | 0.19 | -0.70 | LOX | 0.12 | 0.60 |
|  |  |  | MT1E | 0.17 | 0.87 |
|  |  |  | MT2A* | 0.68 | 0.75 |
|  |  |  | NCAN* | 10.00 | -4.08 |
|  |  |  | NOCT | 0.38 | -0.33 |
|  |  |  | P4HA2 | 0.10 | 0.40 |
|  |  |  | PLIN2 | 0.05 | -0.51 |
|  |  |  | PPARGC1A | 0.22 | -1.28 |
|  |  |  | PPP1R15A* | 0.59 | 0.92 |
|  |  |  | PRDX5 | 0.06 | 0.29 |
|  |  |  | SLC2A1 | 0.19 | -0.17 |
|  |  |  | SLC2A3* | 0.63 | -0.42 |
|  |  |  | SRPX | 0.10 | 0.35 |
|  |  |  | STBD1* | 1.79 | -0.01 |
|  |  |  | SULT2B1 | 0.12 | 0.26 |
|  |  |  | TPI1 | 0.04 | -0.39 |

IDR: IDR value from combined IBC patient FFPE tumor datasets

genes with IDR>0.5 were marked by *

LFC: log_2_ Fold Change in IBC versus non-IBC patient FFPE tumor datasets

**Table S4. Hallmark gene set genes with high ΔIDRs (>2) that were not significantly differentially over-expressed (LFC<1) in IBC versus healthy and non-IBC PBMC for reads mapped to the transcriptome reference sequences.**

| Inflammatory Response | | | | TNFα-via-NFκB | | | | Epithelial Mesenchymal Transition | | | |
| --- | --- | --- | --- | --- | --- | --- | --- | --- | --- | --- | --- |
| ID | IDR | LFC | | ID | IDR | LFC | | ID | IDR | LFC | |
|  |  | Healthy | non-IBC |  |  | Healthy | non-IBC |  |  | Healthy | non-IBC |
| ABCA1* | 0.50 | 0.20 | 0.53 | ABCA1* | 0.50 | 0.20 | 0.53 | ADAM12* | 0.55 | -1.88 | -2.05 |
| ACVR2A* | 0.72 | -1.06 | -0.19 | ATP2B1* | 0.54 | 0.16 | 0.41 | BASP1 | 0.07 | 0.80 | 0.89 |
| ADGRE1 | 0.05 | -1.12 | -0.45 | B4GALT1 | 0.29 | -0.56 | -0.53 | CADM1* | 0.79 | -0.56 | -1.62 |
| ADM* | 0.65 | 0.80 | 0.92 | B4GALT5 | 0.32 | -0.22 | -0.02 | CD44 | 0.35 | -0.78 | -0.83 |
| AHR | 0.05 | -0.12 | 0.01 | BCL6* | 0.53 | 0.59 | -0.17 | COL5A2 | 0.21 | 0.85 | 0.73 |
| AQP9 | 0.17 | 0.12 | 0.63 | CD44 | 0.35 | -0.78 | -0.83 | COL6A3 | 0.19 | -0.33 | -0.78 |
| ATP2B1* | 0.54 | 0.16 | 0.41 | DENND5A | 0.15 | -0.30 | -0.06 | COPA | 0.25 | -0.80 | -0.32 |
| C5AR1 | 0.06 | 0.06 | 0.32 | DRAM1 | 0.10 | -0.59 | -0.71 | EDIL3* | 10.00 | -1.36 | -3.27 |
| CYBB | 0.14 | -1.16 | -0.82 | EGR2* | 0.53 | 0.04 | 0.52 | FBLN2 | 0.18 | -1.47 | -0.84 |
| DCBLD2 | 0.34 | -1.09 | -0.41 | F2RL1 | 0.07 | -0.65 | -0.14 | FBN2 | 0.05 | -0.44 | -0.70 |
| GCH1 | 0.19 | -0.82 | -0.07 | GCH1 | 0.19 | -0.82 | -0.07 | ITGAV | 0.26 | -0.97 | -0.37 |
| GPC3* | 3.99 | -4.43 | -2.87 | GFPT2* | 0.76 | 0.56 | -0.08 | ITGB1 | 0.06 | -1.19 | -0.62 |
| GPR132 | 0.40 | -0.33 | -0.25 | LAMB3* | 0.64 | 0.85 | 0.45 | LAMA2 | 0.25 | -1.85 | -0.85 |
| IL1R1 | 0.15 | 0.01 | -0.31 | LITAF | 0.06 | -0.39 | -0.16 | LAMC1 | 0.08 | -0.78 | -1.00 |
| IRAK2 | 0.16 | -0.56 | -0.36 | NFAT5 | 0.39 | -0.09 | 0.07 | LOXL1* | 0.75 | -0.50 | -0.27 |
| LPAR1 | 0.47 | -1.12 | -1.52 | NFKB1 | 0.48 | -0.44 | -0.17 | NOTCH2 | 0.13 | -0.47 | -0.74 |
| LYN | 0.44 | -0.03 | -0.60 | PDLIM5 | 0.15 | -0.49 | -0.26 | NTM* | 1.19 | -4.30 | -0.93 |
| MSR1 | 0.02 | -0.79 | 0.14 | PLAUR | 0.18 | 0.22 | 0.88 | PDGFRB | 0.13 | -0.66 | -0.30 |
| NFKB1 | 0.48 | -0.44 | -0.17 | PLEK | 0.21 | -0.77 | -0.11 | PLAUR | 0.18 | 0.22 | 0.88 |
| NLRP3 | 0.20 | -0.24 | 0.59 | PLPP3 | 0.44 | -1.43 | -0.74 | PLOD2 | 0.04 | -2.06 | -0.92 |
| PLAUR | 0.18 | 0.22 | 0.88 | PMEPA1* | 1.17 | -0.55 | -1.22 | PMEPA1* | 1.17 | -0.55 | -1.22 |
| PSEN1 | 0.24 | -0.64 | -0.31 | RELB | 0.32 | 0.19 | 0.09 | PMP22* | 0.51 | -0.24 | -0.32 |
| PTAFR | 0.04 | -0.38 | -0.31 | SLC16A6 | 0.42 | -0.18 | 0.27 | SERPINE2 | 0.04 | -1.05 | -0.72 |
| RASGRP1 | 0.34 | -0.94 | -0.52 | SPHK1 | 0.13 | -1.37 | -0.99 | SLIT3* | 10.00 | -1.46 | -2.61 |
| RHOG | 0.12 | -0.15 | -1.02 | TANK | 0.43 | -0.44 | 0.27 | SPOCK1* | 0.53 | -0.46 | -0.99 |
| RNF144B | 0.26 | -0.67 | -0.54 | TSC22D1 | 0.07 | -1.38 | -0.30 | TGFB1 | 0.10 | 0.20 | -0.77 |
| SGMS2 | 0.18 | -0.57 | -0.28 |  |  |  |  | TGFBR3 | 0.16 | -1.02 | -0.67 |
| SLC4A4 | 0.10 | -0.65 | -0.26 |  |  |  |  | TIMP1 | 0.08 | -0.45 | -0.98 |
| SLC7A1* | 0.52 | -0.60 | -0.95 |  |  |  |  | VCAN | 0.17 | -0.27 | -0.24 |
| SPHK1 | 0.13 | -1.37 | -0.99 |  |  |  |  | WIPF1 | 0.10 | -0.74 | -0.75 |
| TIMP1 | 0.08 | -0.45 | -0.98 |  |  |  |  |  |  |  |  |
| TNFRSF1B | 0.11 | -0.65 | -0.74 |  |  |  |  |  |  |  |  |

**Table S4 (Continued).**

| Coagulation | | | | Interferon Gamma Response | | | |
| --- | --- | --- | --- | --- | --- | --- | --- |
| ID | IDR | LFC | | ID | IDR | LFC | |
|  |  | Healthy | non-IBC |  |  | Healthy | non-IBC |
| ARF4 | 0.20 | -0.92 | -0.58 | ARID5B | 0.36 | -0.71 | -0.50 |
| C1S* | 2.72 | -2.35 | -1.69 | C1S* | 2.72 | -2.35 | -1.69 |
| CTSB | 0.14 | -1.12 | -0.83 | CD38 | 0.16 | -1.49 | -1.17 |
| CTSK* | 0.63 | -0.50 | -0.71 | CD74 | 0.10 | -0.91 | -1.18 |
| FYN | 0.29 | -0.62 | -0.41 | CD86 | 0.15 | -1.27 | -0.65 |
| KLF7* | 0.53 | -0.47 | -0.60 | GCH1 | 0.19 | -0.82 | -0.07 |
| MSRB2 | 0.18 | -1.42 | -0.75 | IL7*^1,2^ | 0.53 | -2.27 | -1.34 |
| PLEK | 0.21 | -0.77 | -0.11 | IRF4^3^ | 0.30 | -0.88 | -1.15 |
| PREP | 0.37 | -1.17 | -1.18 | LGALS3BP | 0.17 | -1.33 | -0.85 |
| PRSS23 | 0.04 | -1.93 | -0.80 | NCOA3 | 0.16 | -0.72 | -0.53 |
| RABIF* | 0.58 | -0.79 | -1.01 | NFKB1 | 0.48 | -0.44 | -0.17 |
| RAC1 | 0.17 | -0.98 | -1.40 | OASL | 0.20 | -0.25 | 0.93 |
| SH2B2 | 0.33 | 0.24 | -0.25 | P2RY14 | 0.21 | -1.56 | -0.70 |
| TIMP1 | 0.08 | -0.45 | -0.98 | PLSCR1* | 0.51 | -0.89 | -0.28 |
|  |  |  |  | PNPT1 | 0.23 | -0.85 | -0.22 |
|  |  |  |  | SAMHD1 | 0.12 | -1.03 | -0.98 |
|  |  |  |  | SPPL2A | 0.32 | -0.54 | -0.22 |
|  |  |  |  | SSPN | 0.33 | -0.97 | -0.59 |
|  |  |  |  | ST8SIA4 | 0.36 | -0.68 | -0.06 |
|  |  |  |  | STAT3^4,5^ | 0.17 | -0.57 | -0.31 |
|  |  |  |  | TRIM14 | 0.19 | -0.89 | -0.92 |

IDR: IDR value from combined IBC patient FFPE tumor datasets

genes with IDR>0.5 were marked by *

LFC: log_2_ Fold Change in IBC versus non-IBC patient and healthy donor PBMC datasets
