## Supplemental Figures for "TGIRT-seq of Inflammatory Breast Cancer Tumor and Blood Samples Reveals Widespread Enhanced Transcription Impacting RNA Splicing and Intronic RNAs in Plasma"

### Figure S1

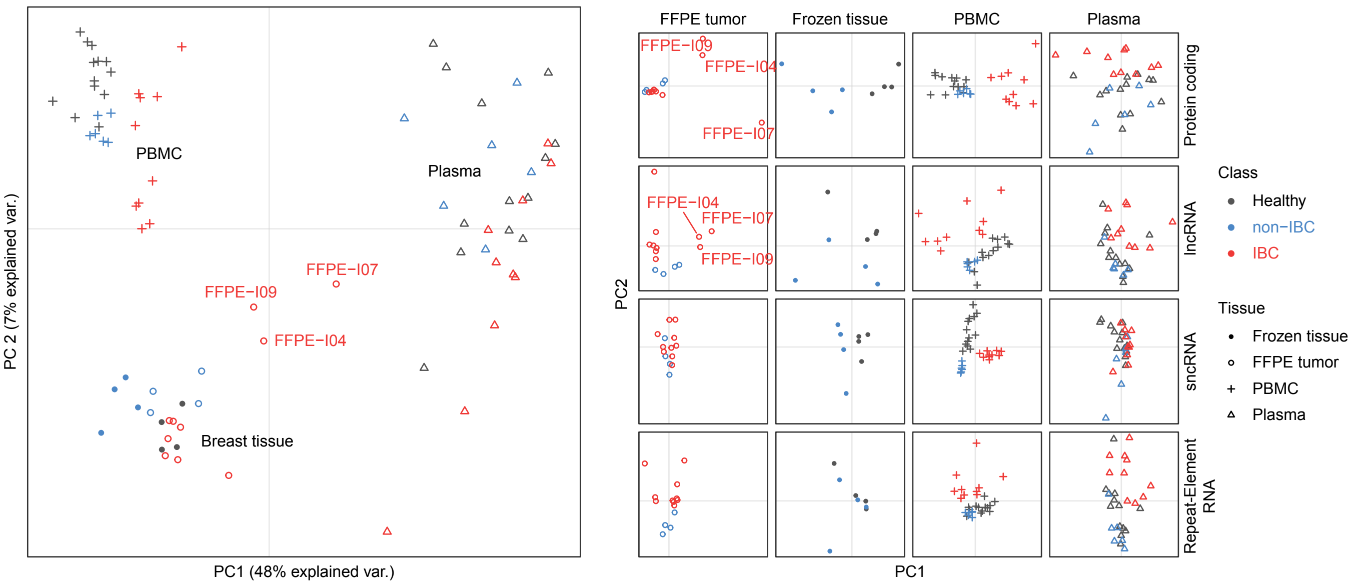

PBMC
Plasma
Breast tissue
FFPE−I09
FFPE−I07
FFPE−I04
PC 2 (7% explained var.)
PC1 (48% explained var.)
FFPE tumor
Frozen tissue
PBMC
Plasma
Protein coding
lncRNA
sncRNA
Repeat-Element RNA
PC2
PC1
Class
Healthy
non−IBC
IBC
Tissue
Frozen tissue
FFPE tumor
PBMC
Plasma

**Fig. S1. Principal Component Analysis.** PCA applied to all genes in all sample sets (left panel) and PCA run separately on genes encoding different categories of RNA biotypes in each sample type (right panels) color coded as shown to the right of the plots. For all samples considered together (left panel), the first principal component (PC1) separated the plasma samples (right) from the tumor and PBMC samples, while the second principal component (PC2) split the PBMC samples (+, top left) from frozen breast tissue and FFPE tumor samples. Seven of the 10 IBC FFPE tumor samples (red open circles) clustered on one side of the breast tissue samples without overlapping the FFPE or frozen non-IBC breast tissue samples (blue open or closed circles, respectively), while the remaining 3 IBC tumor samples (FFPE-I04, FFPE-I07, and FFPE-I09) were outliers with lower gene detection rates than the other FFPE tumor samples (see heat map Fig. 3A). Frozen healthy breast tissue samples were separated from frozen non-IBC breast tissue samples, but overlapped the IBC FFPE tumor samples, limiting qualifying comparisons between frozen and FFPE breast tissue samples. PCAs based on single categories of cellular RNA biotypes (Protein coding, lncRNAs, sncRNAs, and Repeat-Element RNAs, right panels) largely separated IBC patients from healthy donor and non-IBC patient samples in all cases, except for sncRNAs in FFPE tumor samples, likely reflecting limited differential expression of sncRNA between IBC and non-IBC FFPE tumor samples (Fig. 3).

### Figure S2

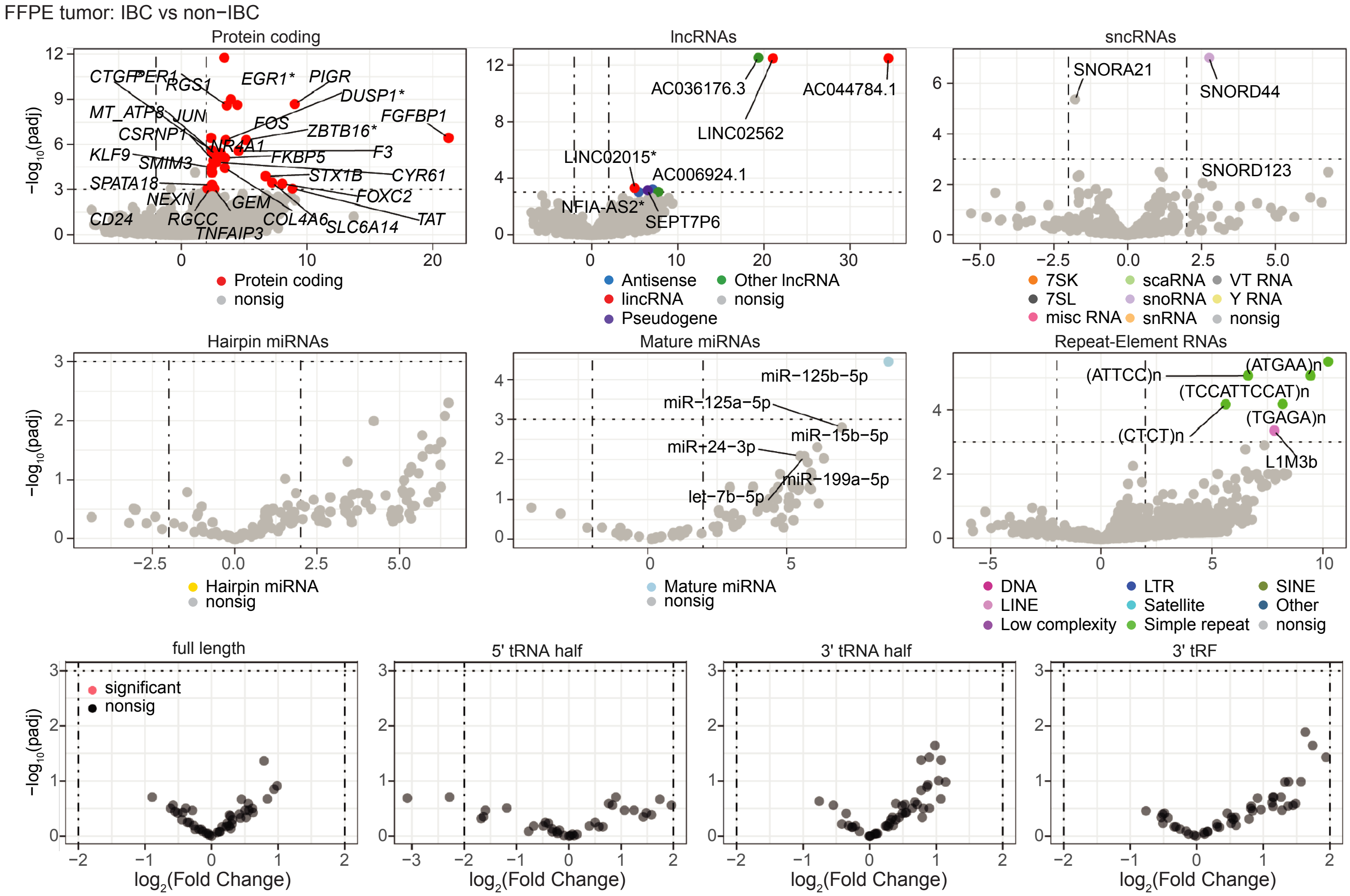
FFPE tumor: IBC vs non−IBC
Protein coding
lncRNAs
sncRNAs
−log10(padj)
CTGF
PER1
RGS1
EGR1*
PIGR
DUSP1*
MT_ATP8
JUN
FGFBP1
CSRNP1
FOS
ZBTB16*
NR4A1
KLF9
FKBP5
F3
SMIM3
STX1B
CYR61
SPATA18
FOXC2
NEXN
GEM
CD24
RGCC
COL4A6
TAT
TNFAIP3
SLC6A14
Protein coding
nonsig
AC036176.3
AC044784.1
LINC02562
LINC02015*
AC006924.1
NFIA-AS2*
SEPT7P6
Antisense
lincRNA
Pseudogene
Other lncRNA
nonsig
SNORA21
SNORD44
SNORD123
7SK
7SL
misc RNA
scaRNA
snoRNA
snRNA
VT RNA
Y RNA
nonsig
Hairpin miRNAs
Mature miRNAs
Repeat-Element RNAs
miR−125b−5p
miR−125a−5p
miR−15b−5p
miR−24−3p
miR−199a−5p
let−7b−5p
(ATTCC)n
(ATGAA)n
(TCCATTCCAT)n
(TGAGA)n
(CTCT)n
L1M3b
Hairpin miRNA
nonsig
Mature miRNA
nonsig
DNA
LINE
Low complexity
LTR
Satellite
Simple repeat
SINE
Other
nonsig
full length
5' tRNA half
3' tRNA half
3' tRF
significant
nonsig
log2(Fold Change)

**Fig. S2. Volcano plots for DESeq2 comparisons of FFPE breast tissue samples.** The *y*-axis indicates the $\log_{10}$-transformed adj. *p*-value while the *x*-axis indicates the DESeq2-estimated LFC. Each point represents one gene with biotypes color coded for those genes with adj. $p \leq 0.001$ and $|LFC|>2$. Significantly over-represented and other notable genes that passed filtering steps and were detected in at least 50% of the samples of the group in which they were over-represented are labeled by their gene symbol. Protein-coding and lncRNA genes with IDR>0.5 are marked with an asterisk. nonsig: genes that are not significantly differentially expressed.

### Figure S3

#### A. Protein-coding genes

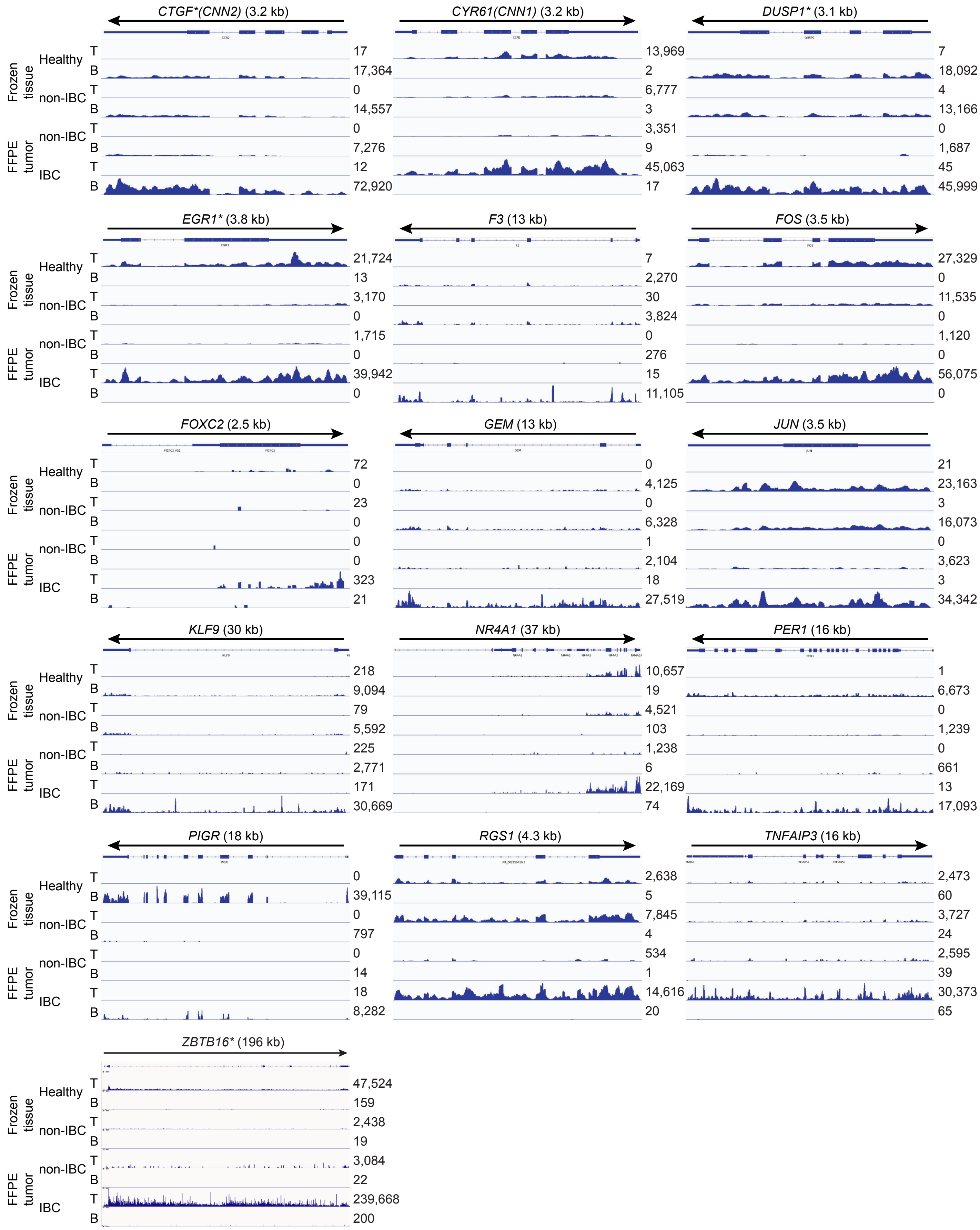

#### Figure S3

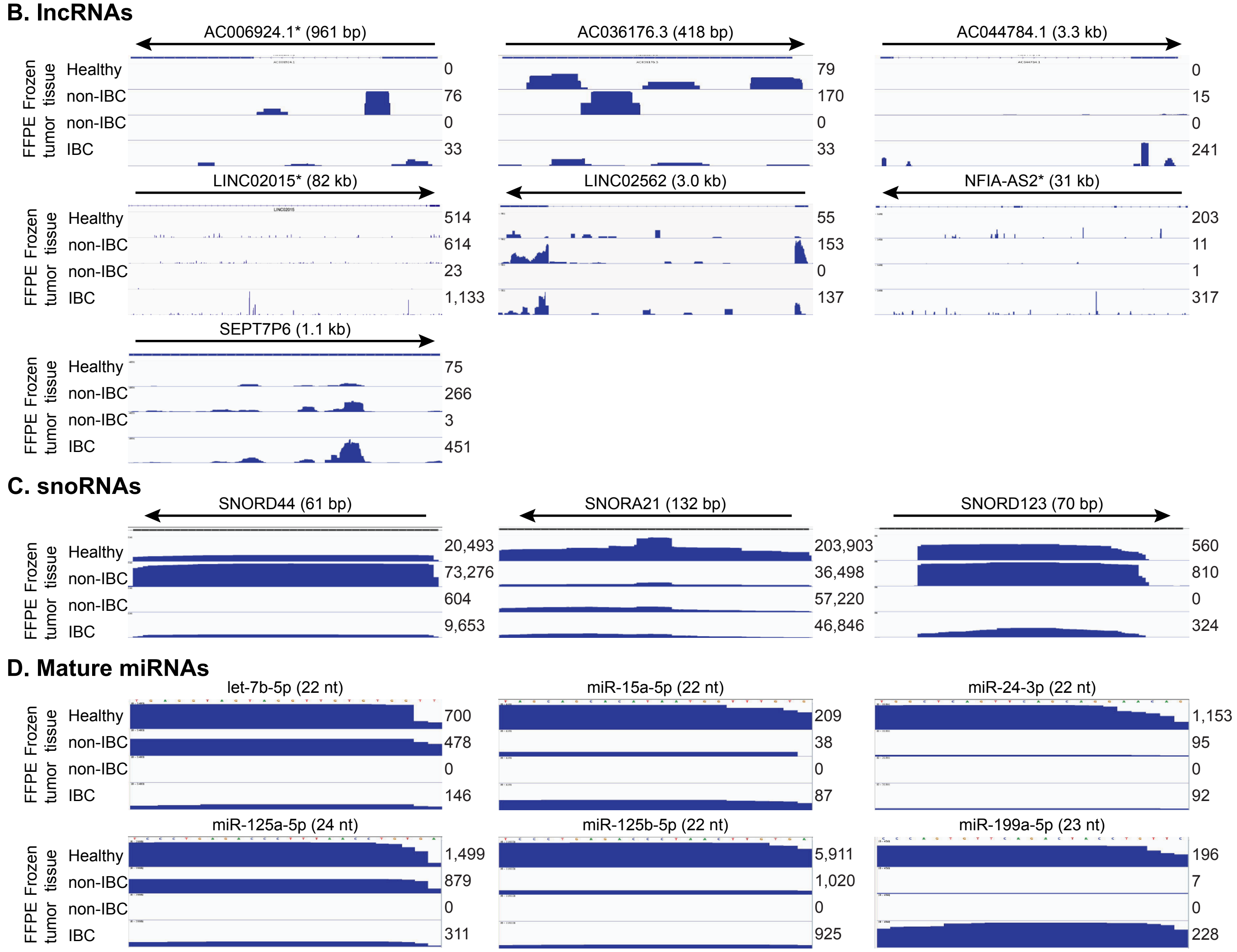
B. lncRNAs
AC006924.1* (961 bp)
AC036176.3 (418 bp)
AC044784.1 (3.3 kb)
Frozen tissue
FFPE tumor
Healthy
non-IBC
non-IBC
IBC
0
76
0
33
79
170
0
33
0
15
0
241
LINC02015* (82 kb)
LINC02562 (3.0 kb)
NFIA-AS2* (31 kb)
514
614
23
1,133
55
153
0
137
203
11
1
317
SEPT7P6 (1.1 kb)
75
266
3
451
C. snoRNAs
SNORD44 (61 bp)
SNORA21 (132 bp)
SNORD123 (70 bp)
20,493
73,276
604
9,653
203,903
36,498
57,220
46,846
560
810
0
324
D. Mature miRNAs
let-7b-5p (22 nt)
miR-15a-5p (22 nt)
miR-24-3p (22 nt)
700
478
0
146
209
38
0
87
1,153
95
0
92
miR-125a-5p (24 nt)
miR-125b-5p (22 nt)
miR-199a-5p (23 nt)
1,499
879
0
311
5,911
1,020
0
925
196
7
0
228

**Fig. S3. IGV plots of differentially expressed or notable genes in breast tissue samples.** (**A**) protein-coding genes, (**B**) lncRNAs, (**C**) snoRNAs, and (**D**) mature miRNAs. T and B, top strand and bottom DNA strands, respectively. The number of reads for RNAs in each IGV plot is indicated to the right. The primary comparison in Results is for FFPE IBC versus FFPE non-IBC patient tumors. Protein-coding and lncRNA genes with IDR>0.5 are marked with an asterisk.

#### Figure S4

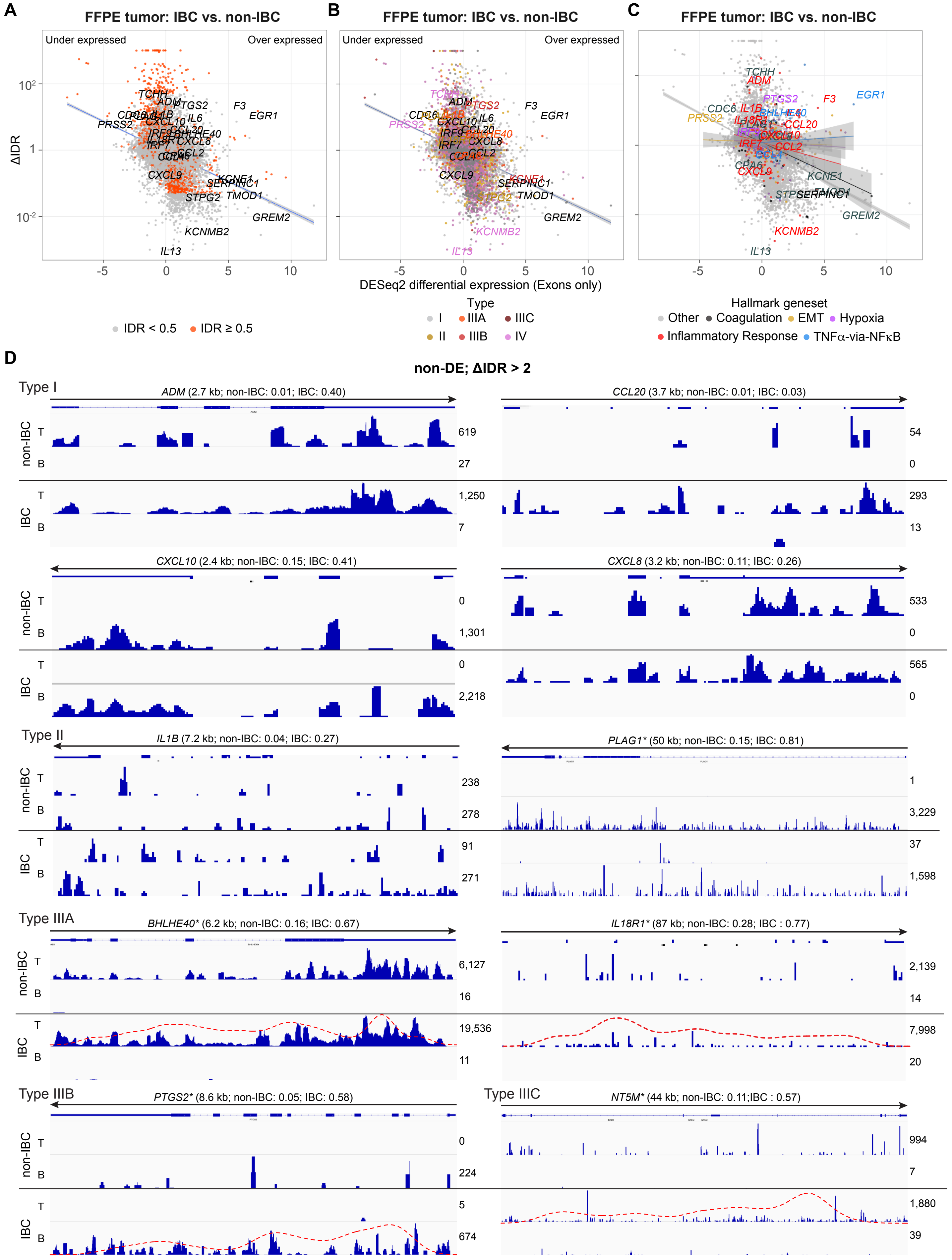

A
FFPE tumor: IBC vs. non-IBC
Under expressed
Over expressed
ΔIDR
IDR < 0.5
IDR ≥ 0.5
B
FFPE tumor: IBC vs. non-IBC
Under expressed
Over expressed
DESeq2 differential expression (Exons only)
Type
I
IIIA
IIIC
II
IIIB
IV
C
FFPE tumor: IBC vs. non-IBC
Hallmark geneset
Other
Coagulation
EMT
Hypoxia
Inflammatory Response
TNFα-via-NFκB
TCHH
ADM
PTGS2
F3
EGR1
CDC6
IL1B
IL6
PRSS2
CXCL10
CCL20
IRF9
BHLHE40
IRF7
CXCL8
CCL2
CCL4
CXCL9
KCNE1
SERPINC1
STPG2
TMOD1
GREM2
KCNMB2
IL13
D
non-DE; ΔIDR > 2
Type I
ADM (2.7 kb; non-IBC: 0.01; IBC: 0.40)
CCL20 (3.7 kb; non-IBC: 0.01; IBC: 0.03)
CXCL10 (2.4 kb; non-IBC: 0.15; IBC: 0.41)
CXCL8 (3.2 kb; non-IBC: 0.11; IBC: 0.26)
Type II
IL1B (7.2 kb; non-IBC: 0.04; IBC: 0.27)
PLAG1* (50 kb; non-IBC: 0.15; IBC: 0.81)
Type IIIA
BHLHE40* (6.2 kb; non-IBC: 0.16; IBC: 0.67)
IL18R1* (87 kb; non-IBC: 0.28; IBC : 0.77)
Type IIIB
PTGS2* (8.6 kb; non-IBC: 0.05; IBC: 0.58)
Type IIIC
NT5M* (44 kb; non-IBC: 0.11;IBC : 0.57)
non-IBC
IBC
T
B

**Fig. S4. High ΔIDR genes in IBC versus non-IBC FFPE tumors samples trend toward lower differential expression for reads mapped to the transcriptome reference. (A-C)** Plots of ΔIDR (*y*-axis) versus differential expression enrichment scores calculated by DESeq2 for reads mapped to the transcriptome reference sequence (*x*-axis). For each panel, different categories of genes are color coded as shown beneath the panels based on: (**A**) IDR in IBC tumor samples; (**B**) gene type based on the ratio of intron to exon read depth across the gene; (**C**) membership in Broad Institute Hallmark gene sets with separate trendlines shown for each gene set to highlight their varying patterns of differential expression and ΔIDRs. Select genes which were either: (i) differentially expressed; (ii) have particularly high or ΔIDR values; or (iii) represent the various intron expression patterns shown in panel (D) are named in the plot. **(D)** IGV plots of genes with high ΔIDRs (>2) that are not differentially expressed in IBC compared to non-IBC FFPE tumor samples for reads mapped to the transcriptome reference sequence. Examples are classified as Type I, II, IIIA, IIIB, and IIIC in IBC FFPE tumors based on definitions in Results illustrated in Fig. 2C. Gene length and IDR values for the gene in non-IBC and IBC datasets are shown in parentheses next to the gene name. The number of reads for RNAs in each IGV plot is indicated to the right. Red dashed lines are density plots of intron body coverage over the length of the gene. Protein-coding and lncRNA genes with IDR>0.5 are marked with an asterisk.

#### Figure S5

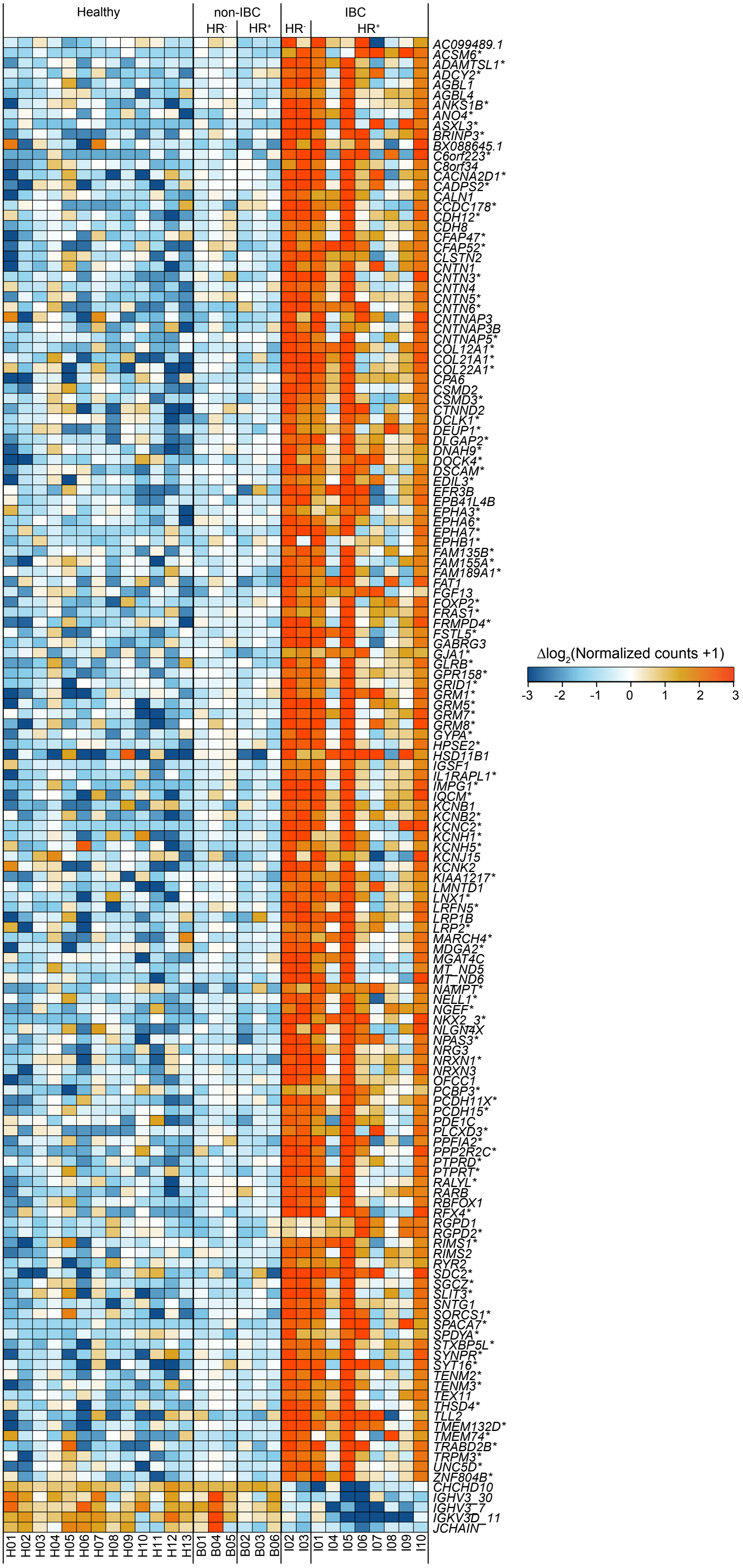

Healthy
non-IBC
HR-
HR+
IBC
HR-
HR+
AC099489.1
ACSM6*
ADAMTSL1*
ADCY2*
AGBL1
AGBL4
ANKS1B*
ANO4*
ASXL3*
BRINP3*
BX088645.1
C6orf223*
C8orf34
CACNA2D1*
CADPS2*
CALN1
CCDC178*
CDH12*
CDH8
CFAP47*
CFAP52*
CLSTN2
CNTN1
CNTN3*
CNTN4
CNTN5*
CNTN6*
CNTNAP3
CNTNAP3B
CNTNAP5*
COL12A1*
COL21A1*
COL22A1*
CPA6
CSMD2
CSMD3*
CTNND2
DCLK1*
DEUP1*
DLGAP2*
DNAH9*
DOCK4*
DSCAM*
EDIL3*
EFR3B
EPB41L4B
EPHA3*
EPHA6*
EPHA7*
EPHB1*
FAM135B*
FAM155A*
FAM189A1*
FAT1
FGF13
FOXP2*
FRAS1*
FRMPD4*
FSTL5*
GABRG3
GJA1*
GLRB*
GPR158*
GRID1*
GRM1*
GRM5*
GRM7*
GRM8*
GYPA*
HPSE2*
HSD11B1
IGSF1
IL1RAPL1*
IMPG1*
IQCM*
KCNB1
KCNB2*
KCNC2*
KCNH1*
KCNH5*
KCNJ15
KCNK2
KIAA1217*
LMNTD1
LNX1*
LRFN5*
LRP1B
LRP2*
MARCH4*
MDGA2*
MGAT4C
MT_ND5
MT_ND6
NAMPT*
NELL1*
NGEF*
NKX2_3*
NLGN4X
NPAS3*
NRG3
NRXN1*
NRXN3
OFCC1
PCBP3*
PCDH11X*
PCDH15*
PDE1C
PLCXD3*
PPFIA2*
PPP2R2C*
PTPRD*
PTPRT*
RALYL*
RARB
RBFOX1
RFX4*
RGPD1
RGPD2*
RIMS1*
RIMS2
RYR2
SDC2*
SGCZ*
SLIT3*
SNTG1
SORCS1*
SPACA7*
SPDYA*
STXBP5L*
SYNPR*
SYT16*
TENM2*
TENM3*
TEX11
THSD4*
TLL2
TMEM132D*
TMEM74*
TRABD2B*
TRPM3*
UNC5D*
ZNF804B*
CHCHD10
IGHV3_30
IGHV3_7
IGKV3D_11
JCHAIN
H01
H02
H03
H04
H05
H06
H07
H08
H09
H10
H11
H12
H13
B01
B04
B05
B02
B03
B06
I02
I03
I01
I04
I05
I06
I07
I08
I09
I10
Δlog2(Normalized counts +1)
-3
-2
-1
0
1
2
3

**Fig. S5. Heat map of the relative abundance for all 149 DE protein-coding genes (adj. *p*≤0.001, |LFC|≥2 in IBC PBMC samples relative to the healthy and non-IBC PBMC samples.** Tiles are colored by expression differences relative to the corresponding genewise means taken across all PBMC samples based on the color code top right. Lanes in the heat map are numbered according to the healthy donor or patient ID number (Table S1). Protein-coding genes with IDR>0.5 are marked with an asterisk.

### Figure S6

**A** PBMC: IBC vs Healthy

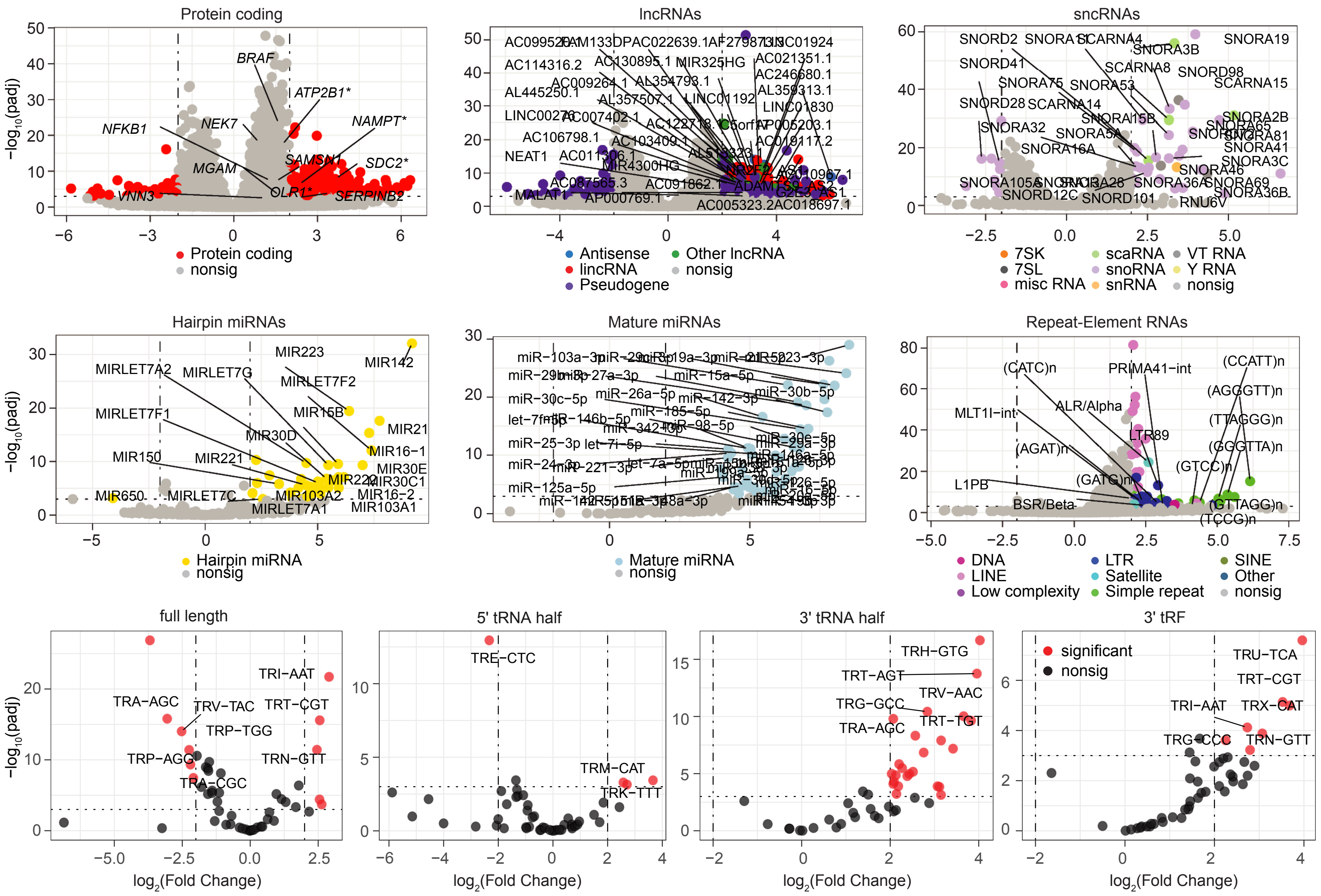

### Figure S6

**B** PBMC: IBC vs non−IBC

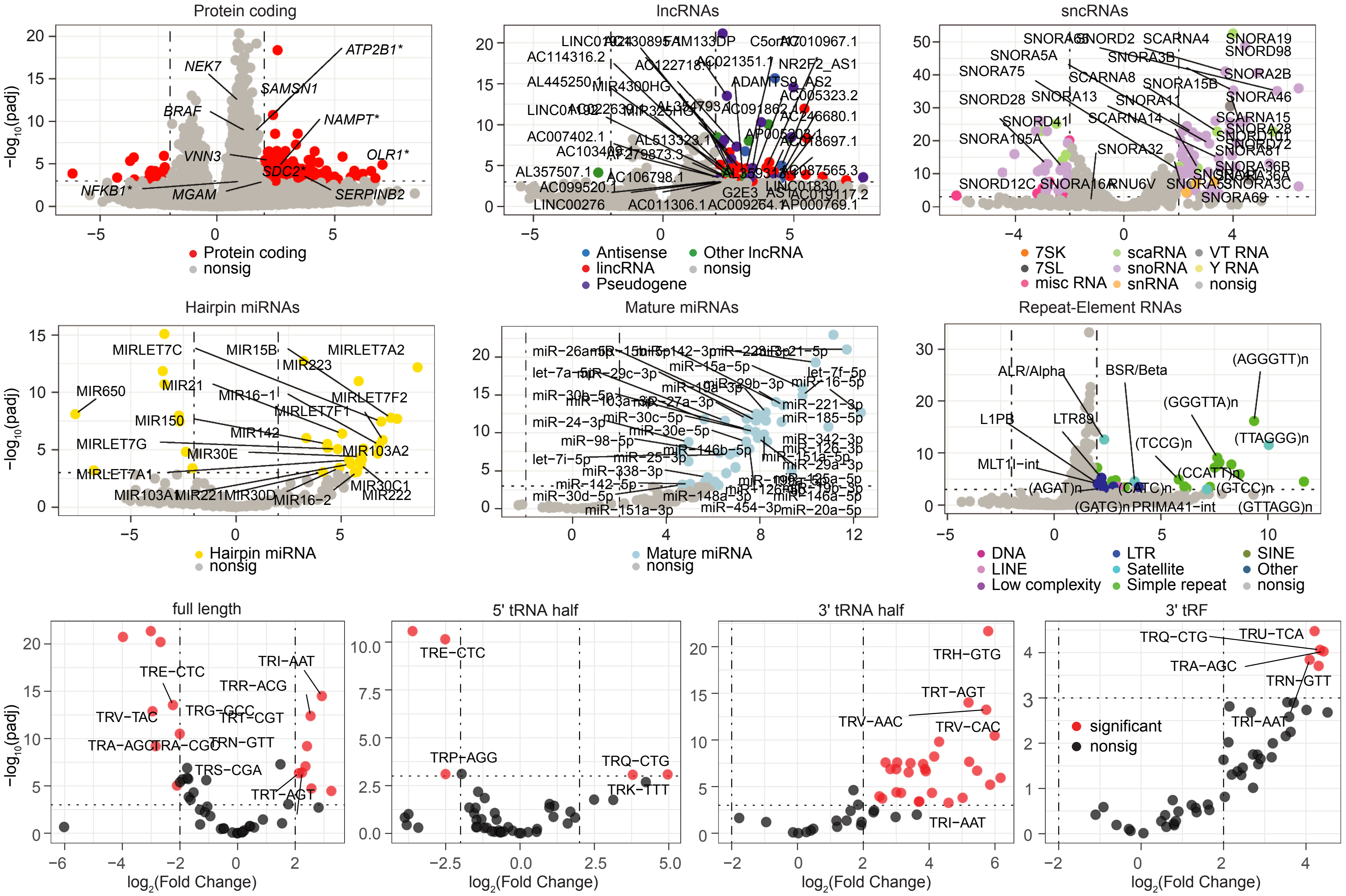

**Fig. S6. Volcano plots for DESeq2 comparisons of PBMC samples.** (**A**) IBC patient vs Healthy donor PBMCs. (**B**) IBC vs non-IBC patient PBMCs. The *y*-axis indicates $log_{10}$-transformed adj. *p*-values, while *x*-axis indicates LFC. Each point represents one gene with biotype color coded for those genes with adj. $p \leq 0.001$ and $|LFC|>2$. Significantly over-represented and other notable genes that passed filtering steps and were detected in at least 50% of the samples of the group in which they were over-represented are labeled by their gene symbol. Protein-coding genes with IDR>0.5 are marked with an asterisk; lncRNA genes with IDR>0.5 were not marked with an asterisk due to crowding of gene names (see Fig. 4 and Suppl. file for IDR status). nonsig: genes that are not significantly differentially expressed.

### Figure S7

#### A. Protein-coding genes

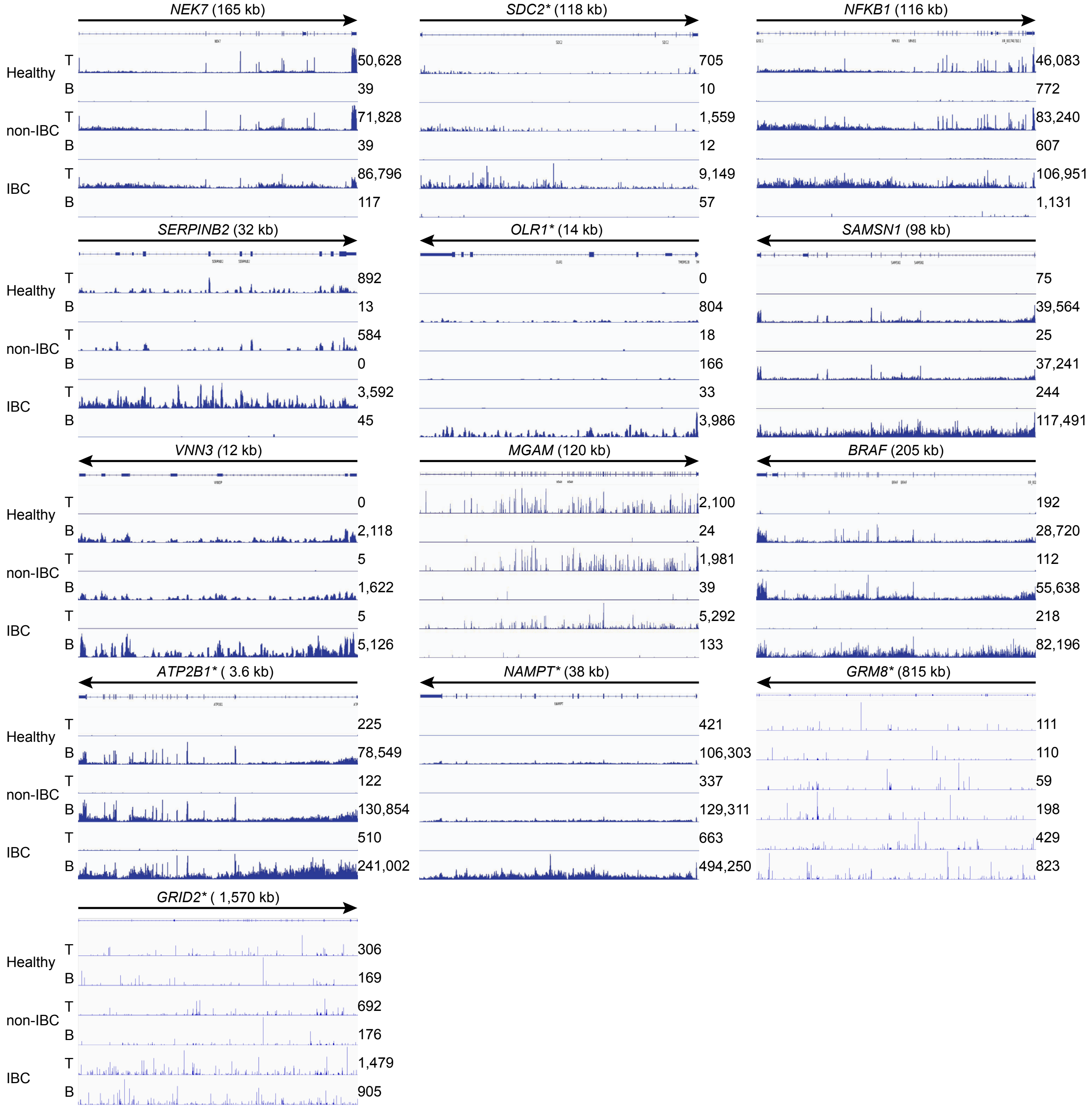

### Figure S7

#### B. lncRNAs

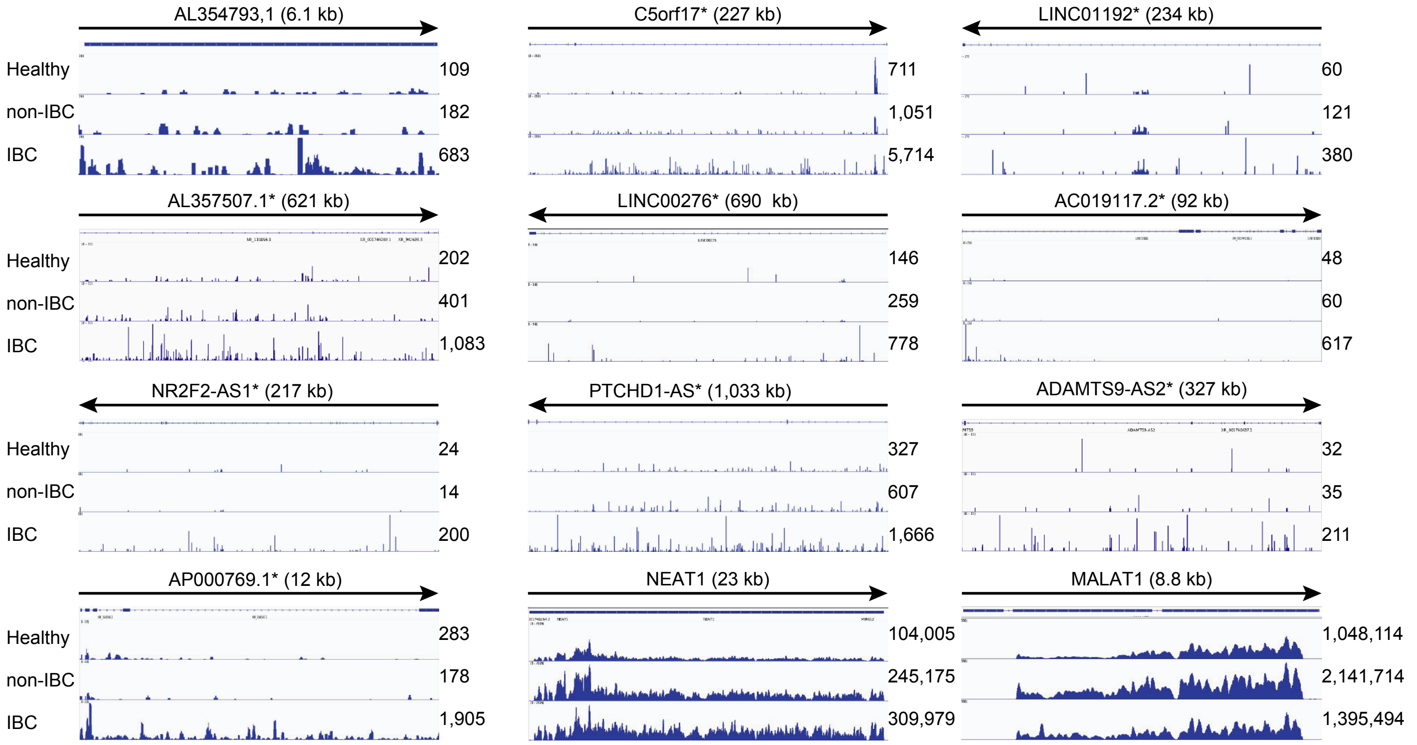


#### C. sncRNAs

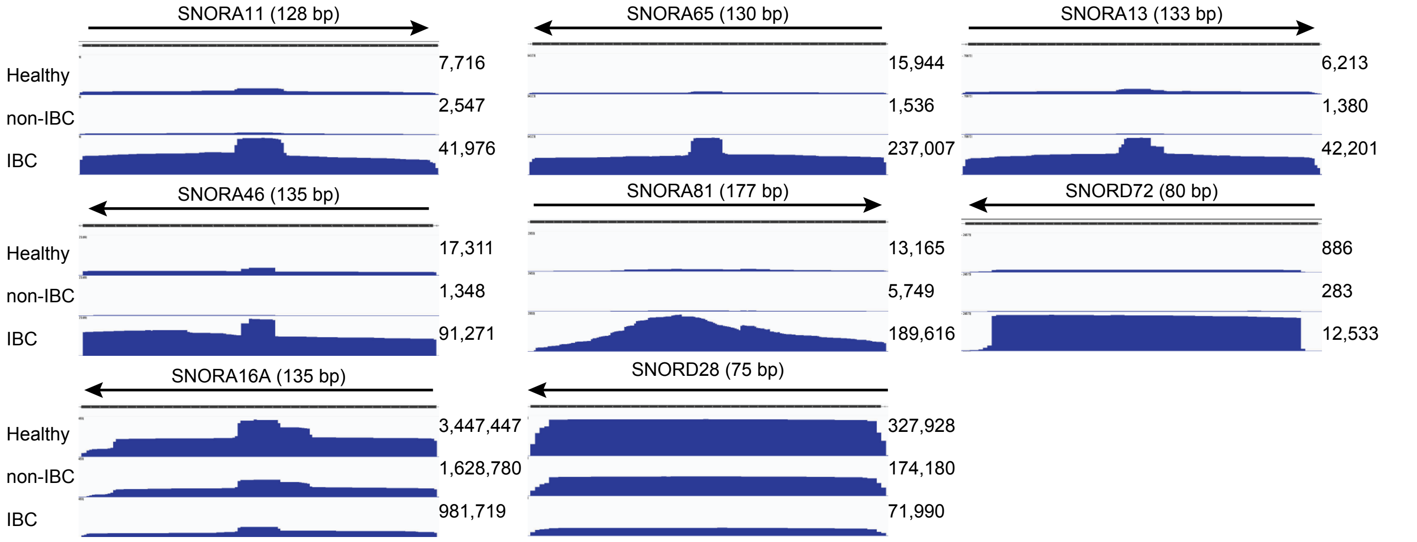

#### Figure S7

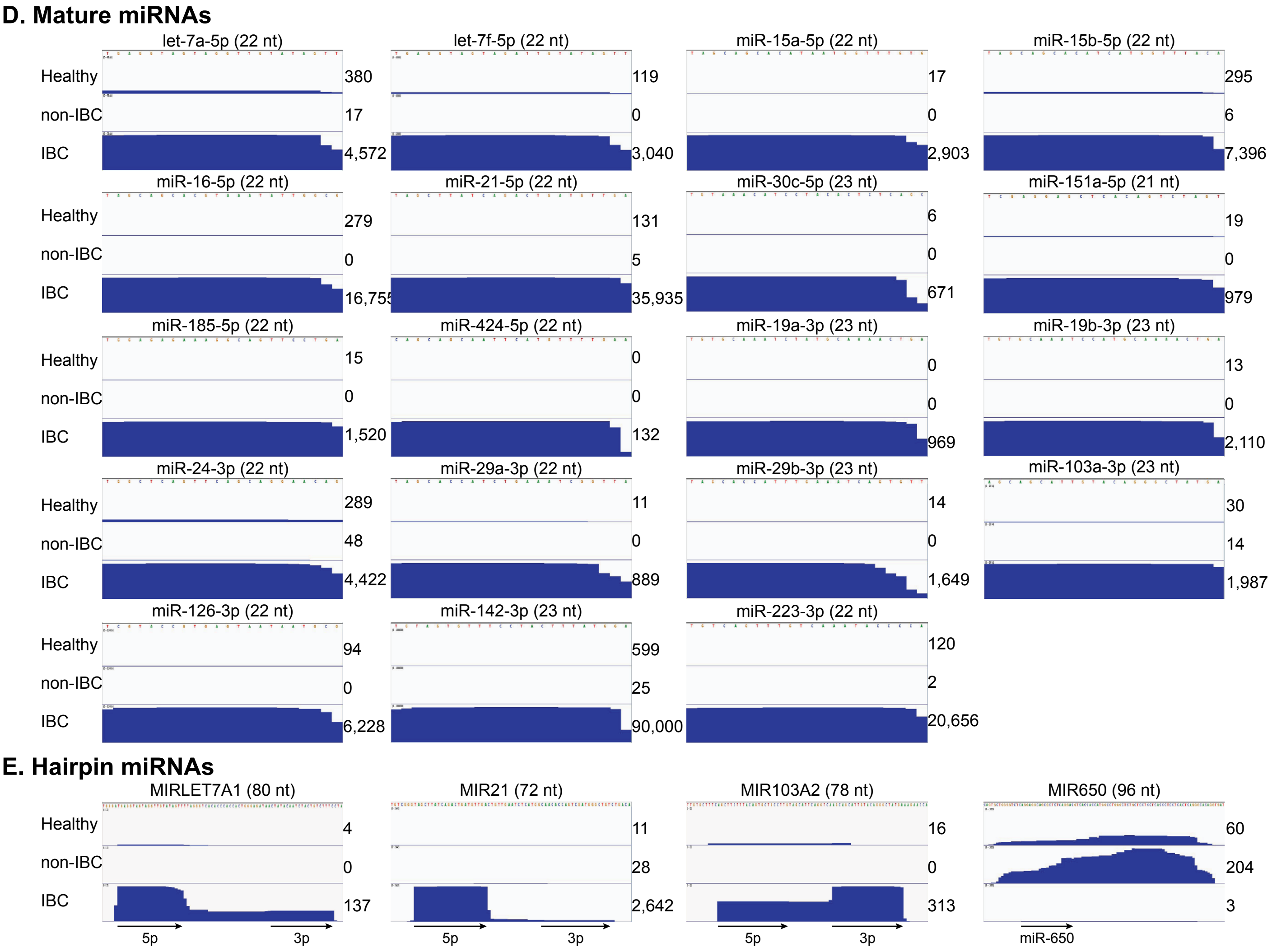
D. Mature miRNAs
let-7a-5p (22 nt)
let-7f-5p (22 nt)
miR-15a-5p (22 nt)
miR-15b-5p (22 nt)
Healthy
non-IBC
IBC
380
17
4,572
119
0
3,040
17
0
2,903
295
6
7,396
miR-16-5p (22 nt)
miR-21-5p (22 nt)
miR-30c-5p (23 nt)
miR-151a-5p (21 nt)
279
0
16,755
131
5
35,935
6
0
671
19
0
979
miR-185-5p (22 nt)
miR-424-5p (22 nt)
miR-19a-3p (23 nt)
miR-19b-3p (23 nt)
15
0
1,520
0
0
132
0
0
969
13
0
2,110
miR-24-3p (22 nt)
miR-29a-3p (22 nt)
miR-29b-3p (23 nt)
miR-103a-3p (23 nt)
289
48
4,422
11
0
889
14
0
1,649
30
14
1,987
miR-126-3p (22 nt)
miR-142-3p (23 nt)
miR-223-3p (22 nt)
94
0
6,228
599
25
90,000
120
2
20,656
E. Hairpin miRNAs
MIRLET7A1 (80 nt)
MIR21 (72 nt)
MIR103A2 (78 nt)
MIR650 (96 nt)
4
0
137
11
28
2,642
16
0
313
60
204
3
5p
3p
miR-650

**Fig. S7. IGV plots of differentially expressed or notable genes in PBMC samples.** (**A**) protein-coding genes, (**B**) lncRNAs, (**C**) sncRNAs, (**D**) mature miRNAs, and (**E**) hairpin miRNAs. T and B, top strand and bottom DNA strands, respectively. The number of reads for RNAs in each IGV plot is indicated to the right. Protein-coding and lncRNA genes with IDR>0.5 are marked with an asterisk.

#### Figure S8

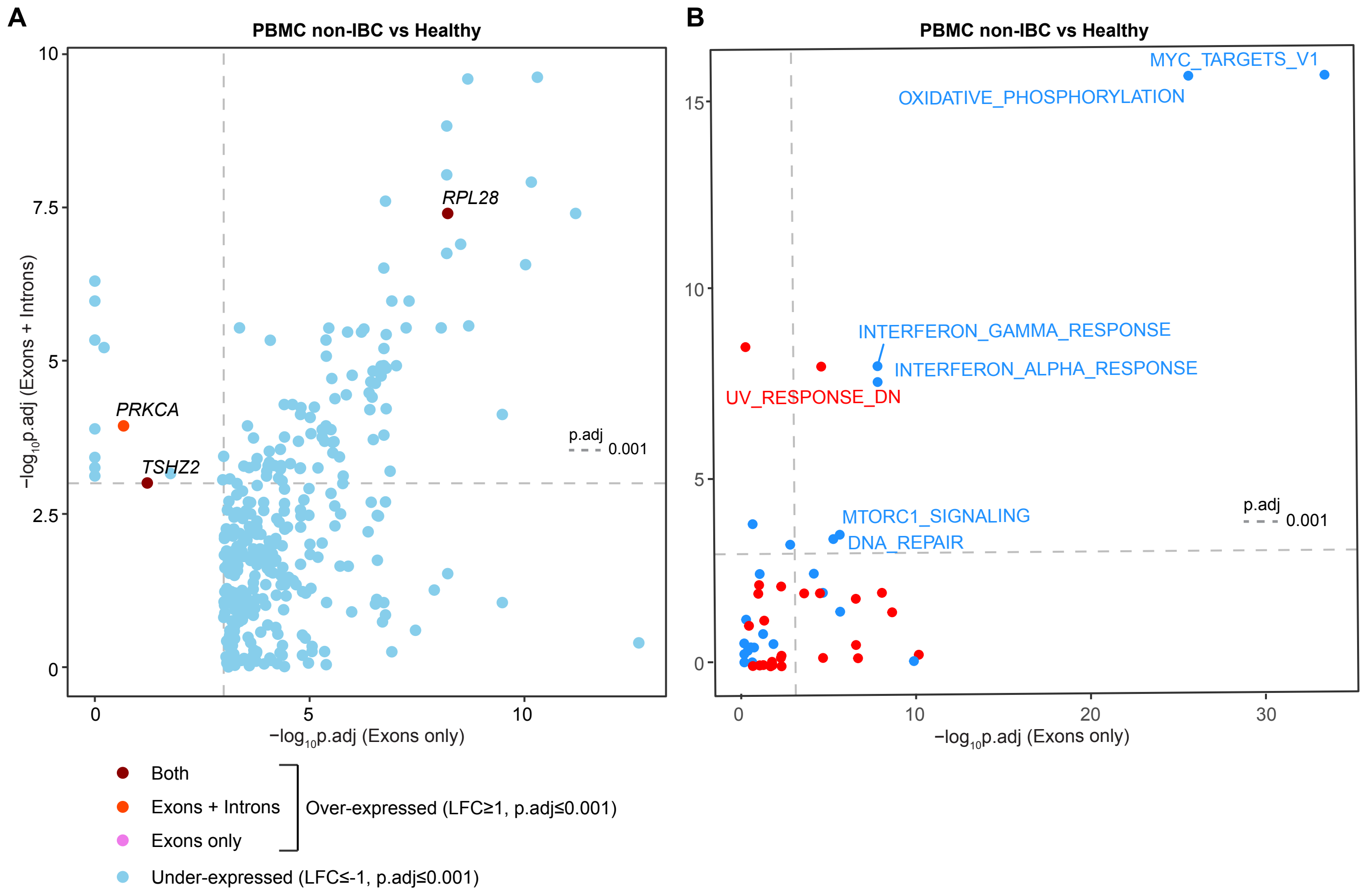

A
PBMC non-IBC vs Healthy
RPL28
PRKCA
TSHZ2
p.adj
0.001
−log10p.adj (Exons + Introns)
−log10p.adj (Exons only)
B
PBMC non-IBC vs Healthy
MYC_TARGETS_V1
OXIDATIVE_PHOSPHORYLATION
INTERFERON_GAMMA_RESPONSE
INTERFERON_ALPHA_RESPONSE
UV_RESPONSE_DN
MTORC1_SIGNALING
DNA_REPAIR
p.adj
0.001
−log10p.adj (Exons only)
Both
Exons + Introns
Exons only
Over-expressed (LFC≥1, p.adj≤0.001)
Under-expressed (LFC≤-1, p.adj≤0.001)

**Fig. S8. Non-IBC patient versus healthy donor PBMC scatter plots.** (**A**) Scatter plot comparing ($log_{10}$-transformed and sign-inverted) FDR-adjusted *p*-values for DE genes in non-IBC patient versus healthy donor PBMCs based on Exon only mapping reads (*x*-axis) versus Exon + Intron mapping reads (*y*-axis). Only genes for with LFC≥1 are shown, and only those for which the adj. *p*-value was ≤0.001 based on at least one of the two mapping strategies (Exons only or Exons + Introns) are named in the plots. (**B**) Scatter plot comparing ($log_{10}$-transformed and sign-inverted) FDR-adjusted *p*-values from GO-MWU non-IBC patient versus healthy donor analysis of BROAD Hallmark gene sets using gene counts generated by Exon only mapping reads (*x*-axis) or Exon + Intron mapping reads (*y*-axis). Gene sets for which the average gene MWU test statistic indicated increased expression in non-IBC are colored red, while those with decreased expression in non-IBC are colored blue. Vertical and horizontal dashed lines indicate adj. $p$=0.001.

#### Figure S9

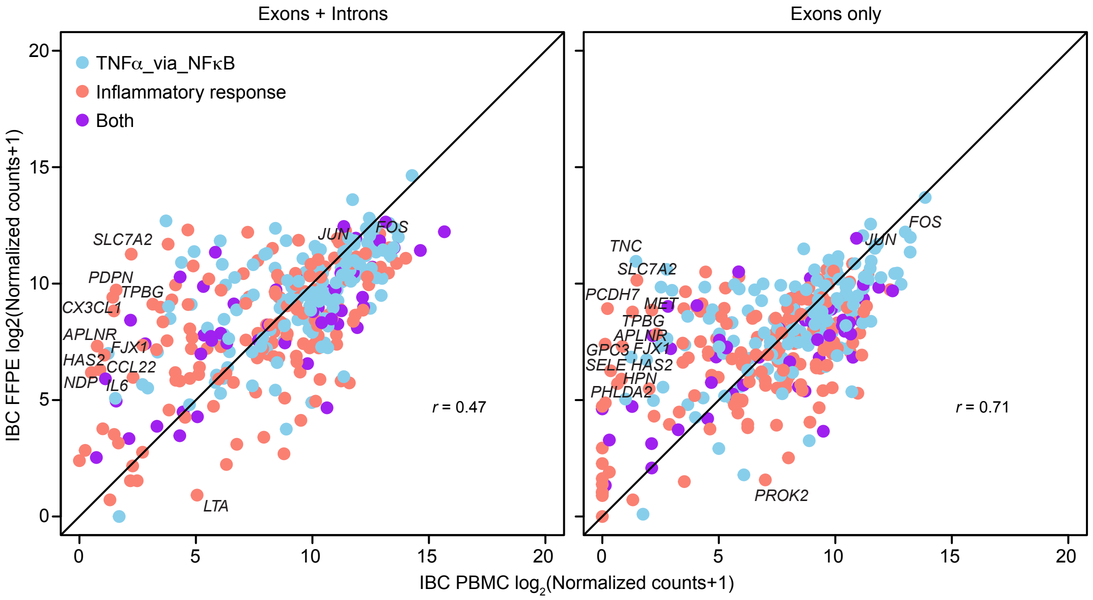

Exons + Introns
Exons only
TNFα_via_NFκB
Inflammatory response
Both
IBC FFPE log2(Normalized counts+1)
IBC PBMC log2(Normalized counts+1)
r = 0.47
r = 0.71
SLC7A2
PDPN
TPBG
CX3CL1
APLNR
FJX1
HAS2
CCL22
NDP
IL6
LTA
JUN
FOS
TNC
PCDH7
MET
GPC3
SELE
HPN
PHLDA2
PROK2
0
5
10
15
20

**Fig. S9. Hallmark TNFα-via-NFκB and Inflammatory Response gene scatter plots for reads mapped to genome and transcriptome reference sequences.** Scatter plots of $log_2$-transformed normalized counts of genes belonging to Hallmark TNFα-via-NFκB (blue) and Inflammatory Response (red) gene sets in IBC FFPE tumor samples (*y*-axis) versus IBC PBMC (*x*-axis) datasets. Genes belonging to both gene sets are colored in purple. Genes that have LFC≥2 are labeled along with *JUN* and *FOS*. Pearson correlation coefficients (*r*) are shown at the bottom right.

#### Figure S10

##### A Exons + Introns

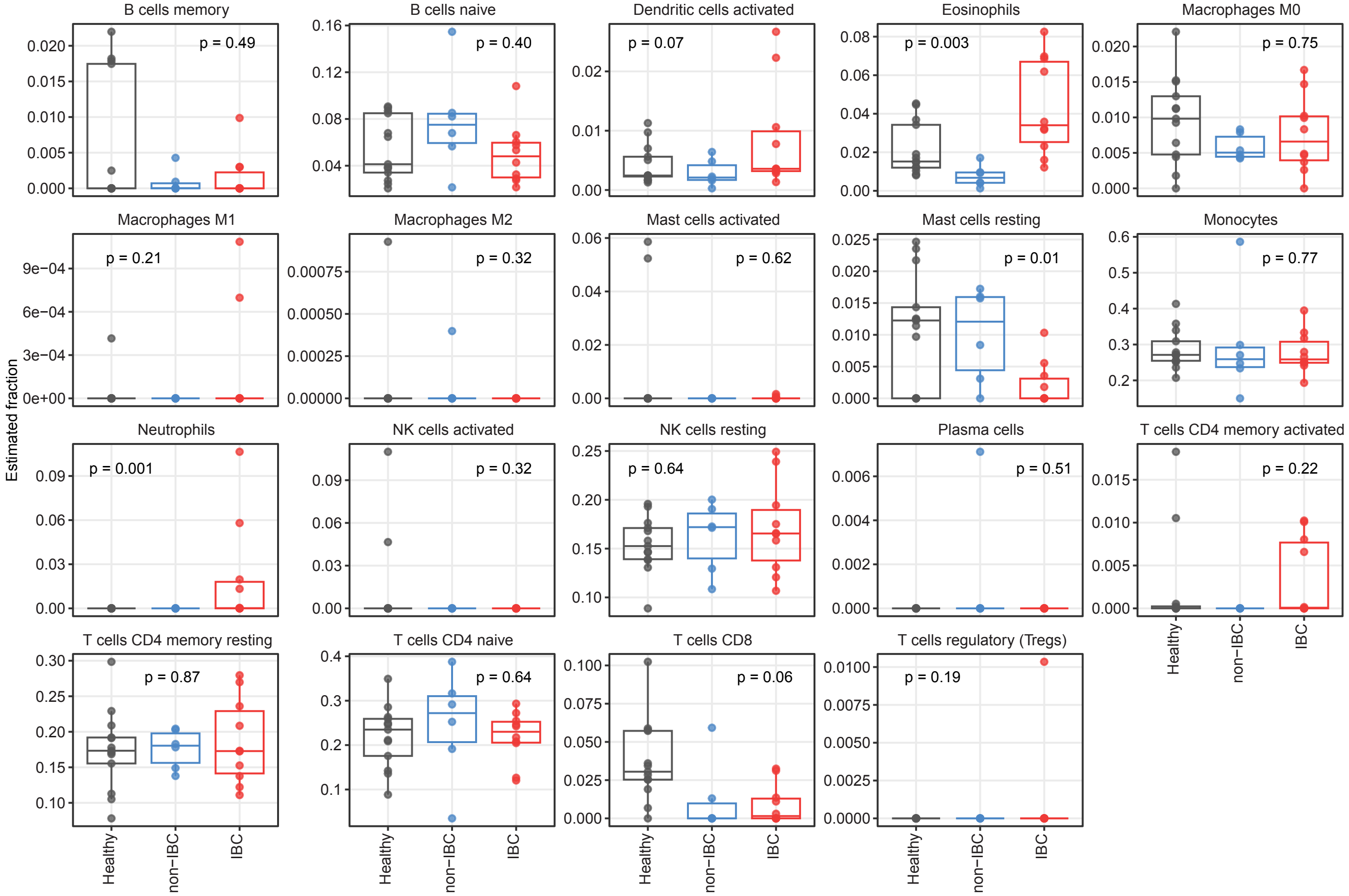


##### B Exons only

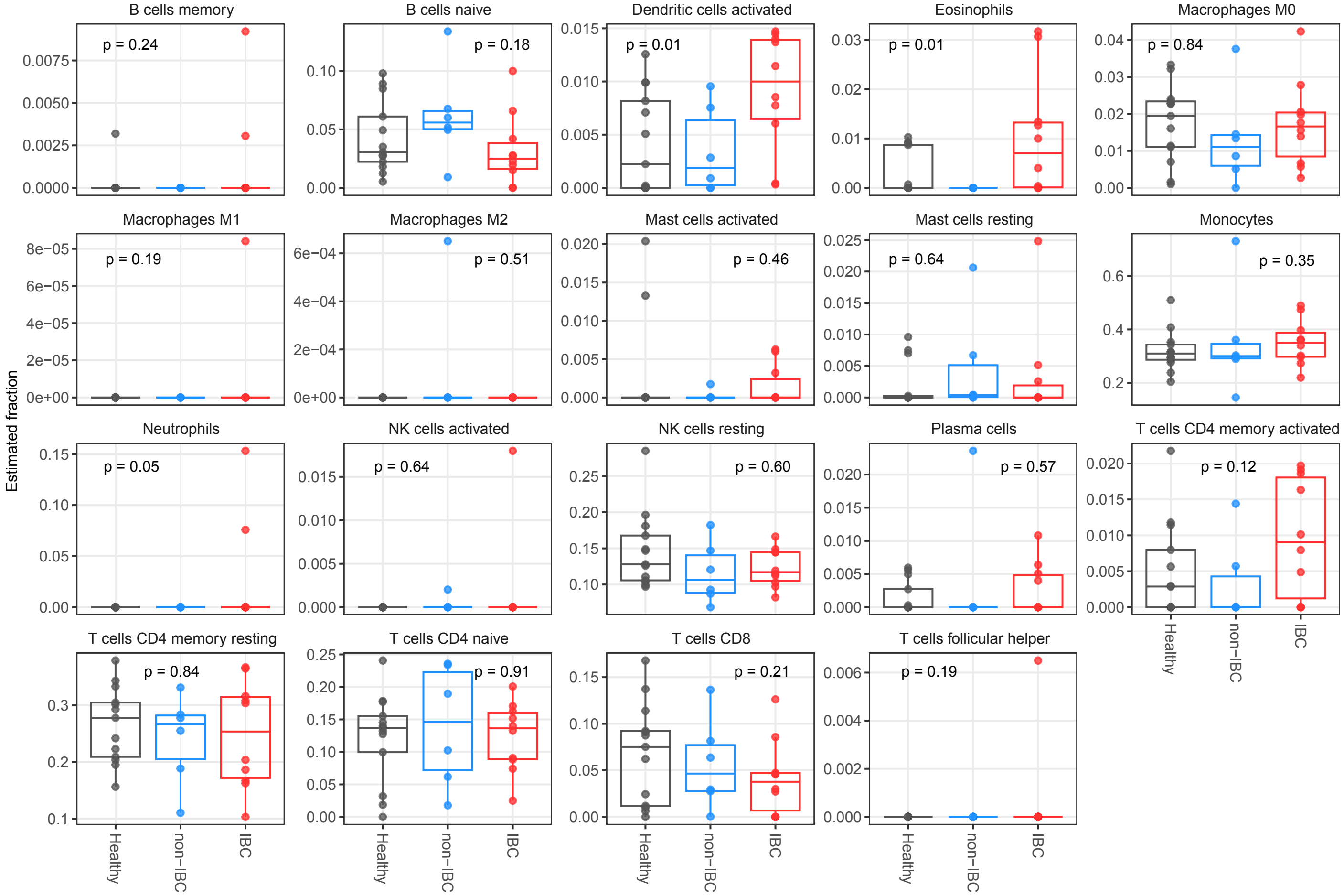

**Fig. S10. CIBERSORTx-estimated fractions of PBMC cell types for all samples grouped by diagnosis.** Points represent individual sample cell type fraction estimates (*y*-axis) separated by patient diagnosis (Healthy, non-IBC, or IBC). Midpoints of boxes indicate median values for each sample group, with bottom and top of boxes indicating $25^{th}$ and $75^{th}$ percentile estimates for within-group distributions of cell type fractions. **(A)** CIBERSORTx analyses conducted using Exon + Intron mapping gene counts, **(B)** CIBERSORTx analyses conducted using Exon only mapping gene counts. Mann-Whitney U-testing indicated significant differences between a number of groups: $p<0.05$ for eosinophil abundances in IBC PBMC samples compared to the combined healthy and non-IBC sample sets using either Exon only or Exon + Intron alignments; for neutrophils, $p=0.0011$ using Exon + Intron alignments and $p=0.053$ using Exon only alignments. Considering Exon only alignments, activated dendritic cells were found to be elevated in IBC ($p=0.011$; for Exon + Intron analysis, $p=0.061$) and significant evidence ($p=0.010$) of reduction in resting mast cells, another class of granulocytes, in IBC PBMCs relative to the combined non-IBC patient and healthy donor PBMC sample sets.

### Figure S11

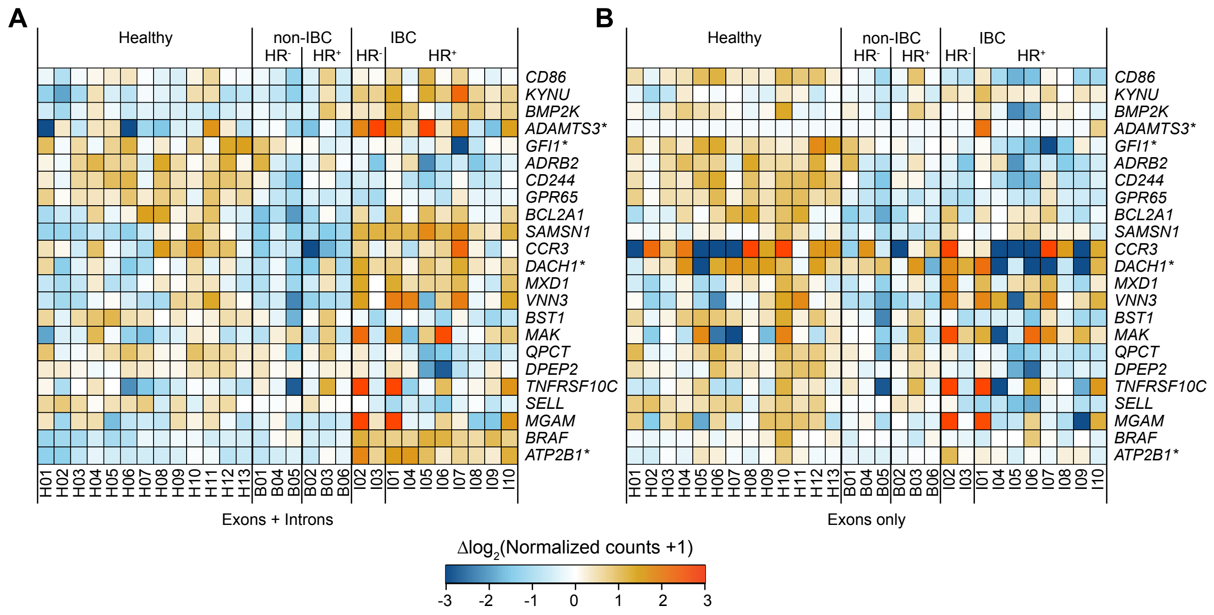
A
Healthy
non-IBC
IBC
HR-
HR+
CD86
KYNU
BMP2K
ADAMTS3*
GFI1*
ADRB2
CD244
GPR65
BCL2A1
SAMSN1
CCR3
DACH1*
MXD1
VNN3
BST1
MAK
QPCT
DPEP2
TNFRSF10C
SELL
MGAM
BRAF
ATP2B1*
H01 H02 H03 H04 H05 H06 H07 H08 H09 H10 H11 H12 H13 B01 B04 B05 B02 B03 B06 I02 I03 I01 I04 I05 I06 I07 I08 I09 I10
Exons + Introns
B
Exons only
Δlog2(Normalized counts +1)
-3 -2 -1 0 1 2 3

**Fig. S11. Heat map of the relative abundance of CIBERSORTx-related genes in PBMC samples.** Relevant CIBERSORTx marker panel genes (those most highly expressed in the CIBERSORTx training set data for activated dendritic cells, eosinophils, neutrophils, or resting mast cells, as well as *BRAF*) and one other gene (*ATP2B1**) potentially relevant to the IBC PBMC response that were differentially expressed (adj. $p \leq 0.001$, $|LFC| \geq 1$) in IBC patient PBMCs compared to either non-IBC patient or healthy donors PBMCs in Exon + Intron alignments are shown. Tiles are colored by expression differences relative to the corresponding genewise means taken across all PBMC samples for (**A**) Exon + Intron alignments and (**B**) Exon only alignments. Lanes in heat maps are numbered according to the healthy donor or patient ID number (Table S1).

### Figure S12

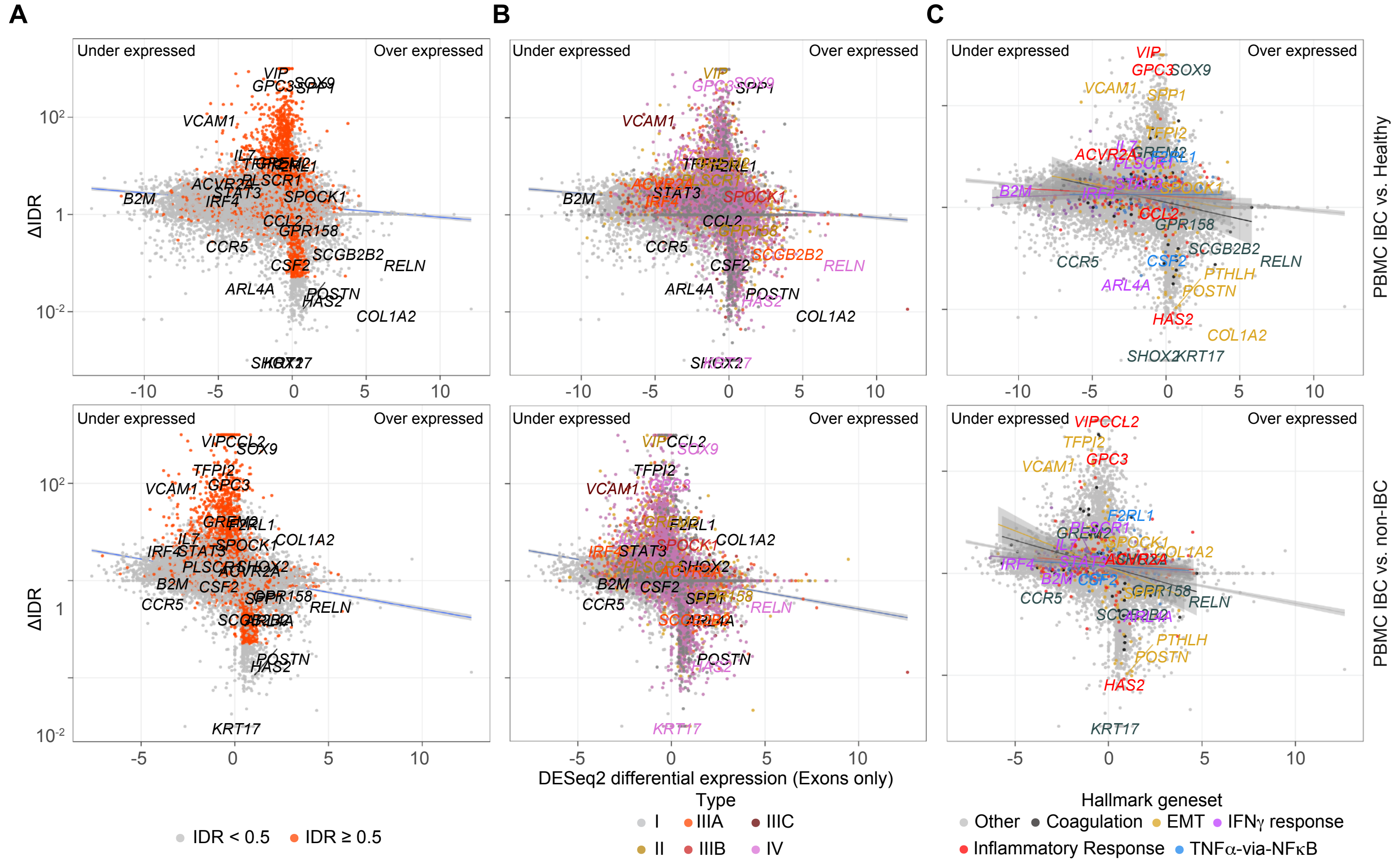

A
B
C
Under expressed
Over expressed
ΔIDR
DESeq2 differential expression (Exons only)
PBMC IBC vs. Healthy
PBMC IBC vs. non-IBC
IDR < 0.5
IDR ≥ 0.5
Type
I
II
IIIA
IIIB
IIIC
IV
Hallmark geneset
Other
Coagulation
EMT
IFNγ response
Inflammatory Response
TNFα-via-NFκB

**Fig. S12. High ΔIDR genes sequenced in IBC versus non-IBC PBMC samples trend toward lower differential expression for reads mapped to the transcriptome reference. (A-C)** ΔIDR (*y*-axis) versus differential expression enrichment scores calculated by DESeq2 for reads mapped to the transcriptome reference sequence (*x*-axis) with genes (and, when applicable, their names) color coded as shown beneath the panels based on (**A**) IDR in IBC PBMC samples; (**B**) Gene type based on the ratio of intron to exon read depth across the gene as defined in Fig.2C; (**C**) Membership in Broad Institute Hallmark gene sets (with separate trendlines shown for each gene set to highlight their varying patterns of differential expression and ΔIDR). Select genes which were either: (i) differentially expressed; (ii) have particularly high or low ΔIDR values; or (iii) represent one of the different intronic expression patterns are named in panels (A-C).

#### Figure S13

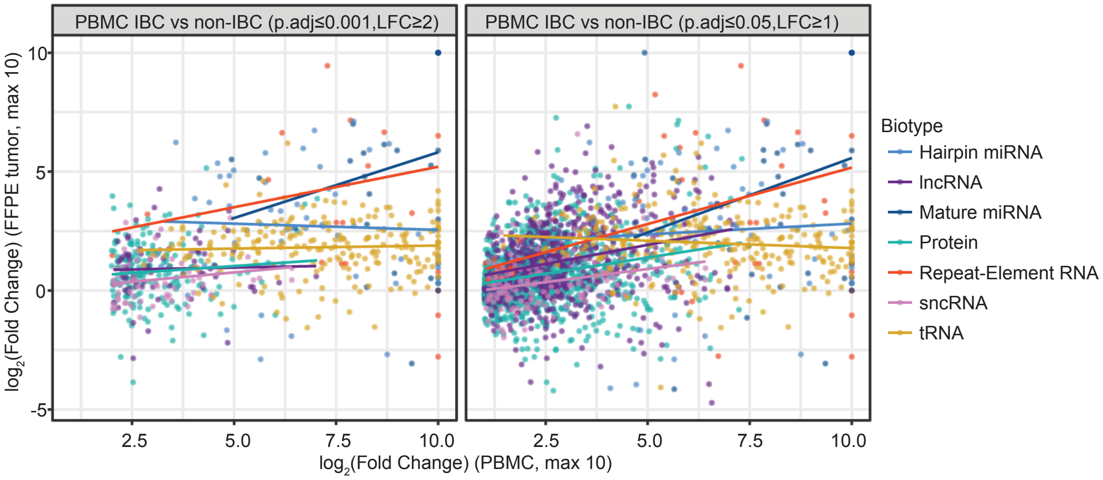

PBMC IBC vs non-IBC (p.adj≤0.001,LFC≥2)
PBMC IBC vs non-IBC (p.adj≤0.05,LFC≥1)
log2(Fold Change) (FFPE tumor, max 10)
10
5
0
-5
2.5
5.0
7.5
10.0
2.5
5.0
7.5
10.0
log2(Fold Change) (PBMC, max 10)
Biotype
Hairpin miRNA
lncRNA
Mature miRNA
Protein
Repeat-Element RNA
sncRNA
tRNA

**Fig. S13. Scatter plots comparing IBC vs non-IBC DE genes LFCs in FFPE tumors and PBMCs.** Each point represents an identified over-expressed gene in IBC vs non-IBC patient PBMCs with adj. $p \leq 0.001$, LFC>2 (left panel) or adj. $p \leq 0.05$, LFC>1 (right panel). The *y*-axis encodes DESeq2 LFC for the FFPE tumor IBC vs non-IBC comparison while the *x*-axis encodes the LFC for PBMC IBC vs non-IBC comparison. Colors indicate different gene biotype indicated to the right of the plots. Trendlines show the linear model fit summarizing relationship between two comparisons for all plotted genes, fit separately for each indicated gene biotype.

#### Figure S14

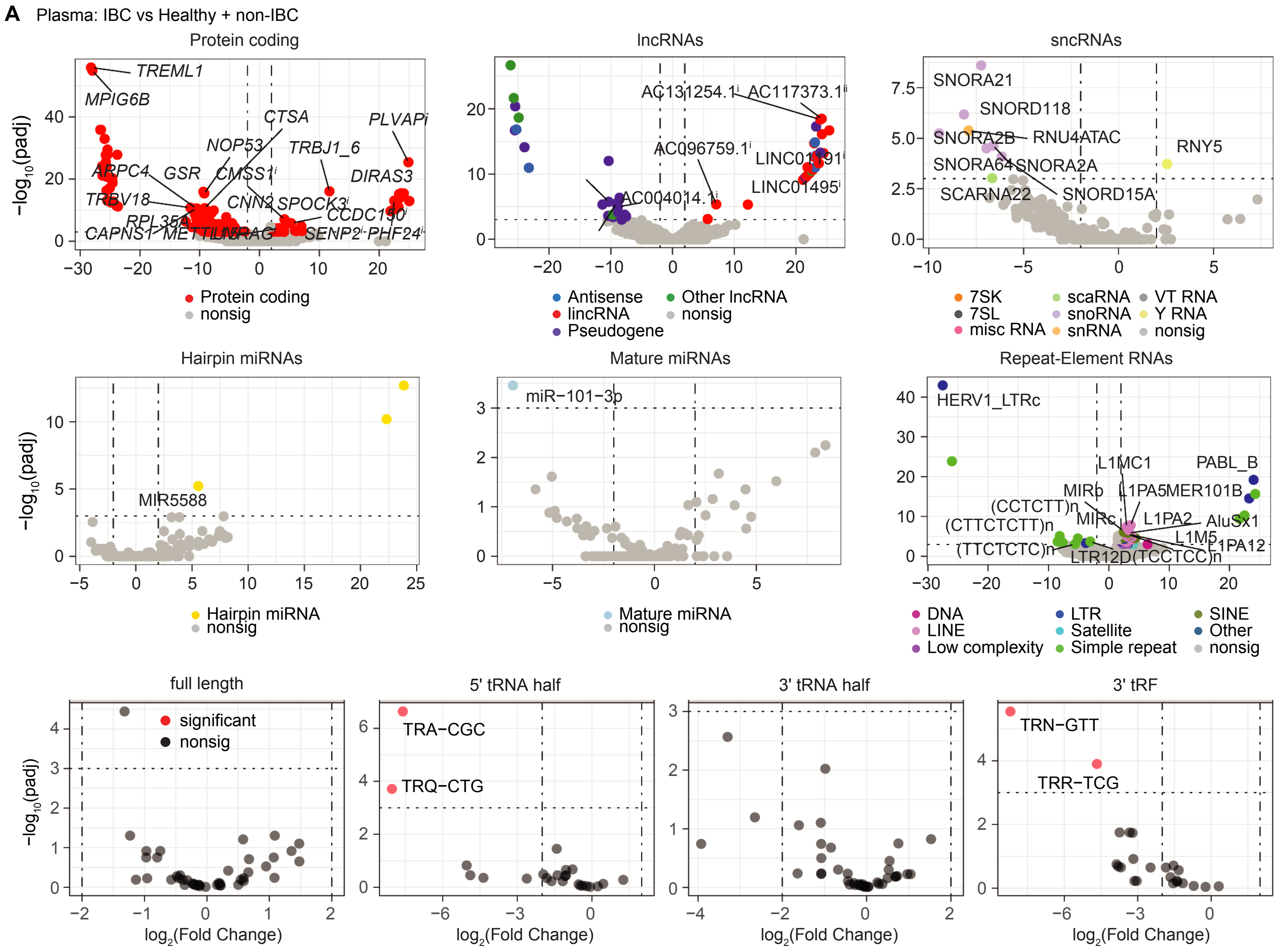
A
Plasma: IBC vs Healthy + non-IBC
Protein coding
lncRNAs
sncRNAs
Hairpin miRNAs
Mature miRNAs
Repeat-Element RNAs
full length
5' tRNA half
3' tRNA half
3' tRF
−log10(padj)
log2(Fold Change)
Protein coding
nonsig
Antisense
Other lncRNA
lincRNA
Pseudogene
7SK
scaRNA
VT RNA
7SL
snoRNA
Y RNA
misc RNA
snRNA
Hairpin miRNA
Mature miRNA
DNA
LTR
SINE
LINE
Satellite
Other
Low complexity
Simple repeat
significant
TREML1
MPIG6B
CTSA
PLVAPi
NOP53
TRBJ1_6
DIRAS3
HERV1_LTRc
PABL_B
SNORA21
SNORD118
RNY5
miR−101−3p
MIR5588
TRA−CGC
TRQ−CTG
TRN−GTT
TRR−TCG

#### Figure S14

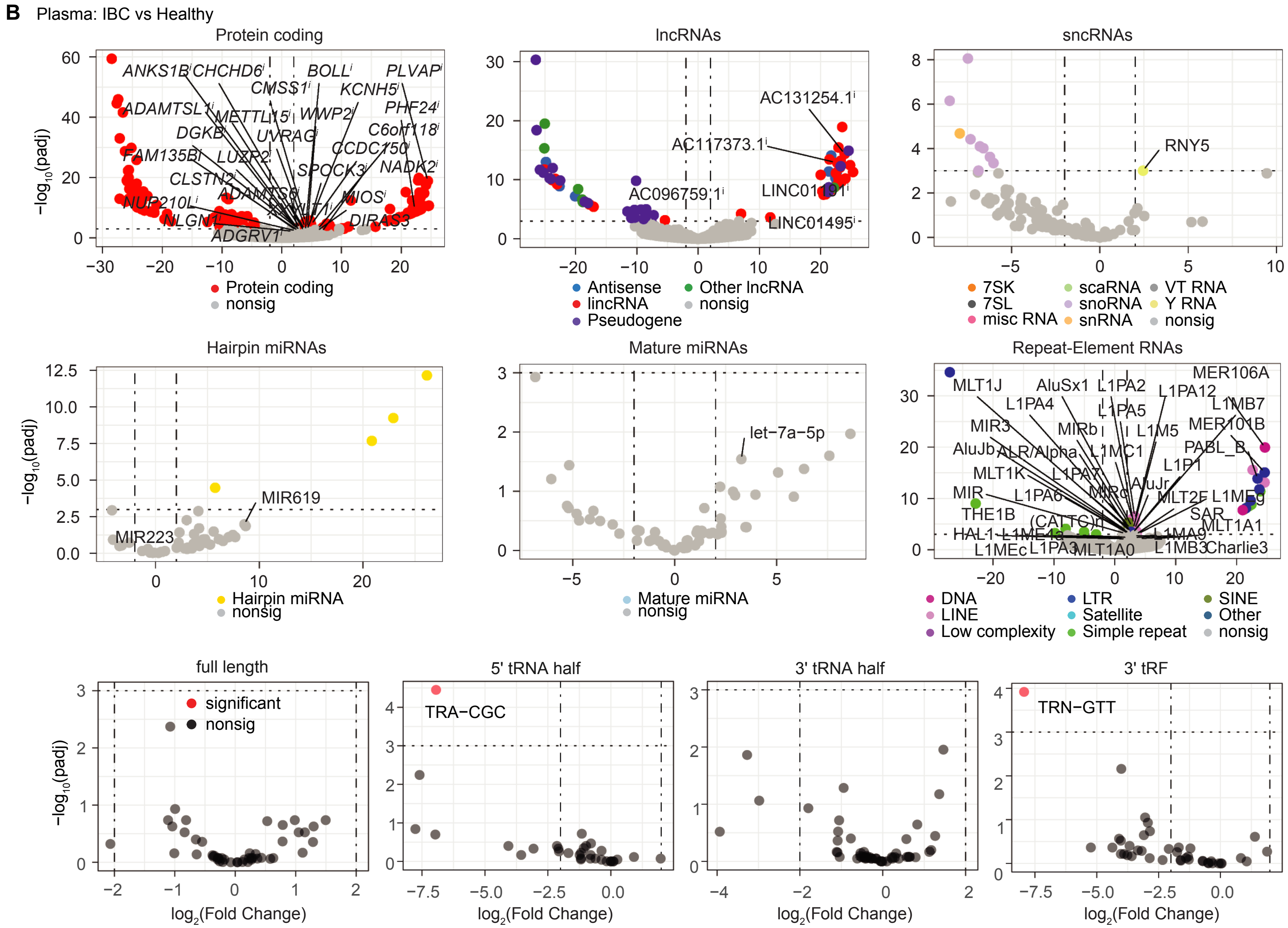
B
Plasma: IBC vs Healthy
Protein coding
lncRNAs
sncRNAs
Hairpin miRNAs
Mature miRNAs
Repeat-Element RNAs
full length
5' tRNA half
3' tRNA half
3' tRF
−log10(padj)
log2(Fold Change)
Protein coding
nonsig
Antisense
Other lncRNA
lincRNA
Pseudogene
7SK
7SL
misc RNA
scaRNA
snoRNA
snRNA
VT RNA
Y RNA
Hairpin miRNA
Mature miRNA
DNA
LINE
Low complexity
LTR
Satellite
Simple repeat
SINE
Other
significant
RNY5
MIR619
MIR223
let−7a−5p
AC131254.1
AC117373.1
AC096759.1
LINC01495
TRA−CGC
TRN−GTT

### Figure S14

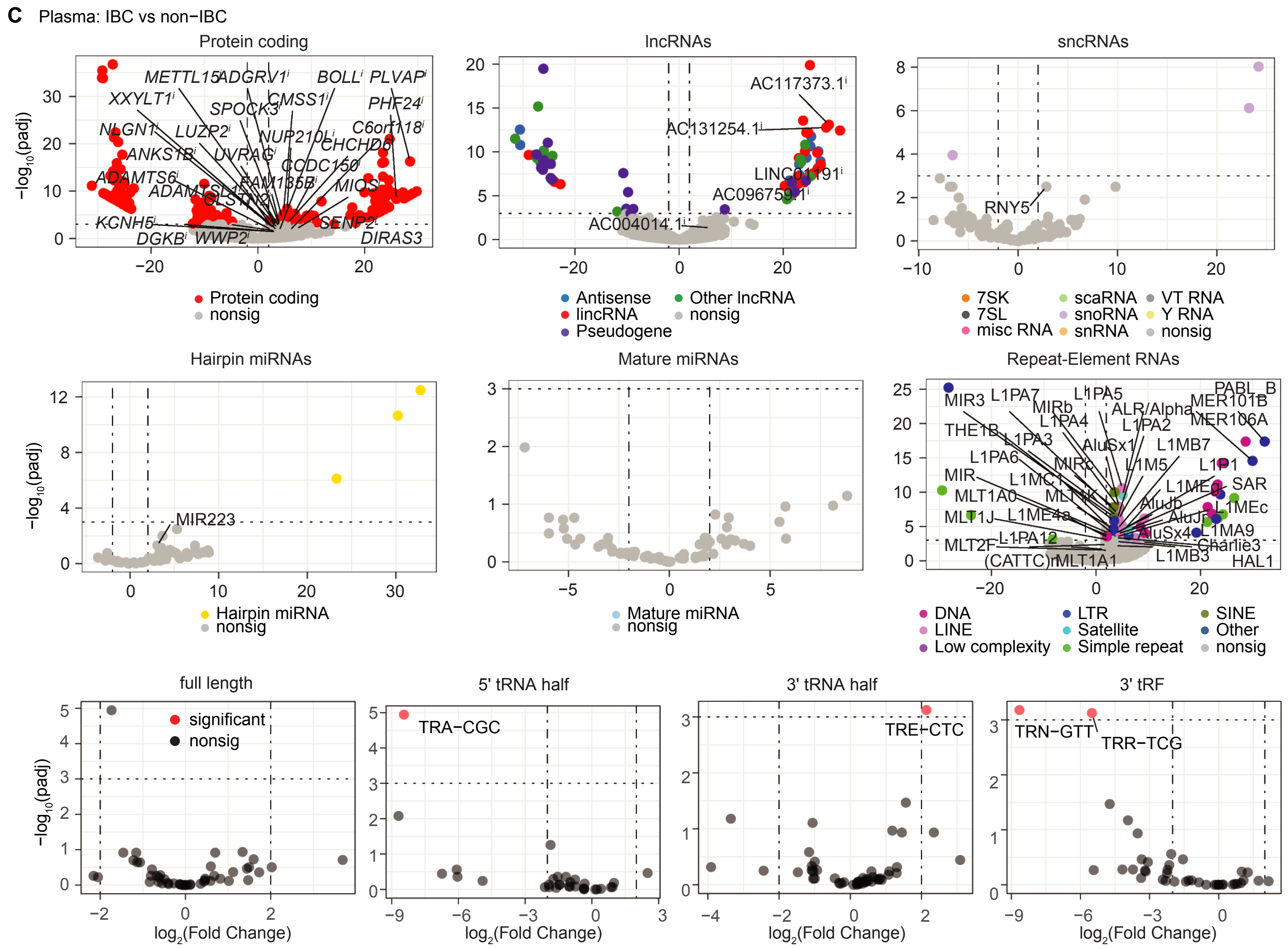

C
Plasma: IBC vs non−IBC
Protein coding
lncRNAs
sncRNAs
Hairpin miRNAs
Mature miRNAs
Repeat-Element RNAs
full length
5' tRNA half
3' tRNA half
3' tRF
−log10(padj)
log2(Fold Change)
Protein coding
nonsig
Antisense
lincRNA
Pseudogene
Other lncRNA
7SK
7SL
misc RNA
scaRNA
snoRNA
snRNA
VT RNA
Y RNA
Hairpin miRNA
Mature miRNA
DNA
LINE
Low complexity
LTR
Satellite
Simple repeat
SINE
Other
significant
AC117373.1
AC131254.1
LINC01191
AC096759.1
AC004014.1
RNY5
MIR223
TRA−CGC
TRE−CTC
TRN−GTT
TRR−TCG

**Fig. S14. Volcano plots for DESeq2 two-group comparisons in plasma samples.** The *y*-axis indicates $\log_{10}$-transformed adj. *p*, while the *x*-axis indicates LFC. Each point represents one gene with biotypes color coded for those genes with adj. $p \leq 0.001$ and $|LFC|>2$. Significantly over-represented and other notable genes that passed filtering steps and were detected in at least 50% of the samples of the group in which they were over-represented are labeled by their gene symbol. i: protein-coding gene contains reads that are mostly derived from intronic regions. nonsig: genes that are not significantly differentially expressed.

### Figure S15

#### A. Protein-coding genes

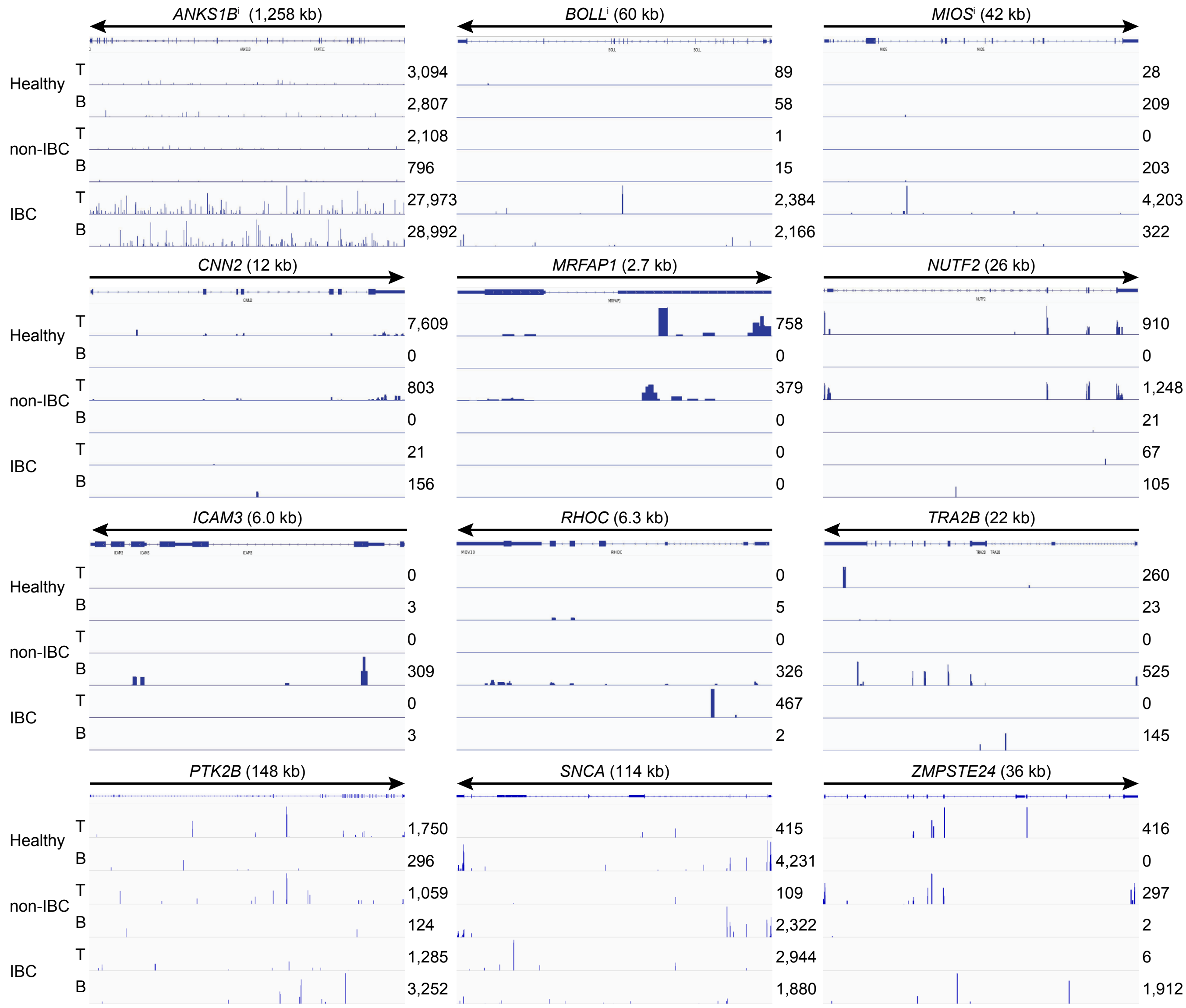

### Figure S15

#### B. lncRNAs

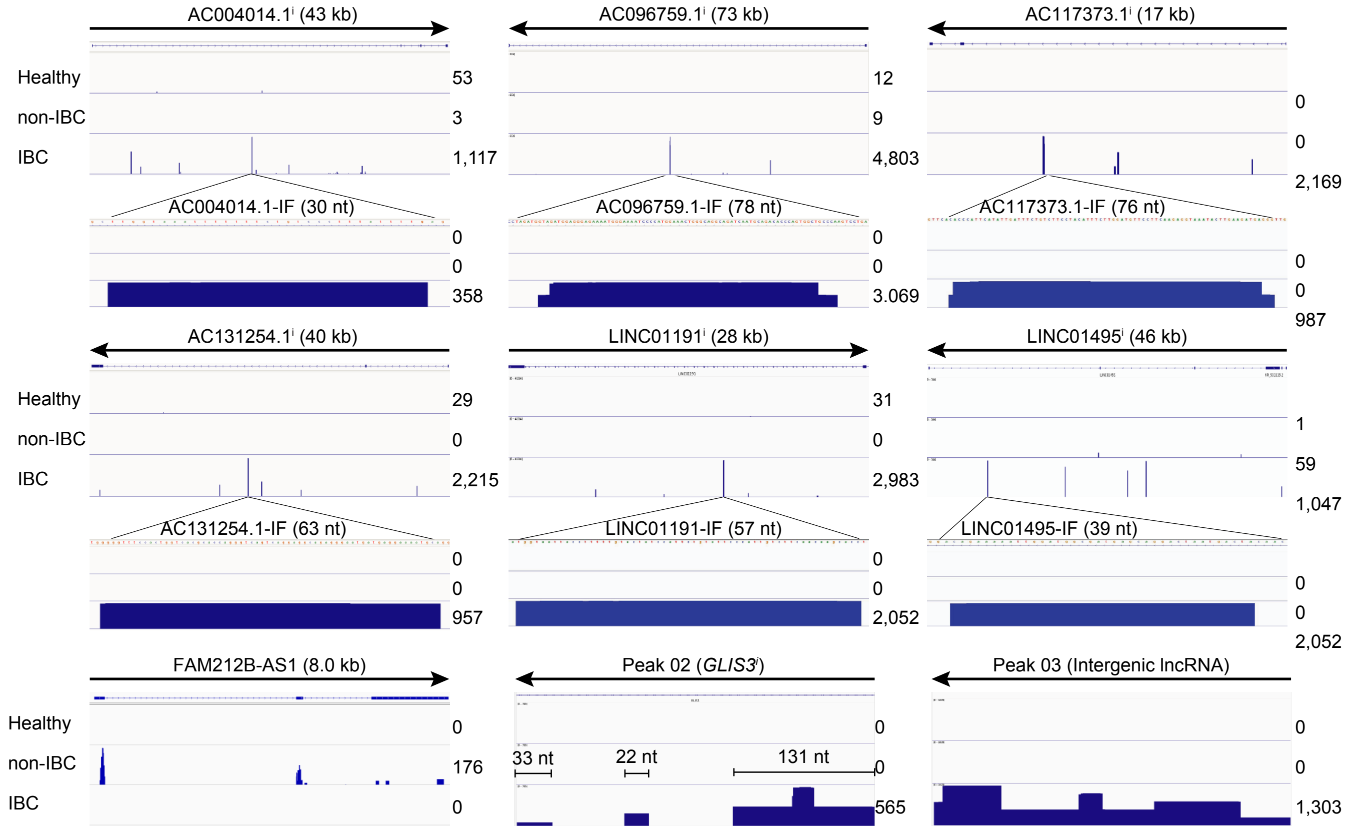


#### C. sncRNAs

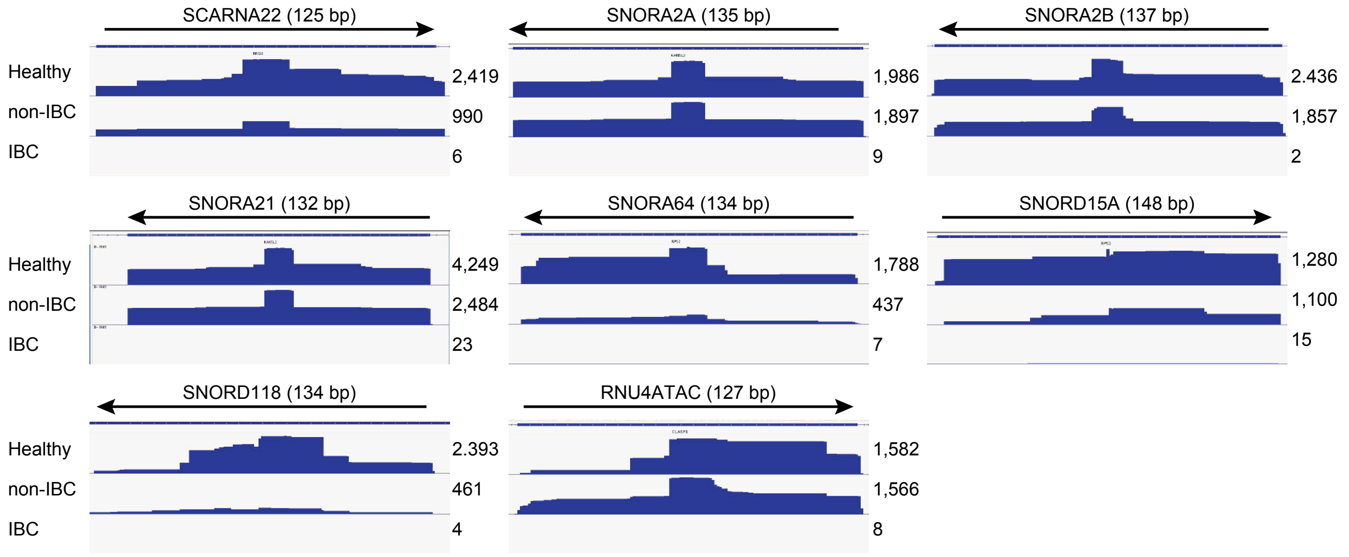

### Figure S15

#### D. miRNAs

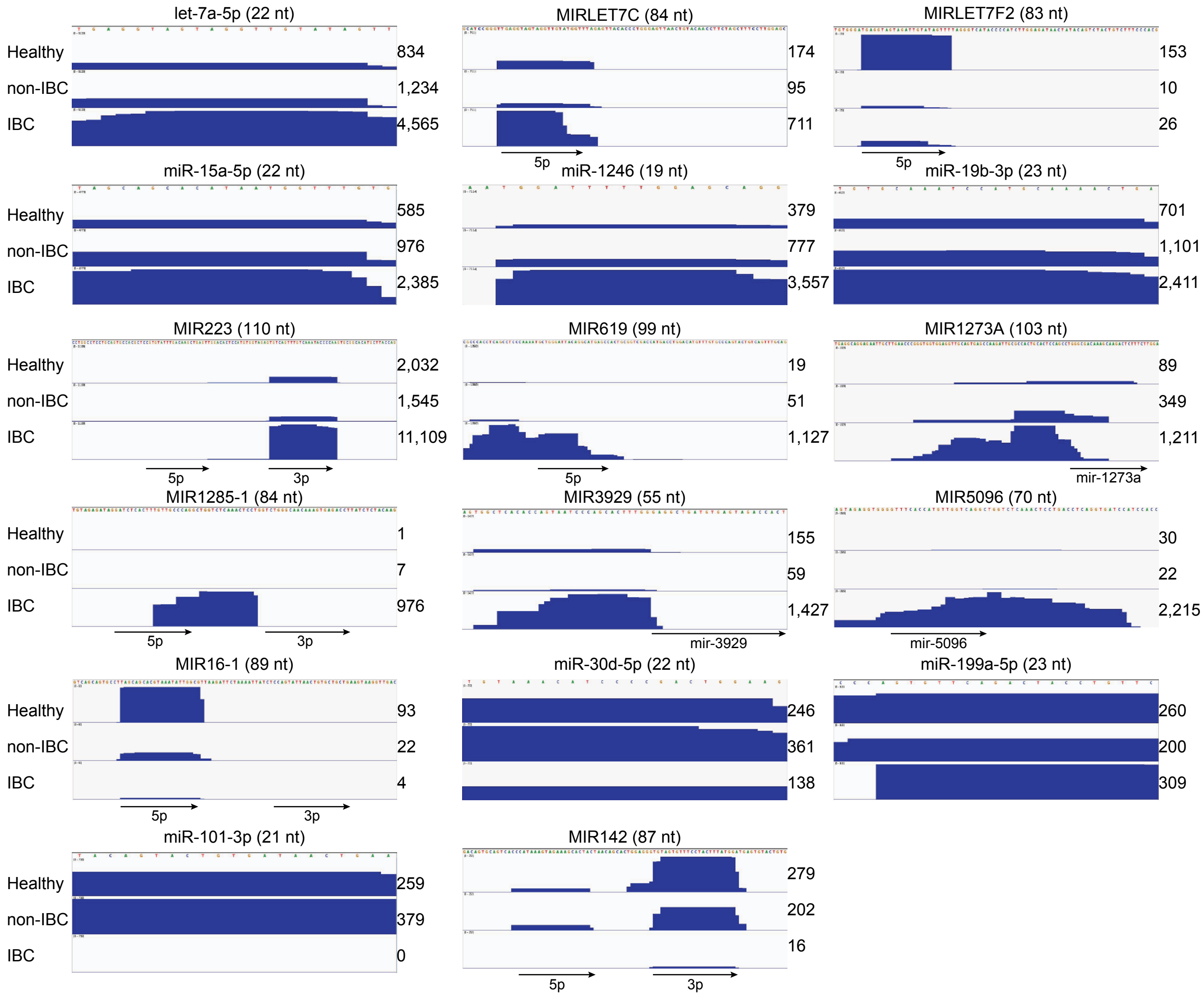

**Fig. S15. IGV plots of differentially expressed or notable genes in plasma samples.** (**A**) protein-coding genes, (**B**) lncRNAs, (**C**) sncRNAs, and (**D**) miRNAs. T and B, top and bottom DNA strands, respectively. The number of reads is indicated to the right. i: protein-coding gene contains reads that are mostly derived from intronic regions.

### Figure S16

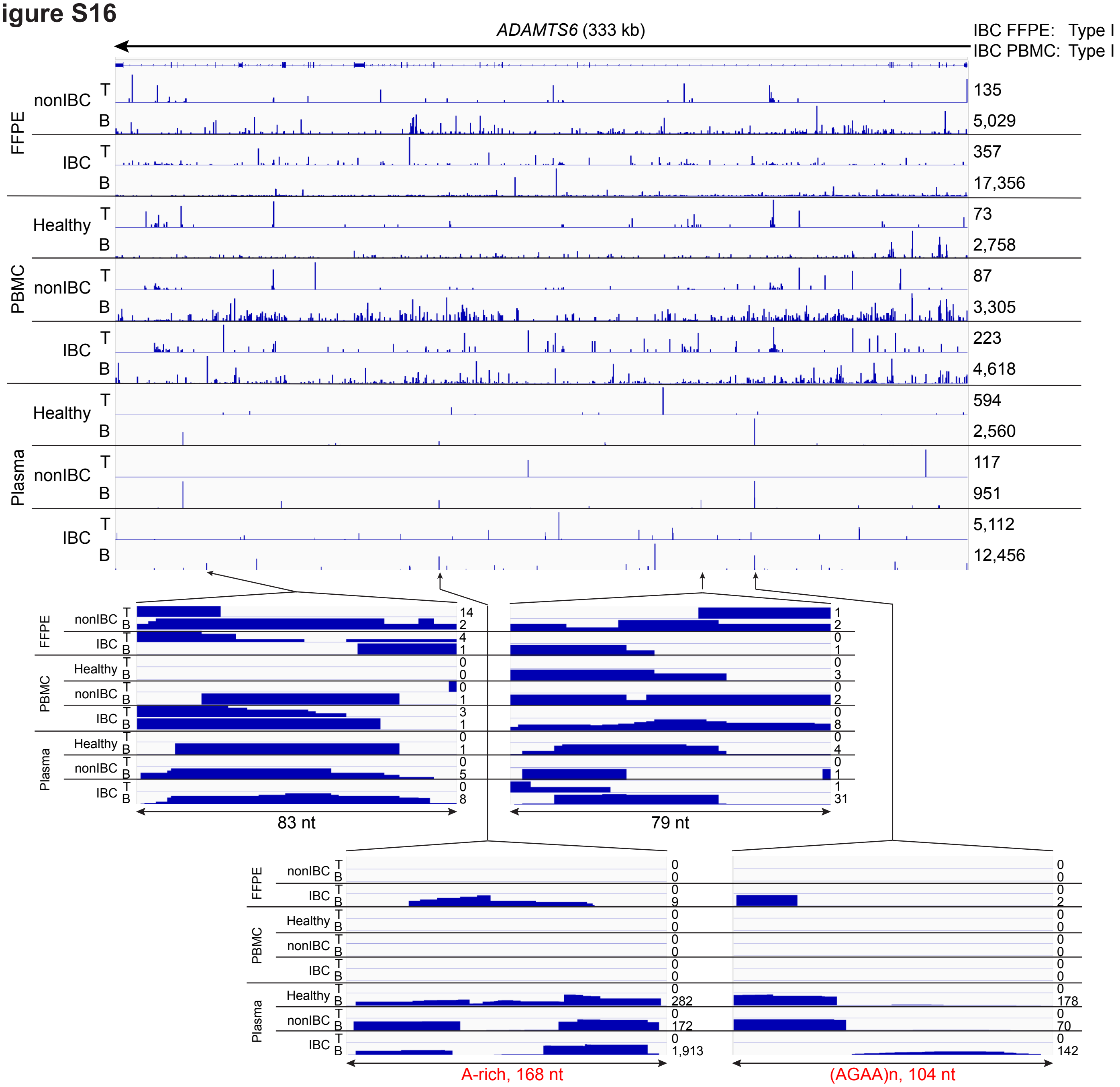

ADAMTS6 (333 kb)
IBC FFPE: Type I
IBC PBMC: Type I
FFPE
nonIBC
IBC
PBMC
Healthy
nonIBC
IBC
Plasma
Healthy
nonIBC
IBC
T
B
135
5,029
357
17,356
73
2,758
87
3,305
223
4,618
594
2,560
117
951
5,112
12,456
83 nt
79 nt
A-rich, 168 nt
(AGAA)n, 104 nt

#### Figure S16

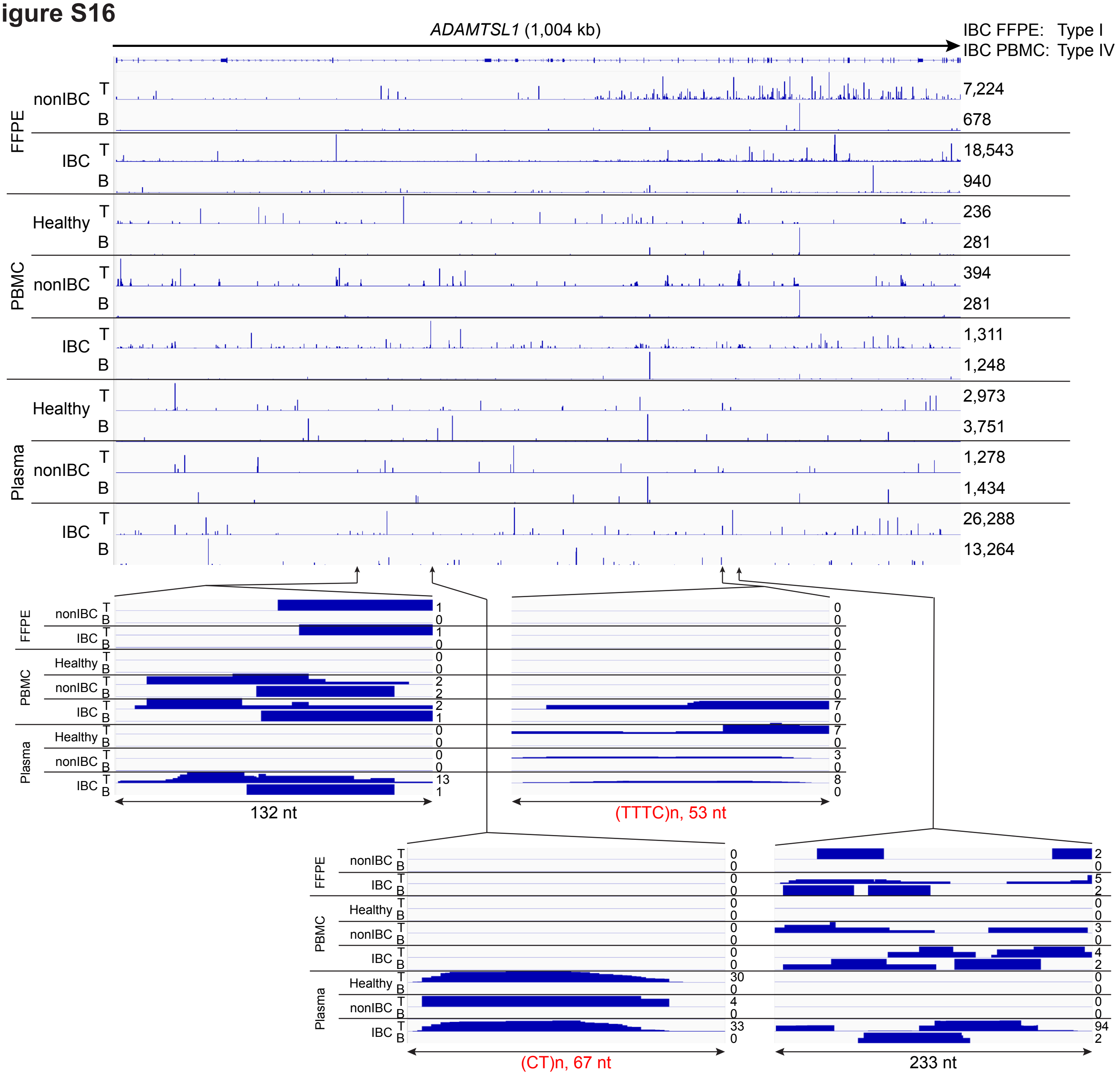
ADAMTSL1 (1,004 kb)
IBC FFPE: Type I
IBC PBMC: Type IV
FFPE
PBMC
Plasma
nonIBC
IBC
Healthy
T
B
7,224
678
18,543
940
236
281
394
281
1,311
1,248
2,973
3,751
1,278
1,434
26,288
13,264
132 nt
(TTTC)n, 53 nt
(CT)n, 67 nt
233 nt

### Figure S16

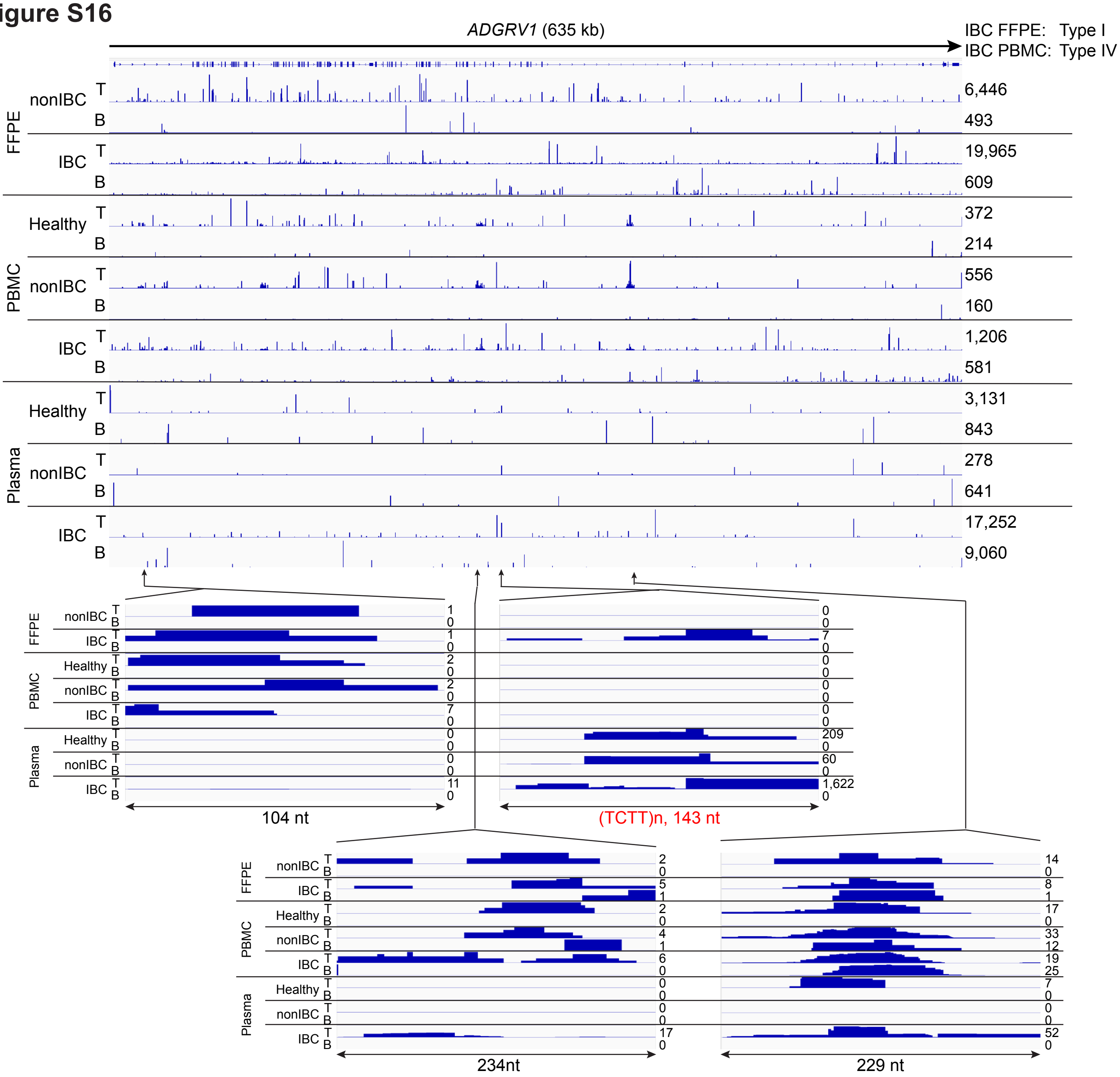

ADGRV1 (635 kb)
IBC FFPE: Type I
IBC PBMC: Type IV
FFPE
nonIBC
IBC
PBMC
Healthy
nonIBC
IBC
Plasma
Healthy
nonIBC
IBC
T
B
6,446
493
19,965
609
372
214
556
160
1,206
581
3,131
843
278
641
17,252
9,060
104 nt
(TCTT)n, 143 nt
234nt
229 nt

**Figure S16**

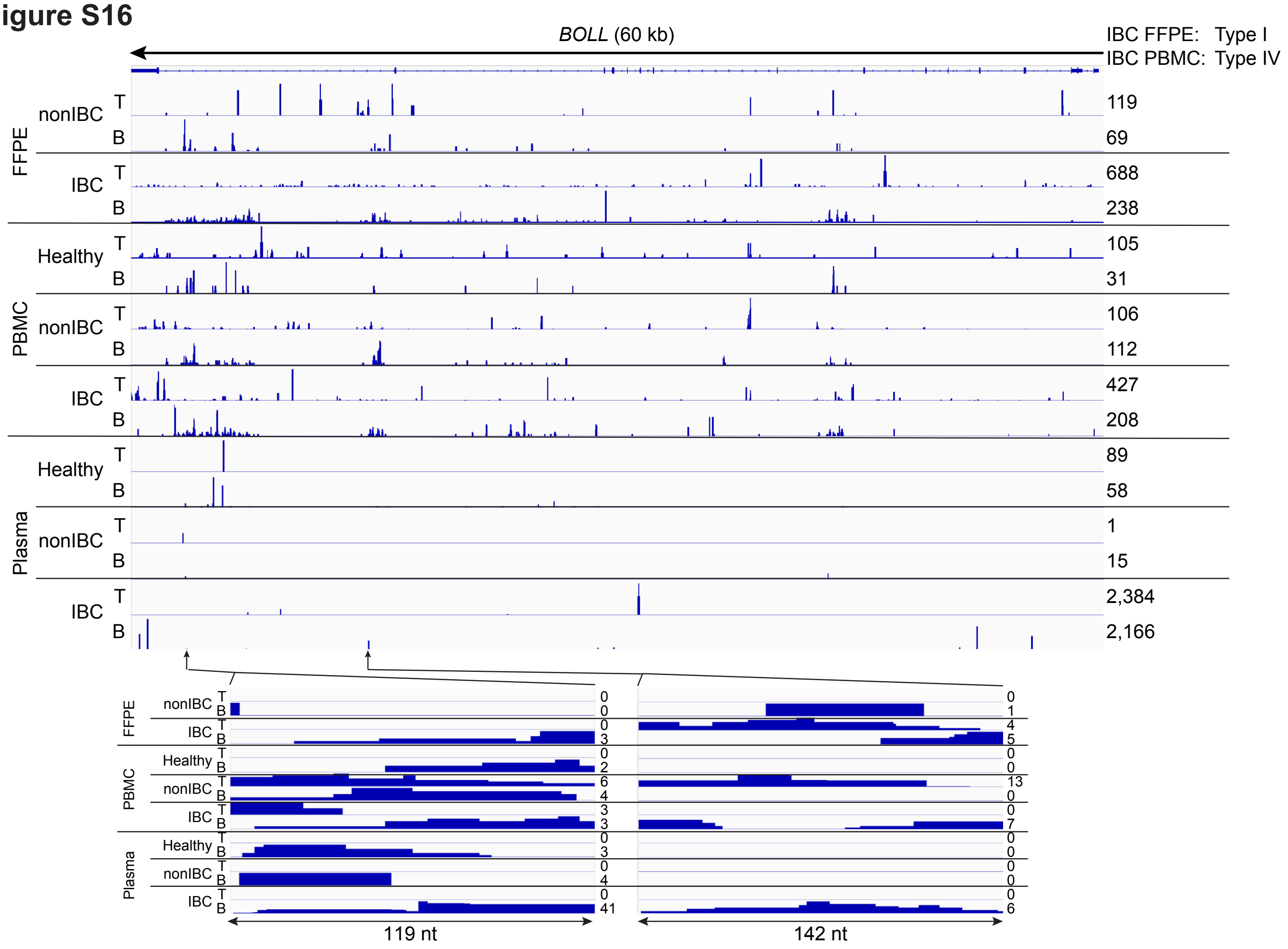
BOLL (60 kb)
IBC FFPE: Type I
IBC PBMC: Type IV
FFPE
nonIBC
IBC
PBMC
Healthy
nonIBC
IBC
Plasma
Healthy
nonIBC
IBC
T
B
119
69
688
238
105
31
106
112
427
208
89
58
1
15
2,384
2,166
119 nt
142 nt

### Figure S16

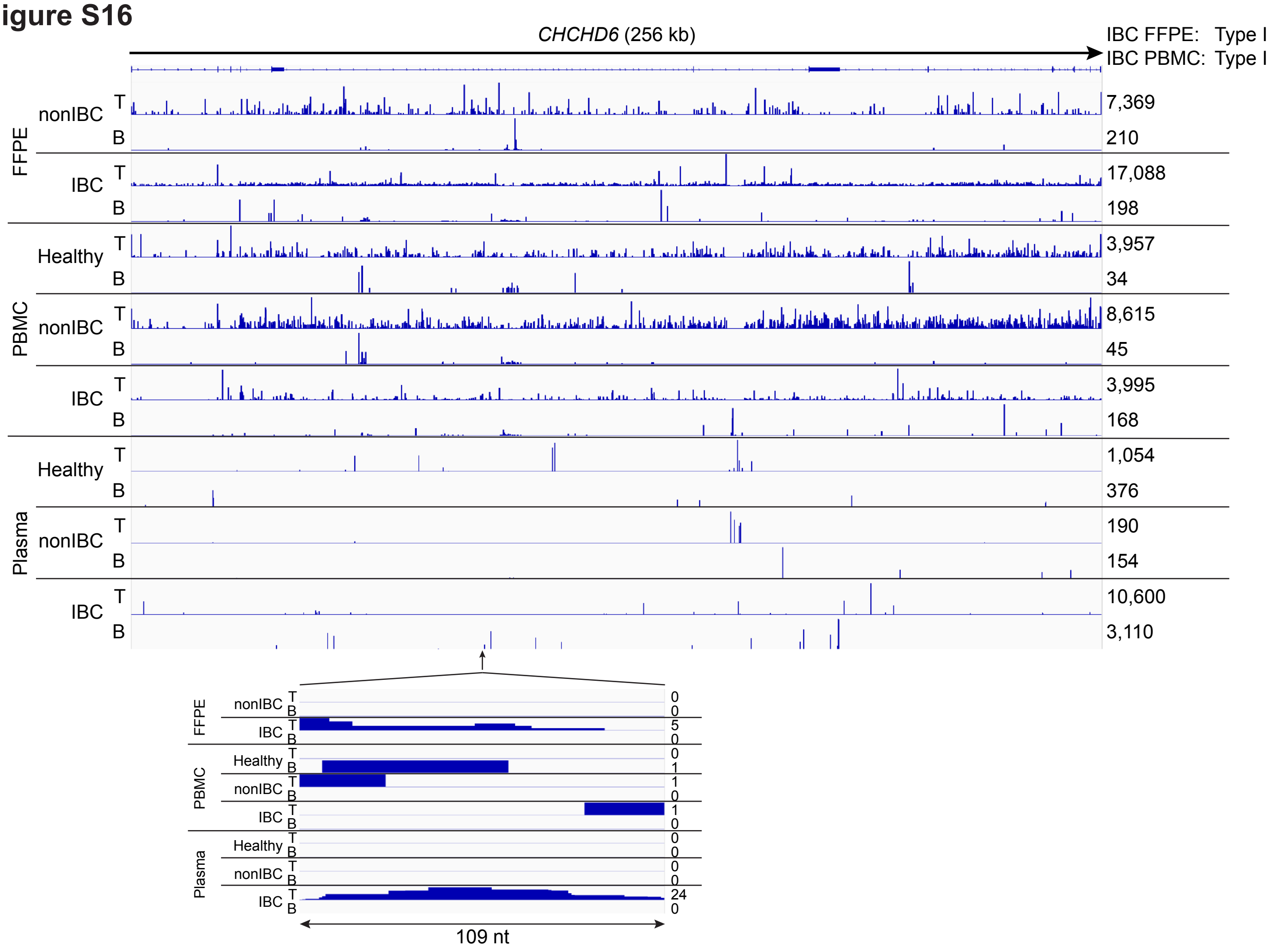
CHCHD6 (256 kb)
IBC FFPE: Type I
IBC PBMC: Type I
FFPE
nonIBC
IBC
PBMC
Healthy
nonIBC
IBC
Plasma
Healthy
nonIBC
IBC
T
B
7,369
210
17,088
198
3,957
34
8,615
45
3,995
168
1,054
376
190
154
10,600
3,110
109 nt

#### Figure S16

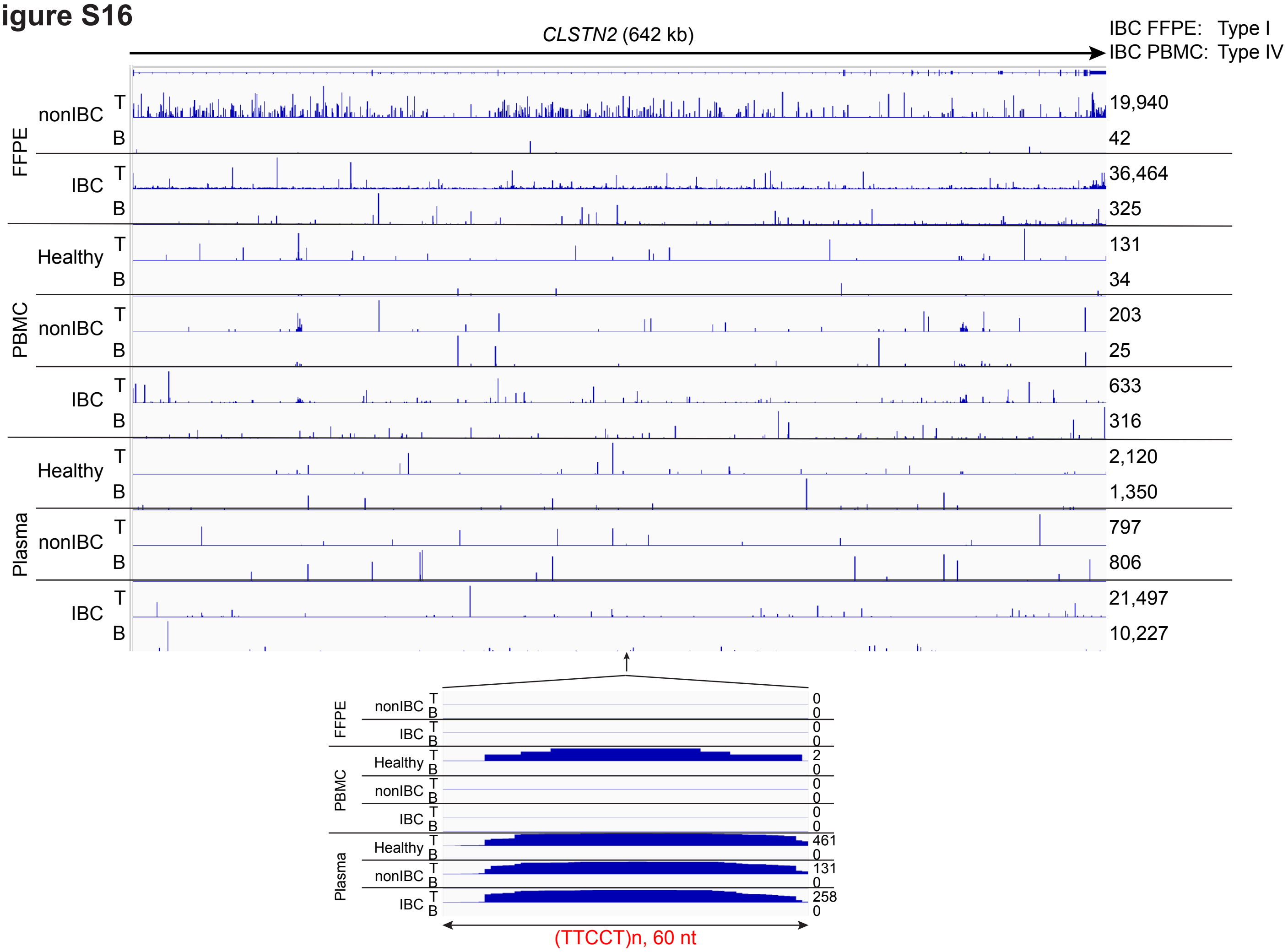

CLSTN2 (642 kb)
IBC FFPE: Type I
IBC PBMC: Type IV
FFPE
nonIBC
T
B
19,940
42
IBC
T
B
36,464
325
PBMC
Healthy
T
B
131
34
nonIBC
T
B
203
25
IBC
T
B
633
316
Plasma
Healthy
T
B
2,120
1,350
nonIBC
T
B
797
806
IBC
T
B
21,497
10,227
(TTCCT)n, 60 nt

#### Figure S16

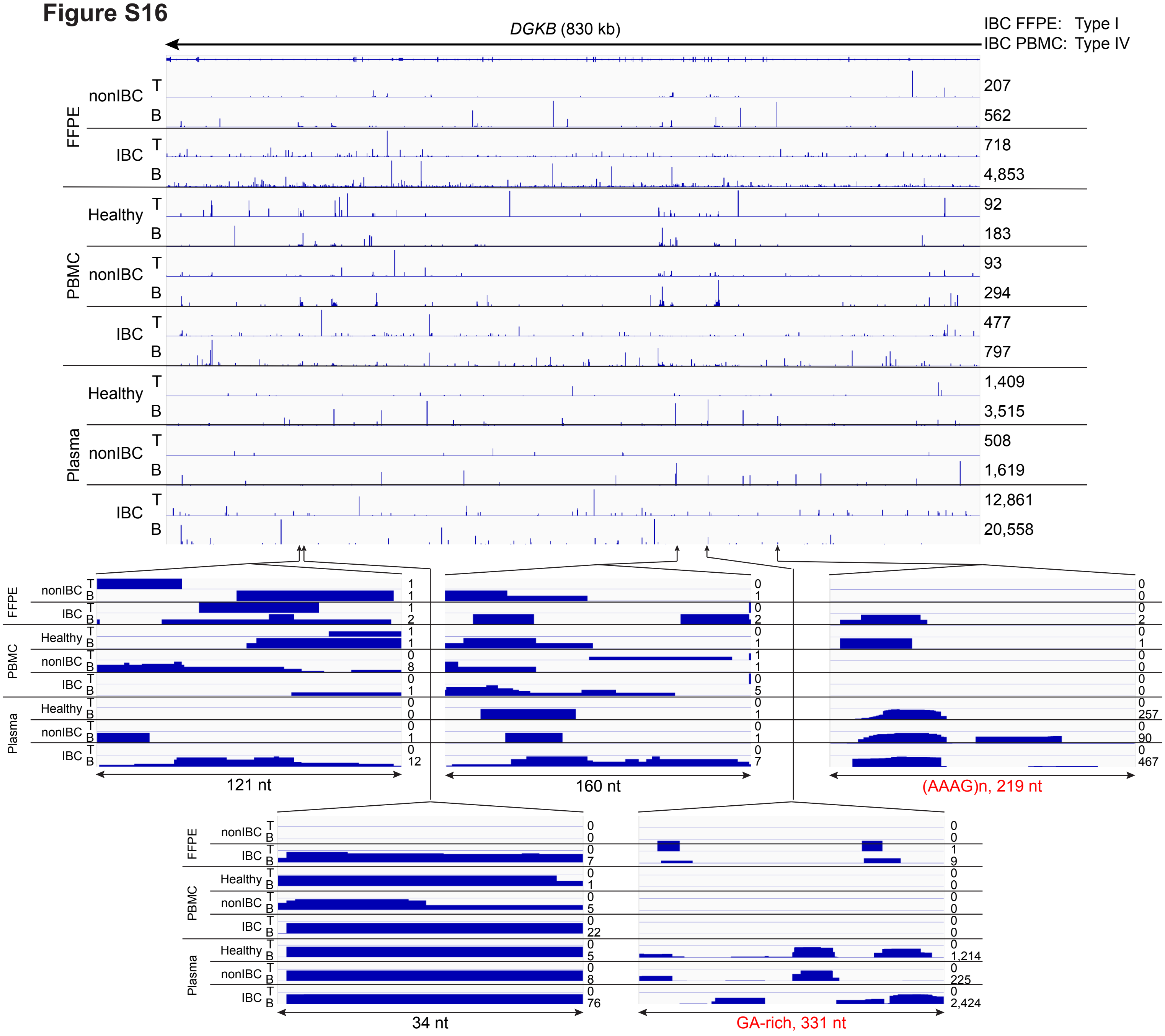

DGKB (830 kb)
IBC FFPE: Type I
IBC PBMC: Type IV
FFPE
PBMC
Plasma
nonIBC
IBC
Healthy
T
B
207
562
718
4,853
92
183
93
294
477
797
1,409
3,515
508
1,619
12,861
20,558
121 nt
160 nt
(AAAG)n, 219 nt
34 nt
GA-rich, 331 nt

### Figure S16

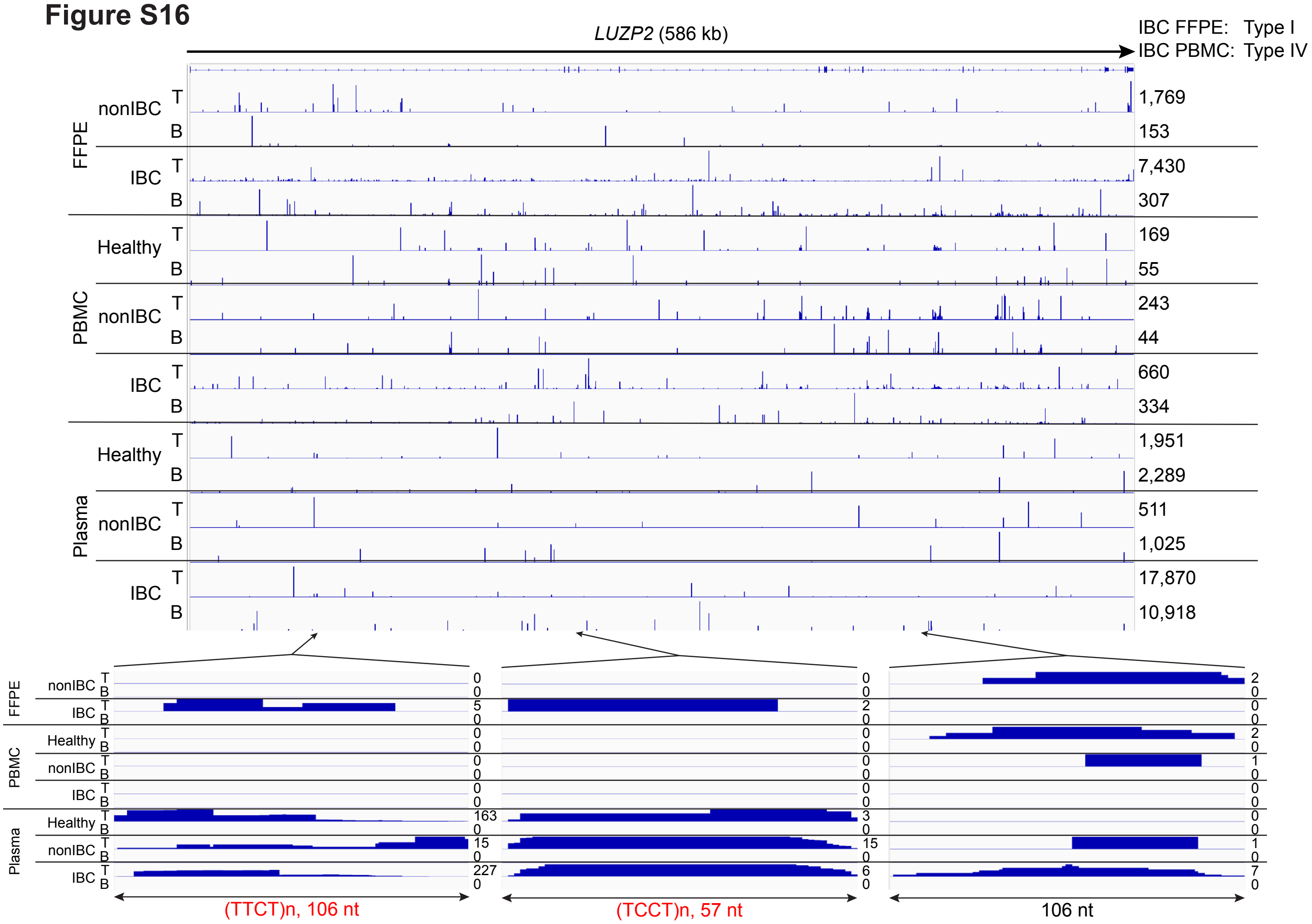
LUZP2 (586 kb)
IBC FFPE: Type I
IBC PBMC: Type IV
FFPE
PBMC
Plasma
nonIBC
IBC
Healthy
nonIBC
IBC
Healthy
nonIBC
IBC
T
B
1,769
153
7,430
307
169
55
243
44
660
334
1,951
2,289
511
1,025
17,870
10,918
(TTCT)n, 106 nt
(TCCT)n, 57 nt
106 nt

### Figure S16

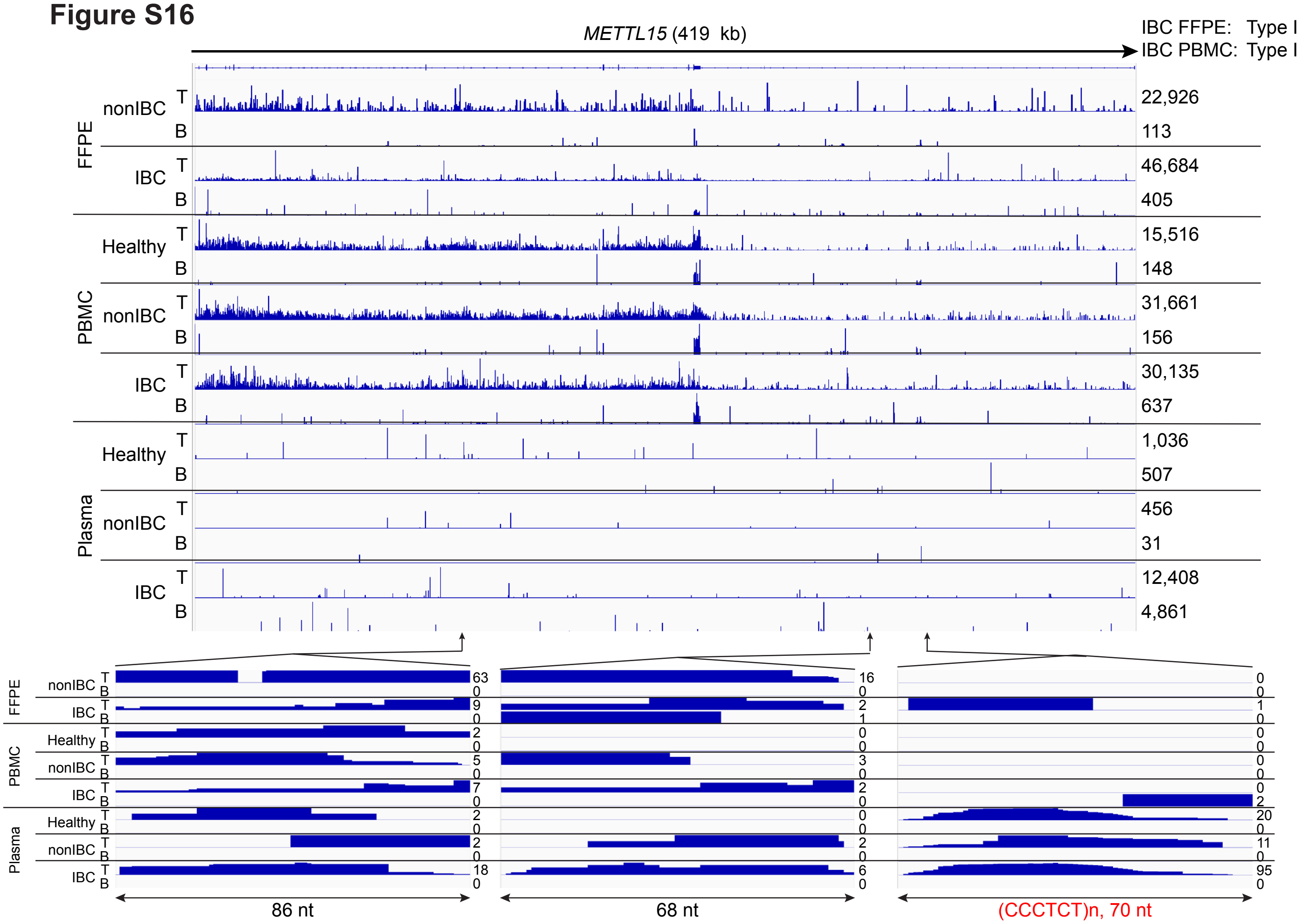

METTL15 (419 kb)
IBC FFPE: Type I
IBC PBMC: Type I
FFPE
nonIBC
IBC
PBMC
Healthy
nonIBC
IBC
Plasma
Healthy
nonIBC
IBC
T
B
22,926
113
46,684
405
15,516
148
31,661
156
30,135
637
1,036
507
456
31
12,408
4,861
86 nt
68 nt
(CCCTCT)n, 70 nt

### Figure S16

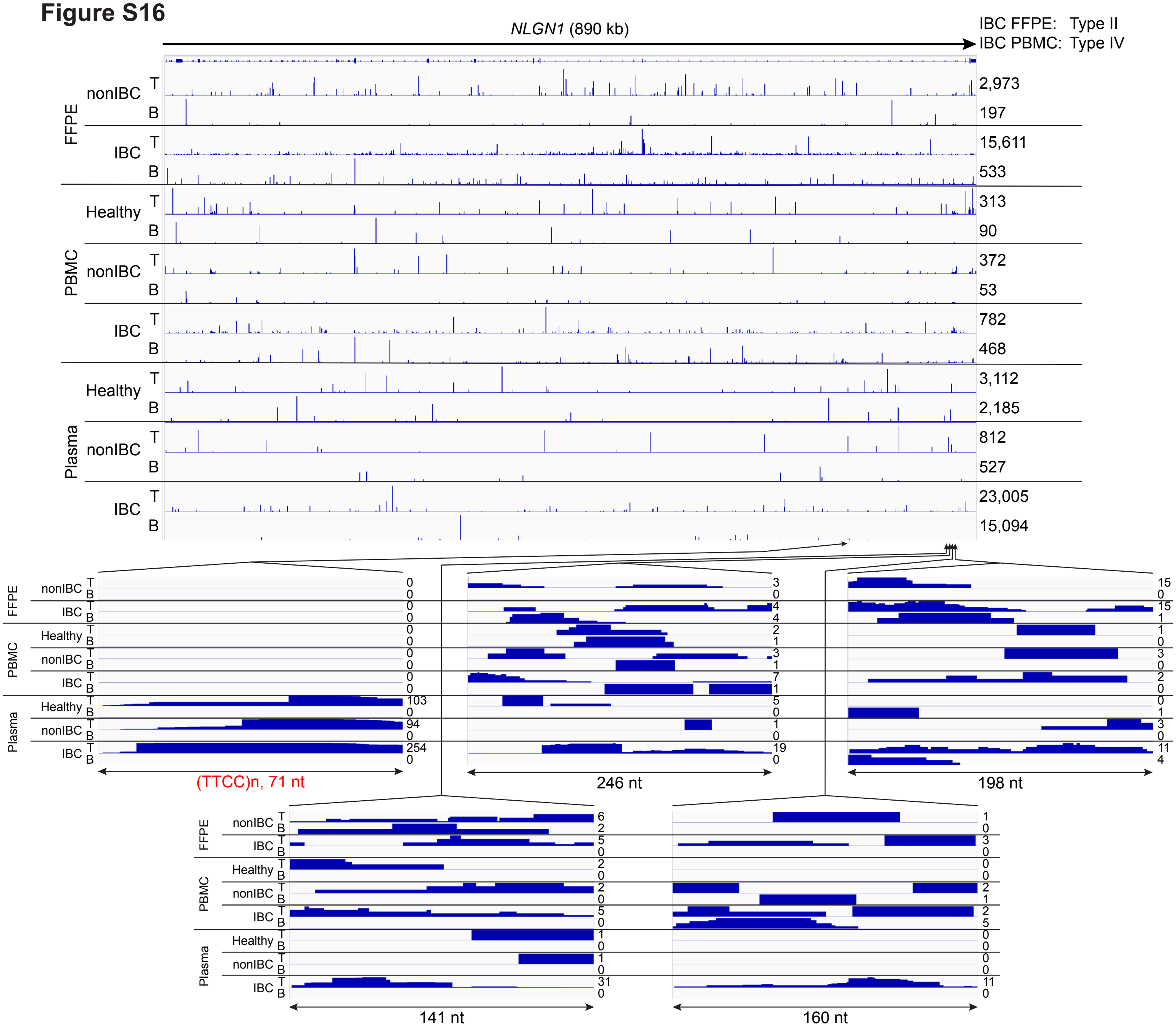

NLGN1 (890 kb)
IBC FFPE: Type II
IBC PBMC: Type IV
FFPE
nonIBC T B
IBC T B
PBMC
Healthy T B
nonIBC T B
IBC T B
Plasma
Healthy T B
nonIBC T B
IBC T B
2,973
197
15,611
533
313
90
372
53
782
468
3,112
2,185
812
527
23,005
15,094
(TTCC)n, 71 nt
246 nt
198 nt
141 nt
160 nt

### Figure S16

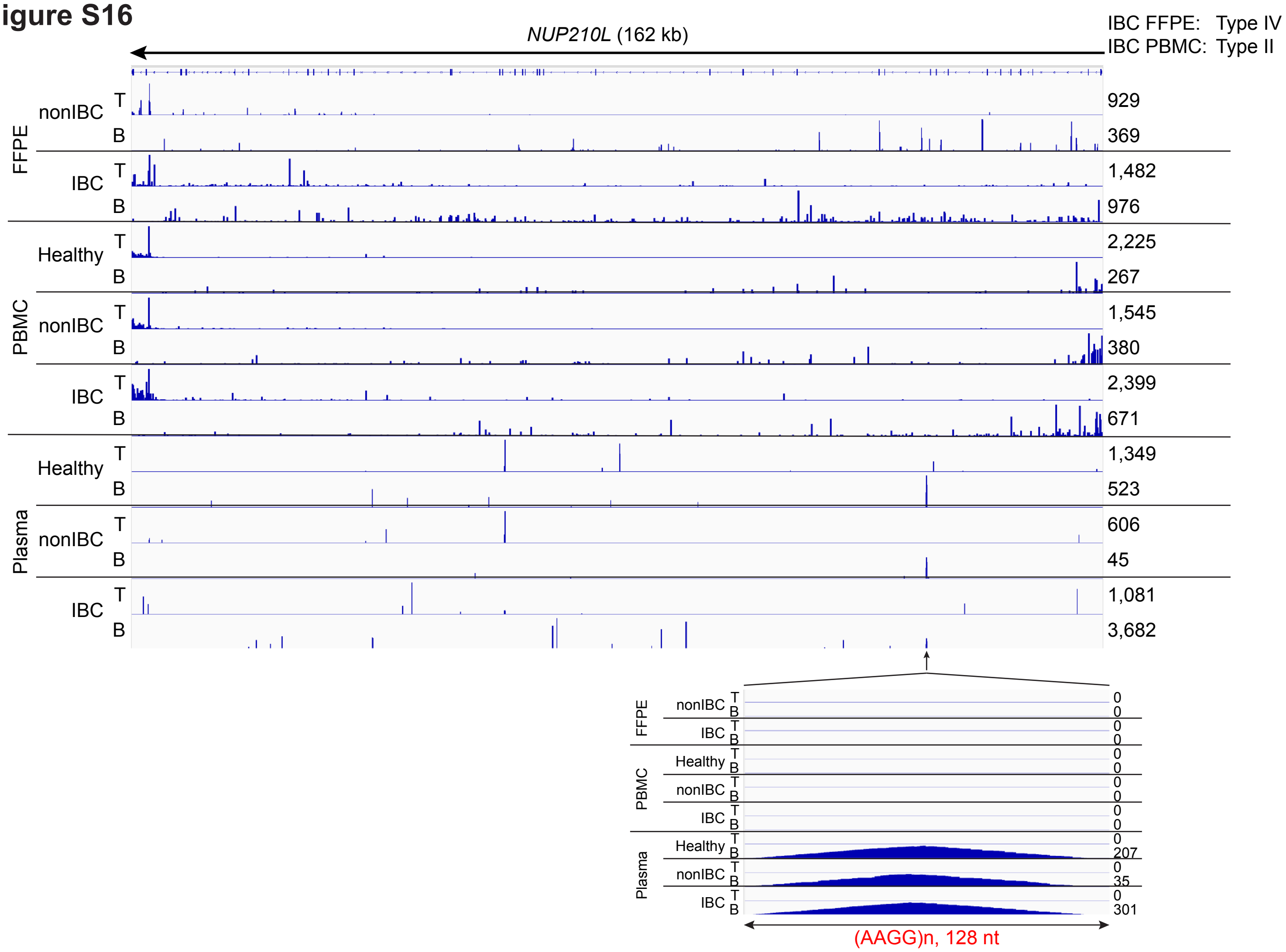

NUP210L (162 kb)
IBC FFPE: Type IV
IBC PBMC: Type II
FFPE
nonIBC
IBC
PBMC
Healthy
nonIBC
IBC
Plasma
Healthy
nonIBC
IBC
T
B
929
369
1,482
976
2,225
267
1,545
380
2,399
671
1,349
523
606
45
1,081
3,682
0
207
35
301
(AAGG)n, 128 nt

### Figure S16

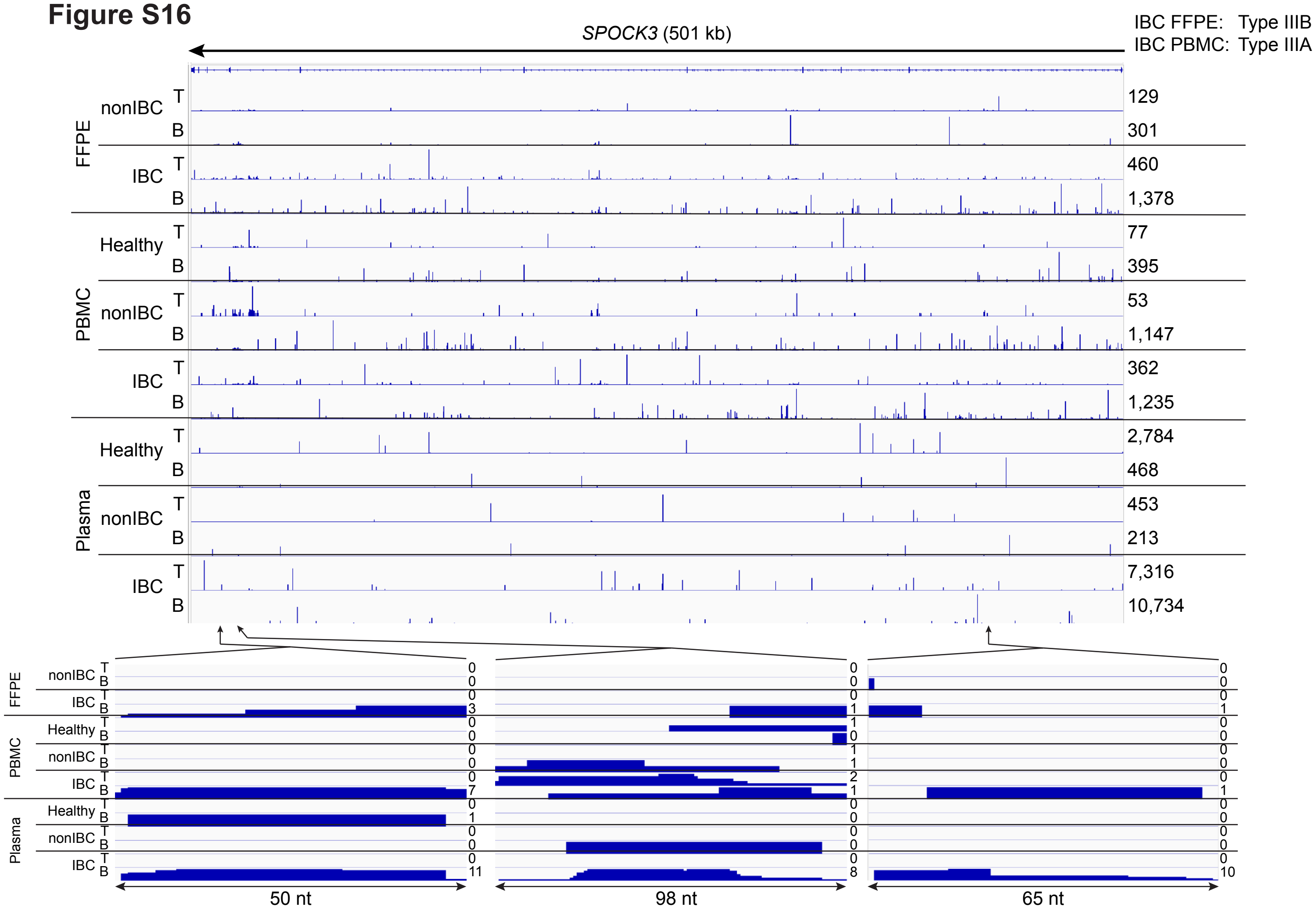

SPOCK3 (501 kb)
IBC FFPE: Type IIIB
IBC PBMC: Type IIIA
FFPE
PBMC
Plasma
nonIBC
IBC
Healthy
T
B
129
301
460
1,378
77
395
53
1,147
362
1,235
2,784
468
453
213
7,316
10,734
50 nt
98 nt
65 nt

#### Figure S16

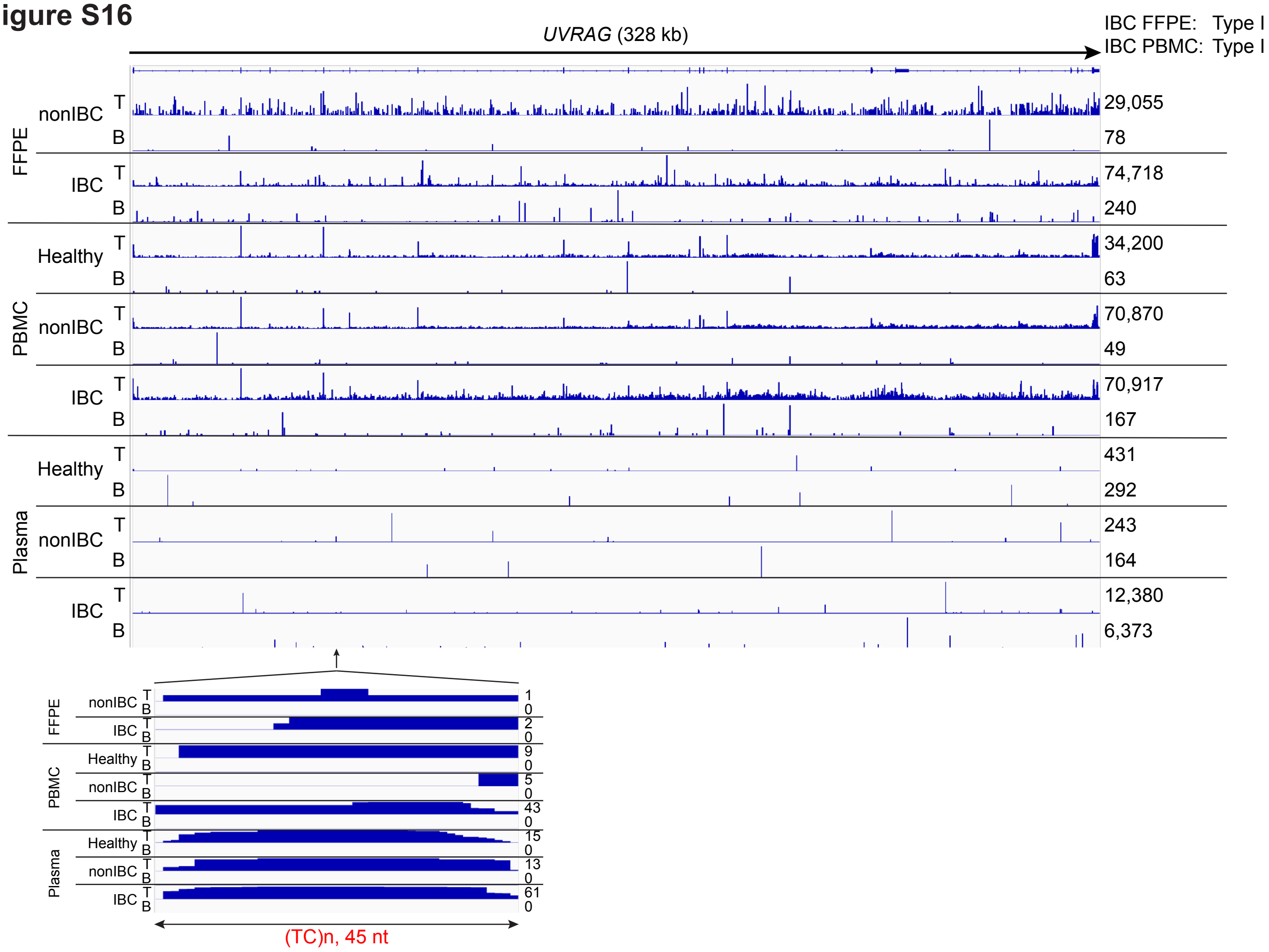

UVRAG (328 kb)
IBC FFPE: Type I
IBC PBMC: Type I
FFPE
nonIBC
IBC
PBMC
Healthy
nonIBC
IBC
Plasma
Healthy
nonIBC
IBC
T
B
29,055
78
74,718
240
34,200
63
70,870
49
70,917
167
431
292
243
164
12,380
6,373
1
0
2
0
9
0
5
0
43
0
15
0
13
0
61
0
(TC)n, 45 nt

### Figure S16

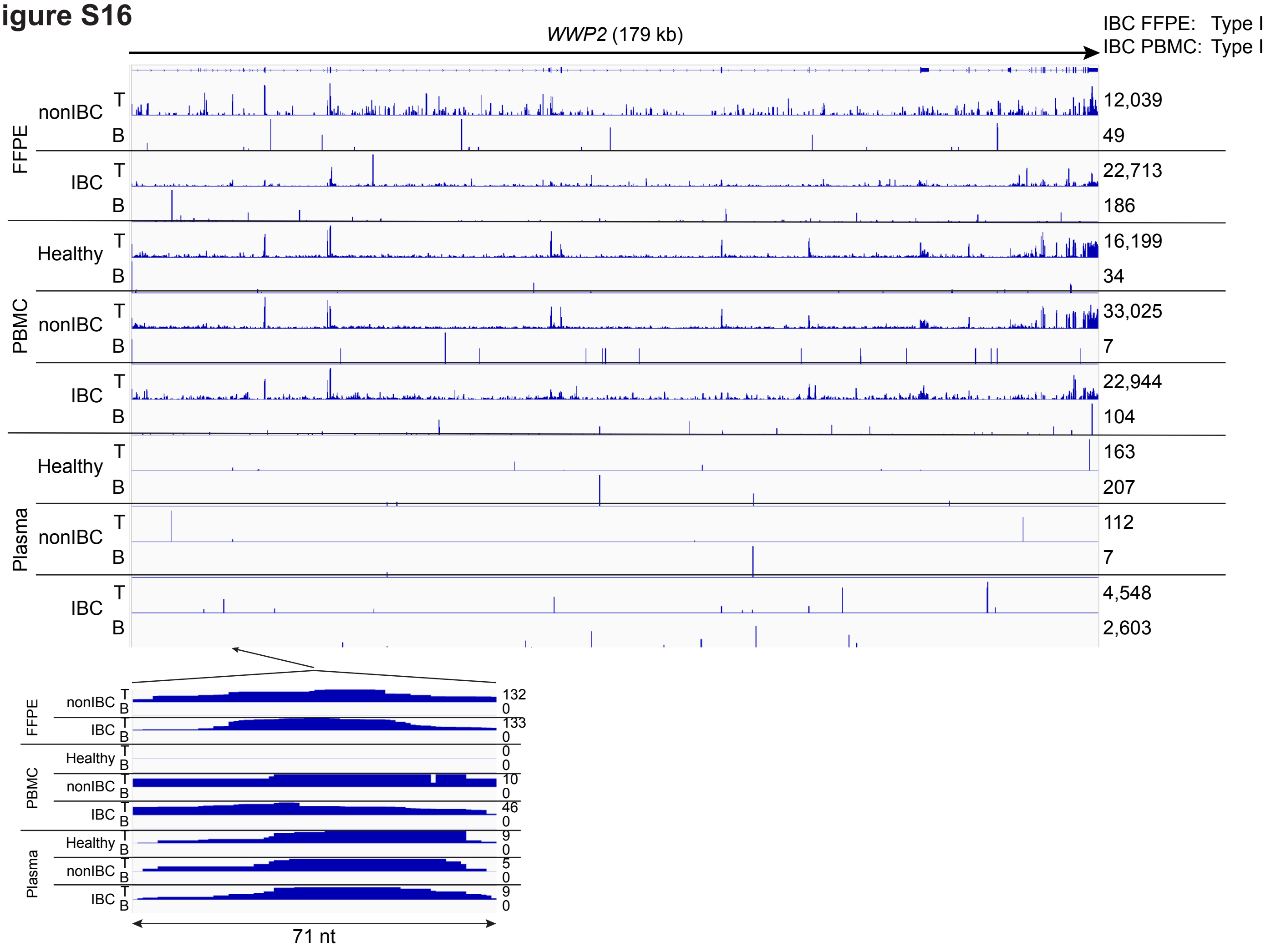

WWP2 (179 kb)
IBC FFPE: Type I
IBC PBMC: Type I
FFPE
nonIBC T B 12,039 49
IBC T B 22,713 186
PBMC
Healthy T B 16,199 34
nonIBC T B 33,025 7
IBC T B 22,944 104
Plasma
Healthy T B 163 207
nonIBC T B 112 7
IBC T B 4,548 2,603
FFPE nonIBC T B 132 0
IBC T B 133 0
PBMC Healthy T B 0 0
nonIBC T B 10 0
IBC T B 46 0
Plasma Healthy T B 9 0
nonIBC T B 5 0
IBC T B 9 0
71 nt

### Figure S16

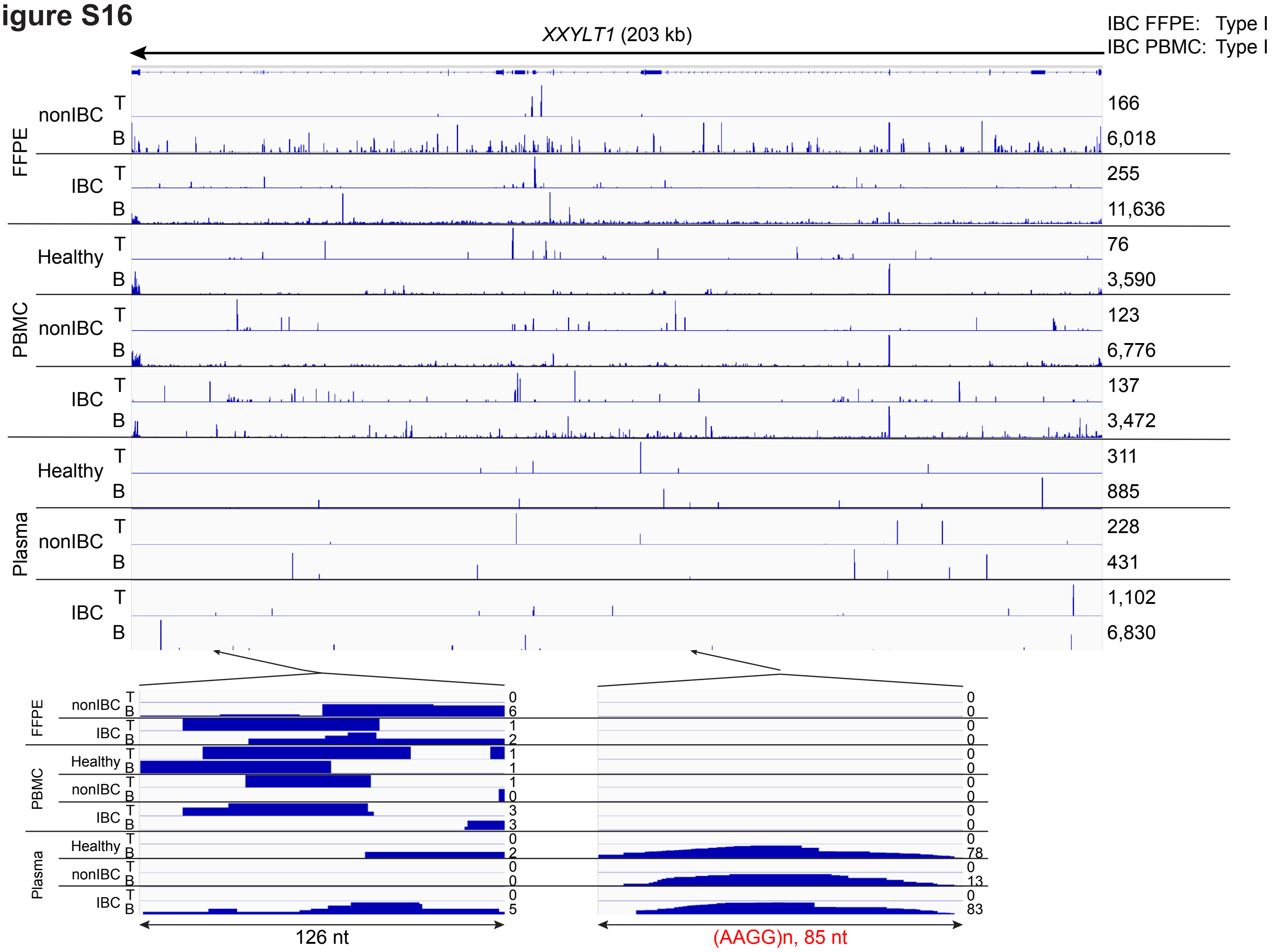
XXYLT1 (203 kb)
IBC FFPE: Type I
IBC PBMC: Type I
FFPE
PBMC
Plasma
nonIBC
IBC
Healthy
T
B
166
6,018
255
11,636
76
3,590
123
6,776
137
3,472
311
885
228
431
1,102
6,830
126 nt
(AAGG)n, 85 nt

**Fig. S16. IGV plots for protein-coding genes whose intron RNAs were significantly over-represented in IBC plasma and for which intron RNA fragments could be traced to tumors or PBMCs.** The IGV plots show coverage in combined datasets for IBC FFPE tumors, PBMC, and plasma with expanded regions at the bottom for intron RNA fragments found in 5 or more IBC plasma samples. The number of reads is indicated to the right. Types of gene based on IDR analysis in IBC FFPE tumor samples and PBMCs are indicated at the top right of each panel. Intron RNA fragments corresponding to RepBase-annotated simple repeats are indicated in red.

### Figure S17

#### A. SINEs

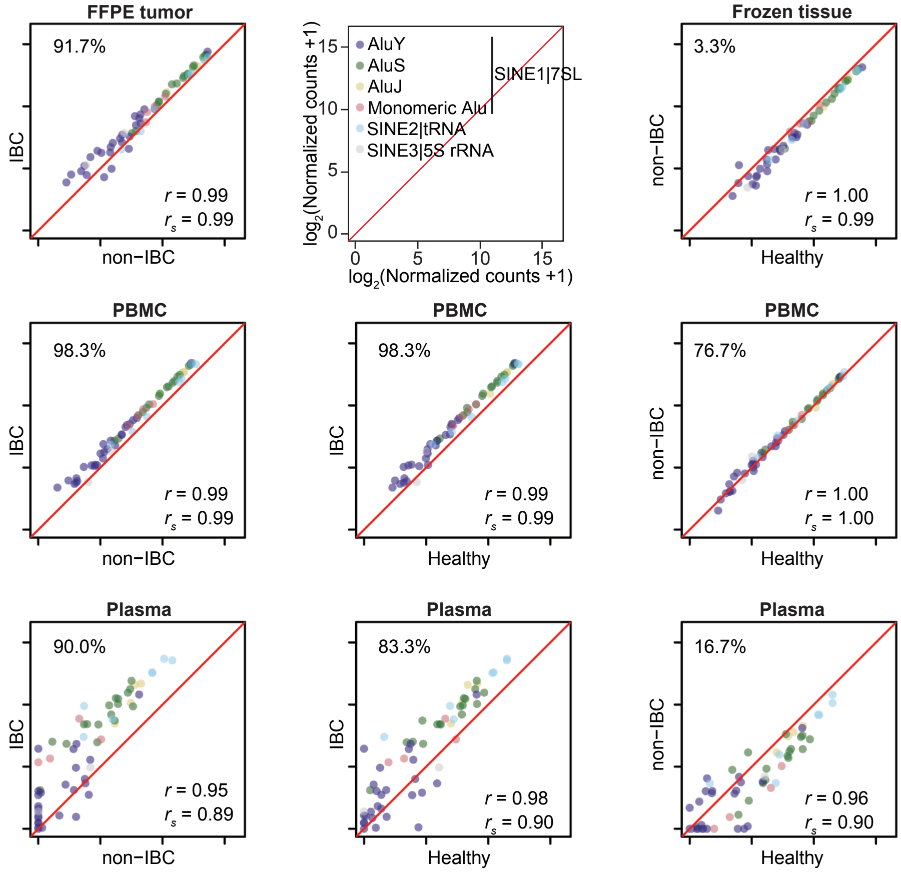


#### B. LTR-containing retroelements

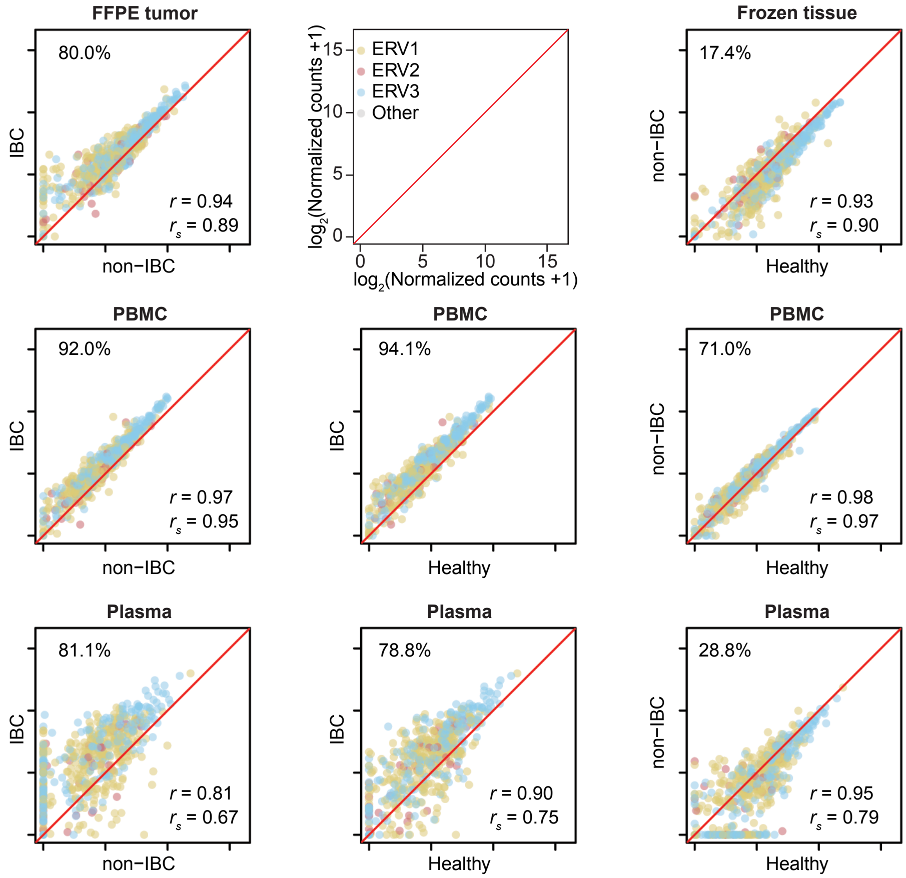

### Figure S17

#### C. DNA elements

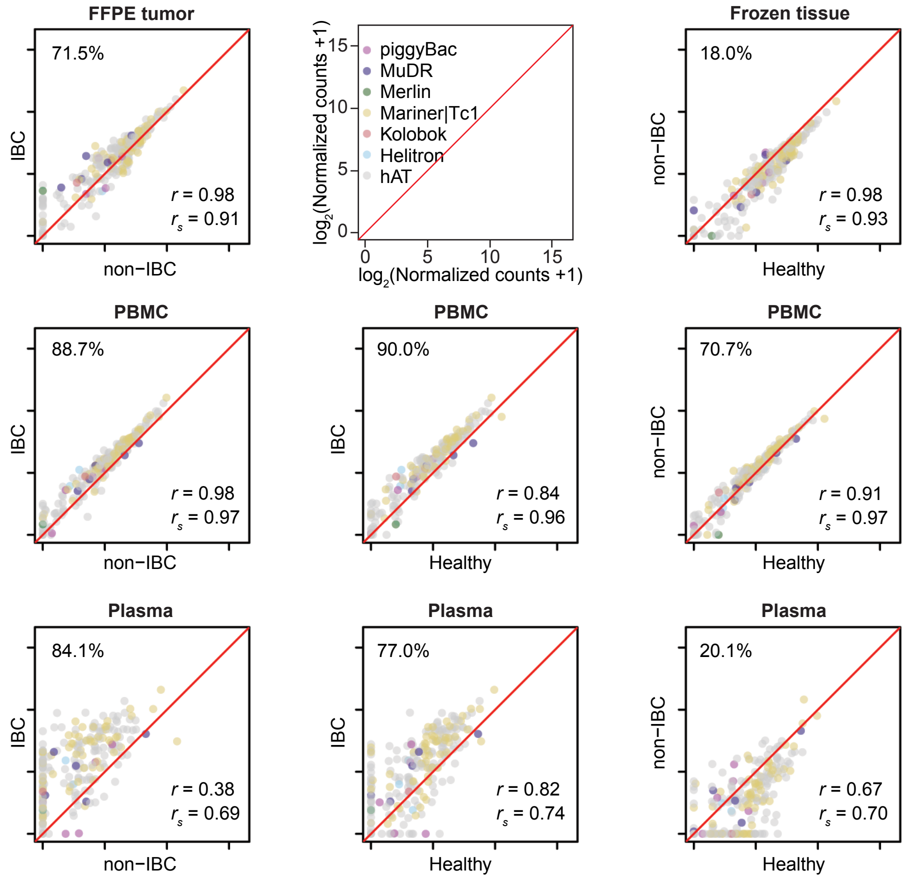


#### D. Other

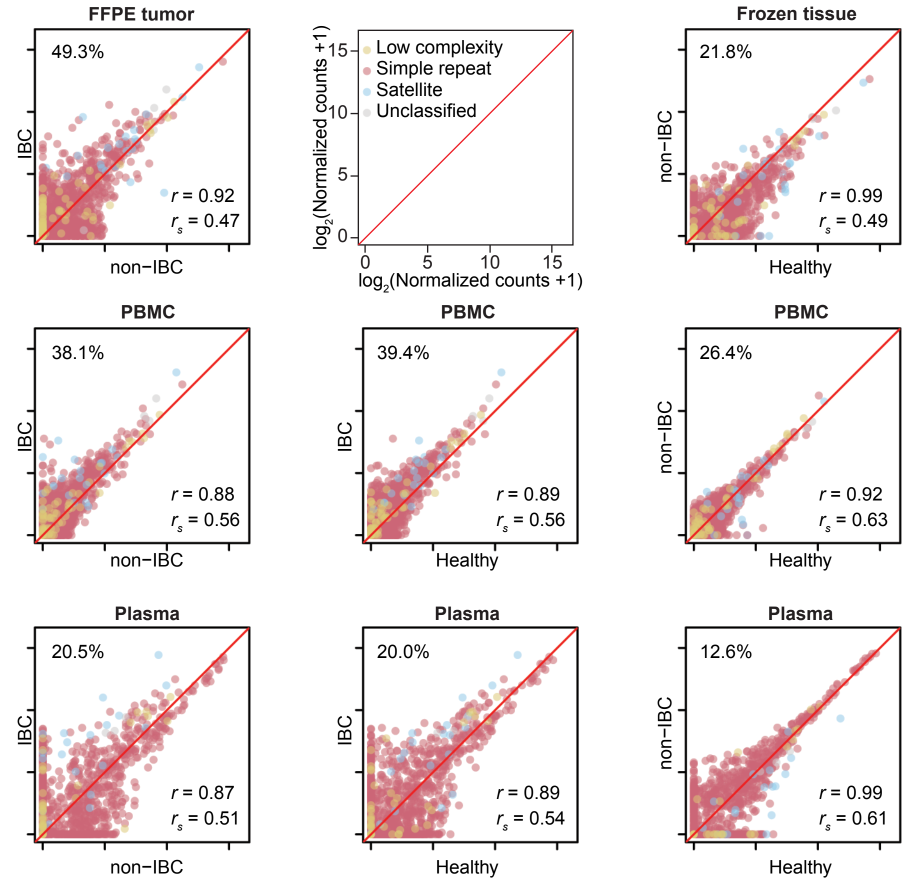

**Fig. S17. Scatter plots comparing the relative abundance of different Repeat-Element RNAs detected in breast tissues, PBMCs or Plasma.** (**A**) SINE elements, (**B**) LTR-containing retroelements, (**C**) DNA elements, and (**D**) other Repeat-Element RNAs. DESeq2-normalized counts of Repeat-Element RNAs were averaged by tissue type and disease status and $log_2$-transformed for pairwise comparisons. The numbers at the top left of each indicate the percentage of Repeat-Element RNAs above the diagonal, a measure of over-expression of Repeat-Element RNAs in each comparison. Pearson's ($r$) and Spearman's ($r_s$) correlation coefficients are shown at the bottom right. Subfamilies of the Repeat-Element RNAs are color coded as indicated in the top center box for each family of repeats.
